# “Prevalence of cancer in patients with cardiovascular diseases and risk factors: a systematic review and meta-analysis”

**DOI:** 10.64898/2026.07.04.26357301

**Authors:** Akhmetzhan Galimzhanov, Elif Beytekin, Leh Chuan Lim, Abdul Basit Ali Zai, Gemina Doolub, Mustafa Aljarshawi, Balamrit Singh Sokhal, Andrija Matetic, Rodrigo Bagur, Louise Y. Sun, Cheng Han Ng, Miguel Nobre Menezes, Sarah Zaman, Bonnie Ky, Mamas A. Mamas

## Abstract

**Background:** No systematic review has been conducted to pool the existing evidence and quantify cancer prevalence rates in cardiovascular diseases (CVDs). We aimed to estimate pooled cancer prevalence in coronary artery disease (CAD), heart failure (HF), atrial fibrillation (AF), hypertension, type 2 diabetes mellitus (DM), stroke, peripheral arterial disease (PAD), and valvular heart diseases (VHD).

**Methods:** PubMed, Web of Science, and Scopus were searched from 2010 to July 2024. The outcomes were proportions of patients with active, any, previous, blood, solid, and metastatic cancer. The prevalence rates were estimated via one-step generalized linear mixed models.

**Results:** Totally, we retrieved 676 studies with enrollment of roughly 180 million participants. The analysis for active cancer included 59 studies with a population of 4,759,695 patients. The pooled prevalence of active cancer was 4.22% (95% confidence interval (CI) 2.18-5.32), 4.43% (95% CI 2.78-6.38), 4.60% (95% CI 1.72-8.13), 4.61% (95% CI, 2.83-6.97), 4.90% (95% CI 3.84-6.37), and 5.55% (95% CI 3.97-7.01) in patients with type 2 DM, chronic HF, any stroke, CAD, VHD, and AF. For any cancer, prevalence rates ranged from 14.10% (95% CI 12.20-15.99) in AF to 7.04% (95% CI 6.05-8.03) in CAD.

**Conclusion:** Pooled prevalence rates demonstrate a measurable burden of cancer among patients with a wide range of CVDs, highlighting the need for multidisciplinary management in this population.

**Registration:** PROSPERO CRD42023494764

## 1 Introduction

Cardiovascular diseases (CVDs) and cancer are the two leading causes of mortality worldwide, accounting for ∼19 million CVD deaths annually (up from ∼13 million in 1990)^1^ and ∼20.0 million new cancer cases with 9.7–9.8 million deaths in 2022..^2^ Although distinct clinical entities, they share common risk factors and biological pathways.^3–5^ Patients with cancer have an increased risk of developing CVD due to both cancer biology and cardiotoxic effects of oncological therapies.^6–8^ Conversely, CVD may promote tumorigenesis through cytokine release, protumorigenic extracellular vesicles, immune dysregulation, and microbial dysbiosis.^3,9^ While substantial epidemiological evidence supports this bidirectional relationship,^10–13^ data on the prevalence of cancer among patients with CVD remain limited. Although several large cohort studies have reported prevalence estimates,^14–16^ no systematic review has synthesized these data across CVD populations.

Quantifying cancer prevalence in patients with CVD is important for defining the burden of coexisting disease, informing surveillance and multidisciplinary management strategies, and guiding the design of future randomized controlled trials (RCTs), in which patients with cancer and other comorbidities are often underrepresented.^17,18^ These estimates may also support healthcare planning, including the development of integrated cardio-oncology services, resource allocation, reimbursement policies, and early detection initiatives.^19^

We therefore conducted a systematic review and meta-analysis to estimate pooled cancer prevalence among patients with a broad spectrum of CVDs and cardiovascular risk factors, including coronary artery disease (CAD), heart failure (HF), atrial fibrillation (AF), hypertension, type 2 diabetes mellitus (DM), stroke, peripheral artery disease (PAD), and valvular heart disease (VHD).

## 2 Methods

### 2.1 Search strategy

This meta-analysis was based on previously published literature and did not involve human participants; therefore, ethical approval was not required. The study protocol was prospectively registered in PROSPERO (CRD42023494764). This publication represents the first stage of the project focused on estimating cancer prevalence in real-world settings. The review followed the Preferred Reporting Items for Systematic Reviews and Meta-Analyses (PRISMA) recommendations.^20^

A systematic search was conducted in PubMed, Web of Science, and Scopus. To obtain contemporary prevalence estimates and manage the large literature volume, we limited inclusion to full-length articles published after 2010. Detailed search terms, keywords, and filters are provided in the Supplementary Methods. No language restrictions were applied. Additional sources, including study registries, high-impact journal websites, and conference proceedings, were also screened to identify eligible studies. The search process flowchart was generated using a dedicated ShinyApp web tool.^21^

### 2.2 Screening and Eligibility Criteria

Eligible studies included adults (>18 years) with CAD (acute/chronic coronary syndromes, PCI), HF (acute/chronic; preserved or reduced ejection fraction), AF (including non-valvular AF), hypertension, type 2 DM, stroke (ischemic or hemorrhagic), PAD, or VHD, according to original study definitions. Pregnancy was excluded.

Studies were required to report baseline cancer prevalence (number and/or proportion). Only patient-level data were included; hospitalization-level studies were excluded because repeated admissions could bias prevalence estimates.^22–24^ Studies limited to in situ disease or benign neoplasms were excluded.

Eligible designs comprised prospective/retrospective cohort, case–control, and cross-sectional observational studies from inpatient or outpatient settings. RCTs, reviews, editorials, correspondence, conference abstracts, and propensity score–based studies were excluded because pseudo-randomization may distort real-world prevalence estimates.^25^ When multiple publications reported the same cohort, the most representative study was selected based on eligibility criteria, enrollment period, sample size, and baseline characteristics. Only studies with sample sizes ≥500 were included, consistent with recommendations for expected cancer prevalence of 3–20%.^26^

Reported outcomes included active cancer (current treatment, active, or life-threatening disease), history of cancer (≥1 year before enrollment, when reported), any cancer (undefined ascertainment), blood cancers (lymphoma, leukemia, and/or myeloma), solid cancers, and metastatic disease. Active cancer prevalence was prioritized because ascertainment methods, coding practices, and look-back periods for other outcomes were inconsistently reported, limiting interpretability.

Title/abstract and full-text screening were conducted in Rayyan.^27^ Disagreements were resolved by consensus among all reviewers.

### 2.3 Data Extraction and Risk of Bias Assessment

Data extraction was performed using the Systematic Review Database Repository Plus platform.^28^ Extracted variables included authors, publication year, study design, setting, enrollment period, country, eligibility criteria, baseline population characteristics (age, sex, race, CV risk factors, cardiovascular diseases, comorbidities), and cancer prevalence data. CV risk factors comprised DM, hypertension, dyslipidemia, obesity, smoking, and alcohol use; cardiovascular diseases included CAD, stroke, HF, AF, PAD, and VHD; comorbidities included cerebrovascular disease, liver abnormalities, CKD, gout, asthma, COPD, thyroid disorders, and anemia.

When only study arm–level statistics were reported, aggregate estimates for the target population were derived using Cochrane Handbook methods.^29^ Means and standard deviations were calculated from medians and interquartile ranges when required, according to Wan et al.^30^

Risk of bias was assessed using the Joanna Briggs Institute (JBI) Prevalence Critical Appraisal Tool, which evaluates methodological quality of prevalence studies, including sampling, measurement reliability, statistical analysis, and response rates.^31^ Data extraction and quality assessment were conducted independently by multiple reviewers, with discrepancies resolved by discussion among all authors.

### 2.4 Statistical analyses

Analyses followed the prespecified protocol (<u>CRD42023494764</u>). To avoid bias associated with traditional two-step arcsine-based meta-analytic methods, pooled prevalence was estimated using one-step generalized linear mixed models.^32,33^ Both median proportions and population-averaged prevalence estimates were calculated. Median estimates summarize included studies, whereas population-averaged estimates are interpretable at the target population level; these estimates apply only to populations meeting our predefined inclusion criteria (individuals with CVD and cardiovascular risk factors) and should not be generalized to broader populations beyond this scope. Confidence intervals (CIs) for population-averaged estimates were obtained by bootstrap resampling (1000 iterations).^32^

Heterogeneity was assessed using tau² statistics. Meta-regression analyses were performed to explore heterogeneity and derive age-adjusted prevalence estimates when ≥10 studies were available.^29^ Subgroup analyses were conducted according to United Nations Sustainable Development Goal geographic regions, with results reported only when ≥3 studies were available.^34^

Although Doi plots and LFK indices were prespecified,^35^ publication bias was assessed using multiple complementary methods, prioritizing the Thompson & Sharp test due to concerns regarding LFK false positives under high heterogeneity.^36^ Such assessment was considered supplementary, consistent with recommendations for meta-analyses of proportions.^37^

Analyses were conducted in R using code proposed by Lin et al. and the *metafor* and *lme4* packages.^32,38,39^ Certainty of evidence was graded according to GRADE Working Group guidance.^40^ Patients and the public were not involved in study design, conduct, reporting, or dissemination. Ethical approval was not required because only published data from studies with participant informed consent were used.

## 3 Results

### 3.1 Study Characteristics

A total of 676 articles (686 cohorts; 180,448,455 patients) were identified (Supplementary References); the study selection process is shown in Figure 1. Of these, 59 studies including 4,759,695 patients reported active cancer prevalence and comprised the main analysis. Study characteristics by UN geographic region are summarized in Table 1. Most cohorts originated from Europe/Northern America (n=41) and Eastern/South-Eastern Asia (n=11), whereas Oceania (n=3) and Northern Africa/Western Asia (n=1) were underrepresented. Mean participant age ranged from 63.6 years in Northern Africa/Western Asia to 77.4 years in Europe/Northern America, while female representation varied from 26.2% in Eastern/South-Eastern Asia to 53.8% in Europe/Northern America.

**Figure 1.**
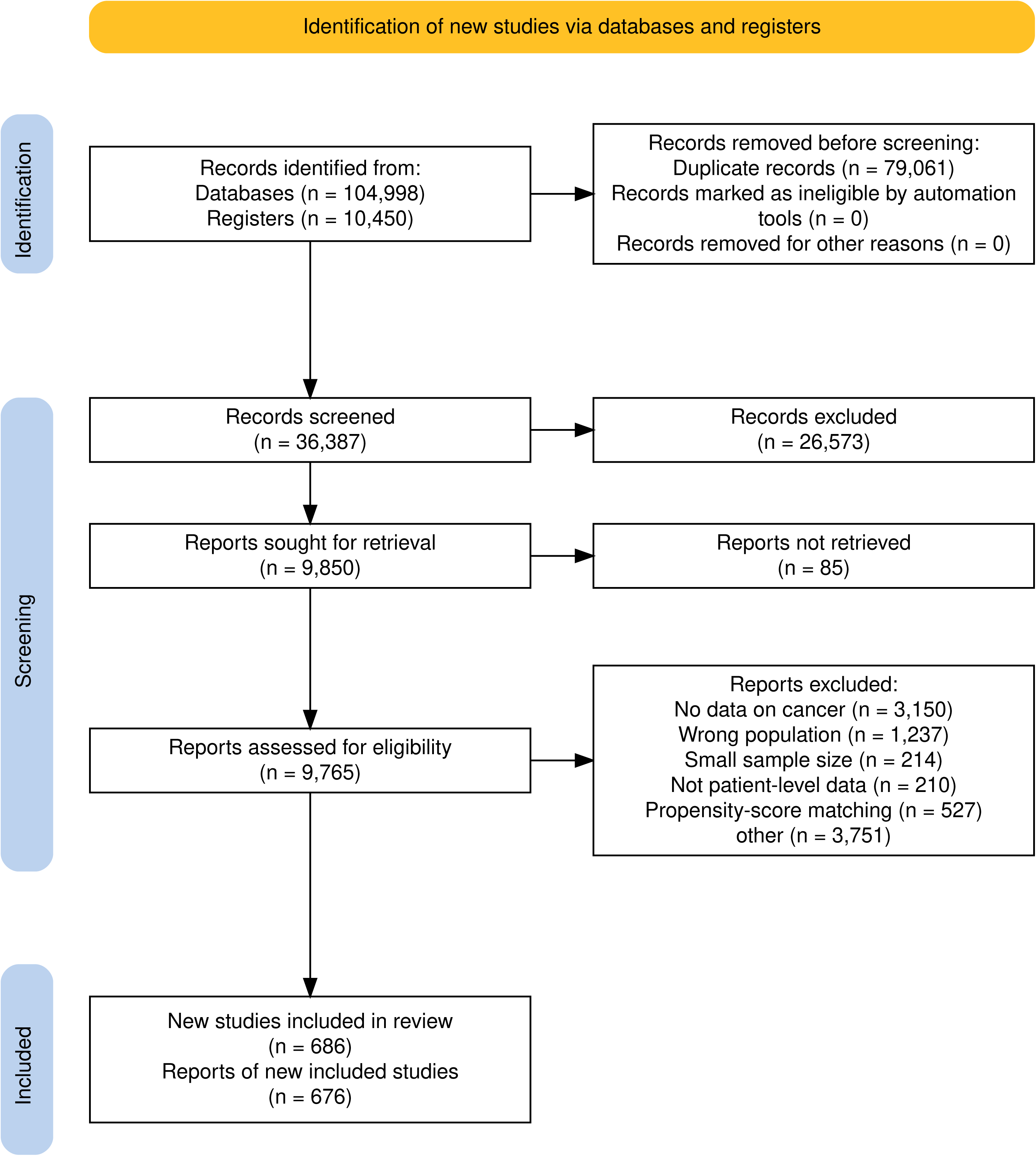
The flow-diagram for the systematic search.

**Table 1.**
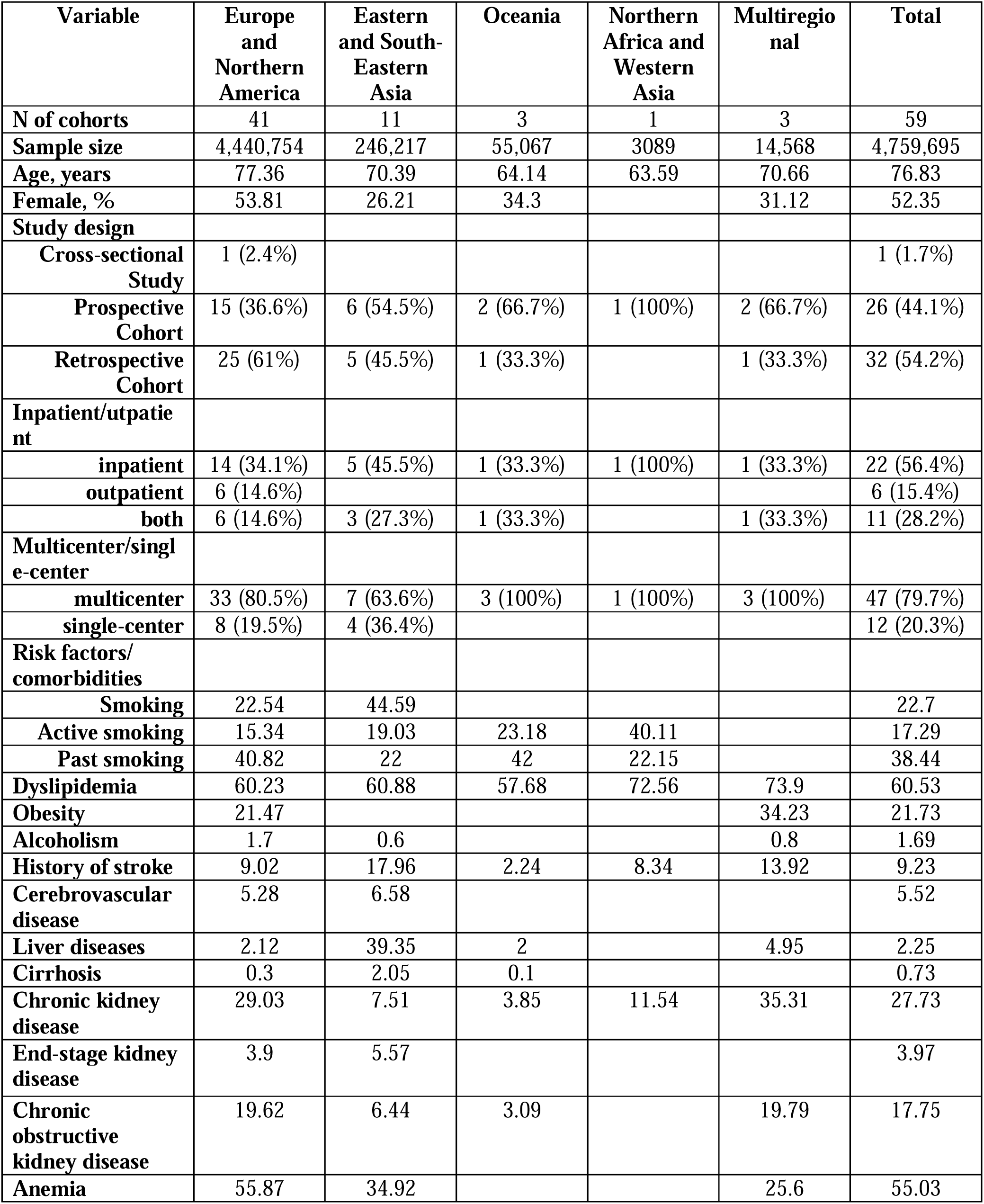
The baseline characteristics of studies with reported prevalence of active cancer across the world geographic regions.

| **Variable** | **Europe and Northern America** | **Eastern and South-Eastern Asia** | **Oceania** | **Northern Africa and Western Asia** | **Multiregional** | **Total** |
| --- | --- | --- | --- | --- | --- | --- |
| **N of cohorts** | 41 | 11 | 3 | 1 | 3 | 59 |
| **Sample size** | 4,440,754 | 246,217 | 55,067 | 3089 | 14,568 | 4,759,695 |
| **Age, years** | 77.36 | 70.39 | 64.14 | 63.59 | 70.66 | 76.83 |
| **Female, %** | 53.81 | 26.21 | 34.3 |  | 31.12 | 52.35 |
| **Study design** |  |  |  |  |  |  |
| **Cross-sectional Study** | 1 (2.4%) |  |  |  |  | 1 (1.7%) |
| **Prospective Cohort** | 15 (36.6%) | 6 (54.5%) | 2 (66.7%) | 1 (100%) | 2 (66.7%) | 26 (44.1%) |
| **Retrospective Cohort** | 25 (61%) | 5 (45.5%) | 1 (33.3%) |  | 1 (33.3%) | 32 (54.2%) |
| **Inpatient/outpatient** |  |  |  |  |  |  |
| **inpatient** | 14 (34.1%) | 5 (45.5%) | 1 (33.3%) | 1 (100%) | 1 (33.3%) | 22 (56.4%) |
| **outpatient** | 6 (14.6%) |  |  |  |  | 6 (15.4%) |
| **both** | 6 (14.6%) | 3 (27.3%) | 1 (33.3%) |  | 1 (33.3%) | 11 (28.2%) |
| **Multicenter/single-center** |  |  |  |  |  |  |
| **multicenter** | 33 (80.5%) | 7 (63.6%) | 3 (100%) | 1 (100%) | 3 (100%) | 47 (79.7%) |
| **single-center** | 8 (19.5%) | 4 (36.4%) |  |  |  | 12 (20.3%) |
| **Risk factors/comorbidities** |  |  |  |  |  |  |
| **Smoking** | 22.54 | 44.59 |  |  |  | 22.7 |
| **Active smoking** | 15.34 | 19.03 | 23.18 | 40.11 |  | 17.29 |
| **Past smoking** | 40.82 | 22 | 42 | 22.15 |  | 38.44 |
| **Dyslipidemia** | 60.23 | 60.88 | 57.68 | 72.56 | 73.9 | 60.53 |
| **Obesity** | 21.47 |  |  |  | 34.23 | 21.73 |
| **Alcoholism** | 1.7 | 0.6 |  |  | 0.8 | 1.69 |
| **History of stroke** | 9.02 | 17.96 | 2.24 | 8.34 | 13.92 | 9.23 |
| **Cerebrovascular disease** | 5.28 | 6.58 |  |  |  | 5.52 |
| **Liver diseases** | 2.12 | 39.35 | 2 |  | 4.95 | 2.25 |
| **Cirrhosis** | 0.3 | 2.05 | 0.1 |  |  | 0.73 |
| **Chronic kidney disease** | 29.03 | 7.51 | 3.85 | 11.54 | 35.31 | 27.73 |
| **End-stage kidney disease** | 3.9 | 5.57 |  |  |  | 3.97 |
| **Chronic obstructive kidney disease** | 19.62 | 6.44 | 3.09 |  | 19.79 | 17.75 |
| **Anemia** | 55.87 | 34.92 |  |  | 25.6 | 55.03 |

Most studies were retrospective cohorts (54.2%), conducted in inpatient settings (56.4%) and across multiple centers (79.7%). The pooled study population had a high burden of traditional CV risk factors and comorbidities, including smoking (22.7%), dyslipidemia (60.5%), obesity (21.7%), CKD (27.7%), COPD (17.8%), and prior stroke (9.2%) (Table 1). Study-level baseline characteristics are provided in Tables S1–S2.

### 3.2 Risk of Bias Assessment

Risk of bias results for the main analysis of active cancer prevalence are shown in Table S3. All studies were based on nationwide or large cohorts with all, consecutive, or random patient samples and were therefore judged as low risk of bias for sample representativeness, recruitment, and sample size. Although all studies reported baseline characteristics, 35/59 did not provide patient flow diagrams and/or losses to follow-up, resulting in unclear risk regarding sample coverage. Measurement reliability was also frequently unclear, as most studies did not report validated or consensus-based cancer ascertainment methods (50/59) or procedures for data quality assessment (55/59). Because none of the included studies were designed primarily to assess cancer prevalence, the final three JBI Prevalence Critical Appraisal Tool domains (statistical analyses for prevalence data, confounding, and subgroup considerations) were considered not applicable.

### 3.3 Cancer Prevalence in CAD and Its Subtypes

The main analysis of active cancer included 16 cohorts (819,468 participants), predominantly multicenter retrospective inpatient studies. Mean age was 67.9±1.7 years, with 26.5% females (Table S4). The population had a high burden of CV risk factors (hypertension 61.0%, DM 32.2%, smoking 53.6%, dyslipidemia 62.6%, obesity 42.8%) and comorbidities (CKD 10.5%, COPD 8.6%, anemia 25.6%; Table S4).

Pooled prevalence estimates for active, any, and previous cancer were 4.61% (95% CI 2.83–6.97; Tau²=0.64), 7.04% (95% CI 6.05–8.03; Tau²=0.70), and 9.72% (95% CI 6.97–14.17; Tau²=0.87), respectively (Graphical Abstract, Figure 2). Prevalence rates were broadly consistent across geographic regions. Meta-regression suggested higher active cancer prevalence in studies with greater PAD and COPD burden (Table S5). Prevalence estimates for solid cancers, hematologic malignancies, and metastatic disease were 4.39% (95% CI 2.04–7.72), 0.62% (95% CI 0.39–0.90), and 1.39% (95% CI 0.81–2.23), respectively (Table 2; Figures S1–S6). Any cancer prevalence increased with age, reaching 10.11% (95% CI 6.25–15.94) in patients >80 years (Table S6).

**Figure 2.**
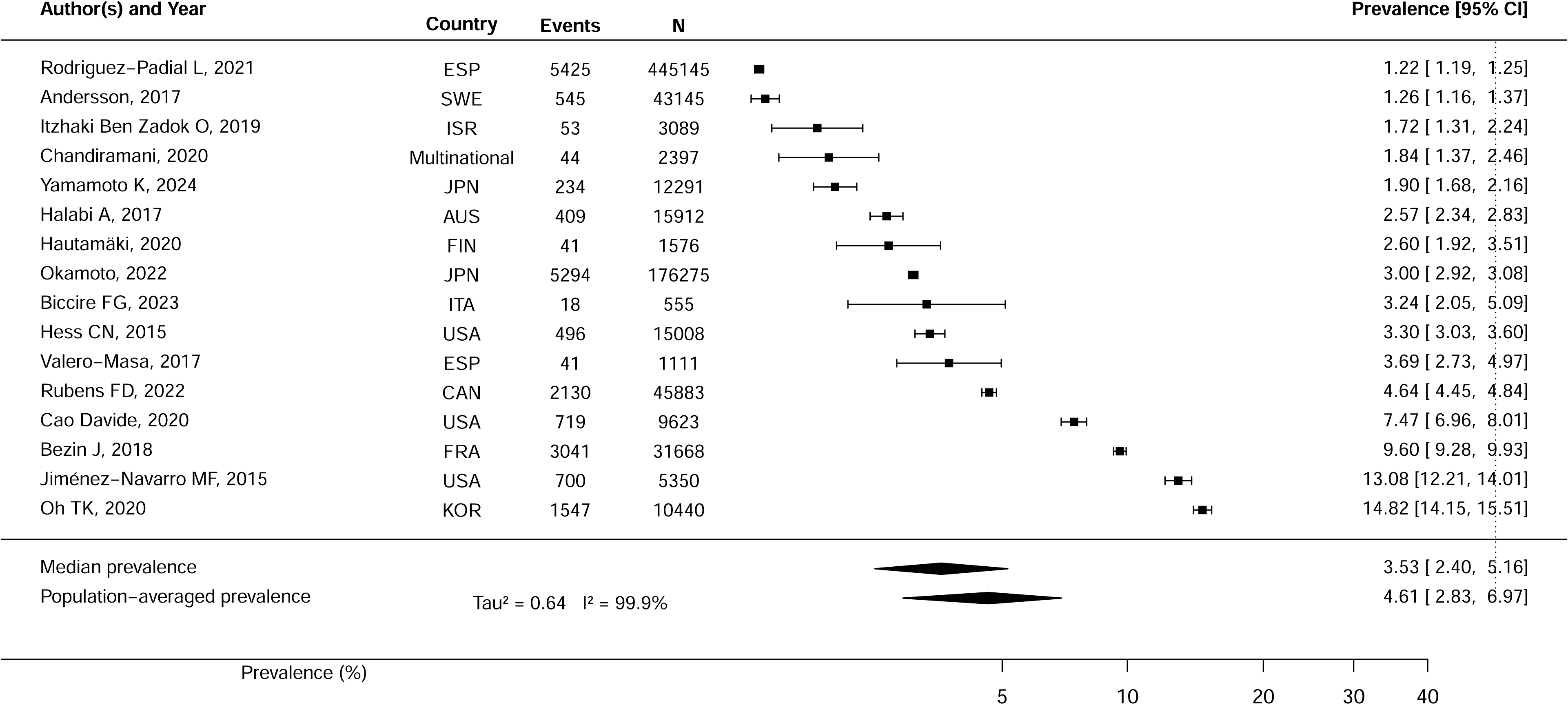
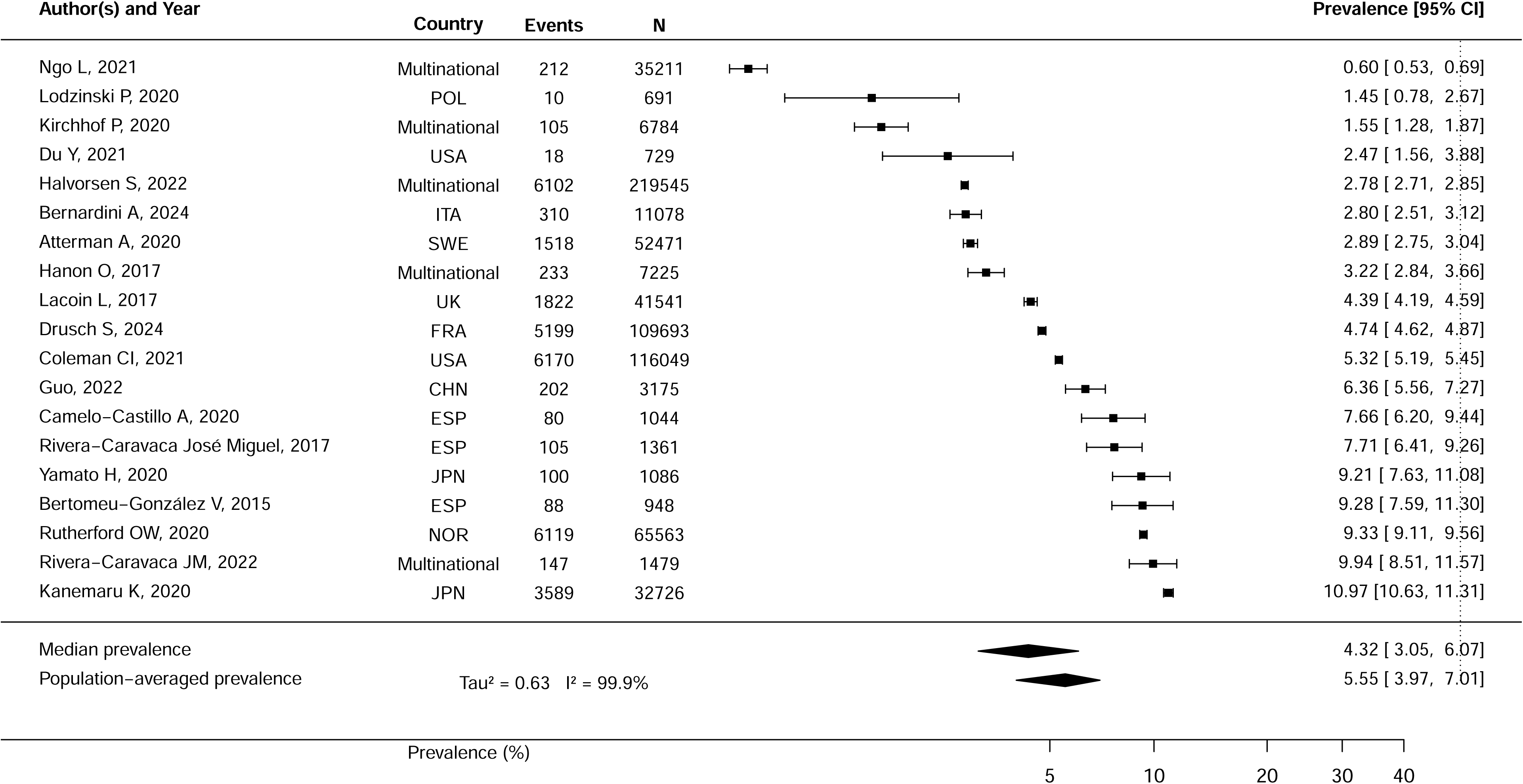
Pooled prevalence of active cancer in coronary artery disease. CI, confidence intervals.

Across CAD subtypes, active cancer prevalence was comparable in acute coronary syndrome, acute myocardial infarction, and PCI populations: 3.12% (95% CI 1.91–5.13), 2.41% (95% CI 1.56–3.03), and 4.53% (95% CI 2.52–7.39), respectively (Table S7; Figures S7–S31). Previous cancer prevalence was highest among PCI patients (13.93%, 95% CI 7.25–24.76; Tau²=1.18). Additional outcome estimates are presented in Figures S7–S31.

### 3.4 Cancer Prevalence in AF and Its Subtypes

The meta-analysis of active cancer in AF included 18 cohorts (642,836 individuals; mean age 75.9±5.2 years; 42.3% females; Table S4). Active cancer prevalence was 5.55% (95% CI 3.97–7.01; Tau²=0.63; Graphical Abstract, Figure 3), with higher rates in Eastern/South-Eastern Asia (8.88%, 95% CI 7.15–10.94; *p*=0.04; 3 studies; Table S7). Prevalence estimates for solid, hematologic, metastatic, and any cancer were 8.43% (95% CI 1.62–15.24), 1.26% (95% CI 0.88–1.72), 2.74% (95% CI 1.68–4.14), and 14.10% (95% CI 12.20–15.99), respectively (Figures S32–S37). Meta-regression indicated higher cancer prevalence in studies with greater burdens of HF, hypertension, and prior stroke (Table S5). Active cancer prevalence increased with age, reaching 2.9% (95% CI 1.80–4.63) and 6.34% (95% CI 3.97–9.99) in populations aged >70 and >80 years, respectively (Table S6).

**Figure 3.**
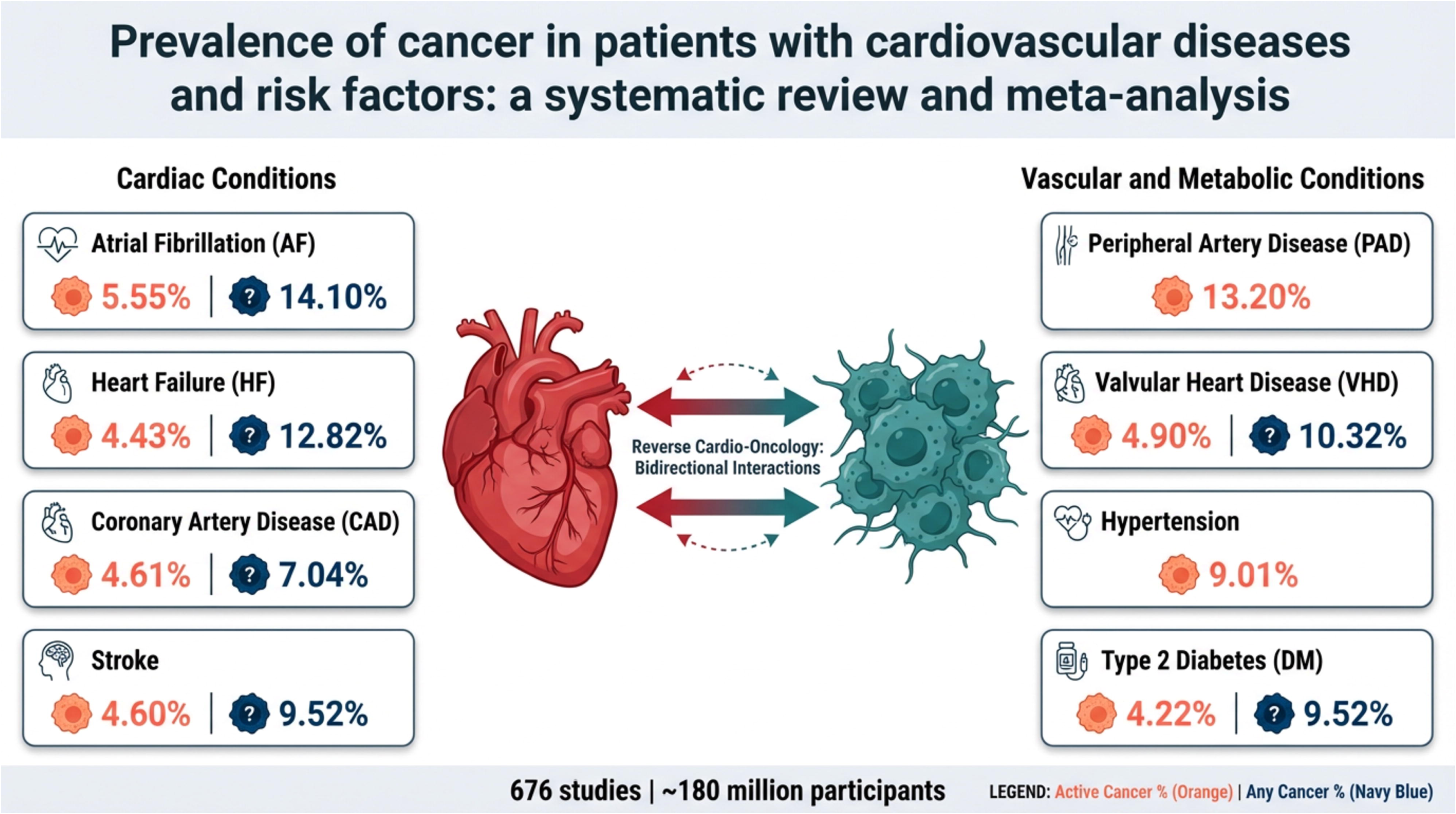
Pooled prevalence of active cancer in atrial fibrillation. CI, confidence intervals.

For non-valvular AF, 8 cohorts (279,408 patients; mean age 77.8 years; 40.6% females) showed an active cancer prevalence of 5.87% (95% CI 3.92–7.74; Tau²=0.39; Table S7). Corresponding prevalence estimates for any, previous, solid, hematologic, and metastatic cancers were 14.03% (95% CI 12.19–15.73), 14.31% (95% CI 9.36–19.94), 11.06% (95% CI 4.97–15.60), 1.27% (95% CI 0.84–1.61), and 2.95% (95% CI 1.57–5.48), respectively (Table S7; Figures S38–S43).

### 3.5 Cancer prevalence in HF and its subtypes

The data for HF were obtained from 7 cohorts with a total population of 3,070,089 participants (mean age 79.3±2.6 years, females 55.8%). The proportion of hypertension, DM, dyslipidemia, CKD, and COPD were 74.9%, 48.7%, 22.6%, 34.4%, and 32.8%, respectively (Table S4).

The pooled prevalence of active malignancies was 4.43% (95% CI 2.78-6.38, Tau^2^ = 0.36) without any identified significant discrepancies across the regions (Table S7, Figure 2, Figure S44). Regarding other outcomes, the highest estimate was identified for history of cancer at 13.06% (95% CI 8.42-18.25), while the prevalence rates for any, solid, blood cancer, and metastases were 12.82% (95% CI 10.60- 14.89), 9.46% (95% CI 5.08-12.59), 1.11% (95% CI 0.39-1.63), and 2.02 (95% CI 1.47-2.57), respectively (Table S7, Figures S45-52). The meta-regression analysis showed a positive linear association between age and prevalence of any cancer (Table S6).

For acute HF, the total prevalence of active cancer was 9.75 (95% CI 7.03-12.36, Figure S53) with other outcomes provided in Figures S54-56.

### 3.6 Cancer prevalence in stroke and its subtypes

Data from 6 cohorts (84,810 participants; mean age 76.2±5.3 years; 47.8% female) were included (Table S4). The pooled prevalence of active cancer in stroke was 4.60% (95% CI 1.72–8.13, Tau²=1.15; Table S7). The prevalence of any, previous, solid, hematologic, and metastatic cancers was 9.52% (95% CI 7.33–12.19), 13.81% (95% CI 10.90–16.60), 7.02% (95% CI 2.86–11.57), 0.65% (95% CI 0.33–0.94), and 1.19% (95% CI 0.80–1.57), respectively (Table S7, Figure 2, Figures S57–62). Any cancer was most prevalent in patients aged >70 and >80 years (7.6%, 95% CI 5.72–10.05; 9.47%, 95% CI 6.53–13.53; Table S6). By subtype, active cancer prevalence was 5.26% (95% CI 2.40–8.91, Tau²=0.61; Figures S63–67) in ischemic stroke, while any cancer prevalence reached 11.49% (95% CI 6.49–16.97, Tau²=0.41; Figure S68) in hemorrhagic stroke.

### 3.7 Cancer prevalence in PAD

Data on active cancer prevalence in PAD were insufficient for meta-analysis. Analysis of 16 studies (6,246,746 participants; mean age 71.2±2.5 years; 42.5% female) showed prevalences of HF, hypertension, DM, dyslipidemia, prior stroke, and CKD of 21.9%, 59.6%, 44.1%, 63.1%, 12.9%, and 30.3%, respectively (Table S8). The pooled prevalence of any cancer was 13.20% (95% CI 9.11–17.61, Tau²=0.78, I²=100%; Table S7, Figure 2, Figure S69), driven mainly by European and North American studies. Metastatic cancer prevalence was 4.44% (95% CI 3.25–5.45; 3 studies, 27,662 patients; Figure S70).

### 3.8 Cancer prevalence in VHD and its subtypes

Eight cohorts (151,839 participants; mean age 81.5±0.9 years; 48.1% female) were analyzed. Common comorbidities included hypertension (92.3%), DM (39.8%), dyslipidemia (42.6%), prior stroke (16.6%), liver disease (3.8%), CKD (32.7%), COPD (37.4%), and anemia (58.3%) (Table S4). Active cancer prevalence in the overall cohort was 4.90% (95% CI 3.84–6.37, Tau²=0.12; Table S7). The prevalence of prior, any, hematologic, and metastatic cancers was 15.69% (95% CI 12.31–18.65), 10.32% (95% CI 8.28–12.35), 1.48% (95% CI 0.99–1.83), and 1.56% (95% CI 0.37–2.94), respectively (Figure 2, Figures S71–75). In aortic stenosis, active, prior, and any cancer prevalence was 4.42% (95% CI 3.64–5.21), 17.74% (95% CI 14.60–20.03), and 10.53% (95% CI 7.70–13.36), respectively; corresponding estimates after TAVI were 4.87% (95% CI 4.11–5.59), 18.28% (95% CI 14.40–20.71), and 10.63% (95% CI 7.12–15.01) (Figures S76–81). Age-stratified prevalence of any cancer is provided in Table S6.

### 3.9 Cancer Prevalence in Hypertension and Type 2 DM

For type 2 DM, active cancer data were derived from 3 cohorts (n=117,866; mean age 72±1.4 years; 40.1% females; Table S4). Active cancer prevalence was 4.22% (95% CI 2.18–5.32; Tau²=0.99; Table S7, Figure S82). Prevalence of any cancer, prior malignancy, and metastases was 9.52% (95% CI 8.25–11.02), 12.72% (95% CI 4.87–23.94), and 1.00% (95% CI 0.53–1.38), respectively (Figures S83–S85).

For hypertension, meta-analysis was limited to any cancer due to scarce data. One study (Alanaeme et al.) contributed weighted survey data (∼97 million individuals), substantially influencing pooled estimates; sensitivity analyses were performed excluding this study. After exclusion, the pooled cohort comprised 721,074 individuals (mean age 69.2±8.4 years), with high prevalence of smoking (48.1%), dyslipidemia (43.4%), DM (29.2%), cerebrovascular disease (19.6%), and asthma (16.7%) (Tables S9–S10).

Pooled prevalence of any cancer in hypertension was 9.30% (95% CI 5.50–13.33; Tau²=1.67; I²=100%; without Alanaeme et al.; Figure 2, Figure S86) and 9.01% (95% CI 5.16–13.54; Tau²=1.72; I²=100%; Table S7; Figure S87). Age distribution is presented in Table S6.

### 3.10 The assessment of publication bias

The results of publication bias assessment were valid for those analyses that included 10 or more studies. No statistically significant asymmetry was detected for meta-analyses for prevalence of active cancer in patients with CAD, AF, and non-valvular AF (Table S11).

### 3.11 Cancer Prevalence in total study population

The prevalence rates of active, any, prior, hematologic, metastatic, and solid cancers was 5.36 (95% CI 4.39-6.33), 10.27 (95% CI 9.62-10.92), 11.72 (10.10-13.49), 0.97 (95% CI 0.73-1.20), 1.73 (95% CI 1.38-2.13), and 7.15 (95% CI 4.77-9.41), respectively (Figures S88-93).

## 4 Discussion

To our knowledge, this is the first systematic review and meta-analysis summarizing cancer prevalence across common cardiovascular (CV) diseases and risk factors globally. Pooled prevalence of active cancer was relatively consistent across conditions, ranging from 4.22% (95% CI 2.18–5.32) in type 2 DM to 5.55% (95% CI 3.97–7.01) in AF. Prevalence of any cancer was highest in AF (14.10%, 95% CI 12.20–15.99), PAD (13.2%, 95% CI 9.11–17.61), and chronic HF (12.82%, 95% CI 10.6–14.9), followed by VHD (10.32%, 95% CI 8.28–12.35), type 2 DM (9.52%, 95% CI 8.25–11.02), hypertension (9.01%, 95% CI 5.16–13.54), and CAD (7.04%, 95% CI 6.05–8.03). We also report pooled estimates for prior malignancy, hematologic and solid cancers, advanced cancer, and demonstrate positive associations between cancer prevalence, increasing age, and comorbidity burden.

Active cancer estimates are likely more clinically interpretable than those for any cancer because most studies explicitly defined active disease as current treatment, recent diagnosis, or life-threatening malignancy. By contrast, definitions of any cancer were often poorly specified regarding look-back periods and ascertainment methods, potentially encompassing both remote and recent malignancies. This variability likely contributed to substantial heterogeneity; therefore, estimates for any cancer should be interpreted cautiously and viewed as hypothesis-generating.

We could not determine whether active cancer prevalence in CVD populations exceeds that of the general population, as such comparisons were beyond our scope. Nevertheless, published population-based studies report markedly lower cancer prevalence estimates, including GLOBOCAN 1-year and 5-year prevalence estimates of approximately 0.18% and 0.68%, respectively,^42^ ∼1.5% 5-year prevalence in multinational registry analyses,^43^ and 1.85% in the French general population.^44^ Our pooled active cancer prevalence was approximately 5%, although differences in age structure and confounding factors may partly explain this discrepancy.

Cancer and CVD share a complex bidirectional relationship.^3,45^ Common risk factors—including aging, smoking, obesity, dyslipidemia, DM, hypertension, sedentary lifestyle, diet, and alcohol use—drive chronic inflammation, oxidative stress, endothelial dysfunction, metabolic derangements, immune dysregulation, DNA damage, and cellular proliferation, promoting both atherosclerosis and carcinogenesis.^3–5,45^ Cancer can promote CVD through systemic inflammation, prothrombotic states, tumor-derived mediators, and treatment-related cardiotoxicity.^3–5,45^

While the cardiovascular consequences of cancer and its therapies are well established,^8,46^ growing translational and epidemiological evidence suggests that CVD itself may facilitate malignant transformation, giving rise to the emerging field of reverse cardio-oncology.^3,9,45,47^ Systematic reviews and large registries consistently report higher cancer incidence among patients with CVD and CV risk factors than in the general population.^10–13^ Proposed mechanisms include hematopoietic reprogramming, protumorigenic cytokines and extracellular vesicles, gut microbiome dysbiosis, and immunosuppressive pathways enabling tumor immune escape.^3,9,48–51^

### 4.1 Study limitations

Several limitations should be acknowledged. First, the limited number of studies precluded age-adjusted prevalence estimates for most populations except AF, restricting comparisons with the general population. Second, heterogeneity in cancer ascertainment across countries, databases, and institutions likely contributed to between-study variability; to mitigate this, we applied the population-averaged approach proposed by Lin et al.^32^ Third, limited data prevented subgroup analyses by cancer stage, sex, and race/ethnicity. Finally, individual patient-level meta-analysis would likely yield more precise estimates than aggregated published data but was not feasible.

## 5 Conclusion

We estimated pooled cancer prevalence across a broad range of CV diseases and risk factors. Active cancer prevalence ranged from 4.22% in type 2 DM to 5.55% in AF, while prevalence of any cancer ranged from 14.10% in AF, 13.2% in PAD, and 12.82% in chronic HF to 7.04% in CAD. These findings underscore the need for integrated, multidisciplinary approaches to screening, risk stratification, and clinical management in this high-risk population.

## Supporting information

Prisma

Supplementary references

## Data Availability

All data produced in the present study are available upon reasonable request to the authors

