## Supplementary Materials for "“Prevalence of cancer in patients with cardiovascular diseases and risk factors: a systematic review and meta-analysis”"

**Supplementary Materials for the article titled “Prevalence of cancer in patients with cardiovascular diseases and risk factors: a systematic review and meta-analysis”.**

**Content**

|  | Page |
| --- | --- |
| Supplementary Methods. Search strategy. | 2 |
| Table S1. The baseline characteristics of studies with reported prevalence of active cancer. | 4 |
| Table S2. The baseline characteristics of total population across the world geographic regions with consideration of all studies. | 8 |
| Table S3. The risk of bias assessment. | 10 |
| Table S4. Baseline characteristics studies included in meta-analysis of prevalence of active cancer categorized by cardiovascular diseases. | 13 |
| Table S5. The results of meta-regression analyses. | 14 |
| Table S6. The age-standardized prevalence ratios of cancer according to meta-regression analyses. | 16 |
| Table S7. The pooled prevalence rates of cancer obtained during the main and sensitivity analyses and subgroup analyses across different geographical regions. | 17 |
| Table S8. Baseline characteristics studies included in meta-analysis of prevalence of any cancer in patients with peripheral artery disease. | 22 |
| Table S9. Baseline characteristics studies included in meta-analysis of prevalence of unspecified cancer in patients with hypertension. | 23 |
| Table S10. Baseline characteristics studies included in meta-analysis of prevalence of any cancer in patients with hypertension after exclusion of Alanaeme et al’s study. | 24 |
| Table S11. The results of publication bias assessment. | 25 |

**Supplementary Methods. Search strategy.**

PubMed - 38,827 results 13.07.24

#1

"Cardiovascular Diseases"[MeSH Terms] OR "Coronary Thrombosis"[MeSH Terms] OR "Myocardial Revascularization"[MeSH Terms] OR "Coronary Stenosis"[MeSH Terms] OR "Acute Coronary Syndrome"[MeSH Terms] OR "Coronary Artery Disease"[MeSH Terms] OR "Coronary Disease"[MeSH Terms] OR "Myocardial Infarction"[MeSH Terms] OR "angina, stable"[MeSH Terms] OR "Myocardial Ischemia"[MeSH Terms] OR "Heart Failure"[MeSH Terms] OR "heart failure, diastolic"[MeSH Terms] OR "heart failure, systolic"[MeSH Terms] OR "Atrial Fibrillation"[MeSH Terms] OR "Stroke"[MeSH Terms] OR "stroke, lacunar"[MeSH Terms] OR "Hemorrhagic Stroke"[MeSH Terms] OR "Embolic Stroke"[MeSH Terms] OR "Thrombotic Stroke"[MeSH Terms] OR "Ischemic Stroke"[MeSH Terms] OR "death, sudden, cardiac"[MeSH Terms] OR "Heart Arrest"[MeSH Terms] OR "Acute Coronary Syndrome"[Title/Abstract] OR "chronic coronary syndrome"[Title/Abstract] OR "Coronary Artery Disease"[Title/Abstract] OR "Myocardial Infarction"[Title/Abstract] OR "ischemic heart disease"[Title/Abstract] OR "ischaemic heart disease"[Title/Abstract] OR "percutaneous coronary intervention"[Title/Abstract] OR "unstable angina"[Title/Abstract] OR "stable angina"[Title/Abstract] OR "Intracranial bleeding"[Title/Abstract] OR "Hemorrhagic Stroke"[Title/Abstract] OR "Ischemic Stroke"[Title/Abstract] OR "Heart Failure"[Title/Abstract] OR "Atrial Fibrillation"[Title/Abstract] OR "Hypertension"[MeSH Terms] OR "Essential Hypertension"[MeSH Terms] OR "Arterial Hypertension"[Title/Abstract] OR "Diabetes Mellitus"[MeSH Terms] OR "Diabetes Complications"[MeSH Terms] OR "diabetes mellitus, type 2"[MeSH Terms] OR "diabetes mellitus, type 1"[MeSH Terms] OR "Diabetes Mellitus"[Title/Abstract] OR "Peripheral Vascular Diseases"[MeSH Terms] OR "Peripheral Arterial Disease"[MeSH Terms] OR "Peripheral Vascular Diseases"[Title/Abstract] OR "Peripheral Arterial Disease"[Title/Abstract] OR "Peripheral Artery Disease"[Title/Abstract] OR "peripheral vascular occlusive disease"[Title/Abstract] OR "Heart Valve Diseases"[MeSH Terms] OR "Aortic Valve Disease"[MeSH Terms] OR "Mitral Valve Insufficiency"[MeSH Terms] OR "Mitral Valve Stenosis"[MeSH Terms] OR "Pulmonary Valve Insufficiency"[MeSH Terms] OR "Pulmonary Valve Stenosis"[MeSH Terms] OR "Tricuspid Valve Insufficiency"[MeSH Terms] OR "Tricuspid Valve Stenosis"[MeSH Terms] OR "valvular heart disease"[Title/Abstract]

#2

"real-world study"[Title/Abstract:~1] OR "real-world cohort"[Title/Abstract:~1] OR "real-world database"[Title/Abstract:~1] OR "real-world dataset"[Title/Abstract:~1] OR "real-world registry"[Title/Abstract:~1] OR "population-based study"[Title/Abstract:~1] OR "population-based cohort"[Title/Abstract:~1] OR "population-based database"[Title/Abstract:~1] OR "population-based dataset"[Title/Abstract:~1] OR "population-based registry"[Title/Abstract:~1] OR "nationwide study"[Title/Abstract:~1] OR "nationwide cohort"[Title/Abstract:~1] OR "nationwide database"[Title/Abstract:~1] OR "nationwide dataset"[Title/Abstract:~1] OR "nation* registr*"[Title/Abstract] OR "administrative registry"[Title/Abstract:~1] OR "administrat* registr*"[Title/Abstract] OR "administrat* database"[Title/Abstract] OR "administrative database"[Title/Abstract:~1] OR "administrat* dataset"[Title/Abstract] OR "administrative dataset"[Title/Abstract:~1] OR "healthcare registry"[Title/Abstract:~1] OR "hospital* registry"[Title/Abstract] OR "Routinely Collected Health Data"[Mesh] OR "Registries"[Mesh] OR "Electronic Health Records"[Mesh] OR "Comorbidity"[Mesh]

#3 #1 AND #2

#4

"Adaptive Clinical Trial"[pt] OR "Address"[pt] OR "Autobiography"[pt] OR "Bibliography"[pt] OR "Biography"[pt] OR "Case Reports"[pt] OR "Clinical Conference"[pt] OR "Clinical Trial Protocol"[pt] OR "Clinical Trial"[pt] OR "Clinical Trial, Phase I"[pt] OR "Clinical Trial, Phase II"[pt] OR "Clinical Trial, Phase III"[pt] OR "Clinical Trial, Phase IV"[pt] OR "Clinical Trial, Veterinary"[pt] OR "Comment"[pt] OR "Congress"[pt] OR "Consensus Development Conference"[pt] OR "Consensus Development Conference, NIH"[pt] OR "Controlled Clinical Trial"[pt] OR "Dictionary"[pt] OR "Directory"[pt] OR "Editorial"[pt] OR "Guideline"[pt] OR "Interactive Tutorial"[pt] OR "Interview"[pt] OR "Lecture"[pt] OR "Legal Case"[pt] OR "Legislation"[pt] OR "Letter"[pt] OR "Meta-Analysis"[pt] OR "News"[pt] OR "Observational Study, Veterinary"[pt] OR "Patient Education Handout"[pt] OR "Personal Narrative"[pt] OR "Portrait"[pt] OR "Practice Guideline"[pt] OR "Preprint"[pt] OR "Pragmatic Clinical Trial"[pt] OR "Randomized Controlled Trial"[pt] OR "Randomized Controlled Trial, Veterinary"[pt] OR "Retracted Publication"[pt] OR "Retraction of Publication"[pt] OR "Review"[pt] OR "Systematic Review"[pt] OR "Webcast"[pt]

#5 #4 NOT #3

###### Table S1. The baseline characteristics of studies with reported prevalence of active cancer.

| **PMID** | **Publication year** | **Study design** | **Country** | **Sample size, n** | **CAD,%** | **HF, %** | **AF, %** | **HTN, %** | **DM, %** | **PAD, %** | **VHD, %** | **Mean age, y** | **Female, %** | **Smoking, %** | **Dyslipidemia, %** | **Obesity, %** | **History of stroke, %** | **Liver diseases, %** | **CKD, %** | **COPD, %** | **Anemia, %** |
| --- | --- | --- | --- | --- | --- | --- | --- | --- | --- | --- | --- | --- | --- | --- | --- | --- | --- | --- | --- | --- | --- |
| 28927174 | 2017 | Prospective | Australia | 15,912 | 100 |  | 10 | 62.6 | 26.9 |  |  | 65.6 | 30.4 |  | 57.6 |  |  |  |  |  |  |
| 28875506 | 2018 | Retrospective | France | 31,668 | 100 | 20.2 |  | 61.4 | 30.9 | 13.4 |  | 65 | 30.2 |  | 18.9 |  |  |  |  | 3.9 |  |
| 28280146 | 2017 | Retrospective | Sweden | 43,145 | 100 | 2.02 |  |  | 12.5 | 0.55 |  | 64 |  |  |  |  | 5.81 |  |  | 1.57 |  |
| 37201612 | 2023 | Prospective | Italy | 555 | 100 | 6.49 | 8.11 | 67.21 | 29.37 | 18.38 |  | 65.6 |  | 69.91 |  |  | 5.59 |  | 11.89 | 7.75 |  |
| 30883428 | 2019 | Prospective | Israel | 3,089 | 100 |  |  | 64.64 | 39.15 | 7.85 |  | 63.59 |  |  | 72.56 |  | 8.34 |  | 11.54 |  |  |
| 32561143 | 2021 | Retrospective | Spain | 445,145 | 100 | 25.56 |  | 53.58 | 32.25 |  | 13.95 | 68.76 |  |  |  |  | 0.92 | 0.38 | 12.92 | 8.74 |  |
| 31775530 | 2020 | Retrospective | Finland | 1,576 | 100 | 29.5 |  |  | 27.6 | 8.1 |  | 69.3 | 30.9 |  |  |  |  | 0.7 | 2.2 | 5 |  |
| 35965069 | 2022 | Retrospective | Japan | 176,275 | 100 |  | 2.4 | 68 | 30 | 4 |  | 68.6 | 25.4 |  | 65.7 |  |  |  | 4.6 | 2.7 |  |
| 27865662 | 2017 | Prospective | Spain | 1,111 | 100 | 2.97 | 4.23 | 54.1 | 21.42 | 4.32 |  | 64.1 | 23.2 |  | 46.17 |  | 4.95 |  | 7.29 | 7.11 |  |
| 28331926 | 2017 | Prospective | Spain | 1,361 | 18.74 | 31.52 | 100 | 82 | 26.67 | 7.42 |  | 76 |  |  | 32.55 |  | 19.62 | 1.32 | 10.58 |  |  |
| 38657869 | 2024 | Retrospective | Italy | 11,078 | 16.9 |  | 100 | 80.2 | 20.5 | 5.8 |  | 77 | 45.7 |  |  |  | 15.2 |  |  | 12.2 |  |
| 34390242 | 2022 | Prospective | China | 3,175 |  |  | 100 |  |  |  |  |  |  |  |  |  |  |  |  |  | 10.8 |
| 34244745 | 2022 | Retrospective | Multinational | 219,545 | 22.8 | 19.2 | 100 | 66.1 | 16.7 | 6.82 |  | 74.4 | 43.2 |  |  |  | 12.7 | 0.92 | 5.43 | 12.4 |  |
| 33963402 | 2021 | Retrospective | Multinational | 35,211 | 9.76 | 8.78 | 100 | 12.31 | 9.34 | 1.43 | 3.88 | 62.29 |  |  |  |  | 1.26 |  | 2.94 | 1.65 |  |
| 28951401 | 2017 | Cross-sectional Study | UK | 41,541 | 26.08 | 18.04 | 100 | 97.68 |  |  |  | 78 |  |  |  |  | 20.02 | 0.25 | 32.92 |  |  |
| 33214605 | 2020 | Retrospective | Japan | 1,086 | 25.1 | 58.4 | 100 | 83.1 | 25.8 | 5.8 |  | 73 | 32.4 |  | 49.7 |  |  |  |  | 6.9 |  |
| 32506417 | 2020 | Retrospective | Spain | 1,044 | 20.11 | 29.02 | 100 | 82.18 | 26.63 |  |  | 76 |  |  | 33.43 |  | 19.54 |  | 10.54 |  |  |
| 32386073 | 2020 | Prospective | Sweden | 52,471 | 34.01 | 31.34 | 100 | 82.94 | 23.75 |  |  | 80.2 |  |  |  | 2.53 | 19.11 | 1.33 | 6.16 | 6.92 |  |
| 32079476 | 2020 | Retrospective | Multinational | 6,784 |  | 18.65 | 100 |  | 19.65 |  |  | 71.5 |  |  |  |  | 13.78 | 2.02 |  |  |  |
| 31983727 | 2020 | Prospective | Japan | 32,726 |  | 37.47 | 100 | 75.22 | 26.99 | 3.43 | 12.24 | 81.5 |  |  | 42.43 |  | 20.83 |  | 20.71 |  |  |
| 31976927 | 2020 | Retrospective | Poland | 691 | 35.31 | 50.65 | 100 | 57.6 |  |  |  |  |  |  |  |  |  |  |  |  |  |
| 38114724 | 2024 | Retrospective | France | 109,693 | 26 | 41.61 | 100 |  | 21.17 |  |  | 83.22 |  |  |  |  | 17.33 | 1.74 | 11.81 |  |  |
| 34374759 | 2022 | Prospective | Spain  France | 1,479 | 23.3 | 35.6 | 100 | 53.4 | 24.1 | 7 | 29.2 | 72 | 36.9 |  | 43.5 |  | 13.5 |  | 10.5 | 9.1 |  |
| 33607944 | 2021 | Retrospective | USA | 729 | 50.1 |  | 100 | 80.2 | 32.4 | 16 | 100 | 76 | 40.1 |  | 71.3 |  | 17.6 |  | 28.6 | 20 | 48 |
| 25962838 | 2015 | Prospective | Spain | 948 | 20.25 | 31.75 | 100 | 81.33 | 31.12 | 6.96 |  | 73.8 | 43.25 |  | 54.85 |  | 14.98 | 1.16 | 21.1 | 16.67 |  |
| 28111055 | 2017 | Prospective | Multinational | 7,225 | 23.4 | 21.3 | 100 | 72.5 | 22.4 | 4.42 |  | 71.5 | 40 |  | 43.4 |  | 8.48 | 2.05 |  | 11.3 |  |
| 33637082 | 2021 | Retrospective | USA | 116,049 |  | 42.43 | 100 | 90.5 | 100 |  | 0 | 72.17 | 40.55 | 12.14 | 81.18 |  |  | 6.83 | 23 | 26.46 | 52.87 |
| 28411112 | 2017 | Prospective | Spain | 6,516 | 29.9 | 57.77 | 48.59 | 84.61 | 42.08 | 9.09 | 28.24 | 80.6 |  |  | 44.05 |  |  |  | 25.05 | 25.68 |  |
| 28990358 | 2018 | Prospective | Multinational | 5,387 | 55.86 |  | 44.6 | 65.29 | 40.52 | 14.3 |  | 69.31 |  |  |  | 34.23 |  | 8.63 | 28.9 | 19.79 |  |
| 33349492 | 2021 | Retrospective | USA | 26,189 | 32.7 |  | 40.7 | 81.7 | 46 | 46.6 |  | 76 | 51.7 |  |  |  |  |  |  | 18 |  |
| 32689614 | 2020 | Retrospective | Japan | 1,570 | 21.29 |  |  | 60.7 | 21.72 |  |  |  |  |  | 32.17 |  | 24.14 |  |  |  |  |
| 32397931 | 2020 | Prospective | France | 1,048 | 15.36 | 8.78 | 30.06 | 73.09 | 20.8 | 7.16 |  | 76.27 |  |  | 37.98 |  | 21.85 |  |  |  |  |
| 32829328 | 2020 | Retrospective | USA | 27,739 |  |  | 19.9 | 69.8 | 27.9 |  |  | 68.89 | 48.2 | 23.3 | 38.2 | 8.6 | 3.4 |  |  |  |  |
| 31760390 | 2019 | Retrospective | Japan | 1,012 | 12.25 |  | 16.8 | 66.8 | 27.96 |  |  | 72.37 |  | 39.92 | 16.9 |  | 16.6 |  |  |  |  |
| 27164672 | 2016 | Retrospective | Finland | 970 | 4.6 | 4.6 | 3.9 | 39.6 | 6 | 1.8 |  |  | 37.2 | 44.8 | 60.2 | 10.9 |  |  |  |  |  |
| 30371219 | 2018 | Prospective | Australia | 3,944 | 33 | 11 | 15 | 71 | 30 | 9 |  | 74.74 | 34 |  | 58 |  | 11 | 2 | 12 | 16 |  |
| 35301107 | 2022 | Retrospective | Multinational | 1,619 | 58.17 | 100 | 47.22 | 77.3 | 46.41 | 9.96 | 17.75 |  |  |  |  |  |  |  |  | 28.39 |  |
| 28741859 | 2017 | Retrospective | USA | 102,746 |  | 100 |  |  |  |  |  | 72.88 |  |  |  |  |  |  |  |  |  |
| 27995602 | 2017 | Prospective | Poland | 765 | 54 | 100 | 47 | 72 | 35 |  | 13 | 69 | 33 |  |  |  | 11 |  | 28 | 16 |  |
| 33243819 | 2020 | Retrospective | Canada | 352,329 |  | 100 | 12.43 | 77.68 | 29.38 | 6.03 | 4.72 | 73.89 | 50 |  |  |  |  | 1.55 | 9.06 | 34.52 |  |
| 36639191 | 2023 | Retrospective | Denmark | 27,947 |  | 100 | 0 | 41.9 | 20.5 | 12.2 | 9.2 | 68.5 | 33.6 |  | 20.2 |  |  |  | 7.4 | 12.8 |  |
| 31587602 | 2019 | Prospective | Italy | 2,791 | 44 | 100 | 29 | 59 | 30 |  |  | 69 | 30 |  | 47 |  |  |  | 38 | 17 |  |
| 26918404 | 2016 | Retrospective | USA | 2,581,892 |  | 100 |  |  | 51.7 |  |  | 80.4 | 56.9 |  |  |  |  | 2.4 | 38.2 |  |  |
| 38533944 | 2024 | Retrospective | Multinational | 6,765 | 34.01 |  | 27.2 | 82.99 | 29.52 | 16.04 | 100 | 81.68 |  | 17.29 |  |  |  |  | 100 |  |  |
| 33753041 | 2021 | Prospective | Japan | 2,588 |  |  | 21.21 | 79.02 | 21.45 | 14.57 | 100 |  |  |  | 43.04 |  | 11.63 | 2.94 |  | 23.8 |  |
| 29116212 | 2017 | Prospective | Japan | 3,815 | 30 | 42 | 22 | 70 | 24 | 7.4 | 100 | 77.8 | 62 |  | 35 |  | 13 |  |  | 10 | 55 |
| 28684438 | 2017 | Prospective | Germany | 1,378 |  |  | 24.38 |  | 34.18 | 20.97 | 100 | 81.78 | 57.6 |  |  |  | 7.91 |  | 61.03 | 23.88 |  |
| 27491609 | 2016 | Prospective | Multinational | 1,019 |  |  | 19.6 | 81.7 | 26.1 |  | 100 | 82.5 | 100 |  |  |  | 7.5 |  | 30.8 | 18.5 |  |
| 32912457 | 2020 | Retrospective | USA | 134,717 | 80.5 | 79.7 | 43.6 | 93.9 | 41.4 | 36.4 | 100 | 81.7 | 47.3 | 27.9 |  | 24.3 | 17 | 3.8 | 29 | 38.9 | 58.5 |
| 32493659 | 2022 | Retrospective | Canada | 45,883 | 100 | 19.4 | 4.4 | 88.29 | 48.54 | 12.08 | 3.06 | 66.03 |  |  | 74.92 | 42.83 |  | 0.69 |  | 27.74 |  |
| 32762879 | 2020 | Retrospective | Korea | 10,440 | 100 | 59.89 |  |  | 77.4 | 37 |  | 65.38 |  |  |  |  |  | 48.37 | 14.53 | 66.88 |  |
| 32466887 | 2020 | Prospective | USA | 9,623 | 100 |  | 8.75 | 91.68 | 48.26 | 9.84 |  | 66.2 |  |  | 90.67 |  |  |  |  |  |  |
| 37722886 | 2024 | Prospective | Japan | 12,291 | 100 | 21.66 | 9.1 | 82.7 | 38.1 | 9.05 |  | 69.3 |  |  |  |  | 12.52 |  |  | 3.96 |  |
| 32223396 | 2020 | Prospective | Multinational | 2,397 | 100 |  |  | 86.6 | 44.9 |  |  | 71.32 | 34.3 |  | 73.9 |  | 14.31 |  | 49.7 |  | 25.6 |
| 26150477 | 2015 | Retrospective | USA | 15,008 | 100 | 15.26 |  | 65.38 | 27.6 | 8.56 | 0.22 | 62.2 |  | 52.97 |  |  |  | 0.47 | 1.77 | 5.48 |  |
| 25882502 | 2015 | Retrospective | USA | 5,350 | 100 | 14.67 |  | 70.17 | 26.64 | 10.54 |  | 66.78 |  |  | 70.24 |  |  |  |  | 11.51 |  |
| 28648964 | 2017 | Retrospective | Belgium | 578 |  | 2.6 | 6.2 | 49.3 | 100 | 5.7 |  | 67 | 4.8 |  | 33.4 |  |  | 6.7 | 19.9 | 4.8 |  |
| 27997863 | 2017 | Prospective | China | 1,239 | 4.84 | 2.5 |  |  | 100 | 4.36 |  | 58.61 |  | 48.4 |  |  |  |  | 14.12 |  |  |
| 31233278 | 2020 | Prospective | Germany | 828 | 78.12 |  | 44.17 | 77.99 | 31.29 | 13.71 | 100 | 75.31 |  | 16.13 | 51.35 |  | 10.18 |  | 42.35 | 23.16 |  |

Abbreviations: CAD, coronary artery disease; HF, heart failure; AF, atrial fibrillation; HTN, hypertension; DM, diabetes mellitus; PAD, peripheral artery disease; VHD, valve heart disease; CKD, chronic kidney disease; COPD, chronic obstructive pulmonary disease.

###### Table S2. The baseline characteristics of total population across the world geographic regions with consideration of all studies.

| **Variable** | **Europe and Northern America** | **Eastern and South-Eastern Asia** | **Oceania** | **Northern Africa and Western Asia** | **Central and Southern Asia** | **Latin America and the Caribbean** | **Sub-Saharan Africa** | **Multiregional** | **Total** |
| --- | --- | --- | --- | --- | --- | --- | --- | --- | --- |
| **N of cohorts** | 482 | 138 | 21 | 16 | 3 | 4 | 1 | 21 | 686 |
| **Sample size** | 151,604651 | 285,45384 | 368,431 | 137,143 | 9770 | 6131 | 566 | 203,455 | 180,875,531 |
| **Age, years** | 59.98 (7.95) | 62.29 (4.18) | 68.46 (4.08) | 65.46 (6.69) | 54.36 (0.39) | 65.36 (9.16) | 65 (15) | 66.97 (3.61) | 60.39 (7.5) |
| **Female, %** | 46.89 | 36.46 | 34.46 | 38.31 | 17.1 | 57 | 52 | 29.97 | 45.96 |
| **Study design** |  |  |  |  |  |  |  |  |  |
| **Cross-sectional Study** | 7 (1.5%) |  | 2 (9.5%) |  | 1 (33.3%) |  |  | 1 (4.8%) | 11  1.6% |
| **Prospective Cohort** | 110 (22.8%) | 51 (37%) | 7 (33.3%) | 6 (37.5%) | 2 (66.7%) | 2 (50%) | 1 (100%) | 15 (71.4%) | 194  28.3% |
| **Retrospective Cohort** | 363 (75.3%) | 87 (63%) | 12 (57.1%) | 10 (62.5%) |  | 2 (50%) |  | 5 (23.8%) | 479  69.8% |
| **Case-Control Study** | 2 (0.4%) |  |  |  |  |  |  |  | 2  0.3% |
| **Inpatient/utpatient** |  |  |  |  |  |  |  |  |  |
| **inpatient** | 218 (45.2%) | 57 (41.3%) | 11 (52.4%) | 9 (56.2%) | 2 (66.7%) | 1 (25%) | 1 (100%) | 10 (47.6%) | 309  64.4% |
| **outpatient** | 42 (8.7%) | 7 (5.1%) | 4 (19%) | 1 (6.2%) |  | 2 (50%) |  |  | 56  11.7% |
| **both** | 70 (14.5%) | 29 (21%) | 1 (4.8%) | 1 (6.2%) |  | 1 (25%) |  | 7 (33.3%) | 109  22.7% |
| **Other** | 5 (1%) | 1 (0.7%) |  |  |  |  |  |  | 6  1.3% |
| **Multicenter/single-center** |  |  |  |  |  |  |  |  |  |
| **multicenter** | 423 (87.8%) | 130 (94.2%) | 19 (90.5%) | 15 (93.8%) | 3 (100%) | 3 (75%) |  | 21 (100%) | 614  89.5% |
| **single-center** | 59 (12.2%) | 8 (5.8%) | 2 (9.5%) | 1 (6.2%) |  | 1 (25%) | 1 (100%) |  | 72  10.5% |
| **Risk factors/ comorbidities** |  |  |  |  |  |  |  |  |  |
| **Smoking** | 23.73 | 30.72 | 39.76 | 46.86 |  | 9.36 | 3 | 43.04 | 23.83 |
| **Active smoking** | 20.61 | 21.67 | 25.73 | 20.11 | 51.73 | 34.8 |  | 14.82 | 21.07 |
| **Past smoking** | 30.03 | 17.56 | 41.2 | 33.99 | 6.33 |  |  | 41.6 | 24.25 |
| **Dyslipidemia** | 58.25 | 52.76 | 37.56 | 58.53 | 9.76 | 30.14 |  | 45.99 | 57.26 |
| **Obesity** | 41.2 | 0.59 | 6.58 | 37.83 | 20.76 |  |  | 29.22 | 40.11 |
| **Alcoholism** | 4 | 24.91 | 3.59 | 2.4 | 9.01 | 5.53 | 9 | 9.43 | 8.85 |
| **History of stroke** | 6.06 | 11.8 | 2.21 | 10.55 | 1.46 | 33.6 | 12 | 9.93 | 6.93 |
| **Cerebrovascular disease** | 8.84 | 14.08 | 7.57 |  |  |  |  | 14.05 | 10.02 |
| **Liver diseases** | 3.22 | 19.03 | 4.73 | 0.83 |  | 1.07 |  | 3.25 | 6.76 |
| **Cirrhosis** | 1.55 | 2.13 | 0.2 | 0.2 |  |  |  |  | 1.79 |
| **Chronic kidney disease** | 22.27 | 9.77 | 10.37 | 16.64 | 4.9 | 32.21 |  | 9.77 | 21.61 |
| **End-stage kidney disease** | 3.38 | 1.34 |  | 15.44 | 0 |  |  | 14 | 2.55 |
| **Bronchial asthma** | 13.58 | 13.57 | 15.39 | 3.73 |  | 1.34 |  |  | 13.58 |
| **Chronic obstructive kidney disease** | 11.32 | 17.73 | 8.37 | 11.46 |  | 10.34 |  | 9.68 | 11.48 |
| **Anemia** | 21.43 | 12.28 | 11.87 | 33.68 |  | 13.4 |  | 19.49 | 20.28 |
| **Hypothyroidism** | 13.67 | 9.58 | 1 |  |  |  |  | 9 | 13.37 |
| **Hyperthyroidism** | 1.32 | 3.52 | 7.42 |  |  |  |  | 3.17 | 2.43 |

###### Table S3. Risk of bias assessment.

| **PMID** | **Was the sample representative of the target population?** | **Were study participants recruited in an appropriate way?** | **Was the sample size adequate?** | **Were the study subjects and the setting described in detail?** | **Was the data analysis conducted with sufficient coverage of the identified sample?** | **Were objective, standard criteria used for the measurement of the condition?** | **Was the condition measured reliably?** | **Was there appropriate statistical analysis?** | **Are all important confounding factors/ subgroups/differences identified and accounted for?** | **Were subpopulations identified using objective criteria?** |
| --- | --- | --- | --- | --- | --- | --- | --- | --- | --- | --- |
| 38114724 | Yes | Yes | Yes | Yes | Unclear | Unclear | Unclear | NA | NA | NA |
| 28280146 | Yes | Yes | Yes | Yes | Unclear | Unclear | Unclear | NA | NA | NA |
| 31587602 | Yes | Yes | Yes | Yes | Unclear | Yes | Unclear | NA | NA | NA |
| 33214605 | Yes | Yes | Yes | Yes | Yes | Yes | Unclear | NA | NA | NA |
| 33607944 | Yes | Yes | Yes | Yes | Unclear | Unclear | Unclear | NA | NA | NA |
| 33637082 | Yes | Yes | Yes | Yes | Yes | Unclear | No | NA | NA | NA |
| 28927174 | Yes | Yes | Yes | Yes | Yes | Unclear | Unclear | NA | NA | NA |
| 28875506 | Yes | Yes | Yes | Yes | Yes | Unclear | Unclear | NA | NA | NA |
| 37201612 | Yes | Yes | Yes | Yes | Unclear | Unclear | Unclear | NA | NA | NA |
| 30883428 | Yes | Yes | Yes | Yes | Unclear | Yes | Unclear | NA | NA | NA |
| 32561143 | Yes | Yes | Yes | Yes | Unclear | Unclear | Unclear | NA | NA | NA |
| 31775530 | Yes | Yes | Yes | Yes | Unclear | Unclear | Unclear | NA | NA | NA |
| 35965069 | Yes | Yes | Yes | Yes | Yes | Unclear | Unclear | NA | NA | NA |
| 27865662 | Yes | Yes | Yes | Yes | Unclear | Unclear | Unclear | NA | NA | NA |
| 28331926 | Yes | Yes | Yes | Yes | Unclear | Unclear | Unclear | NA | NA | NA |
| 38657869 | Yes | Yes | Yes | Yes | Yes | Unclear | Unclear | NA | NA | NA |
| 34390242 | Yes | Yes | Yes | Yes | Unclear | Unclear | Unclear | NA | NA | NA |
| 34244745 | Yes | Yes | Yes | Yes | Yes | Yes | Unclear | NA | NA | NA |
| 33963402 | Yes | Yes | Yes | Yes | Yes | Unclear | Unclear | NA | NA | NA |
| 28951401 | Yes | Yes | Yes | Yes | Yes | Unclear | Unclear | NA | NA | NA |
| 32506417 | Yes | Yes | Yes | Yes | Unclear | Unclear | Unclear | NA | NA | NA |
| 32386073 | Yes | Yes | Yes | Yes | Yes | Yes | Unclear | NA | NA | NA |
| 32079476 | Yes | Yes | Yes | Yes | Yes | Unclear | Unclear | NA | NA | NA |
| 31983727 | Yes | Yes | Yes | Yes | Yes | Yes | Unclear | NA | NA | NA |
| 31976927 | Yes | Yes | Yes | Yes | Unclear | Unclear | Unclear | NA | NA | NA |
| 34374759 | Yes | Yes | Yes | Yes | Unclear | Unclear | Unclear | NA | NA | NA |
| 25962838 | Yes | Yes | Yes | Yes | Unclear | Unclear | Unclear | NA | NA | NA |
| 28111055 | Yes | Yes | Yes | Yes | Unclear | Unclear | Unclear | NA | NA | NA |
| 28411112 | Yes | Yes | Yes | Yes | Yes | Unclear | Unclear | NA | NA | NA |
| 28990358 | Yes | Yes | Yes | Yes | Yes | Unclear | Unclear | NA | NA | NA |
| 33349492 | Yes | Yes | Yes | Yes | Unclear | Unclear | Unclear | NA | NA | NA |
| 32689614 | Yes | Yes | Yes | Yes | Unclear | Unclear | Unclear | NA | NA | NA |
| 32397931 | Yes | Yes | Yes | Yes | Unclear | Unclear | Yes | NA | NA | NA |
| 32829328 | Yes | Yes | Yes | Yes | Unclear | Unclear | Unclear | NA | NA | NA |
| 31760390 | Yes | Yes | Yes | Yes | Yes | Yes | Unclear | NA | NA | NA |
| 27164672 | Yes | Yes | Yes | Yes | Unclear | Unclear | Unclear | NA | NA | NA |
| 30371219 | Yes | Yes | Yes | Yes | Unclear | Unclear | Unclear | NA | NA | NA |
| 35301107 | Yes | Yes | Yes | Yes | Yes | Unclear | Unclear | NA | NA | NA |
| 28741859 | Yes | Yes | Yes | Yes | Yes | Unclear | Unclear | NA | NA | NA |
| 27995602 | Yes | Yes | Yes | Yes | Unclear | Unclear | Unclear | NA | NA | NA |
| 33243819 | Yes | Yes | Yes | Yes | Unclear | Unclear | Unclear | NA | NA | NA |
| 36639191 | Yes | Yes | Yes | Yes | Yes | Unclear | Unclear | NA | NA | NA |
| 26918404 | Yes | Yes | Yes | Yes | Unclear | Unclear | Unclear | NA | NA | NA |
| 38533944 | Yes | Yes | Yes | Yes | Unclear | Unclear | Unclear | NA | NA | NA |
| 33753041 | Yes | Yes | Yes | Yes | Unclear | Unclear | Unclear | NA | NA | NA |
| 29116212 | Yes | Yes | Yes | Yes | Yes | Unclear | Yes | NA | NA | NA |
| 28684438 | Yes | Yes | Yes | Yes | Yes | Unclear | Unclear | NA | NA | NA |
| 27491609 | Yes | Yes | Yes | Yes | Unclear | Unclear | Yes | NA | NA | NA |
| 32912457 | Yes | Yes | Yes | Yes | Unclear | Unclear | Unclear | NA | NA | NA |
| 32493659 | Yes | Yes | Yes | Yes | Unclear | Yes | Unclear | NA | NA | NA |
| 32762879 | Yes | Yes | Yes | Yes | Yes | Unclear | Unclear | NA | NA | NA |
| 32466887 | Yes | Yes | Yes | Yes | Unclear | Unclear | Unclear | NA | NA | NA |
| 37722886 | Yes | Yes | Yes | Yes | Yes | Unclear | Unclear | NA | NA | NA |
| 32223396 | Yes | Yes | Yes | Yes | Unclear | Unclear | Unclear | NA | NA | NA |
| 26150477 | Yes | Yes | Yes | Yes | Yes | Yes | Unclear | NA | NA | NA |
| 25882502 | Yes | Yes | Yes | Yes | Unclear | Unclear | Unclear | NA | NA | NA |
| 28648964 | Yes | Yes | Yes | Yes | Yes | Unclear | Yes | NA | NA | NA |
| 27997863 | Yes | Yes | Yes | Yes | Unclear | Unclear | Unclear | NA | NA | NA |
| 31233278 | Yes | Yes | Yes | Yes | Unclear | Unclear | Unclear | NA | NA | NA |

NA, not applicable.

###### Table S4. Baseline characteristics studies included in meta-analysis of prevalence of active cancer categorized by cardiovascular diseases.

| **Variable** | **CAD** | **ACS** | **AMI** | **PCI** | **AF** | **NVAF** | **CHF** | **AHF** | **any stroke** | **IS** | **VHD** | **DM** |
| --- | --- | --- | --- | --- | --- | --- | --- | --- | --- | --- | --- | --- |
| **N of cohorts** | 16 | 9 | 5 | 8 | 18 | 8 | 7 | 3 | 6 | 5 | 8 | 3 |
| **Sample size** | 819,468 | 718,476 | 669,620 | 47,911 | 642,836 | 279,408 | 3,070,089 | 38,092 | 84,810 | 57,071 | 151,839 | 117,866 |
| **Retrospective Cohort, %** | 94.5 | 97.1 | 99.2 | 45.8 | 78.1 | 87.9 | 99.9 | 68.8 | 36.9 | 6.2 | 93.7 | 98.9 |
| **Prospective Cohort, %** | 5.5 | 2.9 | 0.8 | 54.2 | 15.5 | 12.1 | 0.1 | 31.2 | 63.1 | 93.8 | 6.3 | 1.1 |
| **Cross-sectional, %** |  |  |  |  | 6.5 |  |  |  |  |  |  |  |
| **Multicenter, %** | 96.0 | 99.6 | 99.8 | 31.8 | 99.5 | 99.2 | 99.9 | 100.0 | 97.0 | 95.5 | 100.0 | 98.9 |
| **Single-center, %** | 4.0 | 0.4 | 0.2 | 68.2 | 0.5 | 0.8 | 0.1 |  | 3.0 | 4.5 |  | 1.1 |
| **Inpatient, %** | 100.0 | 100.0 | 100.0 | 100.0 |  |  | 21.5 | 85.9 | 100.0 | 100.0 | 100.0 |  |
| **Outpatient, %** |  |  |  |  | 9.9 | 4.7 | 72.7 |  |  |  |  | 0.5 |
| **Inpatient/Outpatient, %** |  |  |  |  | 90.1 | 95.3 | 5.7 | 14.1 |  |  |  | 99.5 |
| **Mean age (SD), years** | 67.9(1.7) | 68.2(1.4) | 68.4(1.3) | 66.1(3.0) | 75.9(5.2) | 77.8(5.2) | 79.3(2.6) | 75.8(3.1) | 76.2(5.3) | 80(1.2) | 81.5(0.9) | 72(1.4) |
| **Female, %** | 26.5 | 26.4 | 25.6 | 30.8 | 42.3 | 40.9 | 55.8 | 51.7 | 47.8 | 37.2 | 48.1 | 40.1 |
| **HTN, %** | 61 | 58 | 57.8 | 76.8 | 73.2 | 86.6 | 74.9 | 79.9 | 77.4 | 81.1 | 92.3 | 90.3 |
| **DM, %** | 32.2 | 30.3 | 30.4 | 35.1 | 34.7 | 54.6 | 48.7 | 44.6 | 24.9 | 23.4 | 39.8 | 100 |
| **Smoking, %** | 53.6 | 69.9 |  | 53.6 | 12.1 | 12.1 |  |  | 24.6 | 42.3 | 27.3 | 12.5 |
| **Dyslipidemia, %** | 62.6 | 58.6 | 65.4 | 79.9 | 70.2 | 72.1 | 22.6 | 44 | 37.9 | 36 | 42.6 | 80.9 |
| **Obesity, %** | 42.8 |  |  |  | 2.5 |  |  | 34.2 | 4.7 | 2.7 | 24.3 |  |
| **Alcoholism, %** | 1.4 |  |  |  | 1.9 | 0.8 | 1.1 |  | 5.1 | 14.1 | 2 | 0.8 |
| **History of stroke, %** | 1.7 | 1.4 | 1.4 | 12 | 14.7 | 17.7 | 11 |  | 14 | 19.3 | 16.6 |  |
| **Liver disease, %** | 1.4 | 0.4 | 0.4 | 0.5 | 2.3 | 4.3 | 2.3 | 8.6 | 1.3 | 1.3 | 3.8 | 6.8 |
| **CKD, %** | 10.5 | 10.5 | 10.6 | 7.9 | 12.6 | 17.9 | 34.4 | 26.8 | 6.2 | 6.2 | 32.7 | 22.9 |
| **COPD, %** | 8.6 | 6.5 | 6.7 | 5.9 | 14.5 | 25 | 32.8 | 19.6 | 6.9 | 6.9 | 37.4 | 26.4 |
| **Anemia, %** | 25.6 |  |  | 25.6 | 51.7 | 52.9 |  |  |  |  | 58.3 | 52.9 |
| **Hypothyroidism, %** | 1.8 |  |  |  | 10 |  | 2.6 |  |  |  | 25.8 |  |
| **Hyperthyroidism, %** | 0.5 | 0.5 |  | 0.5 | 3.7 |  | 1.1 |  |  |  |  |  |

CAD – Coronary artery disease; ACS – Acute coronary syndrome; AMI – acute myocardial infarction; PCI – Percutaneous coronary intervention; AF, atrial fibrillation; NVAF, non-valvular atrial fibrillation; CHF, chronic heart failure; AHF, acute heart failure; IS, ischemic stroke; VHD – Valvular heart disease; DM, diabetes mellitus; HTN, hypertension; SD, standard deviation; CKD – Chronic kidney disease; COPD – Chronic obstructive pulmonary disease.

###### Table S5. The results of meta-regression analyses.

| **Population** | **Outcome** | **Variable** | **N of studies** | **Beta coefficient** | **P value** |
| --- | --- | --- | --- | --- | --- |
| Coronary artery disease | active cancer | peripheral artery disease, % | 13 | 0.06 | 0.00 |
|  |  | chronic obstructive pulmonary disease, % | 12 | 0.03 | 0.02 |
|  | history of cancer | acute coronary syndrome, % | 22 | -0.03 | 0.00 |
|  |  | history of percutaneous coronary intervention, % | 23 | 0.03 | 0.00 |
|  |  | heart failure, % | 22 | 0.04 | 0.00 |
|  |  | peripheral artery disease, % | 21 | 0.06 | 0.00 |
|  |  | chronic kidney disease, % | 19 | 0.04 | 0.00 |
|  |  | chronic obstructive pulmonary disease, % | 20 | 0.04 | 0.00 |
|  | unspecified cancer | st-elevation myocardial infarction, % | 36 | -0.02 | 0.00 |
|  |  | history of percutaneous coronary intervention, % | 28 | 0.02 | 0.01 |
|  |  | hypertension, % | 77 | 0.02 | 0.00 |
|  |  | diabetes mellitus, % | 84 | 0.02 | 0.02 |
|  |  | peripheral artery disease, % | 65 | 0.03 | 0.03 |
|  |  | age | 78 | 0.05 | 0.01 |
|  |  | smoking active, % | 32 | -0.04 | 0.01 |
|  |  | past smoking, % | 12 | 0.09 | 0.00 |
|  |  | dyslipidemia, % | 50 | 0.01 | 0.04 |
|  |  | history of stroke, % | 41 | 0.06 | 0.01 |
|  |  | both inpatient and outpatients | 73 | 1.00 | 0.02 |
| All types of acute coronary syndrome | history of cancer | chronic obstructive pulmonary disease, % | 11 | 0.15 | 0.02 |
|  | metastasis | prospective design | 14 | -2.01 | 0.01 |
|  |  | heart failure, % | 13 | -0.06 | 0.04 |
|  |  | liver diseases, % | 10 | -0.54 | 0.04 |
|  | unspecified cancer | pr_ami_total | 43 | 0.03 | 0.00 |
|  |  | hypertension, % | 59 | 0.02 | 0.01 |
|  |  | diabetes mellitus, % | 64 | 0.02 | 0.04 |
|  |  | dyslipidemia, % | 42 | 0.02 | 0.00 |
|  |  | history of stroke, % | 29 | 0.09 | 0.03 |
| ST-elevation myocardial infarction | unspecified cancer | age | 11 | 0.21 | 0.01 |
| Acute myocardial infarction | unspecified cancer | prospective design |  |  |  |
|  |  | dyslipidemia, % | 33 | 0.01 | 0.04 |
| Percutaneous coronary intervention | history of cancer | history of percutaneous coronary intervention, % | 10 | 0.04 | 0.00 |
|  |  | hypertension, % | 11 | 0.03 | 0.02 |
|  | unspecified cancer | history of myocardial infarction, % | 22 | 0.03 | 0.01 |
|  |  | history of percutaneous coronary intervention, % | 16 | 0.02 | 0.00 |
|  |  | prospective design | 32 | 0.85 | 0.002 |
| Atrial fibrillation | active cancer | heart failure, % | 16 | 0.03 | 0.04 |
|  |  | hypertension, % | 16 | 0.02 | 0.01 |
|  |  | history of stroke, % | 15 | 0.10 | 0.00 |
|  | history of cancer | peripheral artery disease, % | 11 | 0.02 | 0.02 |
|  | unspecified cancer | prospective design | 74 | -0.46 | 0.05 |
|  |  | non-valvular atrial fibrillation, % | 33 | 0.16 | 0.00 |
|  |  | age | 65 | 0.06 | 0.02 |
|  |  | male_pr | 14 | 0.03 | 0.01 |
|  |  | dyslipidemia, % | 30 | 0.02 | 0.01 |
|  |  | cerebrovascular disease, % | 11 | 0.04 | 0.03 |
| Any chronic heart failure | unspecified cancer | peripheral artery disease, % | 31 | 0.05 | 0.02 |
|  |  | age | 52 | 0.06 | 0.00 |
|  |  | female_pr | 36 | 0.03 | 0.04 |
|  |  | dyslipidemia, % | 22 | 0.02 | 0.01 |
|  |  | history of stroke, % | 34 | 0.07 | 0.01 |
|  |  | cerebrovascular disease, % | 12 | 0.03 | 0.02 |
|  |  | chronic obstructive pulmonary disease, % | 41 | 0.02 | 0.03 |
|  |  | prospective design, % | 56 | -0.77 | 0.00 |
| Chronic heart failure with reduced ejection fraction | unspecified cancer | atrial fibrillation, % | 10 | 0.04 | 0.00 |
|  |  | age | 11 | 0.05 | 0.04 |
| Acute heart failure | unspecified cancer | atrial fibrillation, % | 23 | 0.02 | 0.02 |
|  |  | hypertension, % | 24 | 0.03 | 0.00 |
|  |  | peripheral artery disease, % | 14 | 0.07 | 0.03 |
|  |  | valve heart disease, % | 10 | 0.05 | 0.01 |
|  |  | age | 22 | 0.07 | 0.00 |
|  |  | female_pr | 11 | 0.05 | 0.00 |
|  |  | chronic obstructive pulmonary disease, % | 19 | 0.04 | 0.04 |
| Any stroke | unspecified cancer | heart failure, % | 28 | 0.04 | 0.03 |
|  |  | hemorrhagic stroke, % | 12 | -0.05 | 0.01 |
|  |  | smoking, % | 13 | 0.03 | 0.02 |
|  |  | chronic kidney disease, % | 17 | 0.05 | 0.00 |
| Ischemic stroke | unspecified cancer | heart failure, % | 17 | 0.07 | 0.00 |
|  |  | peripheral artery disease, % | 14 | 0.15 | 0.02 |
|  |  | smoking, % | 10 | 0.04 | 0.00 |
| Diabetes mellitus | history of cancer | age | 11 | 0.15 | 0.00 |
|  | unspecified cancer | atrial fibrillation, % | 17 | 0.07 | 0.00 |
|  |  | hypertension, % | 46 | 0.02 | 0.03 |
|  |  | age | 55 | 0.05 | 0.01 |
|  |  | chronic kidney disease, % | 40 | 0.02 | 0.01 |
| Hypertension | unspecified cancer | age | 13 | 0.10 | 0.00 |
|  |  | history of stroke, % | 10 | 0.19 | 0.01 |
|  |  | chronic kidney disease, % | 10 | 0.09 | 0.02 |

###### Table S6. The age-standardized prevalence ratios of cancer according to meta-regression analyses.

| **Age, years** | **Prevalence ratios, % [95% confidence intervals]** | | | | | | | |
| --- | --- | --- | --- | --- | --- | --- | --- | --- |
|  | **Coronary artery disease** | **Atrial fibrillation** | | **Heart failure** | **Any stroke** | **Valve heart disease** | **Type 2 diabetes mellitus** | **Hypertension** |
|  | Any cancer | Active cancer | Any cancer | Any cancer | Any cancer | Any cancer | Any cancer | Any cancer |
| **20** | 0.49 [0.08-2.98] | 0.05 [0.00-2.41] | 0.26 [0.02-3.37] | 0.53 [0.10-2.76] | 2.43 [0.50-10.93] | 3.38 [0.19-38.4] | 0.82 [0.20-3.30] | 0.07 [0.01-0.62] |
| **30** | 0.82 [0.19-3.41] | 0.11 [0.00-2.70] | 0.53 [0.06-4.29] | 0.94 [0.24-3.60] | 3.07 [0.86-10.36] | 3.9 [0.38-30.9] | 1.39 [0.46-4.07] | 0.19 [0.03-1.07] |
| **40** | 1.37 [0.47-3.90] | 0.26 [0.02-3.02] | 1.1 [0.21-5.44] | 1.67 [0.58-4.69] | 3.86 [1.45-9.86] | 4.67 [0.74-24.3] | 2.34 [1.08-5.01] | 0.54 [0.16-1.86] |
| **50** | 2.29 [1.16-4.48] | 0.58 [0.10-3.39] | 2.25 [0.71-6.90] | 2.95 [1.40-6.11] | 4.85 [2.43-9.46] | 5.48 [1.42-18.8] | 3.92 [2.45-6.22] | 1.53 [0.69-3.33] |
| **60** | 3.8 [2.75-5.24] | 1.3 [0.43-3.84] | 4.56 [2.32-8.76] | 5.16 [3.30-8.00] | 6.09 [3.93-9.30] | 6.42 [2.69-14.5] | 6.5 [5.24-8.04] | 4.23 [2.60-6.82] |
| **70** | 6.25 [5.13-7.59] | 2.9 [1.80-4.63] | 9.02 [7.02-11.51] | 8.89 [7.18-10.95] | 7.6 [5.72-10.05] | 7.51 [4.8-11.5] | 10.6 [8.26-13.50] | 11.16 [6.23-19.19] |
| **80** | 10.11 [6.25-15.94] | 6.34 [3.97-9.99] | 17.07 [12.61-22.69] | 14.87 [11.60-18.87] | 9.47 [6.53-13.53] | 8.77 [6.2-12.26] | 16.8 [10.42-25.96] | 26.33 [11.18-50.37] |

Ta**ble S7. The pooled prevalence rates of cancer obtained during the main and sensitivity analyses and subgroup analyses across different geographical regions.**

| **Population** | **Outcome** | **Sample size** | **Prevalence rates in main analyses** | | **Prevalence in Europe and Northern America** | | **Prevalence in Eastern and South-Eastern Asia** | | **P value for a regional difference** |
| --- | --- | --- | --- | --- | --- | --- | --- | --- | --- |
|  |  |  | **N of studies** | **Prevalence [CI]** | **N of studies** | **Prevalence [CI]** | **N of studies** | **Prevalence [CI]** |  |
| Coronary artery disease | active cancer | 819468 | 16 | 4.61 [2.83, 6.97] | 10 | 5.02 [2.93, 7.52] | 3 | 6.35 [2.29, 11.07] | 0.6 |
|  | blood cancer | 1171292 | 6 | 0.62 [0.39, 0.90] | 5 | 0.69 [0.48, 0.96] |  |  |  |
|  | past cancer | 904033 | 31 | 9.72 [6.97, 14.17] | 20 | 8.98 [7.64, 10.19] | 7 | 14.13 [3.96, 31.38] | 0.46 |
|  | metastases | 1978970 | 17 | 1.39 [0.81, 2.23] | 12 | 1.55 [0.80, 2.79] | 3 | 0.63 [0.16, 1.24] | 0.11 |
|  | solid cancer | 1049613 | 5 | 4.39 [2.04, 7.72] | 3 | 4.35 [1.37, 7.80] |  |  |  |
|  | any cancer | 6997586 | 88 | 7.04 [6.05, 8.03] | 65 | 7.28 [6.21, 8.50] | 13 | 6.80 [4.76, 8.85] | 0.69 |
| Any acute coronary syndromes | active cancer | 718476 | 9 | 3.12 [1.91, 5.13] | 6 | 3.52 [1.64, 6.29] |  |  |  |
|  | blood cancer | 1149700 | 4 | 0.76 [0.56, 1.03] | 4 | 0.77 [0.56, 1.06] |  |  |  |
|  | past cancer | 1068888 | 21 | 8.02 [5.92, 9.96] | 16 | 9.14 [7.00, 11.07] |  |  |  |
|  | metastases | 1998646 | 14 | 1.64 [0.99, 2.62] | 12 | 1.66 [0.90, 2.79] |  |  |  |
|  | solid cancer | 1043029 | 4 | 4.34 [1.54, 8.65] | 3 | 4.35 [1.37, 7.80] |  |  |  |
|  | any cancer | 5664501 | 67 | 6.38 [5.49, 7.19] | 50 | 6.41 [5.46, 7.42] | 11 | 6.60 [4.73, 8.55] | 0.86 |
| Any acute myocardial infarction | active cancer | 669620 | 5 | 2.41 [1.56, 3.03] | 3 | 1.95 [1.23, 2.83] |  |  |  |
|  | blood cancer | 1149700 | 4 | 0.76 [0.56, 1.03] | 4 | 0.77 [0.56, 1.06] |  |  |  |
|  | past cancer | 900893 | 14 | 7.85 [5.98, 9.65] | 12 | 8.70 [7.14, 10.27] |  |  |  |
|  | metastases | 1938021 | 11 | 1.69 [0.77, 3.04] | 10 | 1.79 [0.77, 3.22] |  |  |  |
|  | solid cancer | 1026258 | 3 | 4.34 [1.37, 7.77] | 3 | 4.35 [1.37, 7.80] |  |  |  |
|  | any cancer | 4450896 | 50 | 6.75 [5.78, 7.81] | 36 | 6.70 [5.51, 8.07] | 9 | 7.19 [5.04, 9.43] | 0.7 |
| ST-elevation myocardial infarction | past cancer | 118751 | 7 | 8.78 [6.10, 11.01] | 7 | 8.81 [5.95, 10.94] |  |  |  |
|  | any cancer | 341260 | 11 | 5.62 [3.66, 7.38] | 9 | 5.12 [3.36, 6.76] |  |  | 0.28 |
| non-ST-elevation myocardial infarction | any cancer | 795282 | 3 | 7.84 [4.59, 10.91] |  |  |  |  |  |
| Non-ST-elevation acute coronary syndromes | any cancer | 782209 | 7 | 6.27 [3.50, 9.23] | 5 | 5.51 [2.90, 8.24] |  |  |  |
| Stable coronary artery disease | any cancer | 401742 | 6 | 9.10 [5.12, 13.00] | 5 | 8.84 [4.79, 13.90] |  |  |  |
| History of myocardial infarction | any cancer | 100990 | 6 | 7.26 [3.78, 11.05] | 4 | 4.32 [2.62, 6.10] |  |  |  |
| Percutaneous coronary intervention | active cancer | 47911 | 8 | 4.53 [2.52, 7.39] | 6 | 5.52 [3.22, 9.01] |  |  |  |
|  | past cancer | 98618 | 12 | 13.93 [7.25, 24.76] | 8 | 9.51 [6.72, 11.56] | 3 | 25.27 [4.86, 46.32] | 0.14 |
|  | metastases | 44697 | 4 | 0.58 [0.19, 1.02] |  |  |  |  |  |
|  | any cancer | 1424959 | 32 | 7.34 [5.85, 8.97] | 19 | 7.52 [5.79, 9.96] | 10 | 6.91 [4.47, 9.43] | 0.71 |
| Coronary artery bypass surgery | past cancer | 83425 | 4 | 6.36 [2.83, 11.71] | 4 | 6.38 [2.40, 11.75] |  |  |  |
|  | metastases | 117276 | 3 | 0.94 [0.60, 1.26] |  |  |  |  |  |
|  | any cancer | 603657 | 14 | 6.85 [4.48, 9.24] | 11 | 6.84 [4.26, 10.11] |  |  |  |
| Atrial fibrillation | active cancer | 708399 | 19 | 5.55 [3.97, 7.01] | 14 | 5.30 [3.89, 6.75] | 3 | 8.88 [7.15, 10.94] | <0.001 |
|  | blood cancer | 428505 | 7 | 1.26 [0.88, 1.72] | 6 | 1.37 [0.96, 1.83] |  |  |  |
|  | past cancer | 2564550 | 17 | 15.33 [11.88, 19.97] | 12 | 12.97 [10.26, 16.35] |  |  |  |
|  | metastases | 827119 | 10 | 2.74 [1.68, 4.14] | 9 | 2.85 [1.58, 4.52] |  |  |  |
|  | solid cancer | 397570 | 4 | 8.43 [1.62, 15.24] | 3 | 11.05 [5.82, 15.60] |  |  |  |
|  | any cancer | 9982683 | 76 | 14.10 [12.20, 15.99] | 51 | 15.82 [13.90, 17.70] | 19 | 8.85 [6.37, 11.48] | <0.001 |
| Non-valvular atrial fibrillation | active cancer | 540494 | 10 | 5.87 [3.92, 7.74] | 7 | 5.20 [3.74, 6.90] |  |  |  |
|  | blood cancer | 9230 | 3 | 1.27 [0.84, 1.61] |  |  |  |  |  |
|  | past cancer | 1517472 | 7 | 14.31 [9.36, 19.94] | 4 | 14.30 [7.66, 22.67] |  |  |  |
|  | metastases | 238133 | 5 | 2.95 [1.57, 5.48] | 5 | 2.96 [1.51, 5.50] |  |  |  |
|  | solid cancer | 9230 | 3 | 11.06 [4.97, 15.60] |  |  |  |  |  |
|  | any cancer | 4944432 | 40 | 14.03 [12.19, 15.73] | 27 | 15.53 [13.73, 17.30] | 11 | 11.70 [8.22, 15.68] | 0.07 |
| Any chronic heart failure | active cancer | 3070089 | 7 | 4.43 [2.78, 6.38] | 7 | 4.45 [2.63, 6.24] |  |  |  |
|  | blood cancer | 623938 | 4 | 1.11 [0.39, 1.63] | 4 | 1.12 [0.39, 1.63] |  |  |  |
|  | past cancer | 191153 | 8 | 13.06 [8.42, 18.25] | 6 | 13.40 [7.27, 20.35] |  |  |  |
|  | metastases | 1149839 | 10 | 2.02 [1.47, 2.57] | 9 | 1.94 [1.31, 2.52] |  |  |  |
|  | solid cancer | 647979 | 6 | 9.46 [5.08, 12.59] | 4 | 6.28 [2.10, 8.34] |  |  |  |
|  | any cancer | 5369779 | 56 | 12.82 [10.60, 14.89] | 39 | 13.73 [11.18, 16.59] | 11 | 10.99 [7.29, 14.85] | 0.25 |
| Chronic heart failure with reduced ejection fraction | any cancer | 162116 | 13 | 8.52 [6.29, 10.58] | 10 | 10.07 [8.05, 11.97] |  |  |  |
| Chronic heart failure with preserved ejection fraction | past cancer | 34352 | 3 | 14.19 [12.24, 15.99] |  |  |  |  |  |
|  | any cancer | 3844 | 4 | 8.30 [3.99, 12.09] |  |  |  |  |  |
| Acute heart failure | active cancer | 38092 | 3 | 9.75 [7.03, 12.36] |  |  |  |  |  |
|  | past cancer | 360148 | 6 | 7.33 [4.28, 11.24] |  |  | 4 | 8.96 [5.45, 13.85] |  |
|  | metastases | 104836 | 3 | 1.67 [1.14, 2.20] | 3 | 1.67 [1.14, 2.20] |  |  |  |
|  | any cancer | 594492 | 26 | 10.22 [8.49, 11.94] | 12 | 12.99 [11.10, 15.08] | 11 | 8.70 [5.99, 11.22] | 0.01 |
| Any stroke | active cancer | 84810 | 6 | 4.60 [1.72, 8.13] | 4 | 2.26 [0.55, 3.63] |  |  |  |
|  | blood cancer | 502745 | 6 | 0.65 [0.33, 0.94] | 4 | 0.80 [0.56, 1.06] |  |  |  |
|  | past cancer | 347791 | 7 | 13.81 [10.90, 16.60] | 7 | 13.85 [10.55, 16.46] |  |  |  |
|  | metastases | 538431 | 9 | 1.19 [0.80, 1.57] | 6 | 1.35 [0.94, 1.79] |  |  |  |
|  | solid cancer | 503704 | 7 | 7.02 [2.86, 11.57] | 4 | 10.54 [5.00, 15.48] |  |  |  |
|  | any cancer | 1601069 | 39 | 9.52 [7.33, 12.19] | 30 | 10.49 [7.79, 13.51] | 5 | 5.14 [2.32, 8.87] | 0.02 |
| Ischemic stroke | active cancer | 57071 | 5 | 5.26 [2.40, 8.91] | 3 | 2.83 [1.82, 3.62] |  |  |  |
|  | past cancer | 246470 | 5 | 13.75 [9.34, 16.84] | 5 | 13.79 [9.04, 16.94] |  |  |  |
|  | metastases | 21720 | 3 | 1.46 [1.17, 1.85] |  |  |  |  |  |
|  | solid cancer | 21720 | 3 | 7.46 [3.41, 12.55] |  |  |  |  |  |
|  | any cancer | 1285410 | 27 | 9.35 [6.52, 13.14] | 21 | 10.22 [6.45, 14.41] | 4 | 4.55 [2.14, 9.32] | 0.04 |
| Hemorrhagic stroke | any cancer | 21532 | 5 | 11.49 [6.49, 16.97] | 4 | 11.44 [5.05, 17.93] |  |  |  |
| Peripheral artery disease | metastases | 27662 | 3 | 4.44 [3.25, 5.45] | 3 | 4.45 [3.25, 5.45] |  |  |  |
|  | any cancer | 6246746 | 16 | 13.20 [9.11, 17.61] | 12 | 13.85 [8.68, 19.48] |  |  |  |
| Valve heart disease | active cancer | 151839 | 8 | 4.90 [3.84, 6.37] | 6 | 5.11 [3.70, 7.12] |  |  |  |
|  | blood cancer | 182006 | 3 | 1.48 [0.99, 1.83] | 3 | 1.48 [1.00, 1.83] |  |  |  |
|  | past cancer | 100041 | 8 | 15.69 [12.31, 18.65] | 6 | 17.94 [16.03, 19.87] |  |  |  |
|  | metastases | 182791 | 4 | 1.56 [0.37, 2.94] | 4 | 1.56 [0.37, 2.95] |  |  |  |
|  | any cancer | 355417 | 22 | 10.32 [8.28, 12.35] | 19 | 10.80 [8.67, 12.68] | 3 | 6.72 [2.33, 10.82] | 0.09 |
| Aortic stenosis | active cancer | 151011 | 7 | 4.42 [3.64, 5.21] | 5 | 4.41 [3.34, 5.57] |  |  |  |
|  | past cancer | 55295 | 5 | 17.74 [14.60, 20.03] | 4 | 19.00 [16.81, 20.71] |  |  |  |
|  | any cancer | 216037 | 18 | 10.53 [7.70, 13.36] | 15 | 11.22 [8.05, 13.93] | 3 | 6.72 [2.33, 10.82] | 0.09 |
| Transcatheter aortic valve replacement | active cancer | 146467 | 5 | 4.87 [4.11, 5.59] | 4 | 4.86 [3.80, 5.94] |  |  |  |
|  | past cancer | 34293 | 4 | 18.28 [14.40, 20.71] | 3 | 19.98 [19.92, 20.80] |  |  |  |
|  | any cancer | 42011 | 11 | 10.63 [7.12, 15.01] | 10 | 11.27 [7.19, 15.51] |  |  |  |
| Hypertension | any cancer | 974721074 | 17 | 9.30 [5.50, 13.33] | 8 | 9.80 [5.30, 14.27] | 7 | 7.54 [1.87, 15.61] | 0.59 |
|  | any cancer | 721074 | 16 (1 study omitted) | 9.01 [ 5.16, 13.54] |  |  |  |  |  |
| Diabetes mellitus | active cancer | 117866 | 3 | 4.22 [2.18, 5.32] |  |  |  |  |  |
|  | past cancer | 812634 | 12 | 12.72 [4.87, 23.94] | 5 | 11.87 [6.09, 18.94] | 6 | 14.76 [3.07, 35.03] | 0.74 |
|  | metastases | 411447 | 6 | 1.00 [0.53, 1.38] | 5 | 0.91 [0.46, 1.39] |  |  |  |
|  | any cancer | 32311688 | 67 | 9.52 [8.25, 11.02] | 46 | 10.50 [9.01, 12.09] | 15 | 6.28 [3.94, 10.09] | 0.02 |

###### Table S8. Baseline characteristics studies included in meta-analysis of prevalence of any cancer in patients with peripheral artery disease.

| **Variable** | **Europe and Northern America** | **Eastern and South-Eastern Asia** | **Multi-regional** | **Total** |
| --- | --- | --- | --- | --- |
| N of studies | 12 | 2 | 2 | 16 |
| Total population | 6,131,691 | 112,974 | 2081 | 6,246,746 |
| Age, years | 71.89 | 69.78 | 68.65 | 71.16 |
| Female, % | 41.78 | 51.5 |  | 42.52 |
| Study design |  |  |  |  |
| Prospective Cohort | 9 (75%) | 2 (100%) |  | 11 |
| Retrospective Cohort | 3 (25%) |  | 2 (100%) | 5 |
| Settings |  |  |  |  |
| Inpatient | 4 (33.3%) |  | 2 (100%) | 6 |
| Outpatient | 1 (8.3%) | 1 (50%) |  | 2 |
| Both | 1 (8.3%) |  |  | 1 |
| Multicenter | 11 (91.7%) | 2 (100%) | 2 (100%) | 15 |
| Single-center | 1 (8.3%) |  |  | 1 |
| Stable CAD, % | 20.12 |  | 14 | 20.01 |
| History of MI, % | 18.07 |  | 18.9 | 18.08 |
| Heart failure, % | 22.16 | 5.54 | 10.3 | 21.86 |
| Atrial fibrillation, % | 32.41 |  |  | 32.41 |
| Hypertension, % | 59.04 | 87.13 | 82.25 | 59.56 |
| Diabetes mellitus, % | 43.1 | 97.09 | 39.2 | 44.08 |
| Smoking, % | 27.91 |  |  | 27.91 |
| Smoking Active, % | 33.26 |  | 37.69 | 34.77 |
| Past smoking, % |  |  | 56.12 | 56.12 |
| Dyslipidemia, % | 47.93 | 82.72 | 74.33 | 63.07 |
| Obesity, % | 8.66 |  |  | 8.66 |
| Alcohol, % | 0.8 |  |  | 0.8 |
| History of stroke, % | 12.5 | 13.52 | 7.8 | 12.94 |
| CVD, % | 23.07 |  | 19.1 | 22.99 |
| Liver diseases, % | 2.69 |  |  | 2.69 |
| Cirrhosis, % | 1.59 | 8.4 |  | 3.16 |
| CKD, % | 30.15 | 38.54 | 21.45 | 30.3 |
| COPD, % | 13.62 | 5.65 |  | 13.47 |
| Anemia, % | 22 |  |  | 22 |

CAD – Coronary artery disease; MI – Myocardial infarction; CKD – Chronic kidney disease; COPD – Chronic obstructive pulmonary disease.

###### Table S9. Baseline characteristics studies included in meta-analysis of prevalence of unspecified cancer in patients with hypertension.

| **Variable** | **Europe and Northern America** | **Eastern and South-Eastern Asia** | **Northern Africa and Western Asia** | **Multi-regional** | **Total** |
| --- | --- | --- | --- | --- | --- |
| N of studies | 8 | 7 | 1 | 1 | 17 |
| Total population | 97,868,816 | 248,393 | 1015 | 2850 | 98,121,074 |
| Age, years | 55.5 | 66.74 | 60.2 |  | 55.52 |
| Female, % | 50.76 |  |  |  | 50.76 |
| Study design |  |  |  |  |  |
| Retrospective Cohort | 8 (100%) | 5 (71.4%) |  |  |  |
| Prospective Cohort |  | 2 (28.6%) | 1 (100%) | 1 (100%) |  |
| Settings |  |  |  |  |  |
| Inpatient | 2 (25%) | 1 (14.3%) |  |  |  |
| Inpatient/ outpatient |  | 2 (28.6%) |  | 1 (100%) |  |
| Outpatient | 5 (62.5%) | 3 (42.9%) |  |  |  |
| Other |  | 1 (14.3%) |  |  |  |
| Multicenter | 8 (100%) | 7 (100%) | 1 (100%) | 1 (100%) |  |
| Atrial fibrillation, % | 2.28 | 3.13 | 8.2 | 5.6 | 2.33 |
| Diabetes mellitus, % | 18.06 | 42.93 | 31.5 | 29.8 | 18.11 |
| Smoking, % | 23.77 |  |  |  | 23.77 |
| Smoking active, % | 18.39 | 27.8 |  | 6.8 | 20.19 |
| Past smoking | 41.75 |  |  |  | 41.75 |
| Dyslipidemia, % | 59.96 | 30.46 | 38.1 | 51.6 | 59.93 |
| Obesity, % | 44.19 | 1 |  | 30.1 | 44.18 |
| Alcohol, % | 56.82 |  |  |  | 56.82 |
| History of stroke, % | 5.42 | 1.23 | 7.9 |  | 5.42 |
| CVD, % |  | 27.81 |  | 12.5 | 19.55 |
| Liver diseases, % | 1.28 | 8.52 |  | 4.8 | 4.2 |
| CKD, % | 23.31 | 11.46 | 5.8 | 11.5 | 23.3 |
| End-stage CKD, % |  | 0.41 |  |  | 0.41 |
| Bronchial asthma, % | 13.78 | 5.7 | 7.4 |  | 13.78 |
| COPD, % | 10.03 | 35.92 | 10.5 |  | 10.06 |

CVD – Cerebrovascular disease; CKD – Chronic kidney disease; COPD – Chronic obstructive pulmonary disease.

###### Table S10. Baseline characteristics studies included in meta-analysis of prevalence of any cancer in patients with hypertension after exclusion of Alanaeme et al’s study.

| **Variable** | **Europe and Northern America** | **Eastern and South-Eastern Asia** | **Northern Africa and Western Asia** | **Multi-regional** | **Total** |
| --- | --- | --- | --- | --- | --- |
| N of studies | 7 | 7 | 1 | 1 | 16 |
| Total population | 468,816 | 248,393 | 1015 | 2850 | 721,074 |
| Age, years | 69.98 | 66.74 | 60.2 |  | 69.17 |
| Female, % | 50.76 |  |  |  | 50.76 |
| Study design |  |  |  |  |  |
| Retrospective Cohort | 7 (100%) | 5 (71.4%) |  |  | 12 |
| Prospective Cohort |  | 2 (28.6%) | 1 (100%) | 1 (100%) | 4 |
| Settings |  |  |  |  |  |
| Inpatient | 2 (28.6%) | 1 (14.3%) |  |  | 3 |
| Inpatient/ outpatient |  | 2 (28.6%) |  | 1 (100%) | 3 |
| Outpatient | 4 (57.1%) | 3 (42.9%) |  |  | 7 |
| Other |  | 1 (14.3%) |  | 1 (100%) | 2 |
| Multicenter | 7 (100%) | 7 (100%) | 1 (100%) | 1 (100%) | 16 |
| Atrial fibrillation, % | 2.28 | 3.13 | 8.2 | 5.6 | 2.33 |
| Diabetes mellitus, % | 22.82 | 42.93 | 31.5 | 29.8 | 29.18 |
| Smoking, % | 48.08 |  |  |  | 48.08 |
| Smoking active, % | 18.39 | 27.8 |  | 6.8 | 20.19 |
| Past smoking | 41.75 |  |  |  | 41.75 |
| Dyslipidemia, % | 55.48 | 30.46 | 38.1 | 51.6 | 43.35 |
| Obesity, % | 47.2 | 1 |  | 30.1 | 12.19 |
| History of stroke, % | 2.26 | 1.23 | 7.9 |  | 2.21 |
| CVD, % |  | 27.81 |  | 12.5 | 19.55 |
| Liver diseases, % | 1.28 | 8.52 |  | 4.8 | 4.2 |
| CKD, % | 4.02 | 11.46 | 5.8 | 11.5 | 5.92 |
| End-stage CKD, % |  | 0.41 |  |  | 0.41 |
| Bronchial asthma, % | 18.1 | 5.7 | 7.4 |  | 16.67 |
| COPD, % | 3.38 | 35.92 | 10.5 |  | 11.03 |

CVD – Cerebrovascular disease; CKD – Chronic kidney disease; COPD – Chronic obstructive pulmonary disease.

###### Table S11. The results of publication bias assessment.

| Population | Outcome | N | LFK | Interpretation | Egger test p | Thompson-Sharp test p | Rank test p |
| --- | --- | --- | --- | --- | --- | --- | --- |
| Any acute coronary syndrome | previous cancer | 21 | -1.78 | minor asymmetry | 0.41 | 0.42 | 0.81 |
|  | metastases | 14 | -7.8 | major asymmetry | 0 | 0.1 | 0.11 |
|  | unspecified cancer | 67 | -1.14 | minor asymmetry | 0.57 | 0 | 0.15 |
| Atrial fibrillation | active cancer | 19 | -1.33 | minor asymmetry | 0.61 | 0.26 | 0.51 |
|  | previous cancer | 17 | -5.77 | major asymmetry | 0.11 | 0.3 | 0.46 |
|  | metastases | 10 | 1.67 | minor asymmetry | 0.42 | 0.19 | 0.42 |
|  | unspecified cancer | 74 | -2.6 | major asymmetry | 0.4 | 0 | 0.01 |
| Acute heart failure | unspecified cancer | 26 | 3.02 | major asymmetry | 0.01 | 0.11 | 0.03 |
| Any stroke | unspecified cancer | 39 | -0.24 | no asymmetry | 0.67 | 0.01 | 0.89 |
| Acute myocardial infarction | previous cancer | 14 | -2.23 | major asymmetry | 0.39 | 0.93 | 0.96 |
|  | metastases | 11 | -8.42 | major asymmetry | 0 | 0.02 | 0.24 |
|  | unspecified cancer | 50 | -2.28 | major asymmetry | 0.47 | 0 | 0.47 |
| Heart failure | metastases | 10 | 1.4 | minor asymmetry | 0.94 | 0.45 | 0.93 |
|  | unspecified cancer | 56 | -2.12 | major asymmetry | 0.81 | 0 | 0.24 |
| Aortic stenosis | unspecified cancer | 18 | -3.65 | major asymmetry | 0.06 | 0.03 | 0.16 |
| Coronary artery bypass surgery | unspecified cancer | 14 | -0.51 | no asymmetry | 0.72 | 0.68 | 0.55 |
| Coronary artery disease | active cancer | 16 | 0.78 | no asymmetry | 0.54 | 0.3 | 0.93 |
|  | previous cancer | 31 | -1.19 | minor asymmetry | 0.87 | 0.02 | 0.49 |
|  | metastases | 17 | -8.6 | major asymmetry | 0 | 0.02 | 0.05 |
|  | unspecified cancer | 88 | -2.22 | major asymmetry | 0.12 | 0 | 0.93 |
| Heart failure with reduced ejection fraction | unspecified cancer | 13 | -6.59 | major asymmetry | 0.02 | 0.03 | 0.39 |
| Diabetes mellitus | previous cancer | 12 | -4.8 | major asymmetry | 0.46 | 0.01 | 0.27 |
|  | unspecified cancer | 67 | -0.38 | no asymmetry | 0.47 | 0 | 0.07 |
| Hypertension | unspecified cancer | 17 | -9.48 | major asymmetry | 0.35 | 0.03 | 0.87 |
| Ischemic stroke | unspecified cancer | 27 | -0.28 | no asymmetry | 0.88 | 0.08 | 0.72 |
| Non-valvular atrial fibrillation | active cancer | 10 | 0.36 | no asymmetry | 0.84 | 0.68 | 0.93 |
|  | unspecified cancer | 40 | -4.27 | major asymmetry | 0.09 | 0 | 0.26 |
| Peripheral artery disease | unspecified cancer | 16 | 3.7 | major asymmetry | 0.14 | 0.9 | 0.24 |
| Percutaneous coronary intervention | previous cancer | 12 | -1.07 | minor asymmetry | 0.26 | 0.17 | 0.22 |
|  | unspecified cancer | 32 | -1.02 | minor asymmetry | 0.69 | 0.04 | 0.14 |
| ST-elevation myocardial infarction | unspecified cancer | 11 | -4.98 | major asymmetry | 0.15 | 0 | 0.7 |
| Transcatheter aortic valve intervention | unspecified cancer | 11 | 1.09 | minor asymmetry | 0.75 | 0.32 | 0.94 |
| Valve heart disease | unspecified cancer | 22 | -4.55 | major asymmetry | 0.01 | 0.02 | 0.12 |

LFK, Luis Furuya-Kanamori indexes.
