## Supplementary Figures for "“Prevalence of cancer in patients with cardiovascular diseases and risk factors: a systematic review and meta-analysis”"

**Supplementary Materials for the article titled "Prevalence of cancer in patients with cardiovascular diseases and risk factors: a systematic review and meta-analysis".**

**Content**


| | |
|---|---|
| Figure S1. Pooled prevalence of active cancer in coronary artery disease. | 4 |
| Figure S2. Pooled prevalence of any cancer in coronary artery disease. | 5 |
| Figure S3. Pooled prevalence of previous cancer in coronary artery disease. | 6 |
| Figure S4. Pooled prevalence of blood cancer in coronary artery disease. | 7 |
| Figure S5. Pooled prevalence of metastatic cancer in coronary artery disease. | 8 |
| Figure S6. Pooled prevalence of solid cancer in coronary artery disease. | 9 |
| Figure S7. Pooled prevalence of active cancer in acute coronary syndromes. | 10 |
| Figure S8. Pooled prevalence of any cancer in acute coronary syndromes. | 11 |
| Figure S9. Pooled prevalence of previous cancer in acute coronary syndromes. | 12 |
| Figure S10. Pooled prevalence of blood cancer in acute coronary syndromes. | 13 |
| Figure S11. Pooled prevalence of metastatic cancer in acute coronary syndromes. | 14 |
| Figure S12. Pooled prevalence of solid cancer in acute coronary syndromes. | 15 |
| Figure S13. Pooled prevalence of active cancer in acute myocardial infarction. | 16 |
| Figure S14. Pooled prevalence of any cancer in acute myocardial infarction. | 17 |
| Figure S15. Pooled prevalence of previous cancer in acute myocardial infarction. | 18 |
| Figure S16. Pooled prevalence of blood cancer in acute myocardial infarction. | 19 |
| Figure S17. Pooled prevalence of metastatic cancer in acute myocardial infarction. | 20 |
| Figure S18. Pooled prevalence of solid cancer in acute myocardial infarction. | 21 |
| Figure S19. Pooled prevalence of any cancer in ST-elevation myocardial infarction. | 22 |
| Figure S20. Pooled prevalence of previous cancer in ST-elevation myocardial infarction. | 23 |
| Figure S21. Pooled prevalence of any cancer in nonST-elevation myocardial infarction. | 24 |
| Figure S22. Pooled prevalence of any cancer in nonST-elevation acute coronary syndrome. | 25 |
| Figure S23. Pooled prevalence of any cancer in stable angina. | 26 |
| Figure S24. Pooled prevalence of any cancer in patients with history of myocardial infarction. | 27 |
| Figure S25. Pooled prevalence of active cancer in percutaneous coronary intervention. | 28 |
| Figure S26. Pooled prevalence of any cancer in percutaneous coronary intervention. | 29 |
| Figure S27. Pooled prevalence of previous cancer in percutaneous coronary intervention. | 30 |
| Figure S28. Pooled prevalence of metastatic cancer in percutaneous coronary intervention. | 31 |
| Figure S29. Pooled prevalence of any cancer in coronary artery bypass surgery | 32 |
| Figure S30. Pooled prevalence of previous cancer in coronary artery bypass surgery | 33 |
| Figure S31. Pooled prevalence of metastatic cancer in coronary artery bypass surgery | 34 |
| Figure S32. Pooled prevalence of active cancer in atrial fibrillation. | 35 |
| Figure S33. Pooled prevalence of any cancer in atrial fibrillation. | 36 |
| Figure S34. Pooled prevalence of previous cancer in atrial fibrillation. | 37 |
| Figure S35. Pooled prevalence of blood cancer in atrial fibrillation. | 38 |

| | |
|---|---|
| Figure S36. Pooled prevalence of metastatic cancer in atrial fibrillation. | 39 |
| Figure S37. Pooled prevalence of solid cancer in atrial fibrillation. | 40 |
| Figure S38. Pooled prevalence of active cancer in non-valvular atrial fibrillation. | 41 |
| Figure S39. Pooled prevalence of any cancer in non-valvular atrial fibrillation. | 42 |
| Figure S40. Pooled prevalence of previous cancer in non-valvular atrial fibrillation. | 43 |
| Figure S41. Pooled prevalence of blood cancer in non-valvular atrial fibrillation. | 44 |
| Figure S42. Pooled prevalence of metastatic cancer in non-valvular atrial fibrillation. | 45 |
| Figure S43. Pooled prevalence of solid cancer in non-valvular atrial fibrillation. | 46 |
| Figure S44. Pooled prevalence of active cancer in chronic heart failure | 47 |
| Figure S45. Pooled prevalence of any cancer in chronic heart failure | 48 |
| Figure S46. Pooled prevalence of precious cancer in chronic heart failure | 49 |
| Figure S47. Pooled prevalence of blood cancer in chronic heart failure | 50 |
| Figure S48. Pooled prevalence of metastatic cancer in chronic heart failure | 51 |
| Figure S49. Pooled prevalence of solid cancer in chronic heart failure | 52 |
| Figure S50. Pooled prevalence of any cancer in chronic heart failure with reduced ejection fraction | 53 |
| Figure S51. Pooled prevalence of previous cancer in chronic heart failure with preserved ejection fraction | 54 |
| Figure S52. Pooled prevalence of any cancer in chronic heart failure with preserved ejection fraction | 55 |
| Figure S53. Pooled prevalence of active cancer in acute heart failure. | 56 |
| Figure S54. Pooled prevalence of any cancer in acute heart failure. | 57 |
| Figure S55. Pooled prevalence of previous cancer in acute heart failure. | 58 |
| Figure S56. Pooled prevalence of metastatic cancer in acute heart failure. | 59 |
| Figure S57. Pooled prevalence of active cancer in any acute stroke. | 60 |
| Figure S58. Pooled prevalence of any cancer in any acute stroke. | 61 |
| Figure S59. Pooled prevalence of previous cancer in any acute stroke. | 62 |
| Figure S60. Pooled prevalence of blood cancer in any acute stroke. | 63 |
| Figure S61. Pooled prevalence of metastatic cancer in any acute stroke. | 64 |
| Figure S62. Pooled prevalence of solid cancer in any acute stroke. | 65 |
| Figure S63. Pooled prevalence of active cancer in ischemic stroke. | 66 |
| Figure S64. Pooled prevalence of any cancer in ischemic stroke. | 67 |
| Figure S65. Pooled prevalence of previous cancer in ischemic stroke. | 68 |
| Figure S66. Pooled prevalence of metastatic cancer in ischemic stroke. | 69 |
| Figure S67. Pooled prevalence of solid cancer in ischemic stroke. | 70 |
| Figure S68. Pooled prevalence of any cancer in hemorrhagic stroke. | 71 |
| Figure S69. Pooled prevalence of any cancer in peripheral artery disease. | 72 |
| Figure S70. Pooled prevalence of metastatic cancer in peripheral artery disease. | 73 |
| Figure S71. Pooled prevalence of active cancer in valve heart disease. | 74 |

| | |
|---|---|
| Figure S72. Pooled prevalence of any cancer in valve heart disease. | 75 |
| Figure S73. Pooled prevalence of previous cancer in valve heart disease. | 76 |
| Figure S74. Pooled prevalence of blood cancer in valve heart disease. | 77 |
| Figure S75. Pooled prevalence of metastatic cancer in valve heart disease. | 78 |
| Figure S76. Pooled prevalence of active cancer in aortic stenosis. | 79 |
| Figure S77. Pooled prevalence of any cancer in aortic stenosis. | 80 |
| Figure S78. Pooled prevalence of previous cancer in aortic stenosis. | 81 |
| Figure S79. Pooled prevalence of active cancer in transcatheter aortic valve intervention. | 82 |
| Figure S80. Pooled prevalence of any cancer in transcatheter aortic valve intervention. | 83 |
| Figure S81. Pooled prevalence of previous cancer in transcatheter aortic valve intervention. | 84 |
| Figure S82. Pooled prevalence of active cancer in type 2 diabetes mellitus. | 85 |
| Figure S83. Pooled prevalence of any cancer in type 2 diabetes mellitus. | 86 |
| Figure S84. Pooled prevalence of previous cancer in type 2 diabetes mellitus. | 87 |
| Figure S85. Pooled prevalence of metastatic cancer in type 2 diabetes mellitus. | 88 |
| Figure S86. Pooled prevalence of any cancer in hypertension (one study excluded). | 89 |
| Figure S87. Pooled prevalence of any cancer in hypertension. | 90 |
| Figure S88. Pooled prevalence of active cancer in the total study population. | 91 |
| Figure S89. Pooled prevalence of any cancer in the total study population. | 92 |
| Figure S90. Pooled prevalence of previous cancer in the total study population. | 93 |
| Figure S91. Pooled prevalence of blood cancer in the total study population. | 94 |
| Figure S92. Pooled prevalence of metastatic cancer in the total study population. | 95 |
| Figure S93. Pooled prevalence of solid cancer in the total study population. | 96 |

**Figure S1. Pooled prevalence of active cancer in coronary artery disease.**

| Author(s) and Year | Country | Events | N | Prevalence [95% CI] |
|---|---|---|---|---|
| Rodriguez−Padial L, 2021 | ESP | 5425 | 445145 | 1.22 [ 1.19, 1.25] |
| Andersson, 2017 | SWE | 545 | 43145 | 1.26 [ 1.16, 1.37] |
| Itzhaki Ben Zadok O, 2019 | ISR | 53 | 3089 | 1.72 [ 1.31, 2.24] |
| Chandiramani, 2020 | Multinational | 44 | 2397 | 1.84 [ 1.37, 2.46] |
| Yamamoto K, 2024 | JPN | 234 | 12291 | 1.90 [ 1.68, 2.16] |
| Halabi A, 2017 | AUS | 409 | 15912 | 2.57 [ 2.34, 2.83] |
| Hautamäki, 2020 | FIN | 41 | 1576 | 2.60 [ 1.92, 3.51] |
| Okamoto, 2022 | JPN | 5294 | 176275 | 3.00 [ 2.92, 3.08] |
| Biccire FG, 2023 | ITA | 18 | 555 | 3.24 [ 2.05, 5.09] |
| Hess CN, 2015 | USA | 496 | 15008 | 3.30 [ 3.03, 3.60] |
| Valero−Masa, 2017 | ESP | 41 | 1111 | 3.69 [ 2.73, 4.97] |
| Rubens FD, 2022 | CAN | 2130 | 45883 | 4.64 [ 4.45, 4.84] |
| Cao Davide, 2020 | USA | 719 | 9623 | 7.47 [ 6.96, 8.01] |
| Bezin J, 2018 | FRA | 3041 | 31668 | 9.60 [ 9.28, 9.93] |
| Jiménez−Navarro MF, 2015 | USA | 700 | 5350 | 13.08 [12.21, 14.01] |
| Oh TK, 2020 | KOR | 1547 | 10440 | 14.82 [14.15, 15.51] |
| Median prevalence | | | | 3.53 [ 2.40, 5.16] |
| Population−averaged prevalence | | | | 4.61 [ 2.83, 6.97] |

$Tau^2 = 0.64$   $I^2 = 99.9\%$

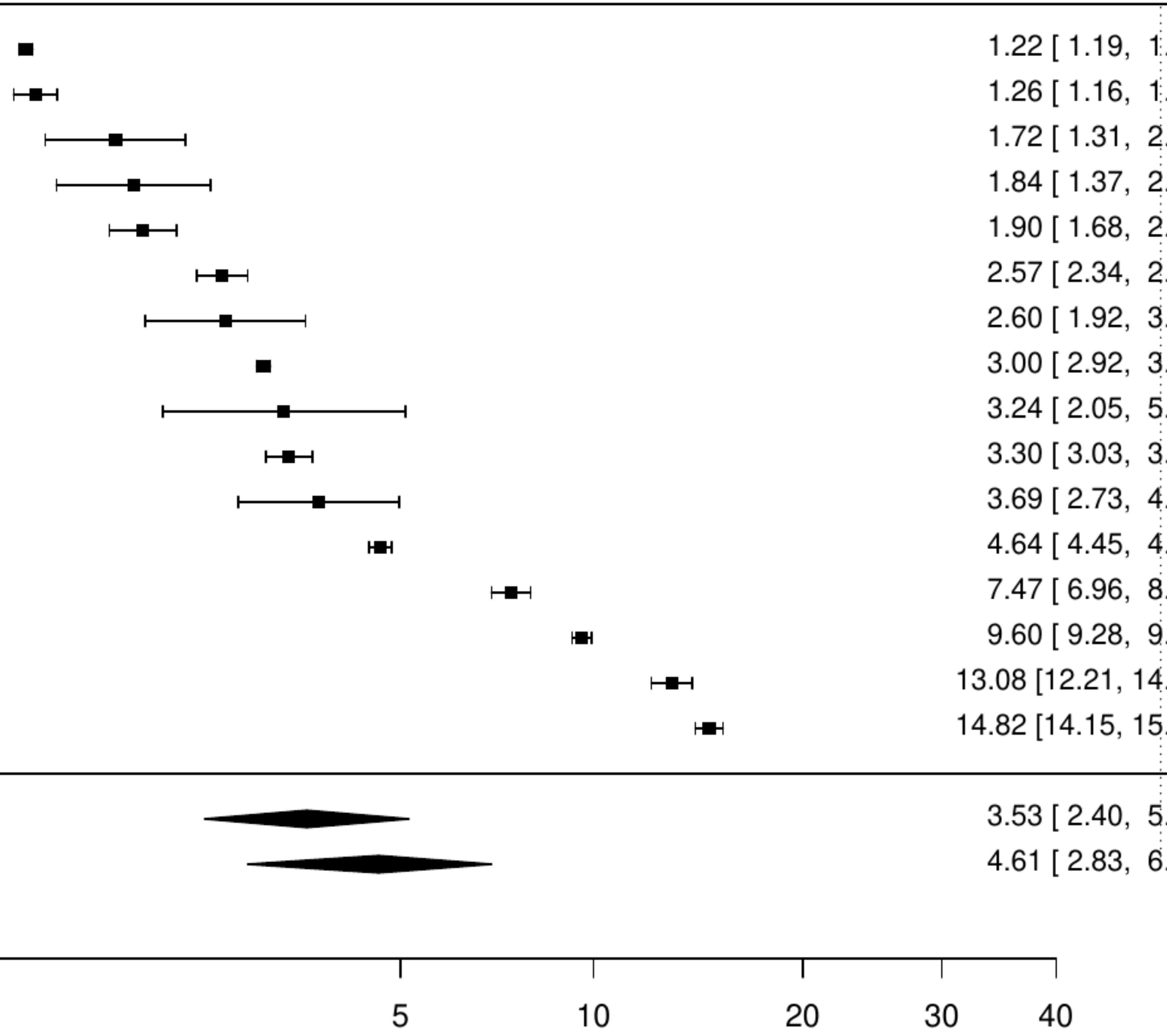

**Figure S2. Pooled prevalence of any cancer in coronary artery disease.**

| Author(s) and Year | Country | Events | N | Prevalence [95% CI] |
|---|---|---|---|---|
| Gupta MD, 2024 | IND | 8 | 5335 | 0.15 [ 0.08, 0.30] |
| Wong CCY, 2021 | AUS | 43 | 10304 | 0.42 [ 0.31, 0.56] |
| Atiya M, 2021 | USA | 1849 | 201116 | 0.92 [ 0.88, 0.96] |
| Shi W, 2021 | CHN | 301 | 24001 | 1.25 [ 1.12, 1.40] |
| Kaul P, 2016 | CAN | 972 | 66008 | 1.47 [ 1.38, 1.57] |
| De Luca, 2017 | ITA | 29 | 1755 | 1.65 [ 1.15, 2.37] |
| Tavella R, 2016 | AUS | 217 | 13071 | 1.66 [ 1.45, 1.89] |
| Rocha JA, 2021 | PRT | 3596 | 212481 | 1.69 [ 1.64, 1.75] |
| Kuehnemund L, 2021 | GER | 16386 | 875735 | 1.87 [ 1.84, 1.90] |
| Vallabhajosyula S, 2020 | USA | 3242 | 163071 | 1.99 [ 1.92, 2.06] |
| Hall M, 2024 | UK | 8667 | 433361 | 2.00 [ 1.96, 2.04] |
| Smilowitz, 2019 | USA | 4482 | 221379 | 2.02 [ 1.97, 2.08] |
| Fowler AJ, 2022 | UK | 893 | 43063 | 2.07 [ 1.94, 2.21] |
| Seki T, 2019 | JPN | 557 | 21409 | 2.60 [ 2.40, 2.82] |
| Erol MK, 2020 | TUR | 54 | 1930 | 2.80 [ 2.15, 3.64] |
| Bucholz EM, 2016 | USA | 1670 | 57574 | 2.90 [ 2.77, 3.04] |
| Guijarro, 2016 | ESP | 16649 | 562837 | 2.96 [ 2.91, 3.00] |
| Itzahki Ben Zadok, 2019 | ISR | 455 | 15024 | 3.03 [ 2.77, 3.31] |
| Kampfer J, 2017 | Multinational | 31 | 1023 | 3.03 [ 2.14, 4.28] |
| van den Broek WWA, 2024 | NLD | 45 | 1320 | 3.41 [ 2.55, 4.54] |
| Goldstone, 2018 | USA | 2116 | 59432 | 3.56 [ 3.41, 3.71] |
| Niknam BA, 2018 | USA | 14805 | 414715 | 3.57 [ 3.51, 3.63] |
| Coll−Fernandez R, 2016 | ESP | 27 | 752 | 3.59 [ 2.47, 5.18] |
| Di Domenicantonio R, 2016 | ITA | 130 | 3608 | 3.60 [ 3.04, 4.26] |
| Johnson SM, 2016 | CAN | 207 | 5578 | 3.71 [ 3.25, 4.24] |
| Sawano M, 2017 | JPN | 487 | 13024 | 3.74 [ 3.43, 4.08] |
| Abdi−Ali A, 2016 | CAN | 3333 | 84373 | 3.95 [ 3.82, 4.08] |
| Altin C, 2022 | TUR | 41 | 1015 | 4.04 [ 2.99, 5.44] |
| Lin GM, 2015 | TW | 30 | 722 | 4.16 [ 2.92, 5.88] |
| Zheng R, 2021 | JPN | 5852 | 138250 | 4.23 [ 4.13, 4.34] |
| Soldati S, 2021 | ITA | 576 | 13540 | 4.25 [ 3.93, 4.61] |
| Hernesniemi, 2019 | FIN | 403 | 9066 | 4.45 [ 4.04, 4.89] |
| Jackel M, 2021 | GER | 28 | 624 | 4.49 [ 3.12, 6.42] |
| Williams MJA, 2019 | NZL | 374 | 8305 | 4.50 [ 4.08, 4.97] |
| Santos, 2021 | PRT | 1588 | 32157 | 4.94 [ 4.71, 5.18] |
| Wu VC, 2021 | TW | 8687 | 170775 | 5.09 [ 4.98, 5.19] |
| Rasia, 2017 | ITA | 124 | 2404 | 5.16 [ 4.34, 6.12] |
| Amann U, 2018 | GER | 90 | 1667 | 5.40 [ 4.41, 6.59] |
| Peyracchia M, 2020 | Multinational | 1106 | 19825 | 5.58 [ 5.27, 5.91] |
| Kim K, 2017 | USA | 1915 | 33027 | 5.80 [ 5.55, 6.06] |
| Nakamura M, 2020 | JPN | 367 | 6267 | 5.86 [ 5.30, 6.47] |
| Kundi, 2019 | USA | 9737 | 166200 | 5.86 [ 5.75, 5.97] |
| Kim MS, 2021 | KOR | 15176 | 258291 | 5.88 [ 5.79, 5.97] |
| Erne P, 2015 | CHE | 2737 | 45091 | 6.07 [ 5.85, 6.29] |
| Wanha W, 2017 | POL | 117 | 1916 | 6.11 [ 5.12, 7.27] |
| Pouche M, 2016 | FRA | 182 | 2894 | 6.29 [ 5.46, 7.23] |
| Akioyamen LE, 2023 | CAN | 3389 | 53841 | 6.29 [ 6.09, 6.50] |
| Alter, 2018 | CAN | 1251 | 19544 | 6.40 [ 6.07, 6.75] |
| De Luca, 2018 | ITA | 331 | 5070 | 6.53 [ 5.88, 7.24] |
| Mackay MH, 2017 | CAN | 2898 | 41792 | 6.93 [ 6.69, 7.18] |
| Nielsen S, 2019 | SWE | 7734 | 110742 | 6.98 [ 6.84, 7.14] |
| Amin AP, 2020 | USA | 30620 | 434172 | 7.05 [ 6.98, 7.13] |
| Saia F, 2016 | ITA | 2777 | 39362 | 7.06 [ 6.81, 7.31] |
| Smolderen Kim G, 2017 | USA | 287 | 4062 | 7.07 [ 6.32, 7.90] |
| Olafiranye O, 2015 | USA | 241 | 3326 | 7.25 [ 6.41, 8.18] |
| Araujo C, 2019 | PRT | 60 | 771 | 7.78 [ 6.09, 9.90] |
| Cilia, 2019 | USA | 850 | 10756 | 7.90 [ 7.41, 8.43] |
| Allonen J, 2020 | FIN | 147 | 1855 | 7.92 [ 6.78, 9.24] |
| Steele AJ, 2018 | UK | 6576 | 82197 | 8.00 [ 7.82, 8.19] |
| Case BC, 2020 | USA | 60127 | 748463 | 8.03 [ 7.97, 8.10] |
| Rymer JA, 2024 | USA | 13087 | 162822 | 8.04 [ 7.91, 8.17] |
| Kazi DS, 2015 | USA | 2735 | 32906 | 8.31 [ 8.02, 8.61] |
| Arai R, 2024 | JPN | 292 | 3411 | 8.56 [ 7.67, 9.55] |
| Tripathi B, 2021 | USA | 1891 | 21738 | 8.70 [ 8.33, 9.08] |
| Shroff GR, 2015 | USA | 3790 | 43168 | 8.78 [ 8.52, 9.05] |
| Takeji Y, 2020 | JPN | 1402 | 15231 | 9.20 [ 8.76, 9.67] |
| Forcadell MJ, 2018 | ESP | 162 | 1733 | 9.35 [ 8.06, 10.81] |
| Puymirat E, 2017 | FRA | 289 | 3079 | 9.39 [ 8.41, 10.47] |
| Belle Loïc, 2017 | FRA | 361 | 3813 | 9.47 [ 8.58, 10.44] |
| Ohya, 2017 | JPN | 163 | 1607 | 10.14 [ 8.76, 11.72] |
| Petersen JK, 2024 | DEN | 14311 | 139137 | 10.29 [10.13, 10.45] |
| Sanchis J, 2019 | ESP | 155 | 1452 | 10.67 [ 9.19, 12.37] |
| Spirito A, 2021 | CHE | 1810 | 16821 | 10.76 [10.30, 11.24] |
| Skalsky K, 2021 | ISR | 1667 | 15153 | 11.00 [10.51, 11.51] |
| Hamood H, 2016 | ISR | 518 | 4655 | 11.13 [10.26, 12.06] |
| Di Martino M, 2015 | ITA | 96 | 778 | 12.34 [10.21, 14.84] |
| Chikwe, 2018 | USA | 2747 | 22245 | 12.35 [11.92, 12.79] |
| Tangri N, 2017 | CAN | 1060 | 8493 | 12.48 [11.79, 13.20] |
| Yamamoto K, 2024 | JPN | 1544 | 12291 | 12.56 [11.99, 13.16] |
| Akyea RK, 2022 | UK | 18311 | 143702 | 12.74 [12.57, 12.92] |
| Raphael CE, 2016 | USA | 2215 | 17165 | 12.90 [12.41, 13.41] |
| Kerola AM, 2022 | FIN | 9930 | 76606 | 12.96 [12.73, 13.20] |
| Solomon J, 2021 | CAN | 65 | 500 | 13.00 [10.33, 16.24] |
| Oikawa T, 2019 | JPN | 123 | 870 | 14.14 [11.98, 16.62] |
| George LK, 2015 | USA | 928 | 5968 | 15.55 [14.65, 16.49] |
| Chamberlain AM, 2015 | USA | 208 | 1159 | 17.95 [15.84, 20.26] |
| Blin P, 2021 | FRA | 47066 | 258260 | 18.22 [18.08, 18.37] |
| Rossini R, 2017 | ITA | 269 | 1082 | 24.86 [22.38, 27.52] |
| Median prevalence | | | | 5.40 [ 4.57, 6.37] |
| Population−averaged prevalence | | | | 7.04 [ 6.05, 8.03] |

Tau² = 0.7 I² = 100%

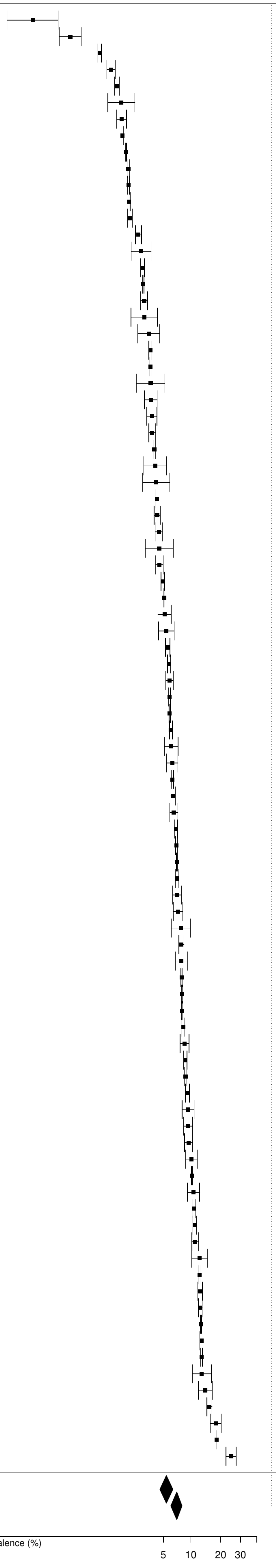

**Figure S3. Pooled prevalence of previous cancer in coronary artery disease.**

| Author(s) and Year | Country | Events | N | Prevalence [95% CI] |
|---|---|---|---|---|
| Erta. FS, 2016 | TUR | 11 | 1034 | 1.06 [ 0.59, 1.91] |
| Kang, 2020 | KOR | 1459 | 108232 | 1.35 [ 1.28, 1.42] |
| Alherbish A, 2017 | CAN | 88 | 6262 | 1.41 [ 1.14, 1.73] |
| Choi IJ, 2021 | KOR | 38 | 1384 | 2.75 [ 2.00, 3.75] |
| Itzhaki Ben Zadok O, 2019 | ISR | 99 | 3089 | 3.20 [ 2.64, 3.89] |
| Sabbag A, 2015 | ISR | 466 | 11501 | 4.05 [ 3.71, 4.43] |
| Pu Aihua, 2017 | CAN | 849 | 20076 | 4.23 [ 3.96, 4.52] |
| Fujita Y, 2022 | JPN | 370 | 7785 | 4.75 [ 4.30, 5.25] |
| Rohla M, 2015 | AUT | 152 | 2890 | 5.26 [ 4.50, 6.14] |
| Nystrom T, 2015 | SWE | 3039 | 53820 | 5.65 [ 5.45, 5.84] |
| Puymirat E, 2016 | FRA | 161 | 2679 | 6.01 [ 5.17, 6.98] |
| Lee SE, 2020 | KOR | 45 | 717 | 6.28 [ 4.72, 8.30] |
| Álvarez−Álvarez B, 2015 | ESP | 381 | 5302 | 7.19 [ 6.52, 7.91] |
| Shore S, 2016 | USA | 240 | 3283 | 7.31 [ 6.47, 8.25] |
| Danchin, 2020 | FRA | 398 | 5147 | 7.73 [ 7.03, 8.49] |
| Teece L, 2023 | UK | 42187 | 512388 | 8.23 [ 8.16, 8.31] |
| Rohla M, 2024 | CHE | 179 | 1996 | 8.97 [ 7.79, 10.30] |
| Nakatsuma, 2018 | JPN | 1109 | 12180 | 9.11 [ 8.61, 9.63] |
| Jacobs JA, 2019 | USA | 58 | 609 | 9.52 [ 7.43, 12.12] |
| Aboal J, 2020 | ESP | 60 | 617 | 9.72 [ 7.62, 12.33] |
| Hwang IC, 2015 | KOR | 860 | 8372 | 10.27 [ 9.64, 10.94] |
| Perl L, 2020 | ISR | 2696 | 25690 | 10.49 [10.13, 10.88] |
| Biccire FG, 2023 | ITA | 59 | 555 | 10.63 [ 8.33, 13.48] |
| Yamaji K, 2018 | CHE | 815 | 7640 | 10.67 [ 9.99, 11.38] |
| Tam DY, 2023 | CAN | 2468 | 22586 | 10.93 [10.53, 11.34] |
| Wang F, 2016 | USA | 263 | 2346 | 11.21 [10.00, 12.55] |
| Palomäki A, 2021 | FIN | 7263 | 60446 | 12.02 [11.76, 12.28] |
| de la Torre Hernandez Jose M, 2021 | ESP | 161 | 1249 | 12.89 [11.14, 14.87] |
| Sanchez F, 2021 | ITA | 146 | 1086 | 13.44 [11.54, 15.60] |
| Dalen M, 2024 | SWE | 461 | 3267 | 14.11 [12.96, 15.35] |
| Hsieh CF, 2015 | TW | 6821 | 9805 | 69.57 [68.65, 70.47] |
| Median prevalence | | | | 7.26 [ 5.33, 9.82] |
| Population−averaged prevalence | Tau² = 0.87 I² = 99.9% | | | 9.72 [ 6.97, 14.17] |

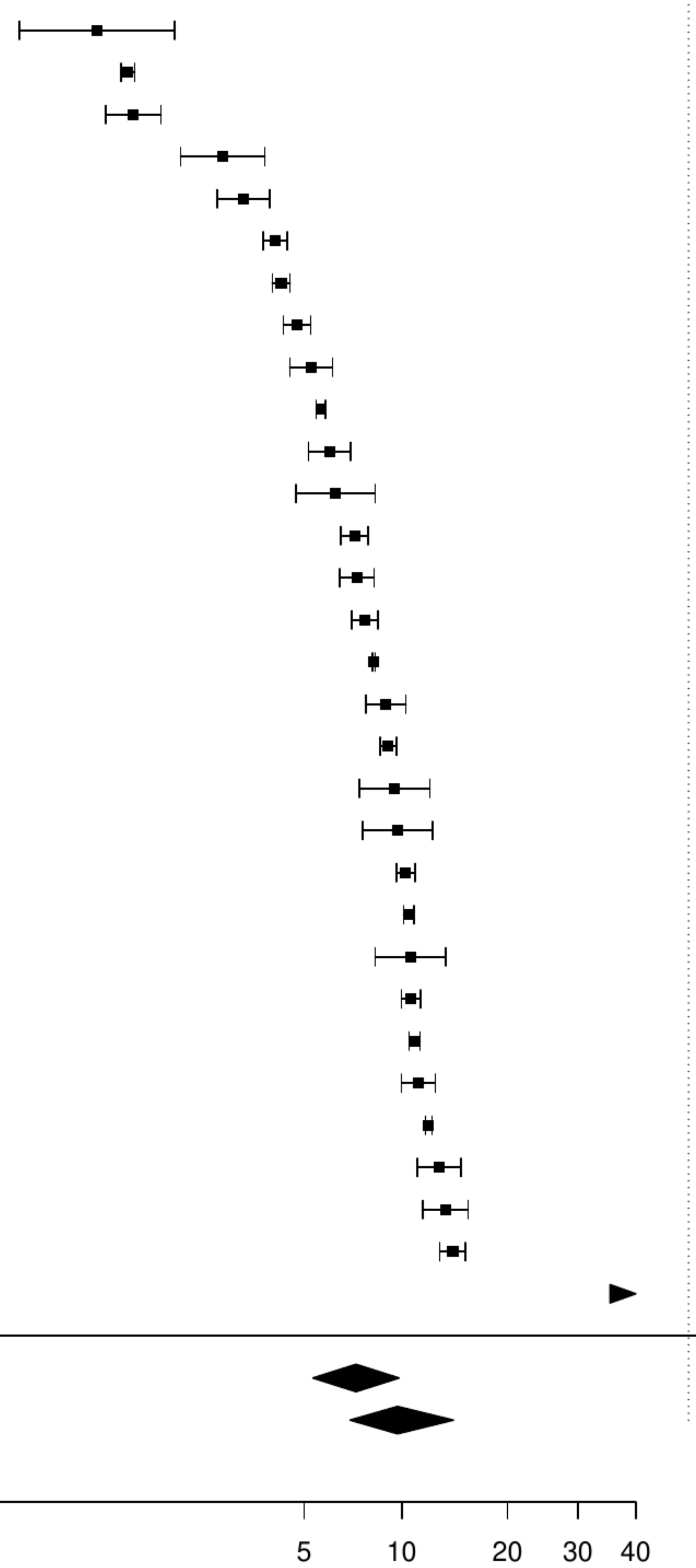

**Figure S4. Pooled prevalence of blood cancer in coronary artery disease.**

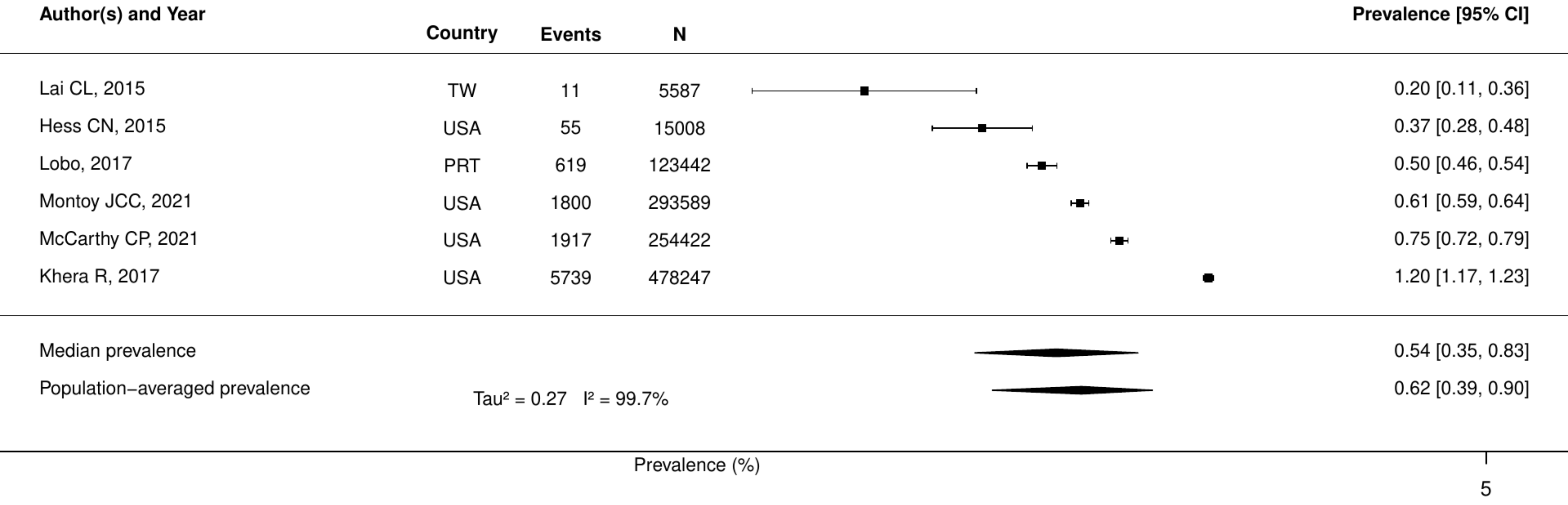

**Figure S5. Pooled prevalence of metastatic cancer in coronary artery disease.**

| Author(s) and Year | Country | Events | N | Prevalence [95% CI] |
|---|---|---|---|---|
| Vallabhajosyula S, 2020 | USA | 49 | 163071 | 0.03 [0.02, 0.04] |
| Seki T, 2019 | JPN | 25 | 21409 | 0.12 [0.08, 0.17] |
| Lai CL, 2015 | TW | 16 | 5587 | 0.29 [0.18, 0.47] |
| Kyto V, 2022 | FIN | 142 | 40654 | 0.35 [0.30, 0.41] |
| Rubens FD, 2022 | CAN | 208 | 45883 | 0.45 [0.40, 0.52] |
| Lobo, 2017 | PRT | 619 | 123442 | 0.50 [0.46, 0.54] |
| Bezin J, 2018 | FRA | 226 | 31668 | 0.71 [0.63, 0.81] |
| Hautamäki, 2020 | FIN | 13 | 1576 | 0.82 [0.48, 1.42] |
| Williams MJA, 2019 | NZL | 71 | 8305 | 0.85 [0.68, 1.08] |
| Alter, 2018 | CAN | 186 | 19544 | 0.95 [0.82, 1.10] |
| Singh, 2022 | USA | 158 | 15128 | 1.04 [0.89, 1.22] |
| Tran, 2018 | CAN | 556 | 41210 | 1.35 [1.24, 1.47] |
| Oh TK, 2020 | KOR | 154 | 10440 | 1.48 [1.26, 1.73] |
| McCarthy CP, 2021 | USA | 4342 | 254422 | 1.71 [1.66, 1.76] |
| Worrall−Carter Linda, 2016 | AUS | 318 | 16771 | 1.90 [1.70, 2.11] |
| Montoy JCC, 2021 | USA | 7404 | 293589 | 2.52 [2.47, 2.58] |
| Wang Y, 2020 | USA | 48910 | 884931 | 5.53 [5.48, 5.57] |
| Median prevalence | | | | 0.72 [0.41, 1.27] |
| Population−averaged prevalence | | | | 1.39 [0.81, 2.23] |

Tau² = 1.41 I² = 100%

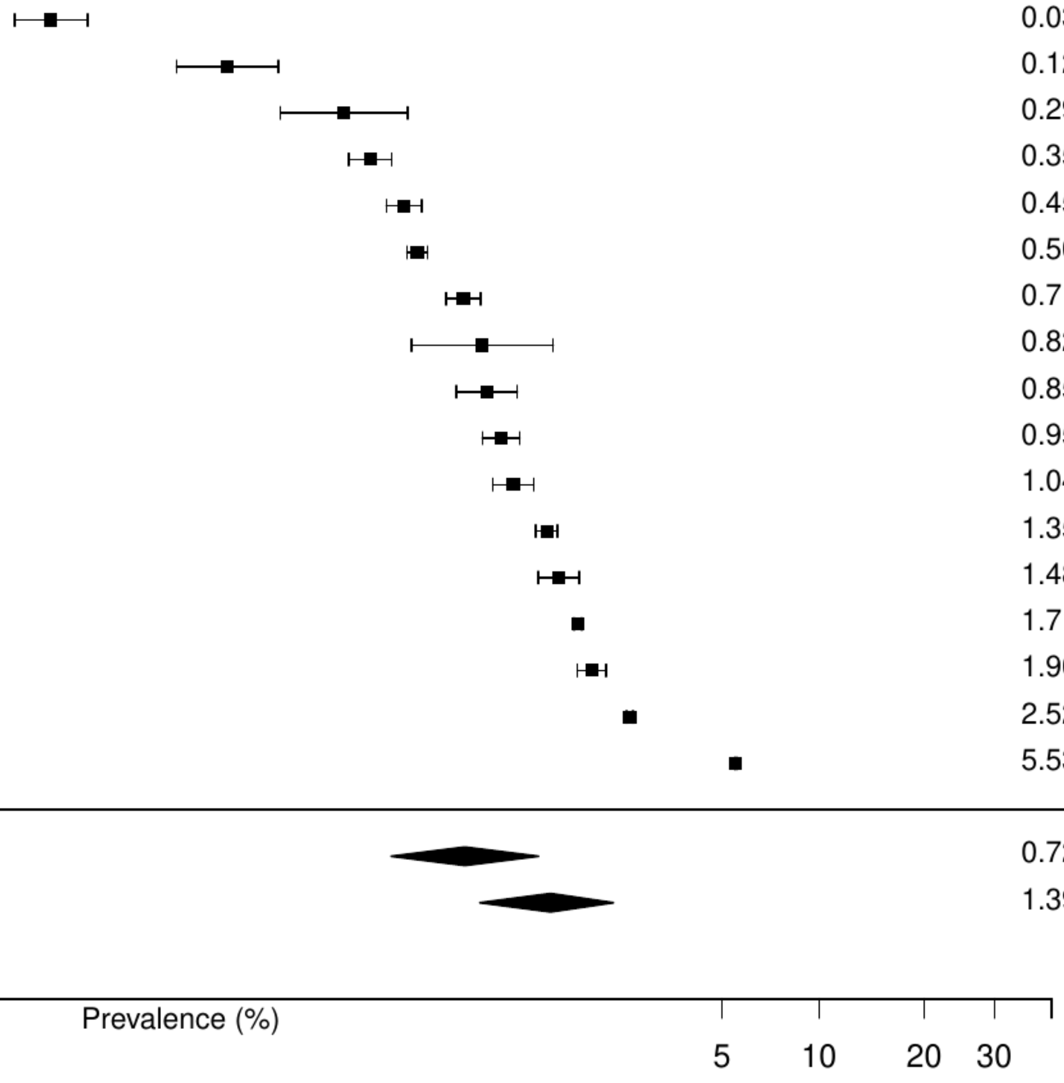

**Figure S6. Pooled prevalence of solid cancer in coronary artery disease.**

| Author(s) and Year | Country | Events | N | Prevalence [95% CI] |
|---|---|---|---|---|
| Montoy JCC, 2021 | USA | 3054 | 293589 | 1.04 [ 1.00, 1.08] |
| McCarthy CP, 2021 | USA | 5204 | 254422 | 2.05 [ 1.99, 2.10] |
| Worrall−Carter Linda, 2016 | AUS | 690 | 16771 | 4.11 [ 3.82, 4.43] |
| Lai CL, 2015 | TW | 288 | 6584 | 4.37 [ 3.91, 4.90] |
| Khera R, 2017 | USA | 49456 | 478247 | 10.34 [10.26, 10.43] |
| Median prevalence | | | | 3.34 [ 1.68, 6.55] |
| Population−averaged prevalence | | | | 4.39 [ 2.04, 7.72] |

$Tau^2 = 0.65$ $I^2 = 100\%$

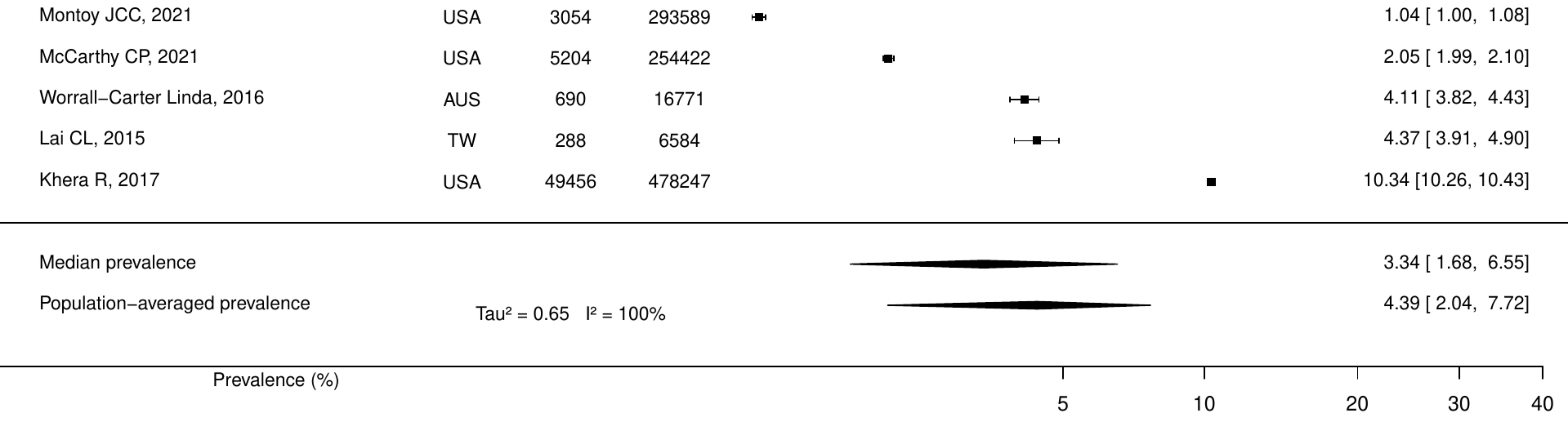

**Figure S7. Pooled prevalence of active cancer in acute coronary syndromes.**

| Author(s) and Year | Country | Events | N | Prevalence [95% CI] |
|---|---|---|---|---|
| Rodriguez−Padial L, 2021 | ESP | 5425 | 445145 | 1.22 [1.19, 1.25] |
| Andersson, 2017 | SWE | 545 | 43145 | 1.26 [1.16, 1.37] |
| Itzhaki Ben Zadok O, 2019 | ISR | 53 | 3089 | 1.72 [1.31, 2.24] |
| Halabi A, 2017 | AUS | 409 | 15912 | 2.57 [2.34, 2.83] |
| Hautamäki, 2020 | FIN | 41 | 1576 | 2.60 [1.92, 3.51] |
| Okamoto, 2022 | JPN | 5294 | 176275 | 3.00 [2.92, 3.08] |
| Biccire FG, 2023 | ITA | 18 | 555 | 3.24 [2.05, 5.09] |
| Valero−Masa, 2017 | ESP | 41 | 1111 | 3.69 [2.73, 4.97] |
| Bezin J, 2018 | FRA | 3041 | 31668 | 9.60 [9.28, 9.93] |
| Median prevalence | | | | 2.63 [1.75, 3.92] |
| Population−averaged prevalence | | | | 3.12 [1.91, 5.13] |

Tau² = 0.39   I² = 99.8%

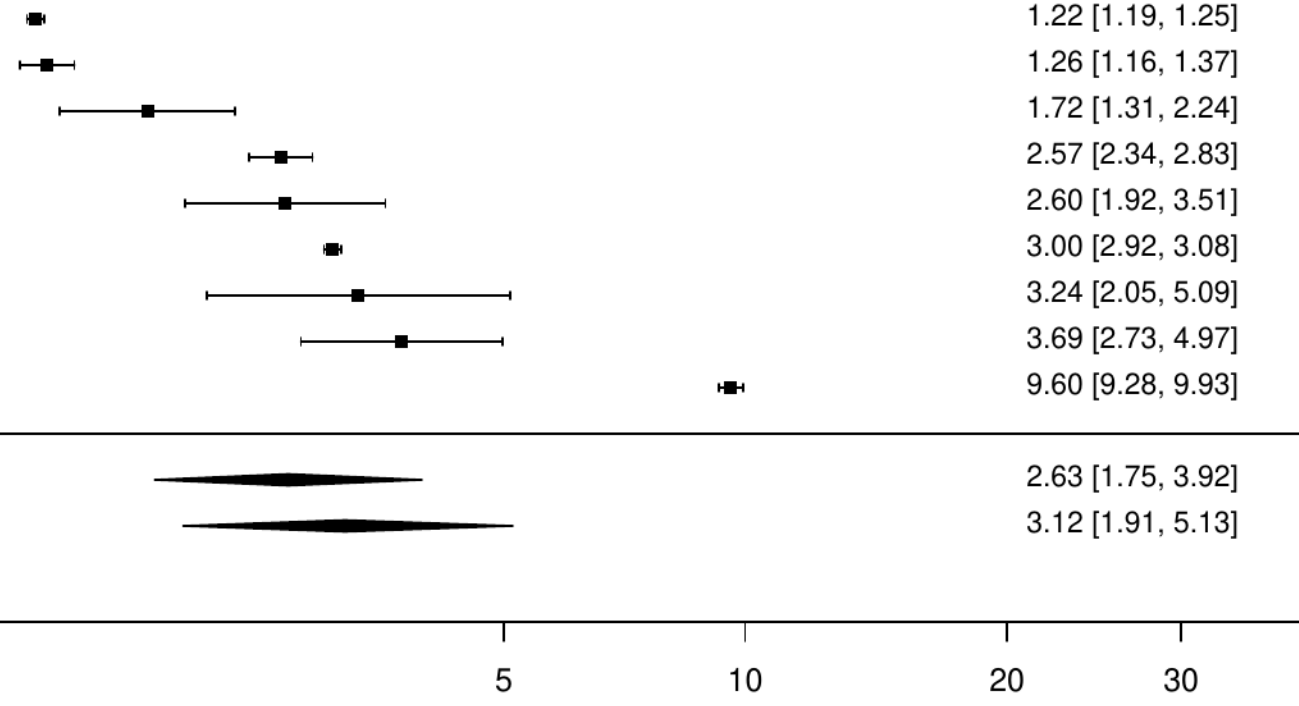

**Figure S8. Pooled prevalence of any cancer in acute coronary syndromes.**

| Author(s) and Year | Country | Events | N | Prevalence [95% CI] |
|---|---|---|---|---|
| Gupta MD, 2024 | IND | 8 | 5335 | 0.15 [ 0.08, 0.30] |
| Carrero JJ, 2017 | SWE | 146 | 36001 | 0.41 [ 0.34, 0.48] |
| Shi W, 2021 | CHN | 301 | 24001 | 1.25 [ 1.12, 1.40] |
| De Luca, 2017 | ITA | 29 | 1755 | 1.65 [ 1.15, 2.37] |
| Tavella R, 2016 | AUS | 217 | 13071 | 1.66 [ 1.45, 1.89] |
| Rocha JA, 2021 | PRT | 3596 | 212481 | 1.69 [ 1.64, 1.75] |
| Kuehnemund L, 2021 | GER | 16386 | 875735 | 1.87 [ 1.84, 1.90] |
| Vallabhajosyula S, 2020 | USA | 3242 | 163071 | 1.99 [ 1.92, 2.06] |
| Hall M, 2024 | UK | 8667 | 433361 | 2.00 [ 1.96, 2.04] |
| Hagen TP, 2015 | NOR | 229 | 10522 | 2.18 [ 1.91, 2.47] |
| Bainey KR, 2018 | CAN | 1523 | 60717 | 2.51 [ 2.39, 2.64] |
| Erol MK, 2020 | TUR | 54 | 1930 | 2.80 [ 2.15, 3.64] |
| Bucholz EM, 2016 | USA | 1670 | 57574 | 2.90 [ 2.77, 3.04] |
| Guijarro, 2016 | ESP | 16649 | 562837 | 2.96 [ 2.91, 3.00] |
| Itzahki Ben Zadok, 2019 | ISR | 455 | 15024 | 3.03 [ 2.77, 3.31] |
| Kampfer J, 2017 | Multinational | 31 | 1023 | 3.03 [ 2.14, 4.28] |
| van den Broek WWA, 2024 | NLD | 45 | 1320 | 3.41 [ 2.55, 4.54] |
| Angerås, 2016 | SWE | 3654 | 104012 | 3.51 [ 3.40, 3.63] |
| Niknam BA, 2018 | USA | 14805 | 414715 | 3.57 [ 3.51, 3.63] |
| Di Domenicantonio R, 2016 | ITA | 130 | 3608 | 3.60 [ 3.04, 4.26] |
| Shiraishi Y, 2016 | JPN | 130 | 3287 | 3.95 [ 3.34, 4.68] |
| Quintana, 2018 | SWE | 79 | 1984 | 3.98 [ 3.21, 4.94] |
| Zheng R, 2021 | JPN | 5852 | 138250 | 4.23 [ 4.13, 4.34] |
| Soldati S, 2021 | ITA | 576 | 13540 | 4.25 [ 3.93, 4.61] |
| Hernesniemi, 2019 | FIN | 403 | 9066 | 4.45 [ 4.04, 4.89] |
| Jackel M, 2021 | GER | 28 | 624 | 4.49 [ 3.12, 6.42] |
| Williams MJA, 2019 | NZL | 374 | 8305 | 4.50 [ 4.08, 4.97] |
| Shoji S, 2020 | JPN | 118 | 2559 | 4.61 [ 3.86, 5.50] |
| Santos, 2021 | PRT | 1588 | 32157 | 4.94 [ 4.71, 5.18] |
| Rasia, 2017 | ITA | 124 | 2404 | 5.16 [ 4.34, 6.12] |
| Peyracchia M, 2020 | Multinational | 1106 | 19825 | 5.58 [ 5.27, 5.91] |
| Kirchberger Inge, 2016 | GER | 106 | 1855 | 5.71 [ 4.75, 6.87] |
| Roffi M, 2020 | CHE | 2558 | 44156 | 5.79 [ 5.58, 6.01] |
| Kim K, 2017 | USA | 1915 | 33027 | 5.80 [ 5.55, 6.06] |
| Kundi, 2019 | USA | 9737 | 166200 | 5.86 [ 5.75, 5.97] |
| Kim MS, 2021 | KOR | 15176 | 258291 | 5.88 [ 5.79, 5.97] |
| Joergensen TS, 2016 | DEN | 5123 | 87188 | 5.88 [ 5.72, 6.03] |
| Wanha W, 2017 | POL | 117 | 1916 | 6.11 [ 5.12, 7.27] |
| Wu VC, 2018 | TW | 11005 | 177058 | 6.22 [ 6.10, 6.33] |
| Ozaki AF, 2022 | CAN | 1500 | 23962 | 6.26 [ 5.96, 6.57] |
| Alter, 2018 | CAN | 1251 | 19544 | 6.40 [ 6.07, 6.75] |
| Ederhy S, 2019 | FRA | 246 | 3664 | 6.71 [ 5.95, 7.57] |
| Iqbal MB, 2016 | CAN | 771 | 11181 | 6.90 [ 6.44, 7.38] |
| Amin AP, 2020 | USA | 30620 | 434172 | 7.05 [ 6.98, 7.13] |
| Smolderen Kim G, 2017 | USA | 287 | 4062 | 7.07 [ 6.32, 7.90] |
| Araujo C, 2019 | PRT | 60 | 771 | 7.78 [ 6.09, 9.90] |
| Allonen J, 2020 | FIN | 147 | 1855 | 7.92 [ 6.78, 9.24] |
| Case BC, 2020 | USA | 60127 | 748463 | 8.03 [ 7.97, 8.10] |
| Rymer JA, 2024 | USA | 13087 | 162822 | 8.04 [ 7.91, 8.17] |
| Yamashita Y, 2017 | JPN | 318 | 3942 | 8.07 [ 7.26, 8.96] |
| Jackevicius CA, 2016 | CAN | 1805 | 21319 | 8.47 [ 8.10, 8.85] |
| Arai R, 2024 | JPN | 292 | 3411 | 8.56 [ 7.67, 9.55] |
| Tripathi B, 2021 | USA | 1891 | 21738 | 8.70 [ 8.33, 9.08] |
| Campanile A, 2024 | ITA | 115 | 1310 | 8.78 [ 7.36, 10.44] |
| Shroff GR, 2015 | USA | 3790 | 43168 | 8.78 [ 8.52, 9.05] |
| Puymirat E, 2017 | FRA | 289 | 3079 | 9.39 [ 8.41, 10.47] |
| Belle Loïc, 2017 | FRA | 361 | 3813 | 9.47 [ 8.58, 10.44] |
| Oberprieler NG, 2022 | NOR | 4251 | 43750 | 9.72 [ 9.44, 10.00] |
| Sanchis J, 2019 | ESP | 155 | 1452 | 10.67 [ 9.19, 12.37] |
| Obayashi Y, 2021 | JPN | 690 | 6228 | 11.08 [10.32, 11.88] |
| Hamood H, 2016 | ISR | 518 | 4655 | 11.13 [10.26, 12.06] |
| Di Martino M, 2015 | ITA | 96 | 778 | 12.34 [10.21, 14.84] |
| Tangri N, 2017 | CAN | 1060 | 8493 | 12.48 [11.79, 13.20] |
| Kerola AM, 2022 | FIN | 9930 | 76606 | 12.96 [12.73, 13.20] |
| Takeuchi T, 2021 | JPN | 462 | 3499 | 13.20 [12.12, 14.37] |
| Al−Hijji Mohammed A, 2018 | USA | 749 | 5660 | 13.23 [12.38, 14.14] |
| Chamberlain AM, 2015 | USA | 208 | 1159 | 17.95 [15.84, 20.26] |
| Median prevalence | Tau² = 0.65 I² = 100% | | | 4.97 [ 4.13, 5.96] |
| Population−averaged prevalence | | | | 6.38 [ 5.49, 7.19] |

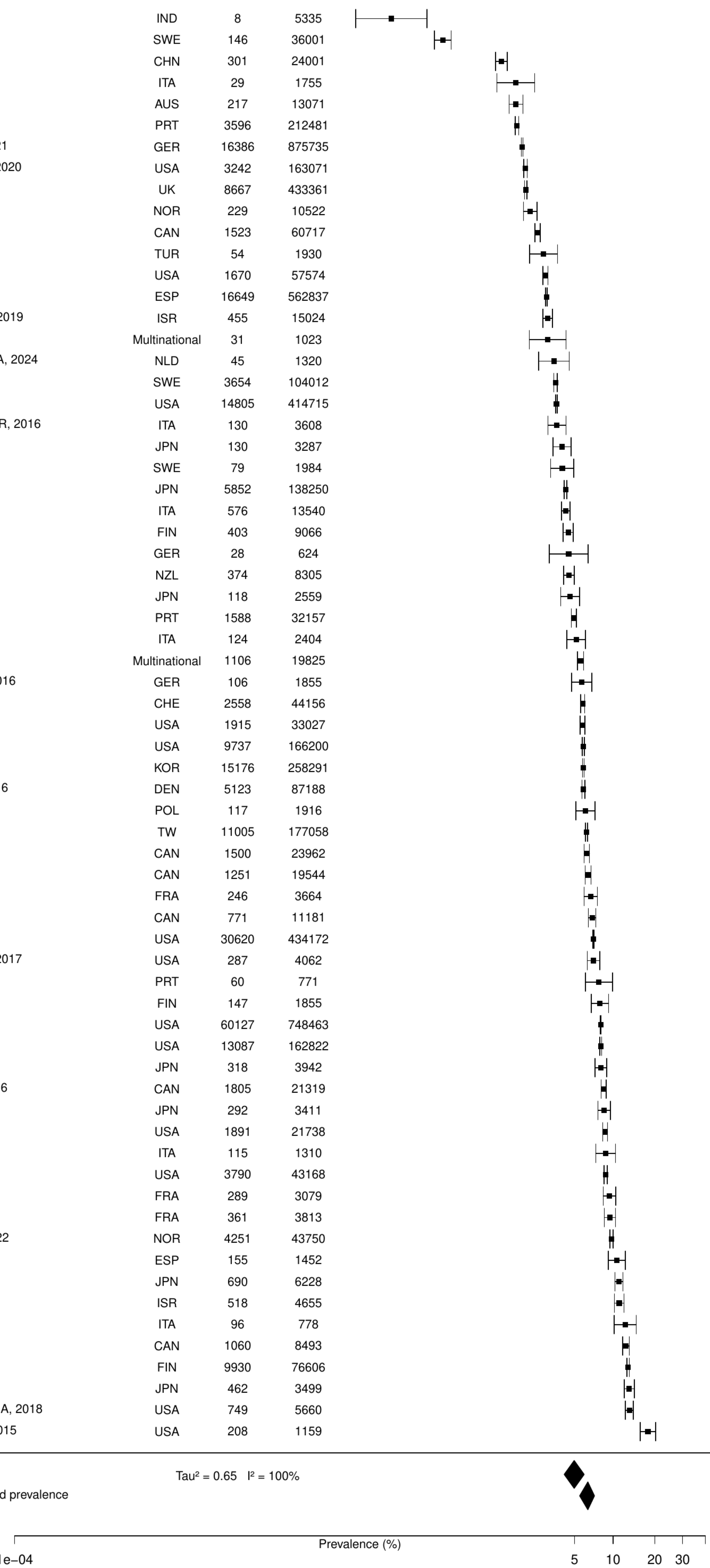

**Figure S9. Pooled prevalence of previous cancer in acute coronary syndromes.**

| Author(s) and Year | Country | Events | N | Prevalence [95% CI] |
|---|---|---|---|---|
| Erta. FS, 2016 | TUR | 11 | 1034 | 1.06 [ 0.59, 1.91] |
| Kang, 2020 | KOR | 1459 | 108232 | 1.35 [ 1.28, 1.42] |
| Szummer K, 2017 | SWE | 1957 | 105674 | 1.85 [ 1.77, 1.93] |
| Jamaly S, 2021 | SWE | 590 | 25384 | 2.32 [ 2.15, 2.52] |
| Szummer, 2018 | SWE | 5346 | 205693 | 2.60 [ 2.53, 2.67] |
| Itzhaki Ben Zadok O, 2019 | ISR | 99 | 3089 | 3.20 [ 2.64, 3.89] |
| Sabbag A, 2015 | ISR | 466 | 11501 | 4.05 [ 3.71, 4.43] |
| Álvarez–Álvarez B, 2015 | ESP | 381 | 5302 | 7.19 [ 6.52, 7.91] |
| Shore S, 2016 | USA | 240 | 3283 | 7.31 [ 6.47, 8.25] |
| Bouisset F, 2021 | FRA | 491 | 6243 | 7.86 [ 7.22, 8.56] |
| Teece L, 2023 | UK | 42187 | 512388 | 8.23 [ 8.16, 8.31] |
| Rohla M, 2024 | CHE | 179 | 1996 | 8.97 [ 7.79, 10.30] |
| Jacobs JA, 2019 | USA | 58 | 609 | 9.52 [ 7.43, 12.12] |
| Aboal J, 2020 | ESP | 60 | 617 | 9.72 [ 7.62, 12.33] |
| Biccire FG, 2023 | ITA | 59 | 555 | 10.63 [ 8.33, 13.48] |
| Wang F, 2016 | USA | 263 | 2346 | 11.21 [10.00, 12.55] |
| Chuang AM, 2018 | AUS | 348 | 3071 | 11.33 [10.26, 12.50] |
| Palomäki A, 2021 | FIN | 7263 | 60446 | 12.02 [11.76, 12.28] |
| Sanchez F, 2021 | ITA | 146 | 1086 | 13.44 [11.54, 15.60] |
| Bjorklund E, 2021 | SWE | 980 | 6558 | 14.94 [14.10, 15.83] |
| Bentzel S, 2024 | SWE | 591 | 3765 | 15.70 [14.57, 16.89] |
| Median prevalence | | | | 6.23 [ 4.43, 8.69] |
| Population–averaged prevalence | Tau² = 0.7 I² = 99.9% | | | 8.02 [ 5.92, 9.96] |

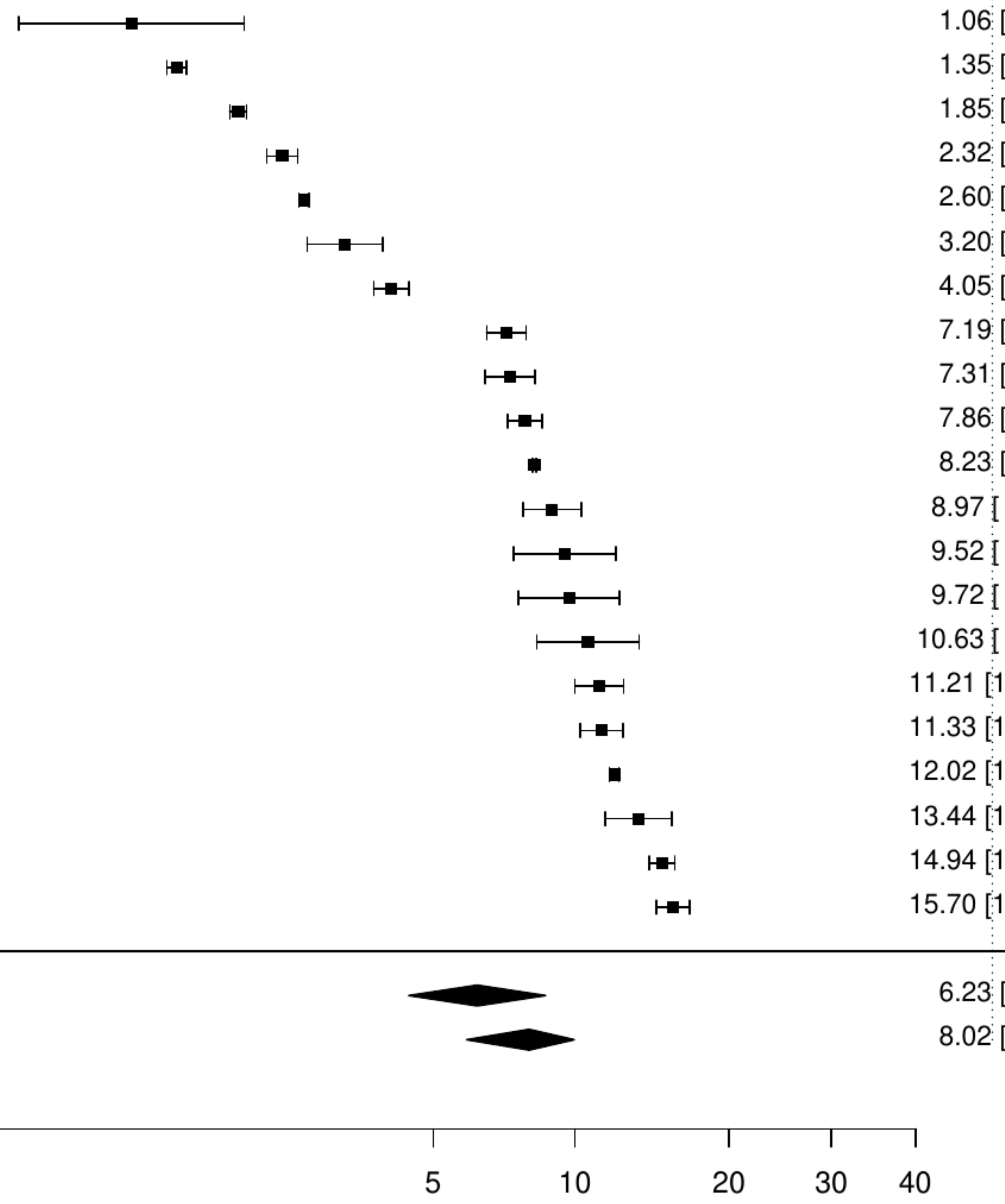

**Figure S10. Pooled prevalence of blood cancer in acute coronary syndromes.**

| Author(s) and Year | Country | Events | N | Prevalence [95% CI] |
|---|---|---|---|---|
| Lobo, 2017 | PRT | 619 | 123442 | 0.50 [0.46, 0.54] |
| Montoy JCC, 2021 | USA | 1800 | 293589 | 0.61 [0.59, 0.64] |
| McCarthy CP, 2021 | USA | 1917 | 254422 | 0.75 [0.72, 0.79] |
| Khera R, 2017 | USA | 5739 | 478247 | 1.20 [1.17, 1.23] |
| Median prevalence | | | | 0.73 [0.53, 1.00] |
| Population−averaged prevalence | | | | 0.76 [0.56, 1.03] |

Tau² = 0.11   I² = 99.5%

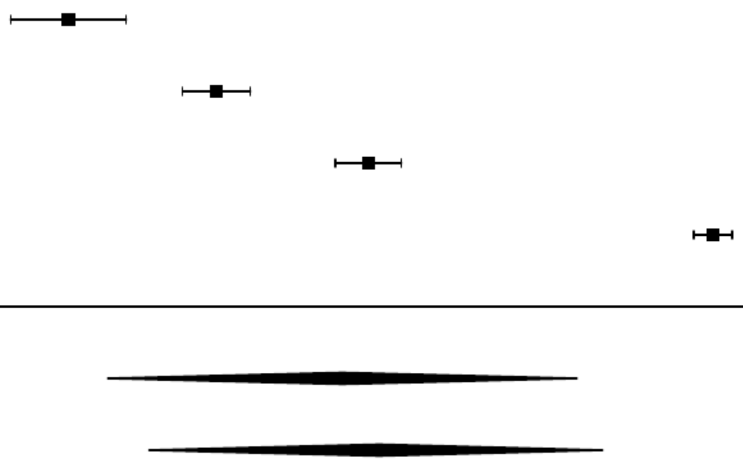

**Figure S11. Pooled prevalence of metastatic cancer in acute coronary syndromes.**

| Author(s) and Year | Country | Events | N | Prevalence [95% CI] |
|---|---|---|---|---|
| Vallabhajosyula S, 2020 | USA | 49 | 163071 | 0.03 [0.02, 0.04] |
| Kyto V, 2022 | FIN | 142 | 40654 | 0.35 [0.30, 0.41] |
| Lobo, 2017 | PRT | 619 | 123442 | 0.50 [0.46, 0.54] |
| Bezin J, 2018 | FRA | 226 | 31668 | 0.71 [0.63, 0.81] |
| Hautamäki, 2020 | FIN | 13 | 1576 | 0.82 [0.48, 1.42] |
| Williams MJA, 2019 | NZL | 71 | 8305 | 0.85 [0.68, 1.08] |
| Alter, 2018 | CAN | 186 | 19544 | 0.95 [0.82, 1.10] |
| Yasaitis LC, 2020 | CAN | 1237 | 112484 | 1.10 [1.04, 1.16] |
| Spoon DB, 2015 | USA | 74 | 6636 | 1.12 [0.89, 1.40] |
| Tran, 2018 | CAN | 556 | 41210 | 1.35 [1.24, 1.47] |
| McCarthy CP, 2021 | USA | 4342 | 254422 | 1.71 [1.66, 1.76] |
| Worrall−Carter Linda, 2016 | AUS | 318 | 16771 | 1.90 [1.70, 2.11] |
| Montoy JCC, 2021 | USA | 7404 | 293589 | 2.52 [2.47, 2.58] |
| Wang Y, 2020 | USA | 48910 | 884931 | 5.53 [5.48, 5.57] |
| Median prevalence | | | | 0.89 [0.49, 1.62] |
| Population−averaged prevalence | | | | 1.64 [0.99, 2.62] |

Tau² = 1.33   I² = 100%

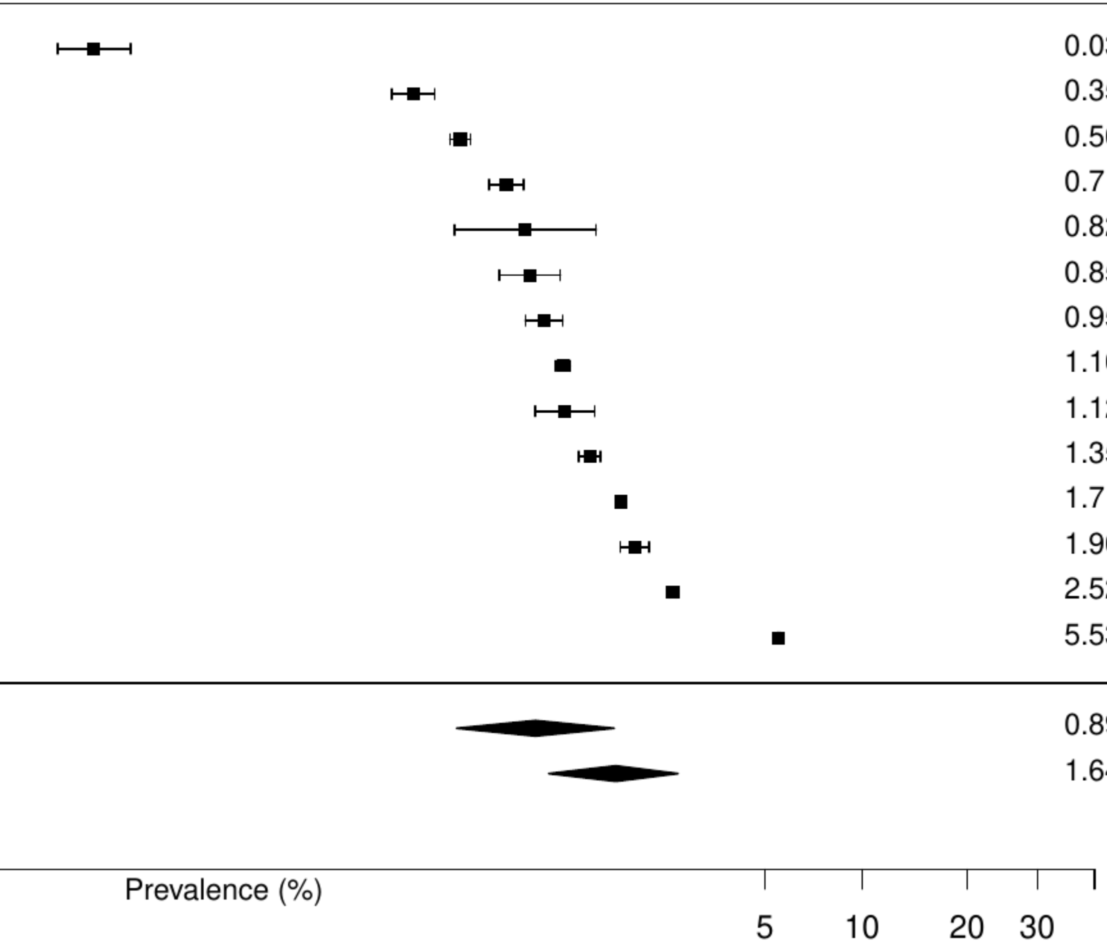

**Figure S12. Pooled prevalence of solid cancer in acute coronary syndromes.**

| Author(s) and Year | Country | Events | N | Prevalence [95% CI] |
|---|---|---|---|---|
| Montoy JCC, 2021 | USA | 3054 | 293589 | 1.04 [ 1.00, 1.08] |
| McCarthy CP, 2021 | USA | 5204 | 254422 | 2.05 [ 1.99, 2.10] |
| Worrall−Carter Linda, 2016 | AUS | 690 | 16771 | 4.11 [ 3.82, 4.43] |
| Khera R, 2017 | USA | 49456 | 478247 | 10.34 [10.26, 10.43] |
| Median prevalence | | | | 3.13 [ 1.33, 7.15] |
| Population−averaged prevalence | | | | 4.34 [ 1.54, 8.65] |

Tau² = 0.79 I² = 100%

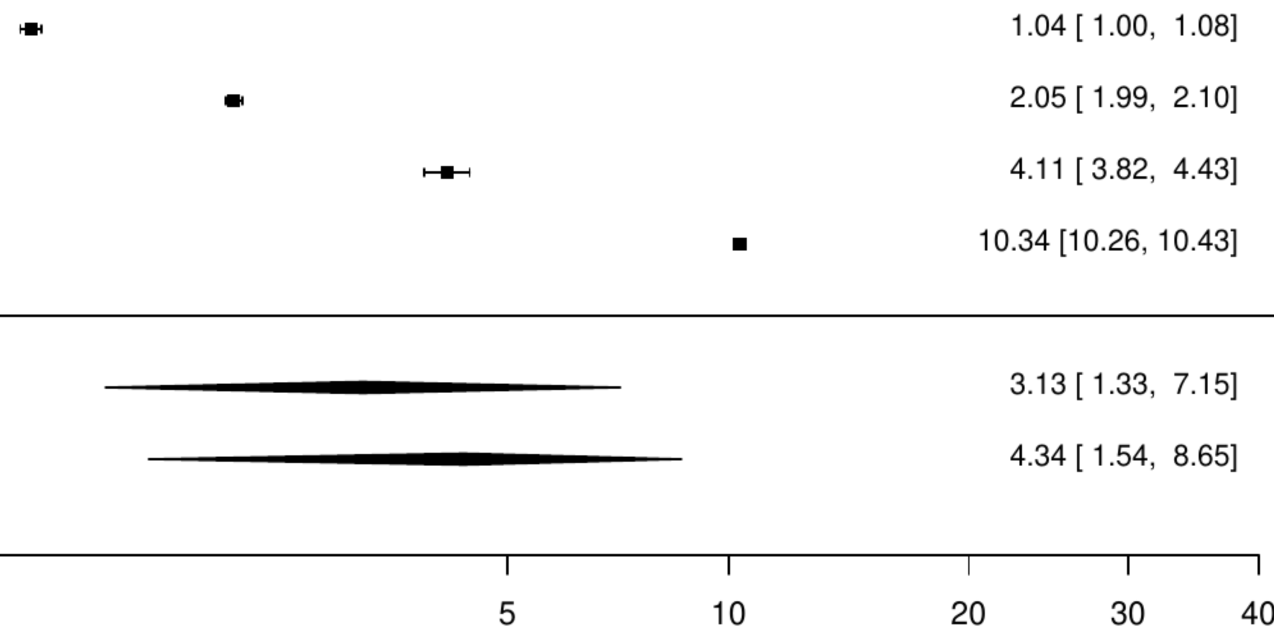

**Figure S13. Pooled prevalence of active cancer in acute myocardial infarction.**

| Author(s) and Year | Country | Events | N | Prevalence [95% CI] |
|---|---|---|---|---|
| Rodriguez−Padial L, 2021 | ESP | 5425 | 445145 | 1.22 [1.19, 1.25] |
| Andersson, 2017 | SWE | 545 | 43145 | 1.26 [1.16, 1.37] |
| Okamoto, 2022 | JPN | 5294 | 176275 | 3.00 [2.92, 3.08] |
| Patel, 2018 | AUS | 121 | 3944 | 3.07 [2.57, 3.65] |
| Valero−Masa, 2017 | ESP | 41 | 1111 | 3.69 [2.73, 4.97] |
| Median prevalence | | | | 2.18 [1.44, 3.29] |
| Population−averaged prevalence | | | | 2.41 [1.56, 3.03] |

$Tau^2 = 0.23$   $I^2 = 99.7\%$

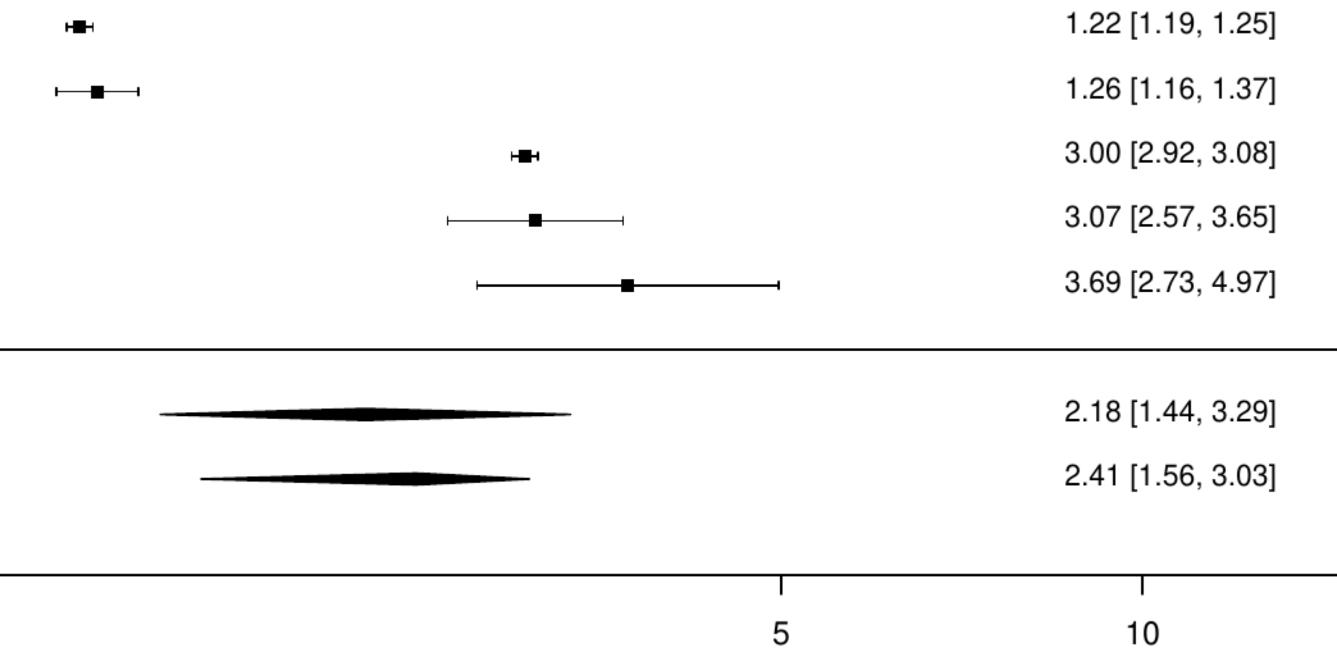

**Figure S14. Pooled prevalence of any cancer in acute myocardial infarction.**

| Author(s) and Year | Country | Events | N | Prevalence [95% CI] |
|---|---|---|---|---|
| Ostrominski JW, 2022 | IND | 7 | 3523 | 0.20 [ 0.09, 0.42] |
| Shi W, 2021 | CHN | 301 | 24001 | 1.25 [ 1.12, 1.40] |
| De Luca, 2017 | ITA | 15 | 923 | 1.63 [ 0.98, 2.68] |
| Kuehnemund L, 2021 | GER | 16386 | 875735 | 1.87 [ 1.84, 1.90] |
| Vallabhajosyula S, 2020 | USA | 3242 | 163071 | 1.99 [ 1.92, 2.06] |
| Hall M, 2024 | UK | 8667 | 433361 | 2.00 [ 1.96, 2.04] |
| Hagen TP, 2015 | NOR | 229 | 10522 | 2.18 [ 1.91, 2.47] |
| Erol MK, 2020 | TUR | 54 | 1930 | 2.80 [ 2.15, 3.64] |
| Bucholz EM, 2016 | USA | 1670 | 57574 | 2.90 [ 2.77, 3.04] |
| Kampfer J, 2017 | Multinational | 31 | 1023 | 3.03 [ 2.14, 4.28] |
| Niknam BA, 2018 | USA | 14805 | 414715 | 3.57 [ 3.51, 3.63] |
| Di Domenicantonio R, 2016 | ITA | 130 | 3608 | 3.60 [ 3.04, 4.26] |
| Ferreira JA, 2021 | PRT | 125 | 3145 | 3.97 [ 3.35, 4.72] |
| Quintana, 2018 | SWE | 79 | 1984 | 3.98 [ 3.21, 4.94] |
| O'Neill DE, 2017 | CAN | 89 | 2105 | 4.23 [ 3.45, 5.18] |
| Zheng R, 2021 | JPN | 5852 | 138250 | 4.23 [ 4.13, 4.34] |
| Soldati S, 2021 | ITA | 576 | 13540 | 4.25 [ 3.93, 4.61] |
| Jackel M, 2021 | GER | 28 | 624 | 4.49 [ 3.12, 6.42] |
| Yeh JS, 2021 | TW | 385 | 8552 | 4.50 [ 4.08, 4.96] |
| Williams MJA, 2019 | NZL | 374 | 8305 | 4.50 [ 4.08, 4.97] |
| Tran, 2018 | CAN | 2269 | 41210 | 5.51 [ 5.29, 5.73] |
| Kirchberger Inge, 2016 | GER | 106 | 1855 | 5.71 [ 4.75, 6.87] |
| Bianco M, 2021 | Multinational | 973 | 16653 | 5.84 [ 5.50, 6.21] |
| Kundi, 2019 | USA | 9737 | 166200 | 5.86 [ 5.75, 5.97] |
| Radovanovic D, 2017 | CHE | 852 | 14515 | 5.87 [ 5.50, 6.26] |
| Kim MS, 2021 | KOR | 15176 | 258291 | 5.88 [ 5.79, 5.97] |
| Wu VC, 2018 | TW | 11005 | 177058 | 6.22 [ 6.10, 6.33] |
| Akioyamen LE, 2023 | CAN | 3389 | 53841 | 6.29 [ 6.09, 6.50] |
| Ederhy S, 2019 | FRA | 246 | 3664 | 6.71 [ 5.95, 7.57] |
| Iqbal MB, 2016 | CAN | 771 | 11181 | 6.90 [ 6.44, 7.38] |
| Alsawas M, 2019 | USA | 1219 | 15777 | 7.73 [ 7.32, 8.15] |
| Araujo C, 2019 | PRT | 60 | 771 | 7.78 [ 6.09, 9.90] |
| Graham GN, 2018 | USA | 501 | 6402 | 7.83 [ 7.19, 8.51] |
| Case BC, 2020 | USA | 60127 | 748463 | 8.03 [ 7.97, 8.10] |
| Glynn L, 2024 | SWE | 24140 | 294018 | 8.21 [ 8.11, 8.31] |
| Watanabe H, 2015 | JPN | 291 | 3536 | 8.23 [ 7.37, 9.18] |
| Arai R, 2024 | JPN | 292 | 3411 | 8.56 [ 7.67, 9.55] |
| Tripathi B, 2021 | USA | 1891 | 21738 | 8.70 [ 8.33, 9.08] |
| Shroff GR, 2015 | USA | 3790 | 43168 | 8.78 [ 8.52, 9.05] |
| Puymirat E, 2017 | FRA | 289 | 3079 | 9.39 [ 8.41, 10.47] |
| Belle Loïc, 2017 | FRA | 361 | 3813 | 9.47 [ 8.58, 10.44] |
| Oberprieler NG, 2022 | NOR | 4251 | 43750 | 9.72 [ 9.44, 10.00] |
| Obayashi Y, 2021 | JPN | 690 | 6228 | 11.08 [10.32, 11.88] |
| Hamood H, 2016 | ISR | 518 | 4655 | 11.13 [10.26, 12.06] |
| Di Martino M, 2015 | ITA | 96 | 778 | 12.34 [10.21, 14.84] |
| Tangri N, 2017 | CAN | 1060 | 8493 | 12.48 [11.79, 13.20] |
| Christensen DM, 2022 | DEN | 14842 | 116481 | 12.74 [12.55, 12.93] |
| Kerola AM, 2022 | FIN | 9930 | 76606 | 12.96 [12.73, 13.20] |
| Takeuchi T, 2021 | JPN | 462 | 3499 | 13.20 [12.12, 14.37] |
| Bagai, 2019 | USA | 22434 | 134716 | 16.65 [16.45, 16.85] |
| Median prevalence | Tau² = 0.57 I² = 100% | | | 5.42 [ 4.44, 6.61] |
| Population−averaged prevalence | | | | 6.75 [ 5.78, 7.81] |

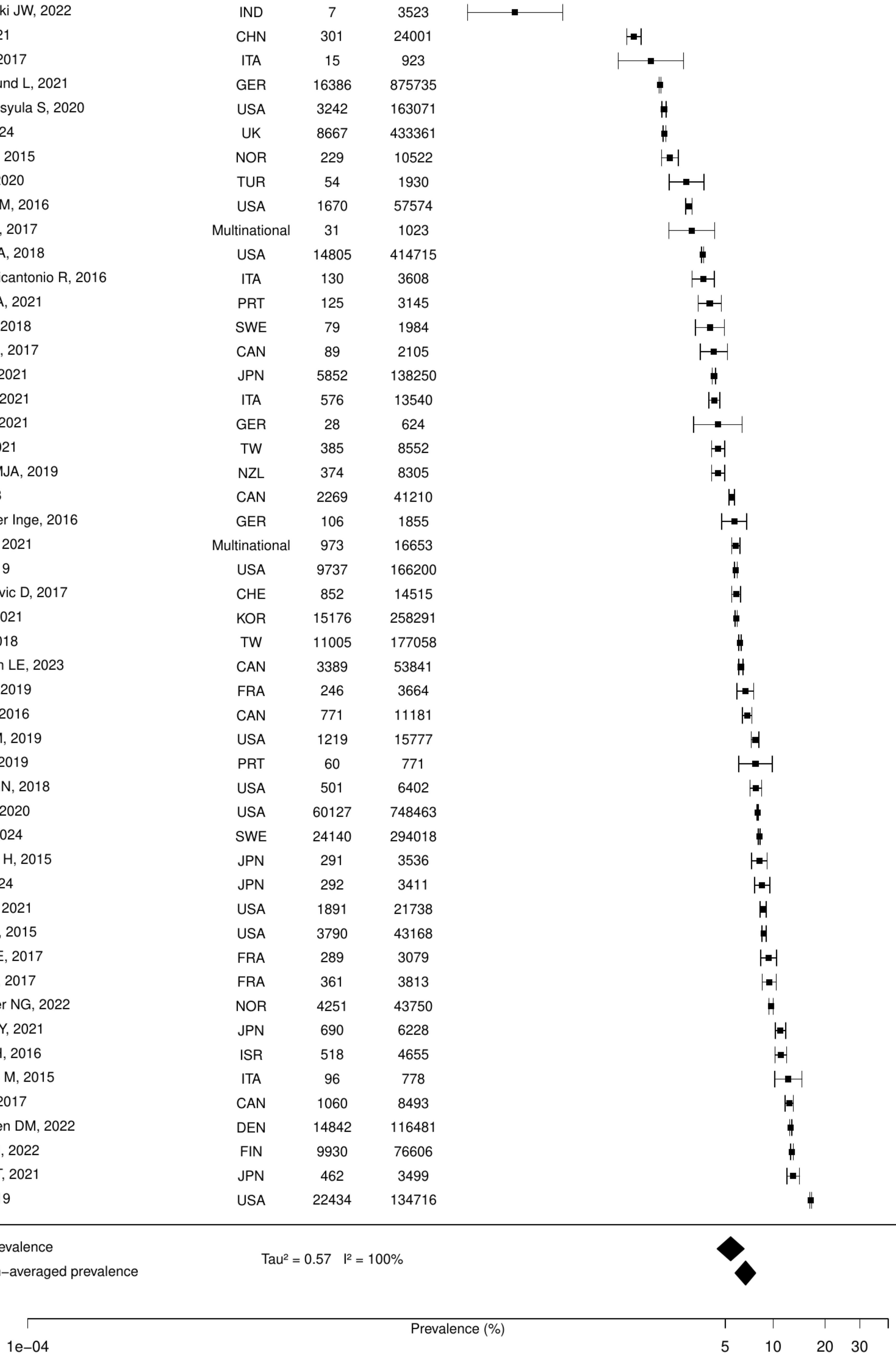

**Figure S15. Pooled prevalence of previous cancer in acute myocardial infarction.**

| Author(s) and Year | Country | Events | N | Prevalence [95% CI] |
|---|---|---|---|---|
| Kang, 2020 | KOR | 1459 | 108232 | 1.35 [ 1.28, 1.42] |
| Jamaly S, 2021 | SWE | 590 | 25384 | 2.32 [ 2.15, 2.52] |
| Choi IJ, 2021 | KOR | 38 | 1384 | 2.75 [ 2.00, 3.75] |
| Puymirat E, 2016 | FRA | 161 | 2679 | 6.01 [ 5.17, 6.98] |
| Shore S, 2016 | USA | 240 | 3283 | 7.31 [ 6.47, 8.25] |
| Abu−Assi Emad, 2016 | ESP | 57 | 767 | 7.43 [ 5.78, 9.51] |
| Danchin, 2020 | FRA | 398 | 5147 | 7.73 [ 7.03, 8.49] |
| Teece L, 2023 | UK | 42187 | 512388 | 8.23 [ 8.16, 8.31] |
| Rohla M, 2024 | CHE | 179 | 1996 | 8.97 [ 7.79, 10.30] |
| Velders MA, 2020 | SWE | 16237 | 175146 | 9.27 [ 9.14, 9.41] |
| Jacobs JA, 2019 | USA | 58 | 609 | 9.52 [ 7.43, 12.12] |
| Wang F, 2016 | USA | 263 | 2346 | 11.21 [10.00, 12.55] |
| Palomäki A, 2021 | FIN | 7263 | 60446 | 12.02 [11.76, 12.28] |
| Sanchez F, 2021 | ITA | 146 | 1086 | 13.44 [11.54, 15.60] |
| Median prevalence | | | | 6.60 [ 4.69, 9.21] |
| Population−averaged prevalence | | | | 7.85 [ 5.98, 9.65] |

$Tau^2 = 0.47$   $I^2 = 99.9\%$

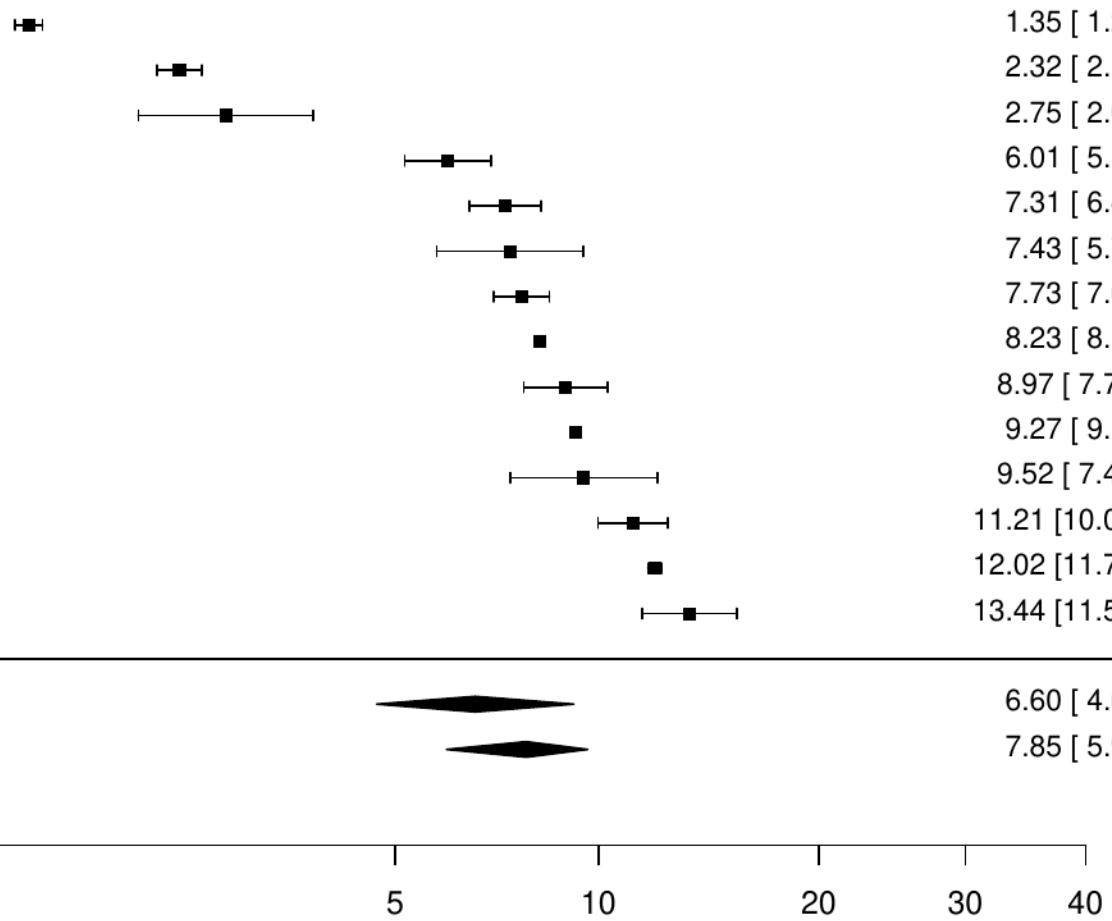

**Figure S16. Pooled prevalence of blood cancer in acute myocardial infarction.**

| Author(s) and Year | Country | Events | N | Prevalence [95% CI] |
|---|---|---|---|---|
| Lobo, 2017 | PRT | 619 | 123442 | 0.50 [0.46, 0.54] |
| Montoy JCC, 2021 | USA | 1800 | 293589 | 0.61 [0.59, 0.64] |
| McCarthy CP, 2021 | USA | 1917 | 254422 | 0.75 [0.72, 0.79] |
| Khera R, 2017 | USA | 5739 | 478247 | 1.20 [1.17, 1.23] |
| Median prevalence | | | | 0.73 [0.53, 1.00] |
| Population−averaged prevalence | | | | 0.76 [0.56, 1.03] |

Tau² = 0.11   I² = 99.5%

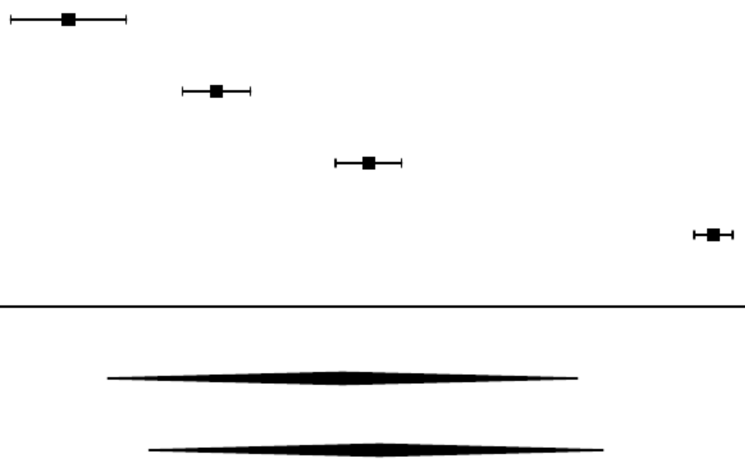

**Figure S17. Pooled prevalence of metastatic cancer in acute myocardial infarction.**

| Author(s) and Year | Country | Events | N | Prevalence [95% CI] |
|---|---|---|---|---|
| Vallabhajosyula S, 2020 | USA | 49 | 163071 | 0.03 [0.02, 0.04] |
| Kerola, 2022 | FIN | 18 | 8934 | 0.20 [0.13, 0.32] |
| Kyto V, 2022 | FIN | 142 | 40654 | 0.35 [0.30, 0.41] |
| Lobo, 2017 | PRT | 619 | 123442 | 0.50 [0.46, 0.54] |
| Williams MJA, 2019 | NZL | 71 | 8305 | 0.85 [0.68, 1.08] |
| Yasaitis LC, 2020 | CAN | 1237 | 112484 | 1.10 [1.04, 1.16] |
| Spoon DB, 2015 | USA | 74 | 6636 | 1.12 [0.89, 1.40] |
| Tran, 2018 | CAN | 556 | 41210 | 1.35 [1.24, 1.47] |
| McCarthy CP, 2021 | USA | 4342 | 254422 | 1.71 [1.66, 1.76] |
| Montoy JCC, 2021 | USA | 7404 | 293589 | 2.52 [2.47, 2.58] |
| Wang Y, 2020 | USA | 48910 | 884931 | 5.53 [5.48, 5.57] |
| Median prevalence | | | | 0.74 [0.34, 1.64] |
| Population−averaged prevalence | | | | 1.69 [0.77, 3.04] |

Tau² = 1.81   I² = 100%

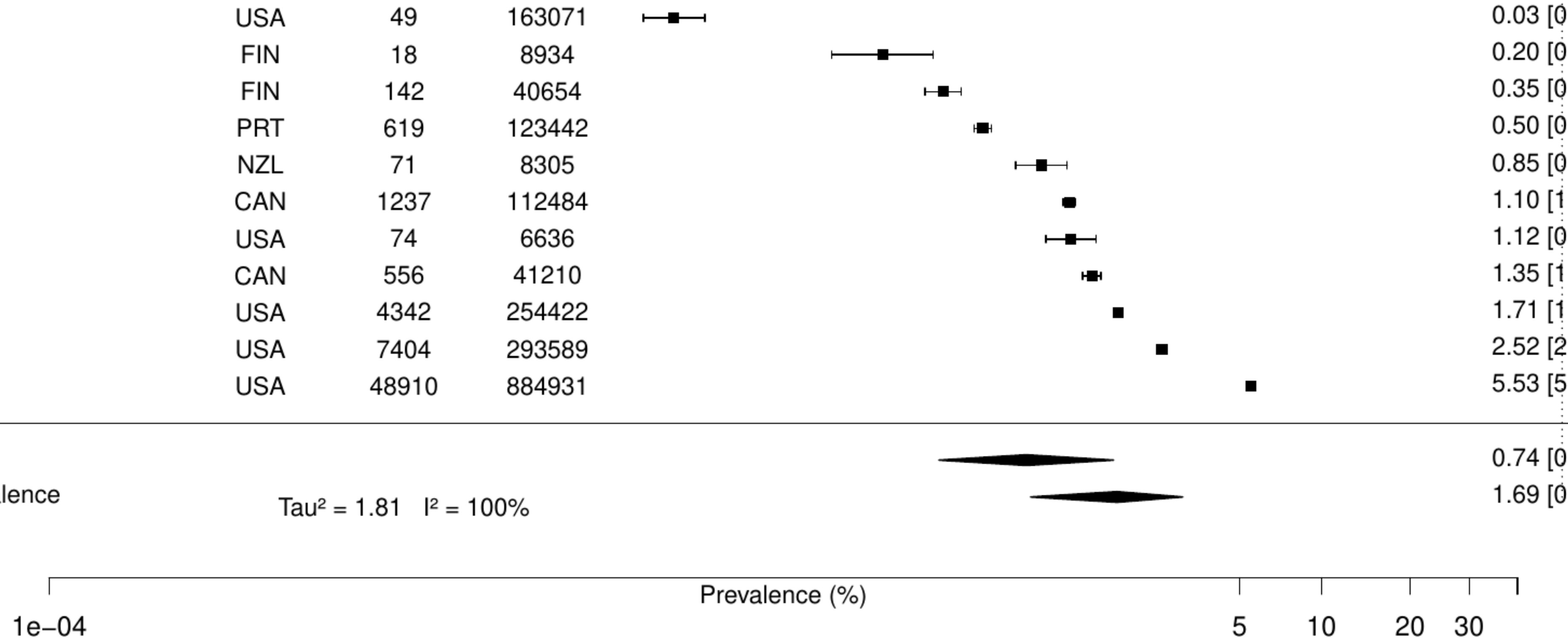

**Figure S18. Pooled prevalence of solid cancer in acute myocardial infarction.**

| Author(s) and Year | Country | Events | N | Prevalence [95% CI] |
|---|---|---|---|---|
| Montoy JCC, 2021 | USA | 3054 | 293589 | 1.04 [ 1.00, 1.08] |
| McCarthy CP, 2021 | USA | 5204 | 254422 | 2.05 [ 1.99, 2.10] |
| Khera R, 2017 | USA | 49456 | 478247 | 10.34 [10.26, 10.43] |
| Median prevalence | | | | 2.85 [ 0.93, 8.41] |
| Population−averaged prevalence | Tau² = 1.01 I² = 100% | | | 4.34 [ 1.37, 7.77] |

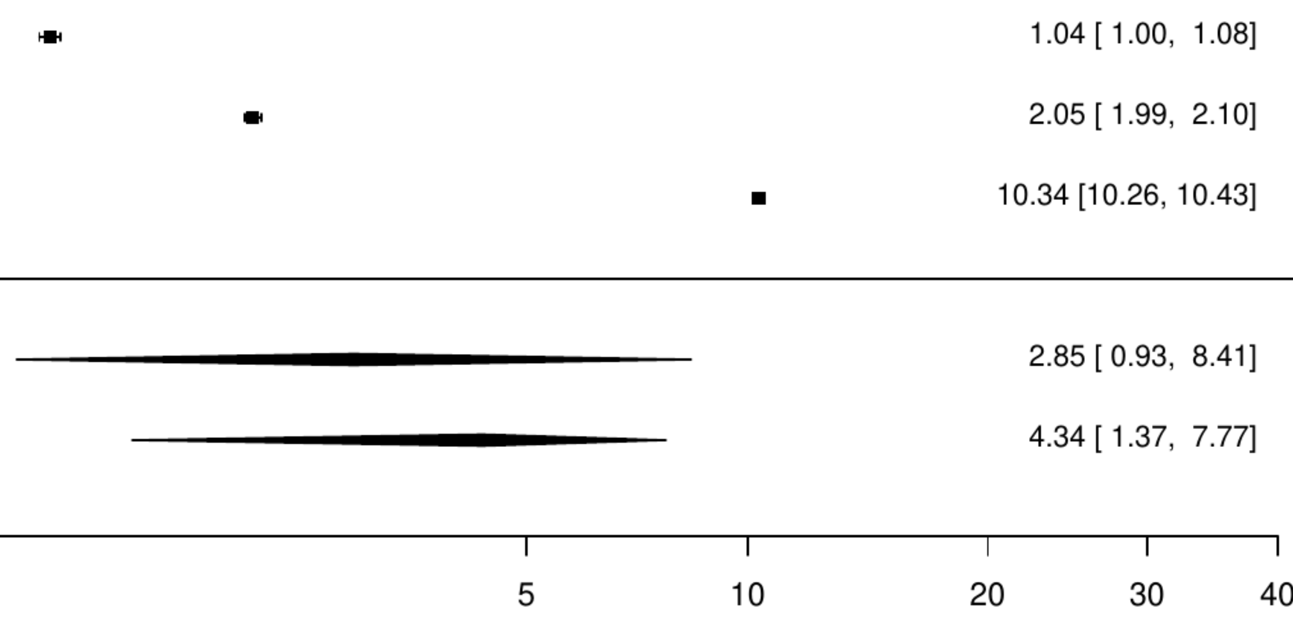

**Figure S19. Pooled prevalence of any cancer in ST-elevation myocardial infarction.**

| Author(s) and Year | Country | Events | N | Prevalence [95% CI] |
|---|---|---|---|---|
| Gupta MD, 2024 | IND | 8 | 5335 | 0.15 [0.08, 0.30] |
| Kaul P, 2016 | CAN | 972 | 66008 | 1.47 [1.38, 1.57] |
| De Luca, 2017 | ITA | 15 | 923 | 1.63 [0.98, 2.68] |
| Di Domenicantonio R, 2016 | ITA | 130 | 3608 | 3.60 [3.04, 4.26] |
| Ferreira JA, 2021 | PRT | 125 | 3145 | 3.97 [3.35, 4.72] |
| Figtree GA, 2021 | SWE | 2970 | 62048 | 4.79 [4.62, 4.96] |
| Radovanovic D, 2016 | CHE | 1000 | 19665 | 5.09 [4.79, 5.40] |
| Brunet T, 2021 | FRA | 297 | 3907 | 7.60 [6.81, 8.48] |
| Rymer JA, 2024 | USA | 13087 | 162822 | 8.04 [7.91, 8.17] |
| Yamashita Y, 2017 | JPN | 318 | 3942 | 8.07 [7.26, 8.96] |
| Rorth R, 2016 | DEN | 885 | 9857 | 8.98 [8.43, 9.56] |
| Median prevalence | | | | 3.44 [1.79, 6.50] |
| Population−averaged prevalence | | | | 5.62 [3.66, 7.38] |

Tau² = 1.26   I² = 99.9%

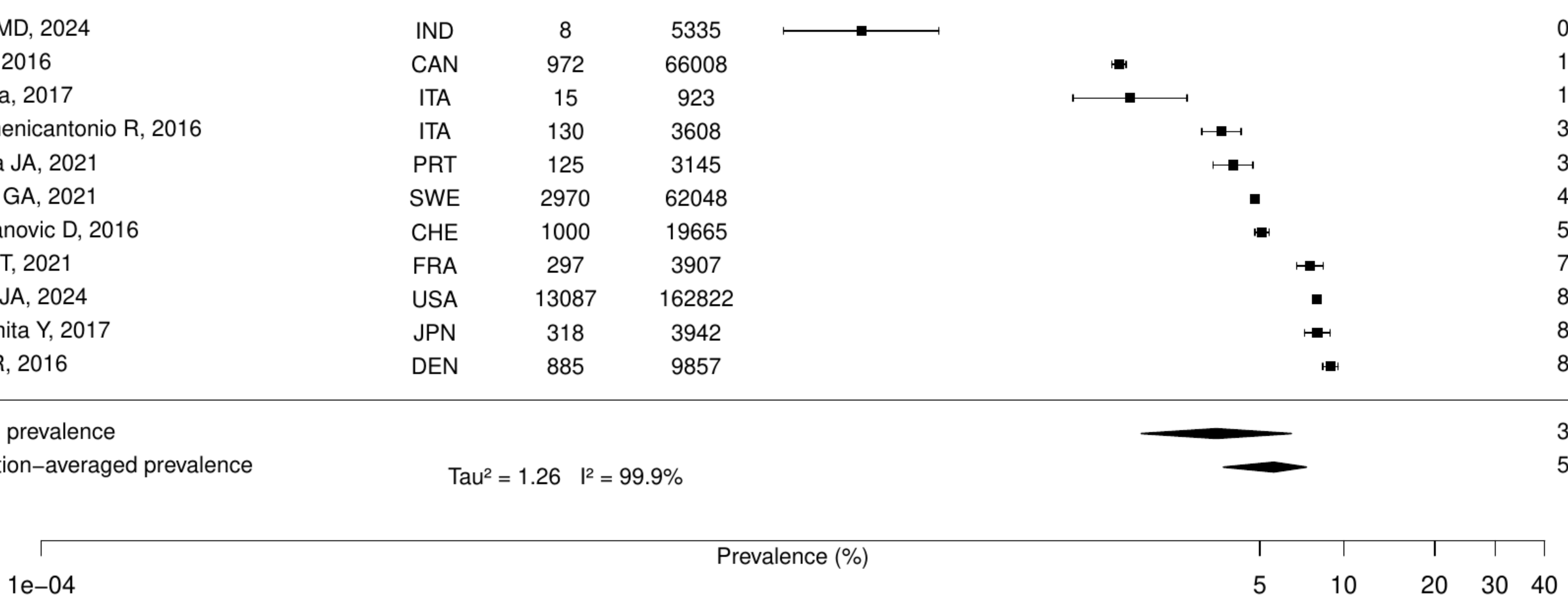

**Figure S20. Pooled prevalence of previous cancer in ST-elevation myocardial infarction.**

| Author(s) and Year | Country | Events | N | Prevalence [95% CI] |
|---|---|---|---|---|
| Szummer K, 2017 | SWE | 1957 | 105674 | 1.85 [ 1.77, 1.93] |
| Abu−Assi Emad, 2016 | ESP | 57 | 767 | 7.43 [ 5.78, 9.51] |
| Bouisset F, 2021 | FRA | 491 | 6243 | 7.86 [ 7.22, 8.56] |
| Rohla M, 2024 | CHE | 179 | 1996 | 8.97 [ 7.79, 10.30] |
| Jacobs JA, 2019 | USA | 58 | 609 | 9.52 [ 7.43, 12.12] |
| Wang F, 2016 | USA | 263 | 2346 | 11.21 [10.00, 12.55] |
| Sanchez F, 2021 | ITA | 146 | 1086 | 13.44 [11.54, 15.60] |
| Median prevalence | | | | 7.58 [ 4.83, 11.70] |
| Population−averaged prevalence | | | | 8.78 [ 6.10, 11.01] |

Tau² = 0.41   I² = 99.1%

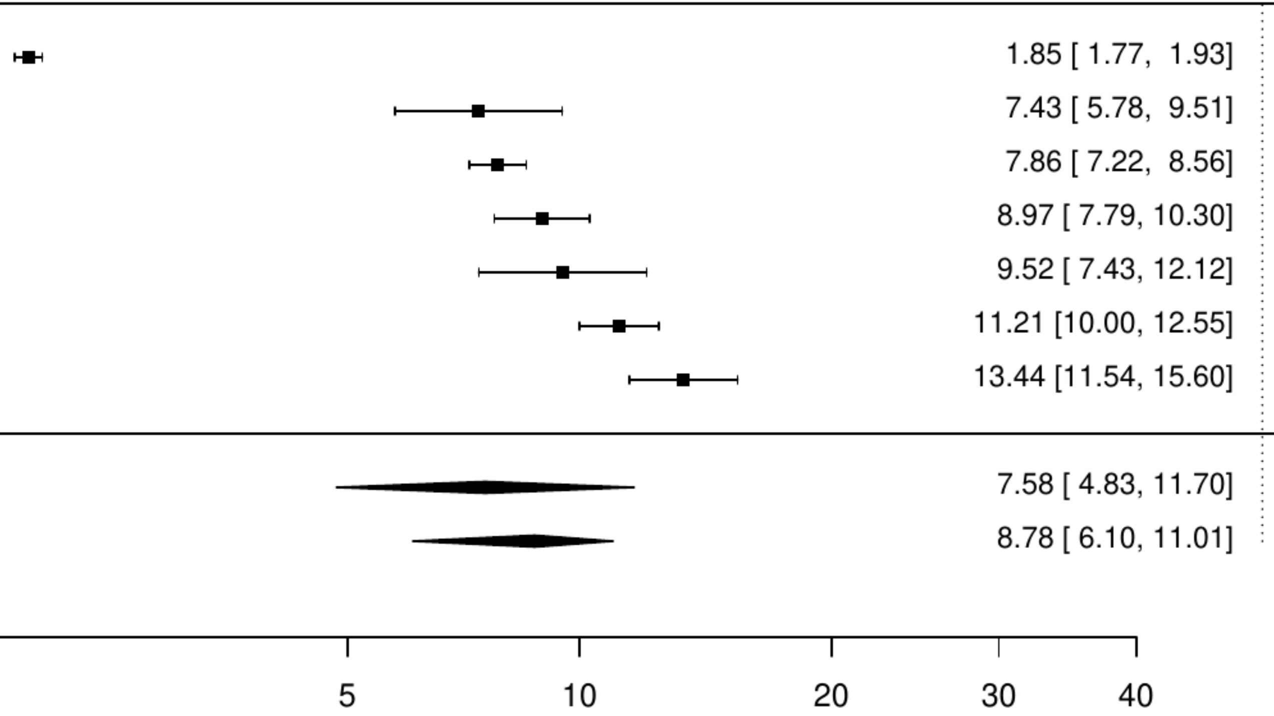

**Figure S21. Pooled prevalence of any cancer in nonST-elevation myocardial infarction.**

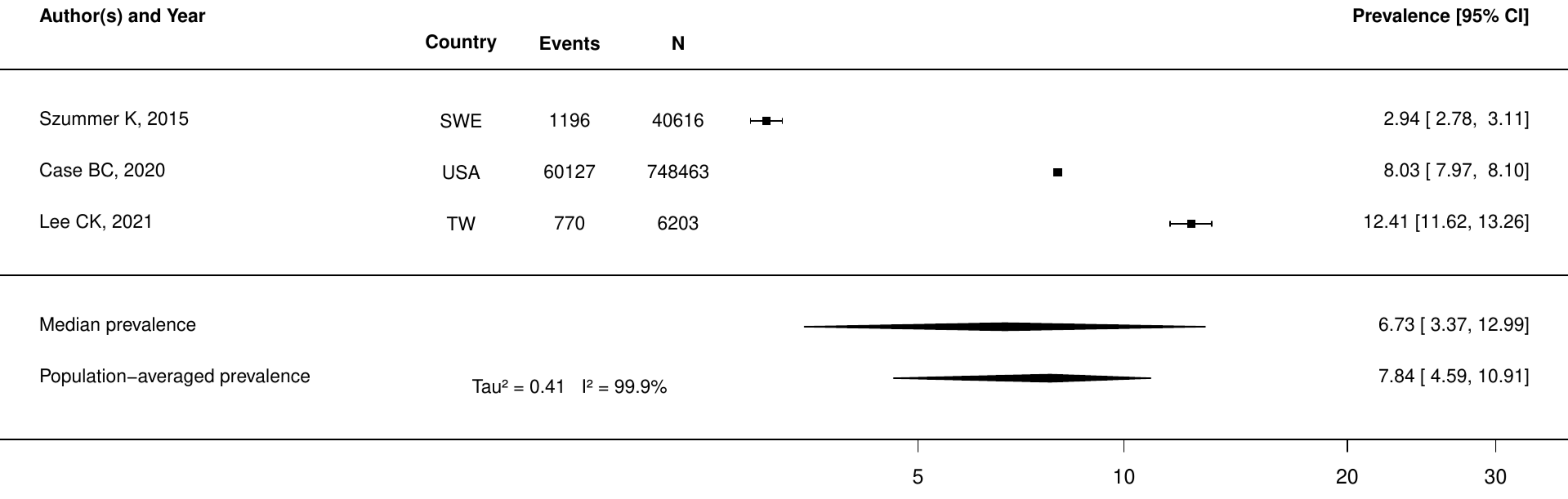

**Figure S22. Pooled prevalence of any cancer in nonST-elevation acute coronary syndrome.**

| Author(s) and Year | Country | Events | N | Prevalence [95% CI] |
|---|---|---|---|---|
| De Luca, 2017 | ITA | 14 | 832 | 1.68 [ 1.00, 2.82] |
| van den Broek WWA, 2024 | NLD | 45 | 1320 | 3.41 [ 2.55, 4.54] |
| Eggers, 2018 | SWE | 730 | 20652 | 3.53 [ 3.29, 3.80] |
| Shiraishi Y, 2016 | JPN | 130 | 3287 | 3.95 [ 3.34, 4.68] |
| Case BC, 2020 | USA | 60127 | 748463 | 8.03 [ 7.97, 8.10] |
| Sanchis J, 2019 | ESP | 155 | 1452 | 10.67 [ 9.19, 12.37] |
| Lee CK, 2021 | TW | 770 | 6203 | 12.41 [11.62, 13.26] |
| Median prevalence | | | | 5.21 [ 3.17, 8.44] |
| Population−averaged prevalence | | | | 6.27 [ 3.50, 9.23] |

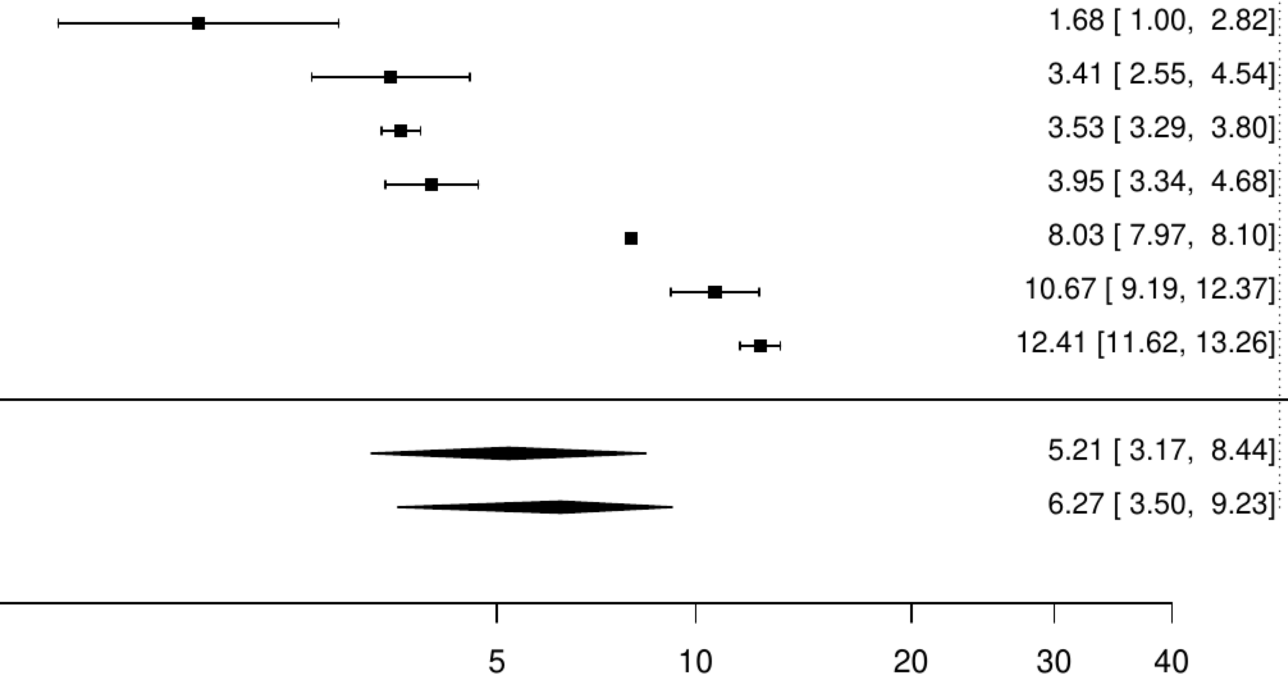

**Figure S23. Pooled prevalence of any cancer in stable angina.**

| Author(s) and Year | Country | Events | N | Prevalence [95% CI] |
|---|---|---|---|---|
| Szpakowski N, 2016 | CAN | 587 | 22917 | 2.56 [ 2.36, 2.77] |
| Christensen DM, 2023 | DEN | 5013 | 82108 | 6.11 [ 5.94, 6.27] |
| De Luca, 2018 | ITA | 331 | 5070 | 6.53 [ 5.88, 7.24] |
| Shiomi H, 2021 | JPN | 2573 | 24951 | 10.31 [ 9.94, 10.70] |
| Iqbal MB, 2022 | CAN | 903 | 8436 | 10.70 [10.06, 11.38] |
| Blin P, 2021 | FRA | 47066 | 258260 | 18.22 [18.08, 18.37] |
| Median prevalence | | | | 7.80 [ 4.75, 12.57] |
| Population−averaged prevalence | | | | 9.10 [ 5.12, 13.00] |

Tau² = 0.44   I² = 99.9%

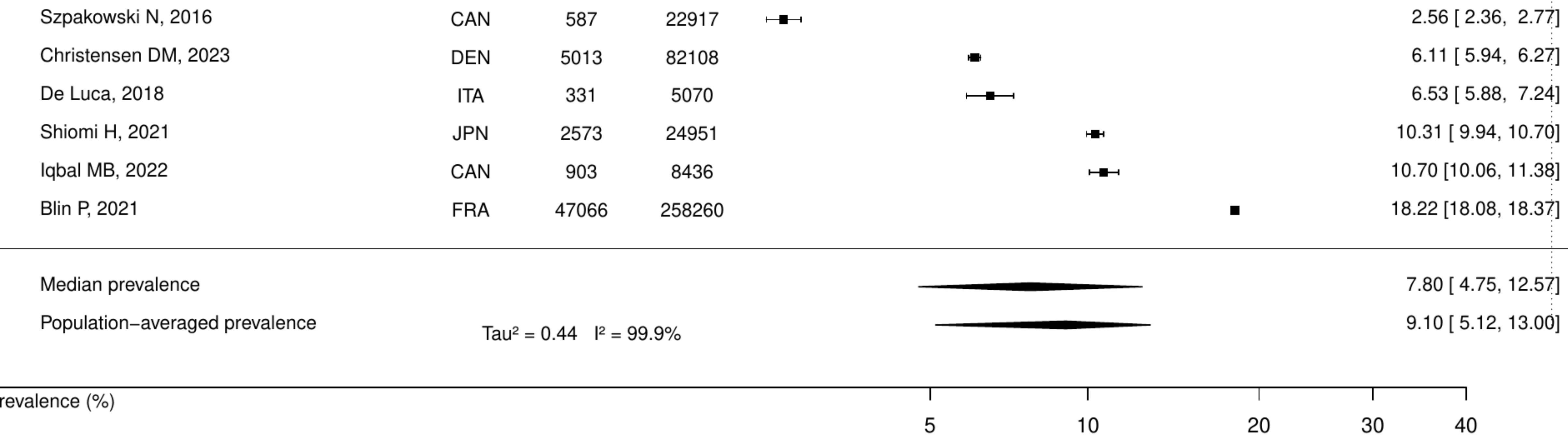

**Figure S24. Pooled prevalence of any cancer in patients with history of myocardial infarction.**

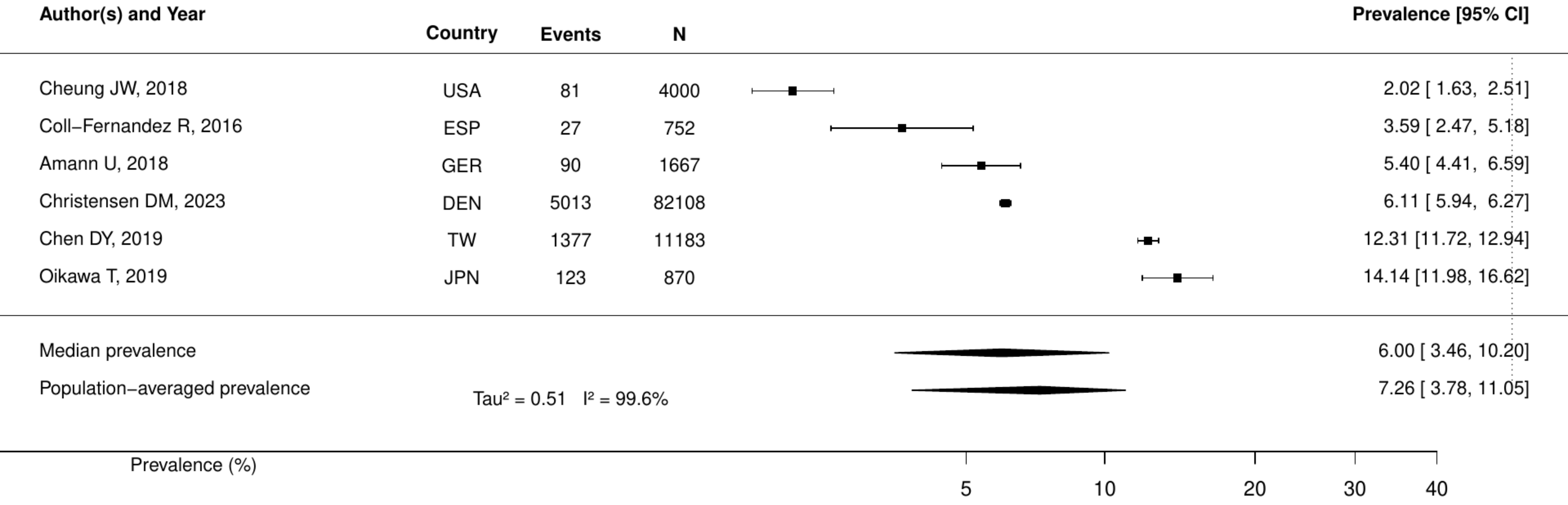

**Figure S25. Pooled prevalence of active cancer in percutaneous coronary intervention.**

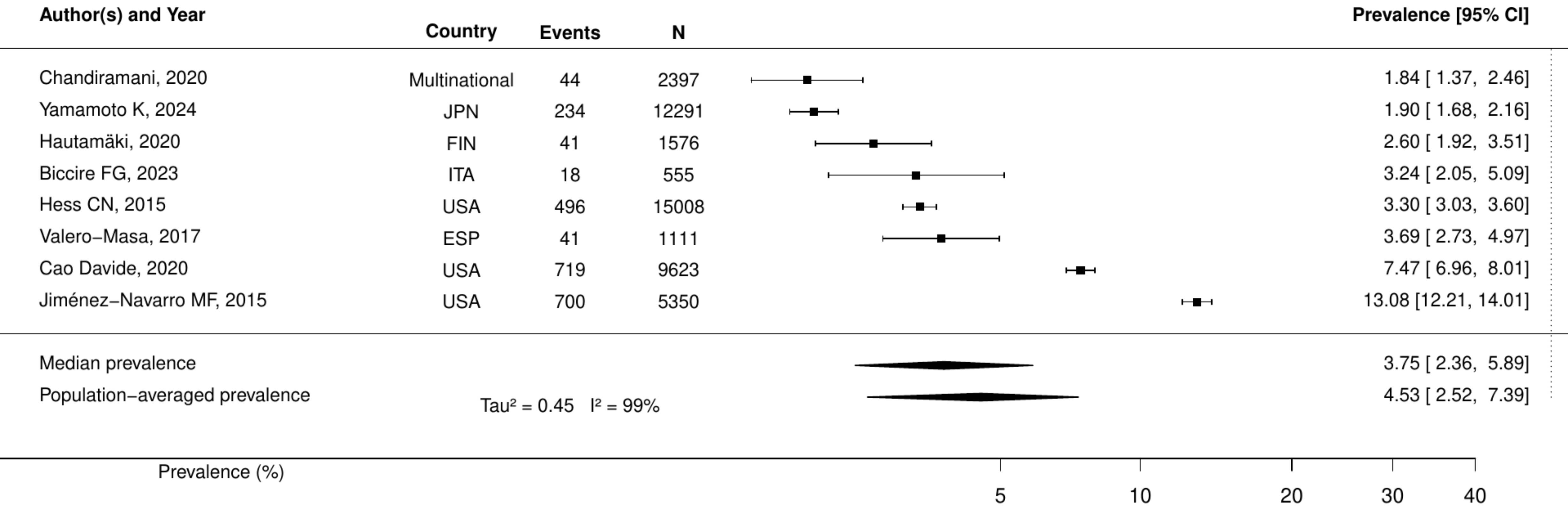

**Figure S26. Pooled prevalence of any cancer in percutaneous coronary intervention.**

| Author(s) and Year | Country | Events | N | Prevalence [95% CI] |
|---|---|---|---|---|
| Wong CCY, 2021 | AUS | 43 | 10304 | 0.42 [ 0.31, 0.56] |
| Koul S, 2018 | SWE | 140 | 7433 | 1.88 [ 1.60, 2.22] |
| Lee, 2020 | KOR | 1749 | 87980 | 1.99 [ 1.90, 2.08] |
| Smilowitz, 2019 | USA | 4482 | 221379 | 2.02 [ 1.97, 2.08] |
| Seki T, 2019 | JPN | 557 | 21409 | 2.60 [ 2.40, 2.82] |
| Terada T, 2017 | CAN | 1018 | 30258 | 3.36 [ 3.17, 3.57] |
| Di Domenicantonio R, 2016 | ITA | 130 | 3608 | 3.60 [ 3.04, 4.26] |
| Sawano M, 2017 | JPN | 487 | 13024 | 3.74 [ 3.43, 4.08] |
| Jackel M, 2021 | GER | 28 | 624 | 4.49 [ 3.12, 6.42] |
| Shoji S, 2020 | JPN | 118 | 2559 | 4.61 [ 3.86, 5.50] |
| Wu VC, 2021 | TW | 8687 | 170775 | 5.09 [ 4.98, 5.19] |
| Rasia, 2017 | ITA | 124 | 2404 | 5.16 [ 4.34, 6.12] |
| D'Ascenzo, 2020 | Multinational | 1126 | 19826 | 5.68 [ 5.37, 6.01] |
| Nakamura M, 2020 | JPN | 367 | 6267 | 5.86 [ 5.30, 6.47] |
| Wanha W, 2017 | POL | 117 | 1916 | 6.11 [ 5.12, 7.27] |
| Mackay MH, 2017 | CAN | 2898 | 41792 | 6.93 [ 6.69, 7.18] |
| Amin AP, 2020 | USA | 30620 | 434172 | 7.05 [ 6.98, 7.13] |
| Saia F, 2016 | ITA | 2777 | 39362 | 7.06 [ 6.81, 7.31] |
| Olafiranye O, 2015 | USA | 241 | 3326 | 7.25 [ 6.41, 8.18] |
| Brunet T, 2021 | FRA | 297 | 3907 | 7.60 [ 6.81, 8.48] |
| Madsen, 2021 | DEN | 616 | 7944 | 7.75 [ 7.19, 8.36] |
| Cilia, 2019 | USA | 850 | 10756 | 7.90 [ 7.41, 8.43] |
| Rymer JA, 2024 | USA | 13087 | 162822 | 8.04 [ 7.91, 8.17] |
| Kazi DS, 2015 | USA | 2735 | 32906 | 8.31 [ 8.02, 8.61] |
| Natsuaki, 2019 | JPN | 1185 | 13058 | 9.07 [ 8.59, 9.58] |
| Ohya, 2017 | JPN | 163 | 1607 | 10.14 [ 8.76, 11.72] |
| Spirito A, 2021 | CHE | 1810 | 16821 | 10.76 [10.30, 11.24] |
| Skalsky K, 2021 | ISR | 1667 | 15153 | 11.00 [10.51, 11.51] |
| Yamamoto K, 2024 | JPN | 1544 | 12291 | 12.56 [11.99, 13.16] |
| Raphael CE, 2016 | USA | 2215 | 17165 | 12.90 [12.41, 13.41] |
| Takeuchi T, 2021 | JPN | 462 | 3499 | 13.20 [12.12, 14.37] |
| Rossini R, 2017 | ITA | 269 | 1082 | 24.86 [22.38, 27.52] |
| Median prevalence | | | | 5.83 [ 4.50, 7.52] |
| Population−averaged prevalence | Tau² = 0.61 I² = 99.9% | | | 7.34 [ 5.85, 8.97] |

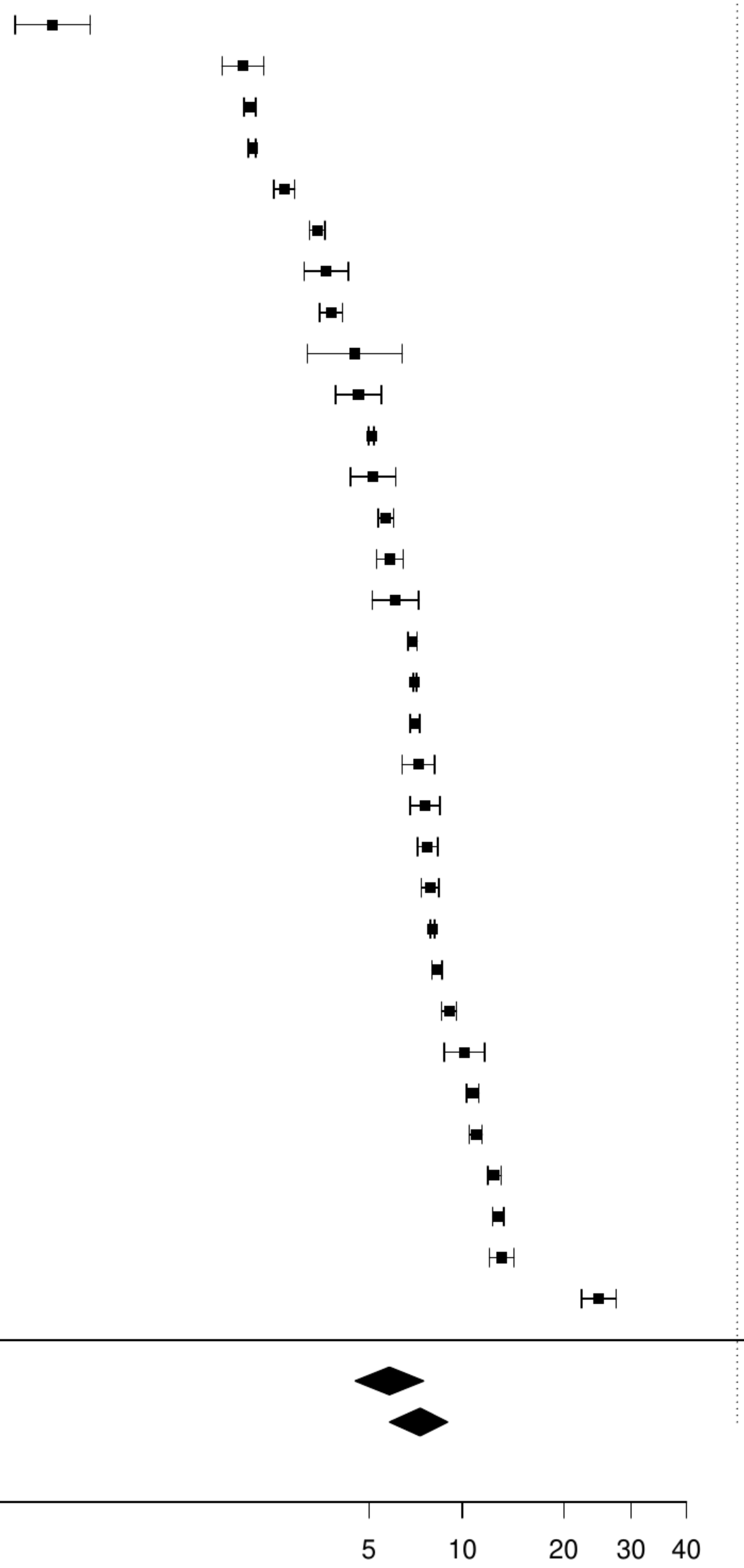

**Figure S27. Pooled prevalence of previous cancer in percutaneous coronary intervention.**

| Author(s) and Year | Country | Events | N | Prevalence [95% CI] |
|---|---|---|---|---|
| Jamaly S, 2021 | SWE | 590 | 25384 | 2.32 [ 2.15, 2.52] |
| Choi IJ, 2021 | KOR | 38 | 1384 | 2.75 [ 2.00, 3.75] |
| Rohla M, 2015 | AUT | 152 | 2890 | 5.26 [ 4.50, 6.14] |
| Rohla M, 2024 | CHE | 179 | 1996 | 8.97 [ 7.79, 10.30] |
| Nakatsuma, 2018 | JPN | 1109 | 12180 | 9.11 [ 8.61, 9.63] |
| Perl L, 2020 | ISR | 2696 | 25690 | 10.49 [10.13, 10.88] |
| Biccire FG, 2023 | ITA | 59 | 555 | 10.63 [ 8.33, 13.48] |
| Yamaji K, 2018 | CHE | 815 | 7640 | 10.67 [ 9.99, 11.38] |
| Wang F, 2016 | USA | 263 | 2346 | 11.21 [10.00, 12.55] |
| de la Torre Hernandez Jose M, 2021 | ESP | 161 | 1249 | 12.89 [11.14, 14.87] |
| Gajanana D, 2020 | USA | 975 | 7499 | 13.00 [12.26, 13.78] |
| Hsieh CF, 2015 | TW | 6821 | 9805 | 69.57 [68.65, 70.47] |
| Median prevalence | | | | 10.08 [ 5.71, 17.17] |
| Population−averaged prevalence | | | | 13.93 [ 7.25, 24.76] |

$Tau^2 = 1.18$   $I^2 = 99.9\%$

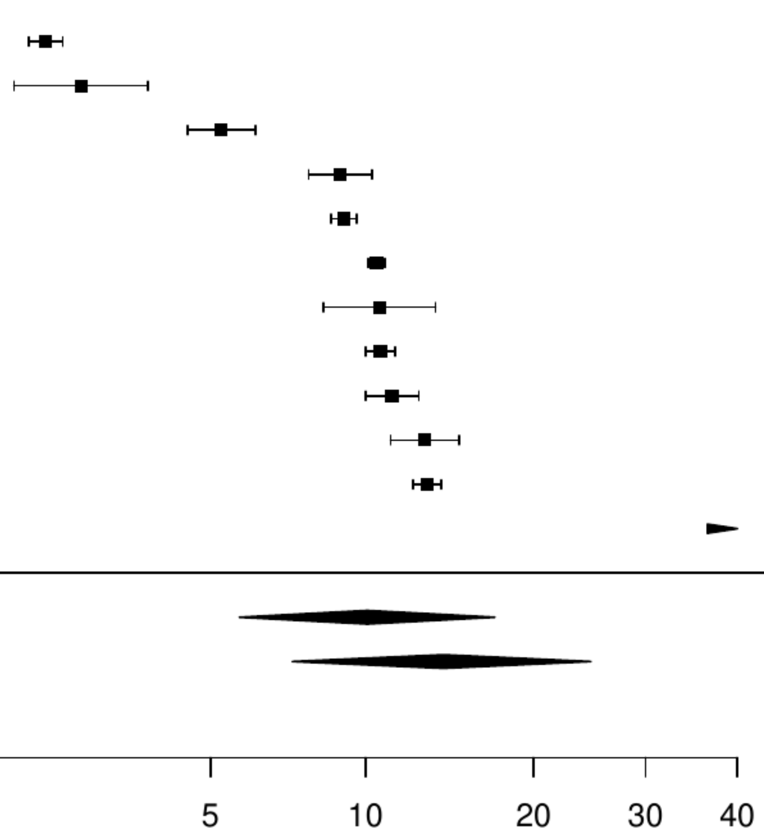


Prevalence (%)

### Figure S28. Pooled prevalence of metastatic cancer in percutaneous coronary intervention.

| Author(s) and Year | Country | Events | N | Prevalence [95% CI] |
|---|---|---|---|---|
| Seki T, 2019 | JPN | 25 | 21409 | 0.12 [0.08, 0.17] |
| Lai CL, 2015 | TW | 16 | 5587 | 0.29 [0.18, 0.47] |
| Hautamäki, 2020 | FIN | 13 | 1576 | 0.82 [0.48, 1.42] |
| Singh, 2022 | USA | 158 | 15128 | 1.04 [0.89, 1.22] |
| Median prevalence | | | | 0.41 [0.17, 0.97] |
| Population−averaged prevalence | | | | 0.58 [0.19, 1.02] |

$Tau^2 = 0.75$   $I^2 = 95.7\%$

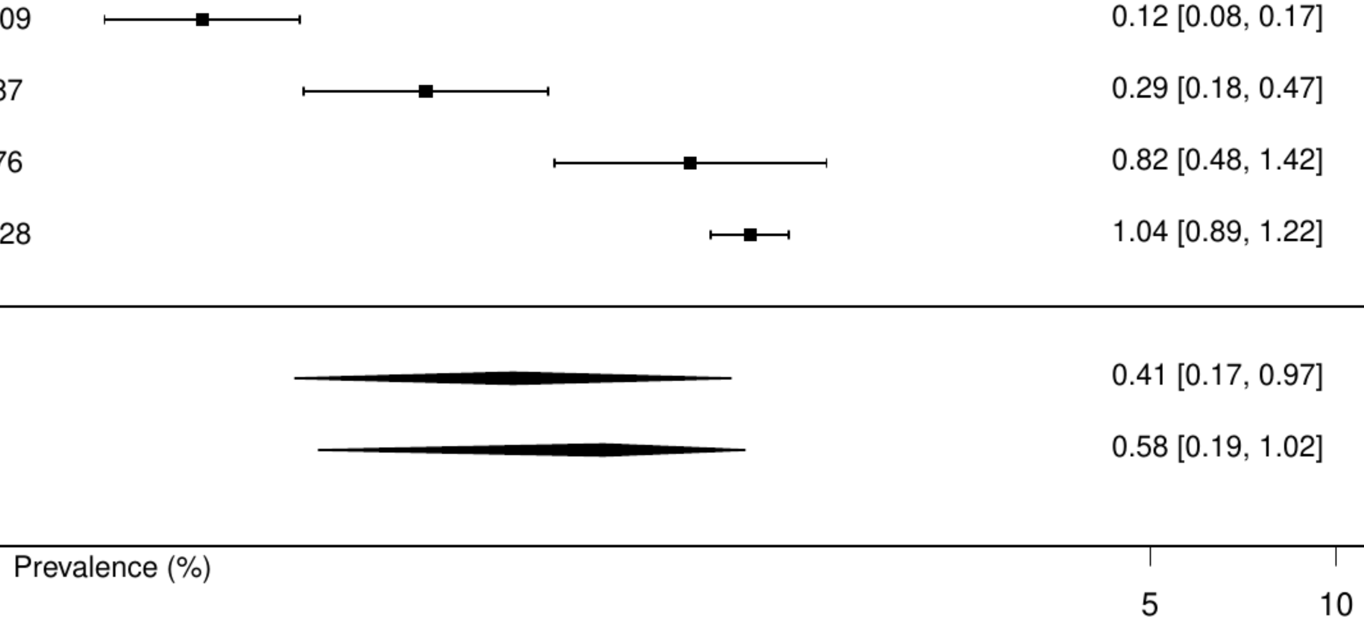

**Figure S29. Pooled prevalence of any cancer in coronary artery bypass surgery**

| Author(s) and Year | Country | Events | N | Prevalence [95% CI] |
|---|---|---|---|---|
| Atiya M, 2021 | USA | 1849 | 201116 | 0.92 [ 0.88, 0.96] |
| Fowler AJ, 2022 | UK | 893 | 43063 | 2.07 [ 1.94, 2.21] |
| Terada T, 2017 | CAN | 163 | 7560 | 2.16 [ 1.85, 2.51] |
| Goldstone, 2018 | USA | 2116 | 59432 | 3.56 [ 3.41, 3.71] |
| Johnson SM, 2016 | CAN | 207 | 5578 | 3.71 [ 3.25, 4.24] |
| Chen JJ, 2017 | TW | 42 | 884 | 4.75 [ 3.53, 6.37] |
| Patel A, 2022 | AUS | 3170 | 60953 | 5.20 [ 5.03, 5.38] |
| Thorsteinsson, 2016 | DEN | 2434 | 38830 | 6.27 [ 6.03, 6.51] |
| Nielsen S, 2019 | SWE | 7734 | 110742 | 6.98 [ 6.84, 7.14] |
| Kytö V, 2021 | FIN | 1104 | 14681 | 7.52 [ 7.10, 7.96] |
| Marui A, 2015 | JPN | 198 | 1877 | 10.55 [ 9.24, 12.02] |
| Chikwe, 2018 | USA | 2747 | 22245 | 12.35 [11.92, 12.79] |
| Solomon J, 2021 | CAN | 65 | 500 | 13.00 [10.33, 16.24] |
| George LK, 2015 | USA | 928 | 5968 | 15.55 [14.65, 16.49] |
| Median prevalence | | | | 5.31 [ 3.52, 7.93] |
| Population−averaged prevalence | Tau² = 0.67 I² = 99.9% | | | 6.85 [ 4.48, 9.24] |

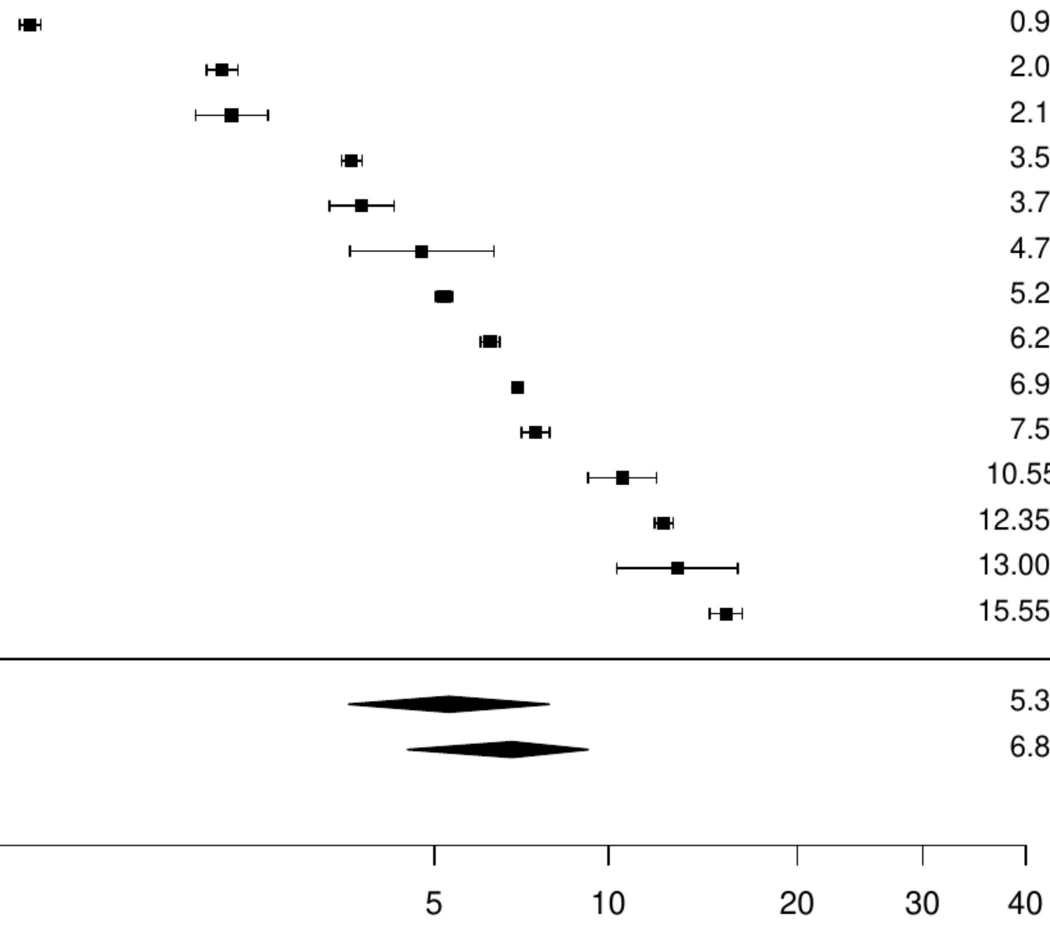

### Figure S30. Pooled prevalence of previous cancer in coronary artery bypass surgery

| Author(s) and Year | Country | Events | N | Prevalence [95% CI] |
|---|---|---|---|---|
| Alherbish A, 2017 | CAN | 88 | 6262 | 1.41 [ 1.14, 1.73] |
| Pu Aihua, 2017 | CAN | 849 | 20076 | 4.23 [ 3.96, 4.52] |
| Nystrom T, 2015 | SWE | 3039 | 53820 | 5.65 [ 5.45, 5.84] |
| Dalen M, 2024 | SWE | 461 | 3267 | 14.11 [12.96, 15.35] |
| Median prevalence | | | | 4.76 [ 2.09, 10.49] |
| Population−averaged prevalence | | | | 6.36 [ 2.83, 11.71] |

Tau² = 0.75 I² = 99.8%

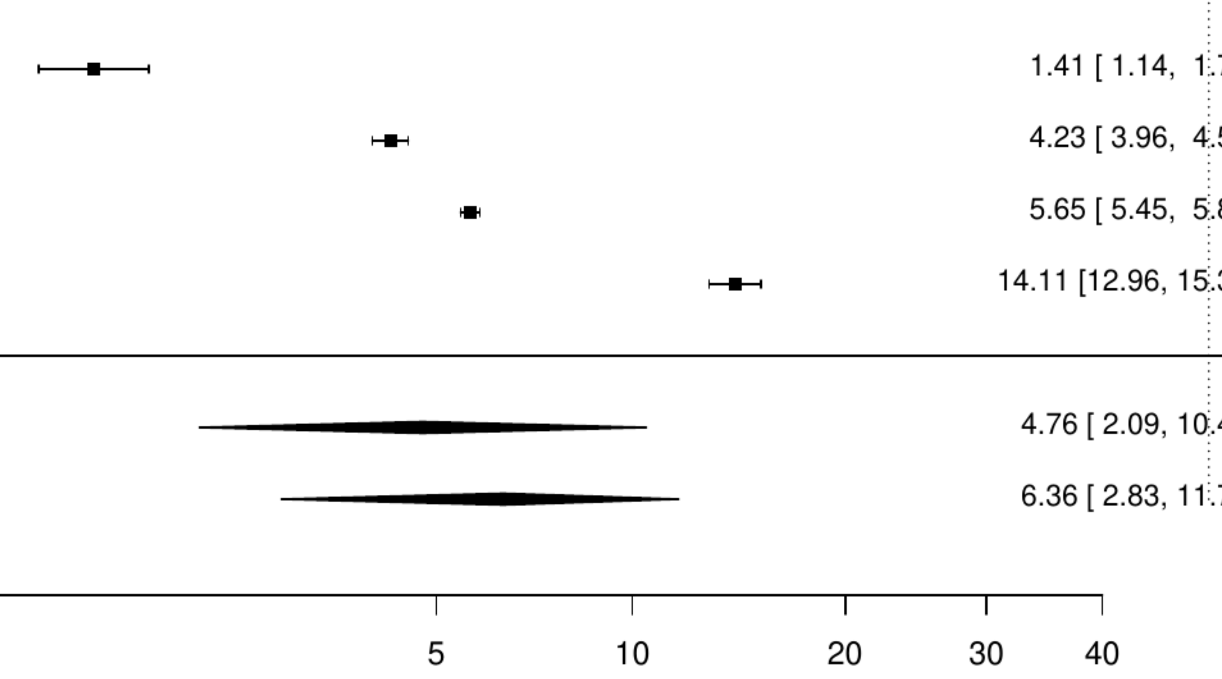

**Figure S31. Pooled prevalence of metastatic cancer in coronary artery bypass surgery**

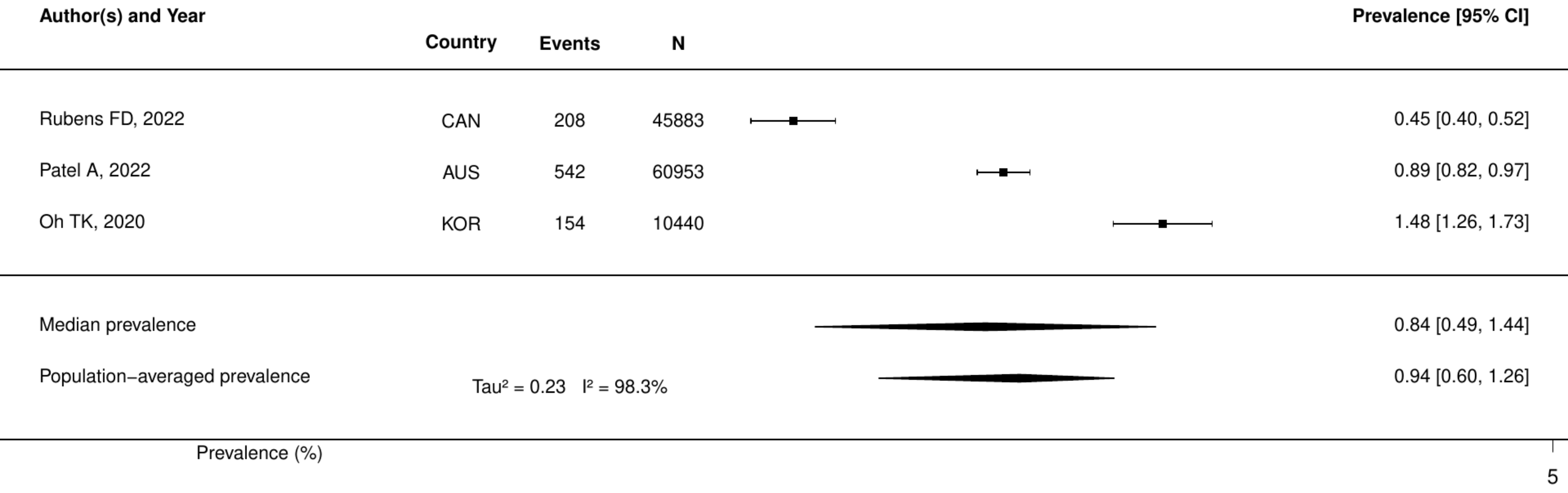

**Figure S32. Pooled prevalence of active cancer in atrial fibrillation.**

| Author(s) and Year | Country | Events | N | Prevalence [95% CI] |
|---|---|---|---|---|
| Ngo L, 2021 | Multinational | 212 | 35211 | 0.60 [ 0.53, 0.69] |
| Lodzinski P, 2020 | POL | 10 | 691 | 1.45 [ 0.78, 2.67] |
| Kirchhof P, 2020 | Multinational | 105 | 6784 | 1.55 [ 1.28, 1.87] |
| Du Y, 2021 | USA | 18 | 729 | 2.47 [ 1.56, 3.88] |
| Halvorsen S, 2022 | Multinational | 6102 | 219545 | 2.78 [ 2.71, 2.85] |
| Bernardini A, 2024 | ITA | 310 | 11078 | 2.80 [ 2.51, 3.12] |
| Atterman A, 2020 | SWE | 1518 | 52471 | 2.89 [ 2.75, 3.04] |
| Hanon O, 2017 | Multinational | 233 | 7225 | 3.22 [ 2.84, 3.66] |
| Lacoin L, 2017 | UK | 1822 | 41541 | 4.39 [ 4.19, 4.59] |
| Drusch S, 2024 | FRA | 5199 | 109693 | 4.74 [ 4.62, 4.87] |
| Coleman CI, 2021 | USA | 6170 | 116049 | 5.32 [ 5.19, 5.45] |
| Guo, 2022 | CHN | 202 | 3175 | 6.36 [ 5.56, 7.27] |
| Camelo−Castillo A, 2020 | ESP | 80 | 1044 | 7.66 [ 6.20, 9.44] |
| Rivera−Caravaca José Miguel, 2017 | ESP | 105 | 1361 | 7.71 [ 6.41, 9.26] |
| Yamato H, 2020 | JPN | 100 | 1086 | 9.21 [ 7.63, 11.08] |
| Bertomeu−González V, 2015 | ESP | 88 | 948 | 9.28 [ 7.59, 11.30] |
| Rutherford OW, 2020 | NOR | 6119 | 65563 | 9.33 [ 9.11, 9.56] |
| Rivera−Caravaca JM, 2022 | Multinational | 147 | 1479 | 9.94 [ 8.51, 11.57] |
| Kanemaru K, 2020 | JPN | 3589 | 32726 | 10.97 [10.63, 11.31] |
| Median prevalence | | | | 4.32 [ 3.05, 6.07] |
| Population−averaged prevalence | Tau² = 0.63 I² = 99.9% | | | 5.55 [ 3.97, 7.01] |

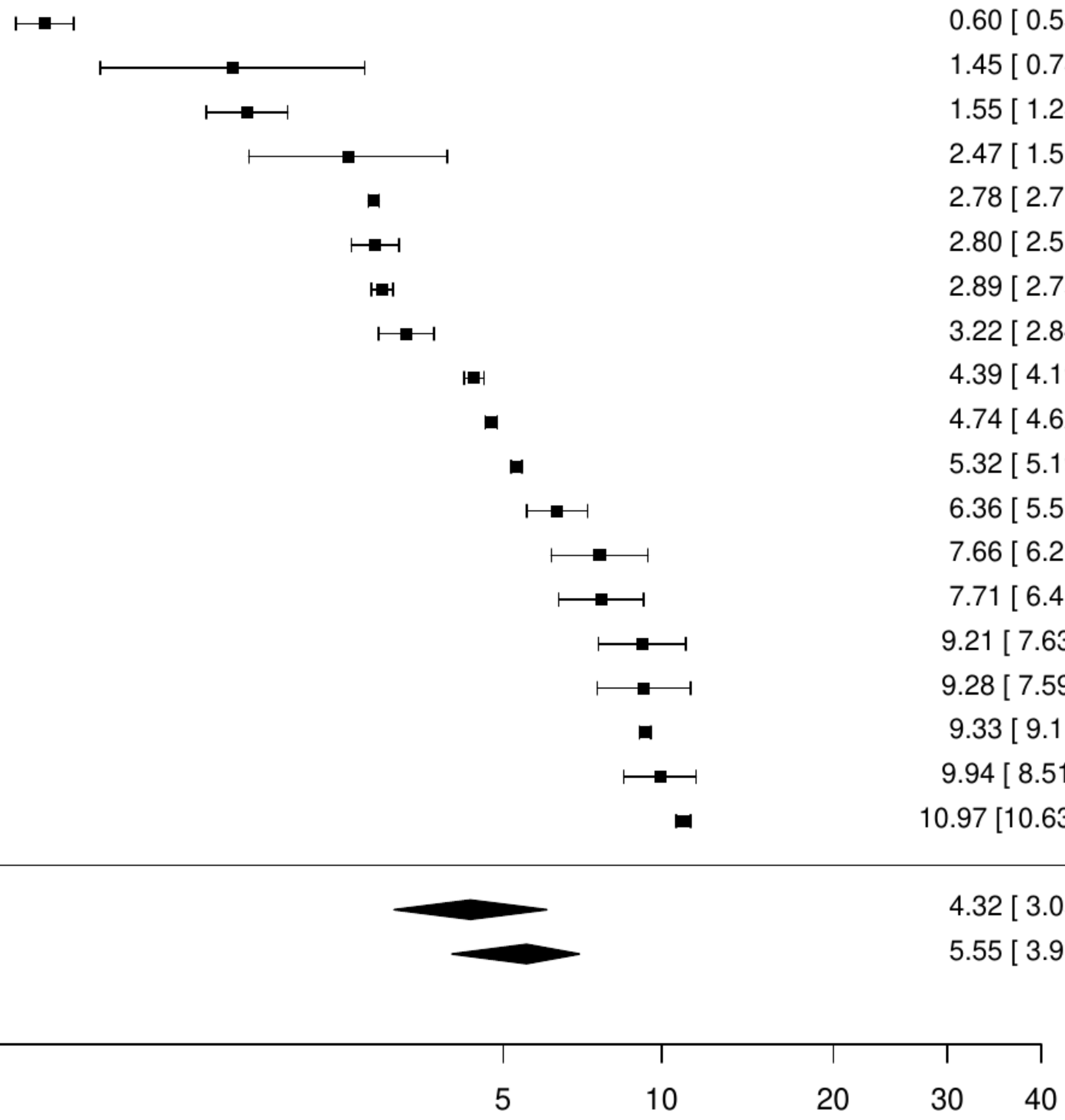

**Figure S33. Pooled prevalence of any cancer in atrial fibrillation.**

| Author(s) and Year | Country | Events | N | Prevalence [95% CI] |
|---|---|---|---|---|
| Deng F, 2020 | CHN | 260 | 15867 | 1.64 [ 1.45, 1.85] |
| Keller, 2019 | GER | 18335 | 1106019 | 1.66 [ 1.63, 1.68] |
| Hu WS, 2019 | TW | 249 | 14212 | 1.75 [ 1.55, 1.98] |
| Fauchier L, 2016 | FRA | 163 | 8962 | 1.82 [ 1.56, 2.12] |
| Bucci T, 2023 | Multinational | 95 | 4121 | 2.31 [ 1.89, 2.81] |
| Tanaka K, 2020 | Multinational | 30 | 1165 | 2.58 [ 1.81, 3.66] |
| Katsumata, 2021 | JPN | 34 | 1173 | 2.90 [ 2.08, 4.03] |
| Jurin I, 2020 | Croatia | 39 | 1000 | 3.90 [ 2.86, 5.29] |
| Potpara TS, 2020 | Multinational | 119 | 2671 | 4.46 [ 3.74, 5.31] |
| Lima FV, 2020 | USA | 35751 | 749776 | 4.77 [ 4.72, 4.82] |
| Chan YH, 2020 | TW | 3368 | 70408 | 4.78 [ 4.63, 4.94] |
| Jaspers Focks J, 2016 | NLD | 61 | 1157 | 5.27 [ 4.12, 6.72] |
| Lip Gregory Y H, 2014 | Multinational | 162 | 3049 | 5.31 [ 4.57, 6.17] |
| Chao TF, 2017 | TW | 19420 | 352656 | 5.51 [ 5.43, 5.58] |
| Cressman AM, 2015 | CAN | 11786 | 166742 | 7.07 [ 6.95, 7.19] |
| Giuseppe Boriani 1, 2018 | Multinational | 818 | 11021 | 7.42 [ 6.95, 7.93] |
| Furukawa Y, 2020 | JPN | 68 | 899 | 7.56 [ 6.01, 9.48] |
| Chen EYH, 2023 | Hong Kong | 1478 | 19292 | 7.66 [ 7.29, 8.05] |
| Chew DS, 2022 | CAN | 5066 | 64093 | 7.90 [ 7.70, 8.12] |
| Balsam P, 2018 | POL | 211 | 2657 | 7.94 [ 6.97, 9.03] |
| Mu Chen, 2023 | CHN | 22185 | 268401 | 8.27 [ 8.16, 8.37] |
| Bertomeu−Gonzalez, 2021 | ESP | 168 | 1956 | 8.59 [ 7.43, 9.91] |
| Tsuda T, 2023 | JPN | 127 | 1396 | 9.10 [ 7.70, 10.72] |
| Proietti, 2019 | ITA | 2189 | 24040 | 9.11 [ 8.75, 9.48] |
| Lin SM, 2023 | TW | 2334 | 24835 | 9.40 [ 9.04, 9.77] |
| Ferroni, 2019 | ITA | 1698 | 17920 | 9.48 [ 9.06, 9.91] |
| Denas G, 2017 | ITA | 3181 | 33488 | 9.50 [ 9.19, 9.82] |
| Romiti, 2024 | Multinational | 3164 | 32560 | 9.72 [ 9.40, 10.04] |
| Cho, 2020 | KOR | 886 | 8883 | 9.97 [ 9.37, 10.61] |
| Tan ESJ, 2022 | Singapore | 4171 | 40602 | 10.27 [ 9.98, 10.57] |
| Grymonprez M, 2023 | BEL | 31363 | 297818 | 10.53 [10.42, 10.64] |
| Koretsune Y, 2019 | JPN | 3589 | 32726 | 10.97 [10.63, 11.31] |
| Buckley, 2021 | USA | 150334 | 1366422 | 11.00 [10.95, 11.05] |
| Rivera−Caravaca JM, 2022 | ESP | 98 | 870 | 11.26 [ 9.33, 13.54] |
| Kjerpeseth LJ, 2019 | NOR | 3601 | 30820 | 11.68 [11.33, 12.05] |
| Elvira Ruiz, 2019 | ESP | 282 | 2361 | 11.94 [10.70, 13.32] |
| Ra.kauskas Gediminas, 2017 | Lithuania | 63 | 515 | 12.23 [ 9.67, 15.36] |
| Proietti M, 2016 | ITA | 76 | 558 | 13.62 [11.02, 16.72] |
| Cihan Ay, 2022 | AUT | 15504 | 112827 | 13.74 [13.54, 13.94] |
| Mostaza JM, 2021 | ESP | 133 | 961 | 13.84 [11.80, 16.17] |
| Martínez−Montesinos, 2023 | ESP | 150 | 1050 | 14.29 [12.30, 16.54] |
| Fernando SM, 2020 | CAN | 221 | 1541 | 14.34 [12.68, 16.18] |
| Shiroto T, 2020 | JPN | 493 | 3221 | 15.31 [14.10, 16.59] |
| Huang JY, 2023 | CHN | 8054 | 52490 | 15.34 [15.04, 15.65] |
| Rodriguez−Bernal CL, 2018 | ESP | 2994 | 18715 | 16.00 [15.48, 16.53] |
| Peacock W Frank, 2017 | USA | 7189 | 44793 | 16.05 [15.71, 16.39] |
| Pastori D, 2021 | FRA | 399344 | 2435541 | 16.40 [16.35, 16.44] |
| O'Neal WT, 2018 | USA | 64016 | 388045 | 16.50 [16.38, 16.61] |
| Wilkinson C, 2021 | UK | 10225 | 61177 | 16.71 [16.42, 17.01] |
| Garcia−Sempere A, 2019 | ESP | 3878 | 22629 | 17.14 [16.65, 17.63] |
| Dhamane, 2023 | USA | 28438 | 163073 | 17.44 [17.26, 17.62] |
| Pastori D, 2020 | ITA | 367 | 2092 | 17.54 [15.97, 19.23] |
| Chung SC, 2023 | UK | 39780 | 225173 | 17.67 [17.51, 17.82] |
| Falsetti L, 2020 | ITA | 254 | 1430 | 17.76 [15.87, 19.83] |
| Kim, 2020 | KOR | 30571 | 171228 | 17.85 [17.67, 18.04] |
| Penttila T, 2019 | FIN | 10408 | 54568 | 19.07 [18.75, 19.41] |
| Lehto, 2017 | FIN | 10408 | 54568 | 19.07 [18.75, 19.41] |
| Gamble DT, 2020 | UK | 373 | 1928 | 19.35 [17.64, 21.17] |
| Dalgaard, 2020 | USA | 6727 | 34174 | 19.68 [19.27, 20.11] |
| Moskowitz A, 2017 | USA | 329 | 1646 | 19.99 [18.13, 21.99] |
| Williams BA, 2017 | USA | 1575 | 7877 | 19.99 [19.13, 20.89] |
| Loikas D, 2017 | SWE | 17894 | 89400 | 20.02 [19.75, 20.28] |
| Fu EL, 2024 | USA | 17029 | 84221 | 20.22 [19.95, 20.49] |
| van Rein, 2019 | DEN | 55186 | 272315 | 20.27 [20.11, 20.42] |
| Teppo, 2023 | FIN | 49028 | 239222 | 20.49 [20.33, 20.66] |
| Renoux C, 2017 | CAN | 31430 | 147622 | 21.29 [21.08, 21.50] |
| Freeman JV, 2019 | USA | 4657 | 21595 | 21.57 [21.02, 22.12] |
| Kohsaka S, 2020 | JPN | 18219 | 79887 | 22.81 [22.52, 23.10] |
| Andersson T, 2021 | SWE | 71813 | 294470 | 24.39 [24.23, 24.54] |
| Ryden A, 2018 | SWE | 2464 | 10039 | 24.54 [23.71, 25.40] |
| Komen J, 2017 | SWE | 1678 | 6765 | 24.80 [23.79, 25.85] |
| McIntyre, 2018 | CAN | 165 | 548 | 30.11 [26.41, 34.08] |
| Abu, 2020 | USA | 377 | 1235 | 30.53 [28.02, 33.15] |
| Weber Courtney, 2021 | AUS | 17412 | 52447 | 33.20 [32.80, 33.60] |
| Shah RU, 2020 | USA | 235 | 632 | 37.18 [33.50, 41.02] |
| Bezabhe WM, 2022 | AUS | 7642 | 18813 | 40.62 [39.92, 41.32] |
| Median prevalence | Tau² = 0.69 I² = 100% | | | 11.60 [ 9.81, 13.66] |
| Population−averaged prevalence | | | | 14.10 [12.20, 15.99] |

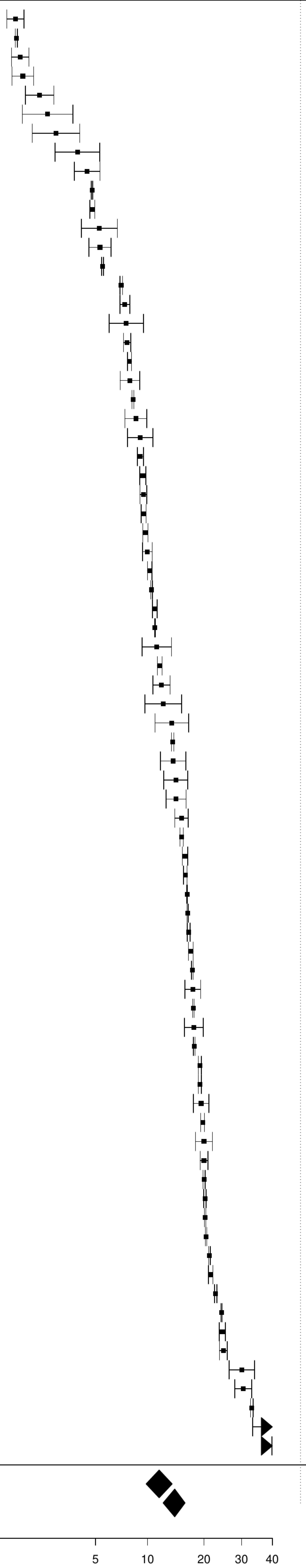

**Figure S34. Pooled prevalence of previous cancer in atrial fibrillation.**

| Author(s) and Year | Country | Events | N | Prevalence [95% CI] |
|---|---|---|---|---|
| Di Monaco A, 2022 | ITA | 68 | 1260 | 5.40 [ 4.28, 6.79] |
| Ajabnoor AM, 2023 | UK | 11379 | 177065 | 6.43 [ 6.31, 6.54] |
| Ortiz MR, 2020 | ESP | 55 | 574 | 9.58 [ 7.43, 12.28] |
| Corica B, 2024 | Multinational | 2388 | 24574 | 9.72 [ 9.35, 10.09] |
| Friberg Leif, 2018 | SWE | 33238 | 312341 | 10.64 [10.53, 10.75] |
| Okumura K, 2020 | JPN | 3600 | 32726 | 11.00 [10.67, 11.34] |
| Abdel−Qadir H, 2022 | CAN | 39305 | 347632 | 11.31 [11.20, 11.41] |
| Chen Q, 2024 | NLD | 40224 | 320139 | 12.56 [12.45, 12.68] |
| de la Torre Hernandez Jose M, 2021 | ESP | 161 | 1249 | 12.89 [11.14, 14.87] |
| Vanbeselaere, 2016 | BEL | 237 | 1830 | 12.95 [11.49, 14.57] |
| Bhonsale A, 2024 | USA | 9510 | 67221 | 14.15 [13.89, 14.41] |
| Sogaard M, 2024 | DEN | 5406 | 36223 | 14.92 [14.56, 15.29] |
| Melgaard L, 2020 | DEN | 4887 | 28770 | 16.99 [16.56, 17.42] |
| Saliba W, 2015 | ISR | 6710 | 32912 | 20.39 [19.96, 20.83] |
| Kim K, 2021 | KOR | 138902 | 615724 | 22.56 [22.45, 22.66] |
| Noseworthy PA, 2022 | USA | 153663 | 562850 | 27.30 [27.18, 27.42] |
| Abbas SS, 2021 | AUS | 521 | 1206 | 43.20 [40.43, 46.02] |
| Median prevalence | | | | 13.91 [10.83, 17.68] |
| Population−averaged prevalence | | | | 15.33 [11.88, 19.97] |

Tau² = 0.36 I² = 100%

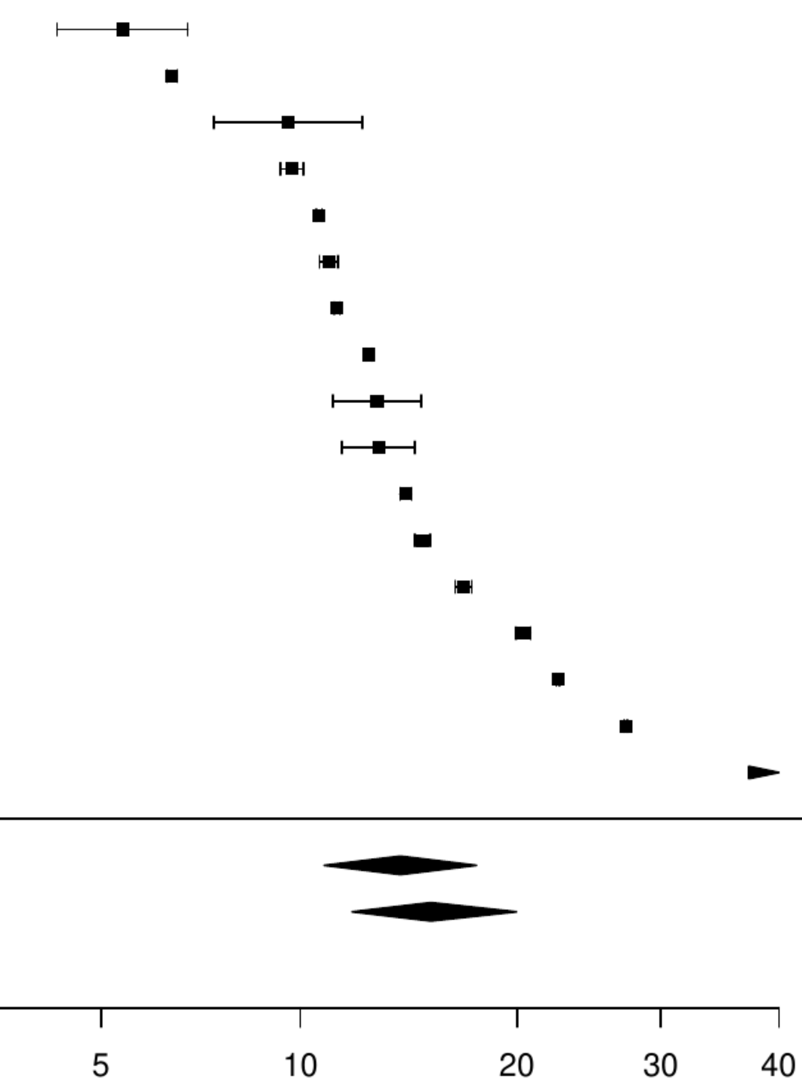

**Figure S35. Pooled prevalence of blood cancer in atrial fibrillation.**

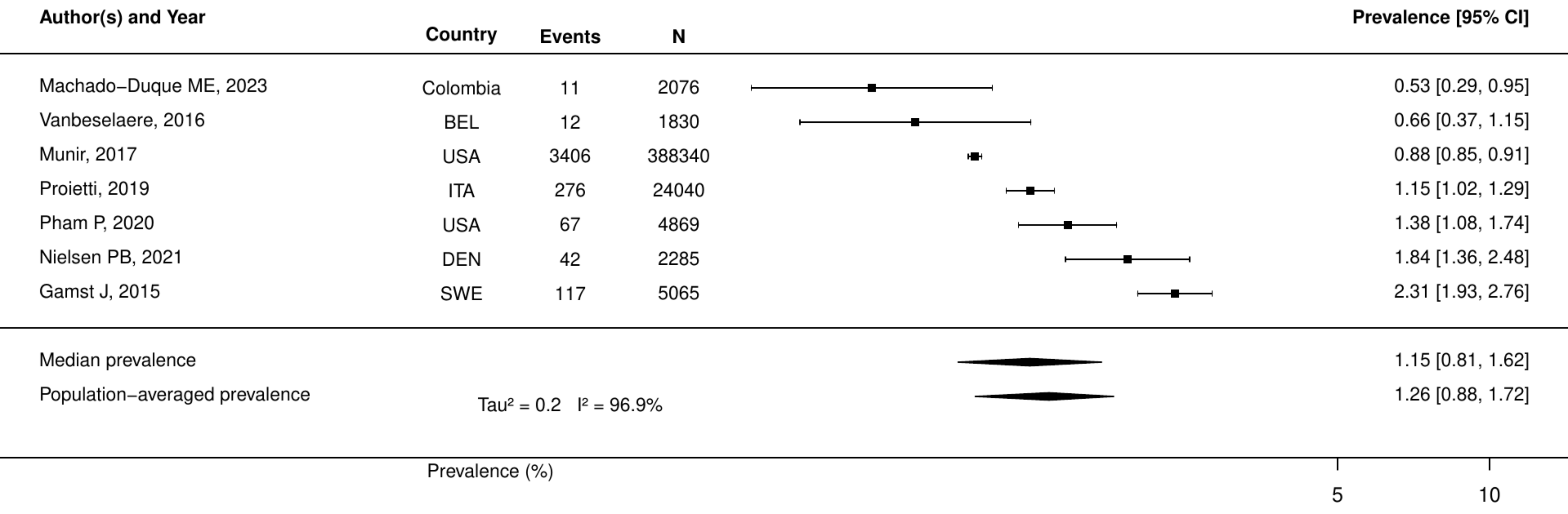

### Figure S36. Pooled prevalence of metastatic cancer in atrial fibrillation.

| Author(s) and Year | Country | Events | N | Prevalence [95% CI] |
|---|---|---|---|---|
| König, 2018 | GER | 646 | 161502 | 0.40 [0.37, 0.43] |
| Rome, 2021 | USA | 342 | 35309 | 0.97 [0.87, 1.08] |
| Munir, 2017 | USA | 5307 | 388340 | 1.37 [1.33, 1.40] |
| Lutsey PL, 2020 | USA | 3266 | 167275 | 1.95 [1.89, 2.02] |
| Field, 2019 | KOR | 303 | 15276 | 1.98 [1.77, 2.22] |
| Proietti, 2019 | ITA | 503 | 24040 | 2.09 [1.92, 2.28] |
| Jun, 2017 | CAN | 496 | 17988 | 2.76 [2.53, 3.01] |
| Gamst J, 2015 | DEN | 149 | 5065 | 2.94 [2.51, 3.44] |
| Ryden A, 2018 | SWE | 514 | 10039 | 5.12 [4.71, 5.57] |
| Nielsen PB, 2021 | DEN | 172 | 2285 | 7.53 [6.51, 8.68] |
| Median prevalence | | | | 2.06 [1.27, 3.35] |
| Population−averaged prevalence | | | | 2.74 [1.68, 4.14] |

$Tau^2 = 0.64$   $I^2 = 99.8\%$

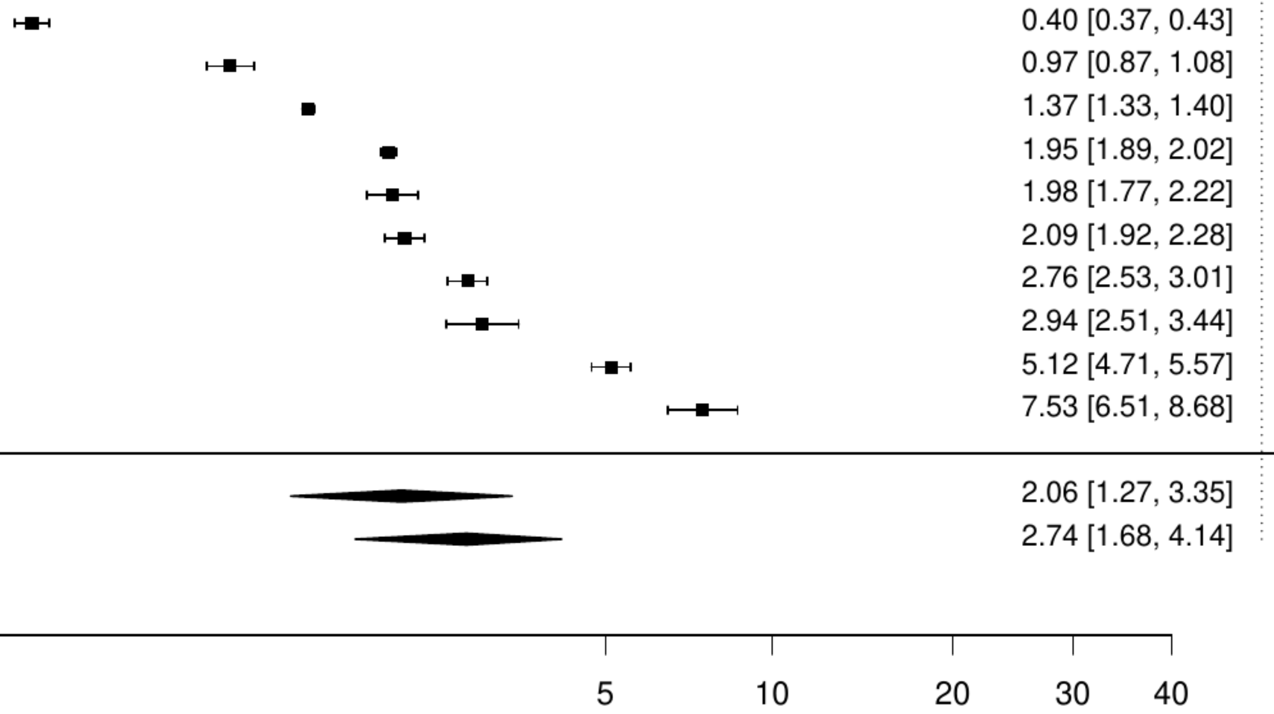

**Figure S37. Pooled prevalence of solid cancer in atrial fibrillation.**

| Author(s) and Year | Country | Events | N | Prevalence [95% CI] |
|---|---|---|---|---|
| Machado−Duque ME, 2023 | Colombia | 21 | 2076 | 1.01 [ 0.66, 1.55] |
| Munir, 2017 | USA | 8169 | 388340 | 2.10 [ 2.06, 2.15] |
| Nielsen PB, 2021 | DEN | 323 | 2285 | 14.14 [12.77, 15.63] |
| Pham P, 2020 | USA | 780 | 4869 | 16.02 [15.02, 17.08] |
| Median prevalence | | | | 4.90 [ 1.46, 15.15] |
| Population−averaged prevalence | | | | 8.43 [ 1.62, 15.24] |

$Tau^2 = 1.6$ $I^2 = 99.9\%$

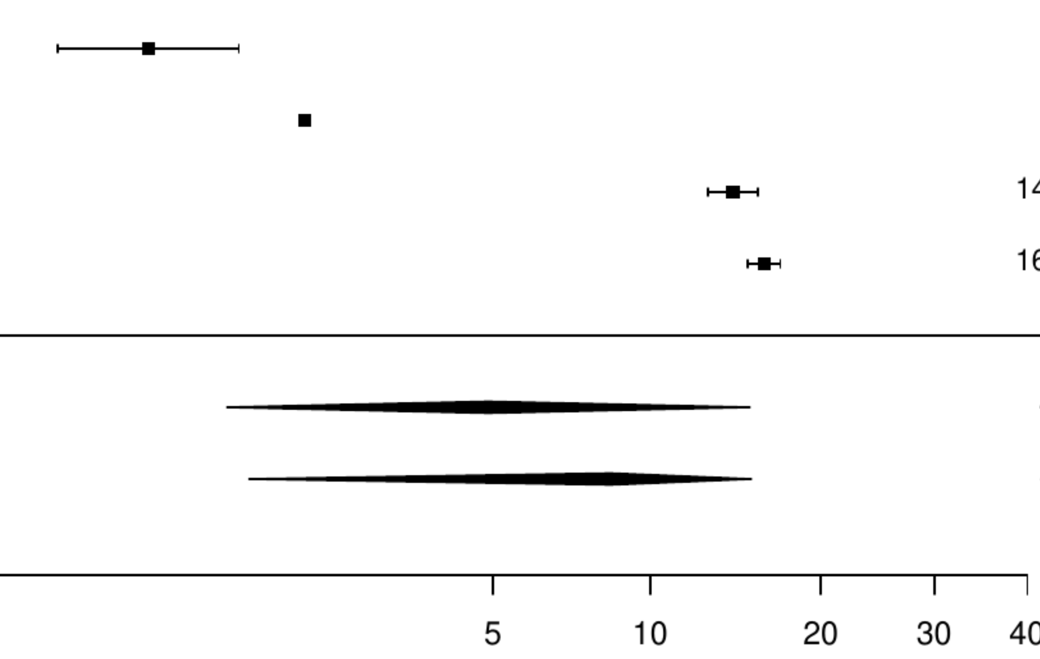

**Figure S38. Pooled prevalence of active cancer in non-valvular atrial fibrillation.**

| Author(s) and Year | Country | Events | N | Prevalence [95% CI] |
|---|---|---|---|---|
| Kirchhof P, 2020 | Multinational | 105 | 6784 | 1.55 [ 1.28, 1.87] |
| Halvorsen S, 2022 | Multinational | 6102 | 219545 | 2.78 [ 2.71, 2.85] |
| Bernardini A, 2024 | ITA | 310 | 11078 | 2.80 [ 2.51, 3.12] |
| Lacoin L, 2017 | UK | 1822 | 41541 | 4.39 [ 4.19, 4.59] |
| Drusch S, 2024 | FRA | 5199 | 109693 | 4.74 [ 4.62, 4.87] |
| Coleman CI, 2021 | USA | 6170 | 116049 | 5.32 [ 5.19, 5.45] |
| Camelo−Castillo A, 2020 | ESP | 80 | 1044 | 7.66 [ 6.20, 9.44] |
| Yamato H, 2020 | JPN | 100 | 1086 | 9.21 [ 7.63, 11.08] |
| Bertomeu−González V, 2015 | ESP | 88 | 948 | 9.28 [ 7.59, 11.30] |
| Kanemaru K, 2020 | JPN | 3589 | 32726 | 10.97 [10.63, 11.31] |
| Median prevalence | | | | 5.02 [ 3.45, 7.24] |
| Population−averaged prevalence | | | | 5.87 [ 3.92, 7.74] |

$Tau^2 = 0.39$ $I^2 = 99.9\%$

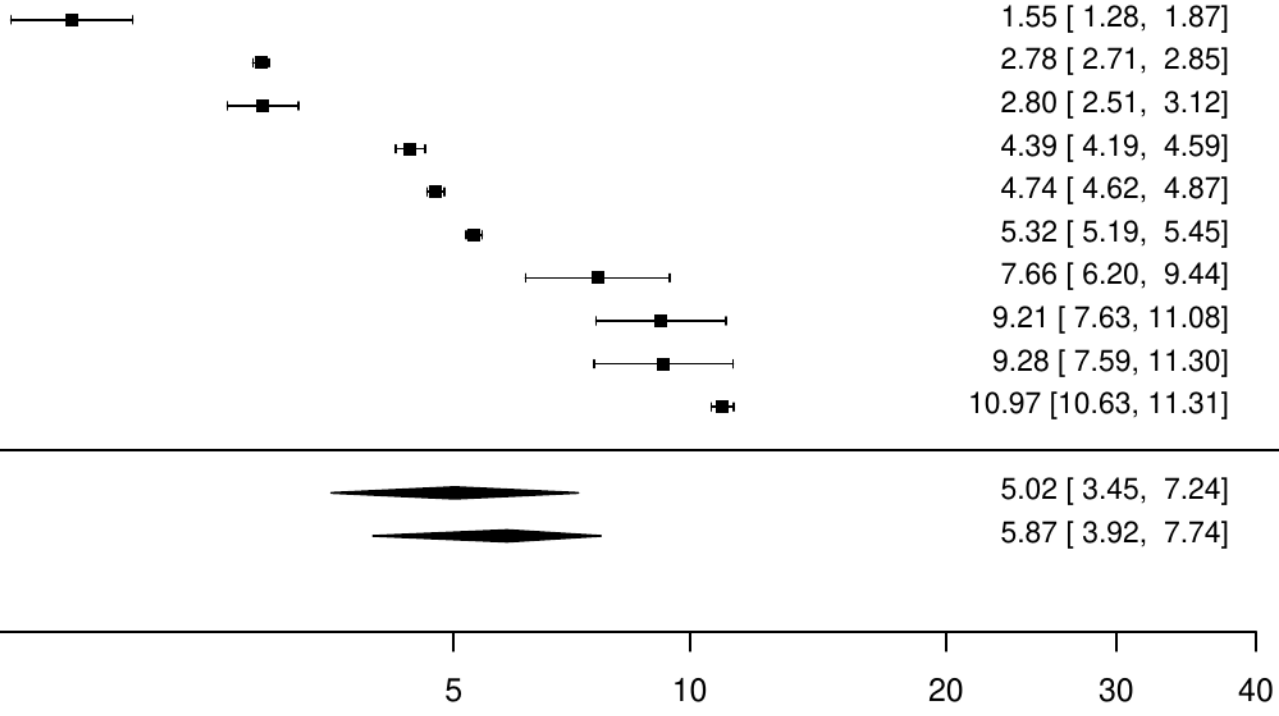

**Figure S39. Pooled prevalence of any cancer in non-valvular atrial fibrillation.**

| Author(s) and Year | Country | Events | N | Prevalence [95% CI] |
|---|---|---|---|---|
| Tanaka K, 2020 | Multinational | 30 | 1165 | 2.58 [ 1.81, 3.66] |
| Jurin I, 2020 | Croatia | 39 | 1000 | 3.90 [ 2.86, 5.29] |
| Potpara TS, 2020 | Multinational | 119 | 2671 | 4.46 [ 3.74, 5.31] |
| Lee SR, 2024 | KOR | 8778 | 165624 | 5.30 [ 5.19, 5.41] |
| Chen EYH, 2023 | Hong Kong | 1478 | 19292 | 7.66 [ 7.29, 8.05] |
| Balsam P, 2018 | POL | 211 | 2657 | 7.94 [ 6.97, 9.03] |
| Bertomeu−Gonzalez, 2021 | ESP | 168 | 1956 | 8.59 [ 7.43, 9.91] |
| Tsuda T, 2023 | JPN | 127 | 1396 | 9.10 [ 7.70, 10.72] |
| Denas G, 2017 | ITA | 3832 | 40411 | 9.48 [ 9.20, 9.77] |
| Romiti, 2024 | Multinational | 3164 | 32560 | 9.72 [ 9.40, 10.04] |
| Chan YH, 2022 | TW | 10407 | 106044 | 9.81 [ 9.64, 9.99] |
| Cho, 2020 | KOR | 886 | 8883 | 9.97 [ 9.37, 10.61] |
| Tan ESJ, 2022 | Singapore | 4171 | 40602 | 10.27 [ 9.98, 10.57] |
| Grymonprez M, 2023 | BEL | 31363 | 297818 | 10.53 [10.42, 10.64] |
| Koretsune Y, 2019 | JPN | 3589 | 32726 | 10.97 [10.63, 11.31] |
| Rivera−Caravaca JM, 2022 | ESP | 98 | 870 | 11.26 [ 9.33, 13.54] |
| Durand Madeleine, 2020 | CAN | 29645 | 255851 | 11.59 [11.46, 11.71] |
| Kjerpeseth LJ, 2019 | NOR | 3601 | 30820 | 11.68 [11.33, 12.05] |
| Elvira Ruiz, 2019 | ESP | 282 | 2361 | 11.94 [10.70, 13.32] |
| Mostaza JM, 2021 | ESP | 133 | 961 | 13.84 [11.80, 16.17] |
| Martínez−Montesinos, 2023 | ESP | 150 | 1050 | 14.29 [12.30, 16.54] |
| Huang JY, 2023 | CHN | 8054 | 52490 | 15.34 [15.04, 15.65] |
| Sogaard M, 2017 | DEN | 40396 | 253741 | 15.92 [15.78, 16.06] |
| Rodriguez−Bernal CL, 2018 | ESP | 2994 | 18715 | 16.00 [15.48, 16.53] |
| Peacock W Frank, 2017 | USA | 7189 | 44793 | 16.05 [15.71, 16.39] |
| O'Neal WT, 2018 | USA | 64016 | 388045 | 16.50 [16.38, 16.61] |
| Steinberg BA, 2015 | USA | 14823 | 86671 | 17.10 [16.85, 17.35] |
| Norhammar A, 2016 | SWE | 387725 | 2248327 | 17.25 [17.20, 17.29] |
| Dhamane, 2023 | USA | 28438 | 163073 | 17.44 [17.26, 17.62] |
| Pastori D, 2020 | ITA | 367 | 2092 | 17.54 [15.97, 19.23] |
| Yao, 2017 | USA | 12757 | 72568 | 17.58 [17.30, 17.86] |
| Dalgaard, 2020 | USA | 6727 | 34174 | 19.68 [19.27, 20.11] |
| Williams BA, 2017 | USA | 1575 | 7877 | 19.99 [19.13, 20.89] |
| Lehto M, 2024 | FIN | 15089 | 74008 | 20.39 [20.10, 20.68] |
| Renoux C, 2017 | CAN | 31430 | 147622 | 21.29 [21.08, 21.50] |
| Jackson, 2018 | USA | 4049 | 18398 | 22.01 [21.42, 22.61] |
| Ajabnoor AM, 2022 | UK | 42434 | 192265 | 22.07 [21.89, 22.26] |
| Kohsaka S, 2020 | JPN | 18219 | 79887 | 22.81 [22.52, 23.10] |
| Shih CJ, 2016 | TW | 1638 | 6772 | 24.19 [23.18, 25.22] |
| Komen J, 2017 | SWE | 1678 | 6765 | 24.80 [23.79, 25.85] |
| Median prevalence | Tau² = 0.31 I² = 100% | | | 12.80 [10.98, 14.88] |
| Population−averaged prevalence | | | | 14.03 [12.19, 15.73] |

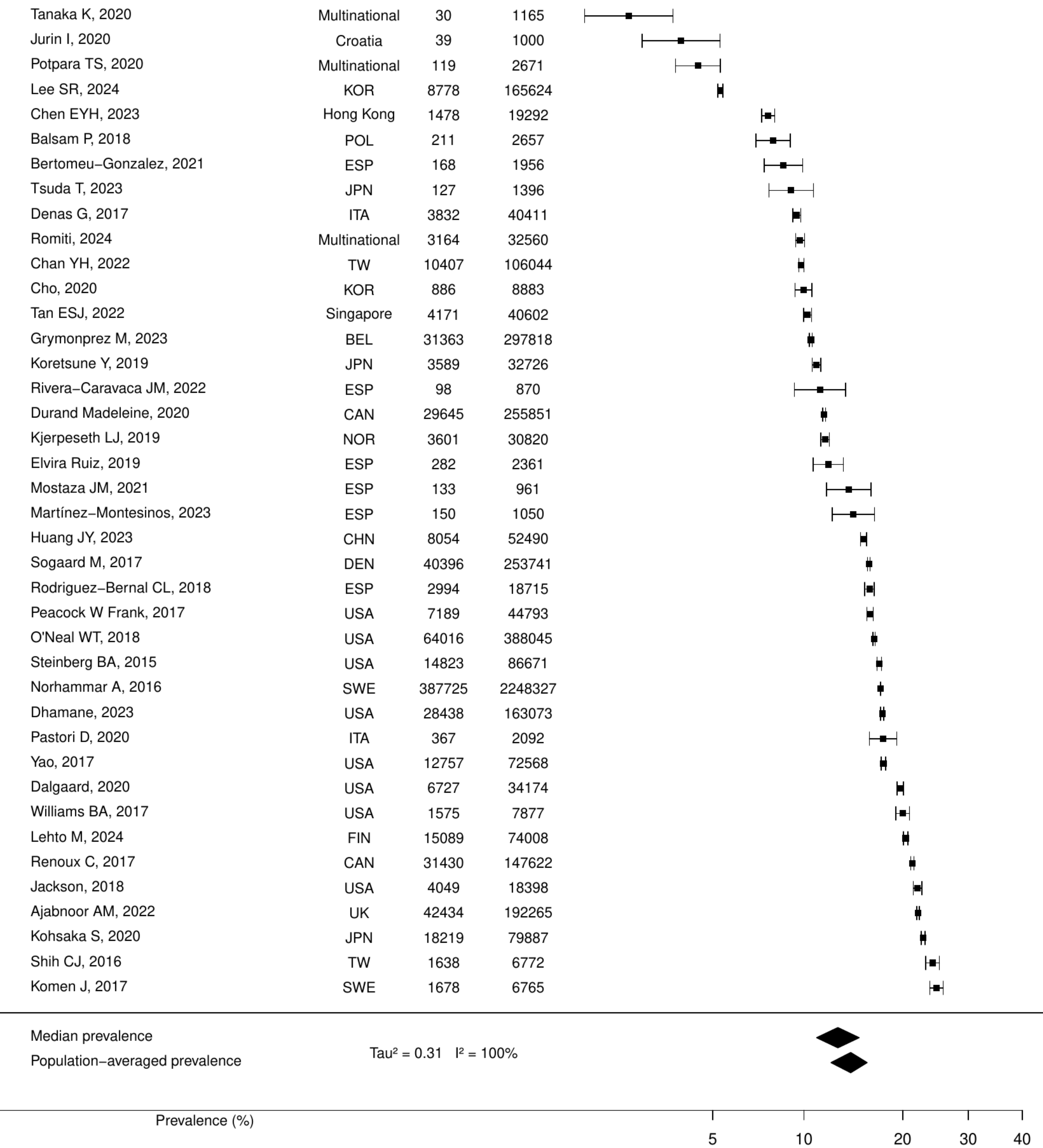

**Figure S40. Pooled prevalence of previous cancer in non-valvular atrial fibrillation.**

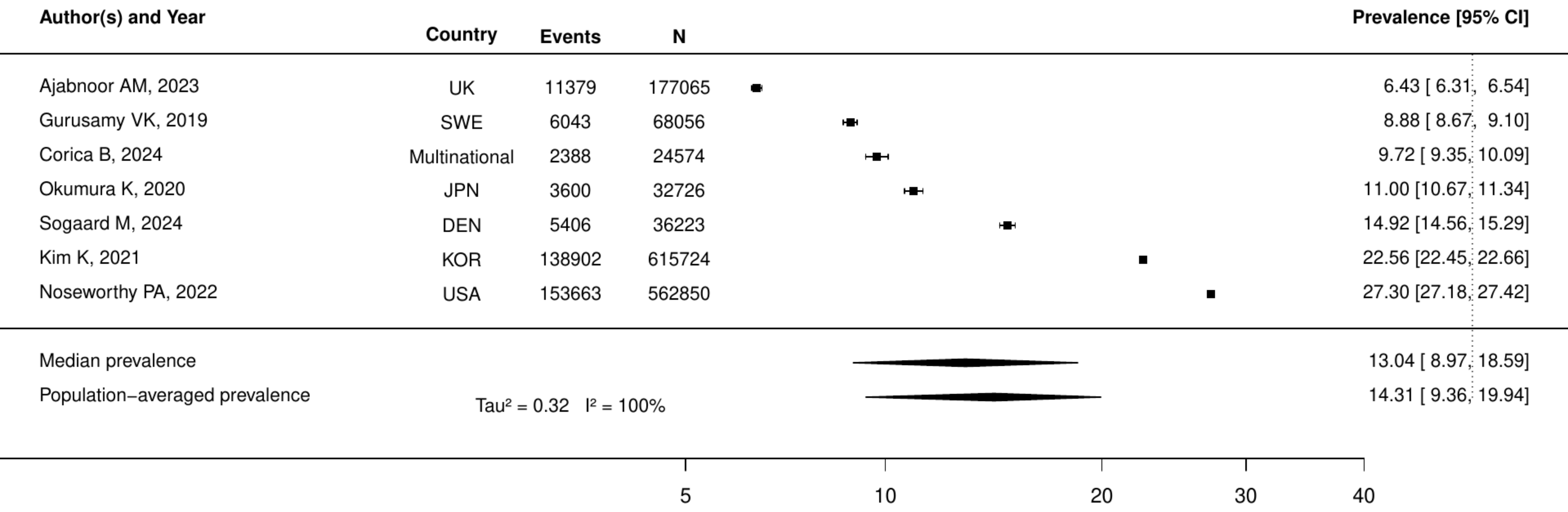

**Figure S41. Pooled prevalence of blood cancer in non-valvular atrial fibrillation.**

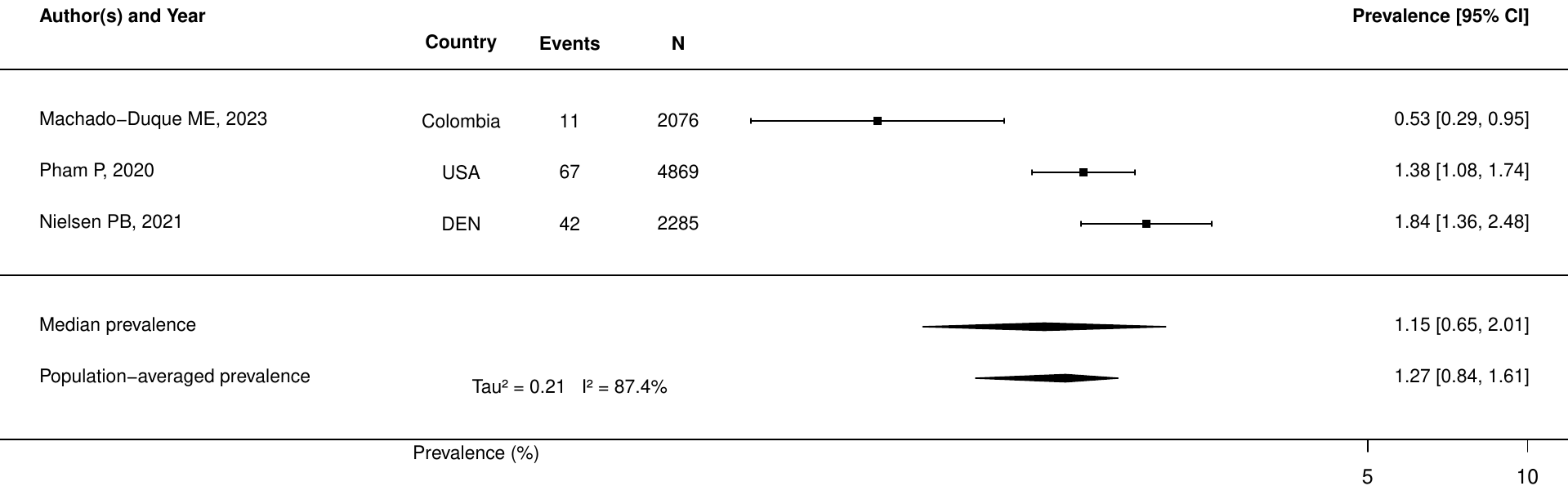

**Figure S42. Pooled prevalence of metastatic cancer in non-valvular atrial fibrillation.**

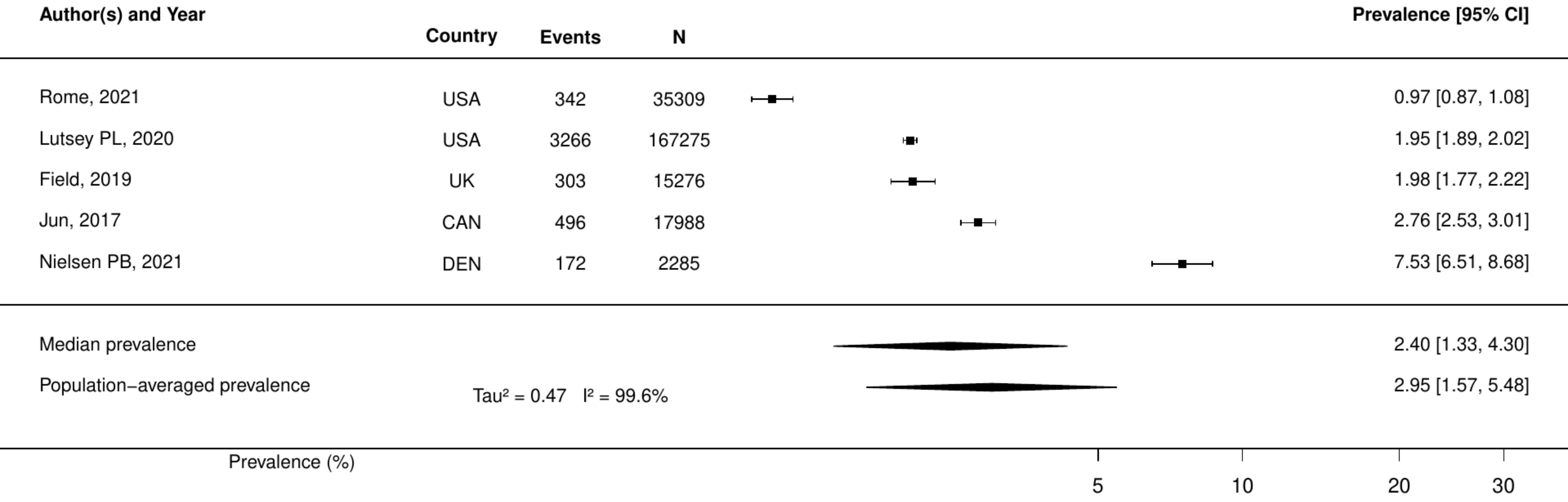

**Figure S43. Pooled prevalence of solid cancer in non-valvular atrial fibrillation.**

| Author(s) and Year | Country | Events | N | Prevalence [95% CI] |
|---|---|---|---|---|
| Machado−Duque ME, 2023 | Colombia | 21 | 2076 | 1.01 [ 0.66, 1.55] |
| Nielsen PB, 2021 | DEN | 323 | 2285 | 14.14 [12.77, 15.63] |
| Pham P, 2020 | USA | 780 | 4869 | 16.02 [15.02, 17.08] |
| Median prevalence | | | | 6.45 [ 1.48, 24.02] |
| Population−averaged prevalence | | | | 11.06 [ 4.97, 15.60] |

Tau² = 1.79 I² = 99.7%

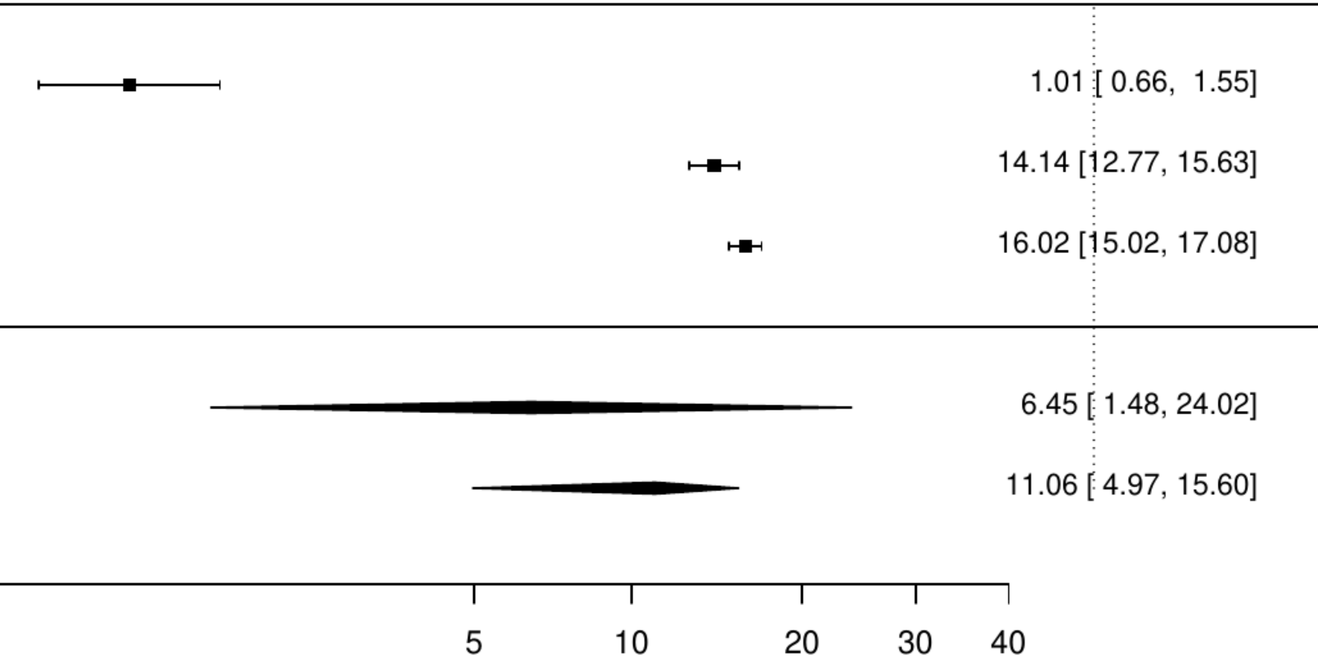

### Figure S44. Pooled prevalence of active cancer in chronic heart failure

| Author(s) and Year | Country | Events | N | Prevalence [95% CI] |
|---|---|---|---|---|
| Wiskar K, 2017 | USA | 1712 | 102746 | 1.67 [1.59, 1.75] |
| Vergaro, 2019 | ITA | 50 | 2791 | 1.79 [1.36, 2.36] |
| Zaprutko J, 2017 | POL | 25 | 765 | 3.27 [2.22, 4.79] |
| Vinter N, 2023 | DEN | 1159 | 27947 | 4.15 [3.92, 4.39] |
| Greene, 2022 | Multinational | 71 | 1619 | 4.39 [3.49, 5.50] |
| Spatz ES, 2016 | USA | 188478 | 2581892 | 7.30 [7.27, 7.33] |
| Sun LY, 2020 | CAN | 29291 | 352329 | 8.31 [8.22, 8.41] |
| Median prevalence | | | | 3.80 [2.45, 5.86] |
| Population−averaged prevalence | | | | 4.43 [2.78, 6.38] |

Tau² = 0.36   I² = 100%

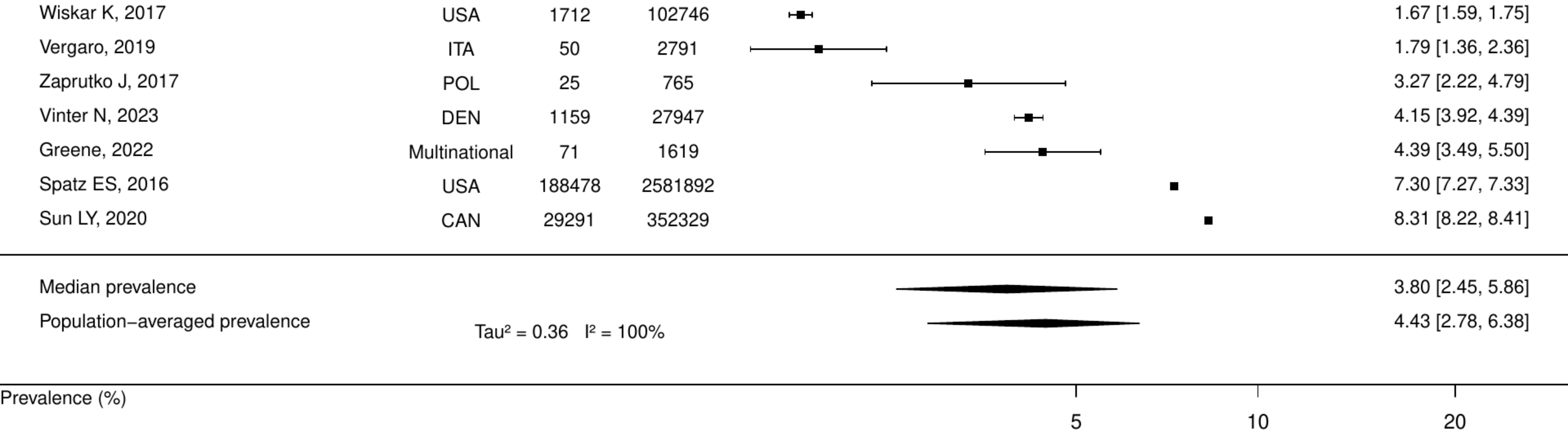

**Figure S45. Pooled prevalence of any cancer in chronic heart failure**

| Author(s) and Year | Country | Events | N | Prevalence [95% CI] |
|---|---|---|---|---|
| Ozlek B, 2019 | TUR | 13 | 819 | 1.59 [ 0.92, 2.71] |
| Arundel C, 2016 | USA | 148 | 7578 | 1.95 [ 1.66, 2.29] |
| Wang H, 2024 | CHN | 7759 | 327472 | 2.37 [ 2.32, 2.42] |
| Chen YL, 2017 | TW | 38 | 1311 | 2.90 [ 2.12, 3.96] |
| Stein G, 2020 | Singapore | 35 | 1070 | 3.27 [ 2.36, 4.52] |
| MacDonald, 2020 | Multinational | 216 | 6480 | 3.33 [ 2.92, 3.80] |
| Rivera−Toquica, 2024 | Colombia | 49 | 1384 | 3.54 [ 2.69, 4.65] |
| Antonides CFJ, 2022 | Multinational | 67 | 1821 | 3.68 [ 2.91, 4.65] |
| Arora S, 2017 | USA | 11774 | 301892 | 3.90 [ 3.83, 3.97] |
| Kormos, 2019 | USA | 853 | 18539 | 4.60 [ 4.31, 4.91] |
| Lima FV, 2020 | USA | 35751 | 749776 | 4.77 [ 4.72, 4.82] |
| Arutyunov AG, 2015 | RUS | 125 | 2498 | 5.00 [ 4.21, 5.93] |
| Tisminetzky, 2018 | USA | 6186 | 114553 | 5.40 [ 5.27, 5.53] |
| Crespo−Leiro MG, 2020 | ESP | 208 | 3587 | 5.80 [ 5.08, 6.61] |
| Li L, 2018 | USA | 9206 | 150959 | 6.10 [ 5.98, 6.22] |
| Cocchieri A, 2015 | ITA | 75 | 1192 | 6.29 [ 5.05, 7.82] |
| Braga JR, 2019 | CAN | 811 | 12814 | 6.33 [ 5.92, 6.76] |
| Ibrahim, 2020 | USA | 45 | 692 | 6.50 [ 4.89, 8.60] |
| Blecker S, 2016 | USA | 339 | 4723 | 7.18 [ 6.48, 7.95] |
| Yamada S, 2021 | JPN | 196 | 2721 | 7.20 [ 6.29, 8.24] |
| Kundi, 2019 | USA | 28076 | 348619 | 8.05 [ 7.96, 8.14] |
| Jawitz, 2020 | USA | 1537 | 18307 | 8.40 [ 8.00, 8.81] |
| Omersa D, 2019 | Slovenia | 144058 | 1656810 | 8.69 [ 8.65, 8.74] |
| Trullàs JC, 2016 | ESP | 122 | 1202 | 10.15 [ 8.57, 11.99] |
| Obata H, 2021 | JPN | 64 | 617 | 10.37 [ 8.20, 13.04] |
| Sicras−Mainar, 2022 | ESP | 2055 | 19762 | 10.40 [ 9.98, 10.83] |
| Guzman, 2018 | ESP | 259 | 2364 | 10.96 [ 9.76, 12.28] |
| Wienbergen H, 2019 | Multinational | 104 | 949 | 10.96 [ 9.12, 13.11] |
| Kusunose K, 2021 | JPN | 10306 | 93692 | 11.00 [10.80, 11.20] |
| Baldi Ileana, 2017 | ITA | 5743 | 51061 | 11.25 [10.98, 11.52] |
| Vaduganathan M, 2020 | USA | 565 | 4970 | 11.37 [10.52, 12.28] |
| Nakagawa A, 2021 | JPN | 112 | 960 | 11.67 [ 9.78, 13.86] |
| Iorio, 2018 | ITA | 283 | 2314 | 12.23 [10.96, 13.63] |
| Yamauchi T, 2016 | JPN | 628 | 4812 | 13.05 [12.13, 14.03] |
| Bosco−Lévy P, 2020 | FRA | 971 | 7387 | 13.14 [12.39, 13.93] |
| Chen X, 2019 | SWE | 5681 | 42987 | 13.22 [12.90, 13.54] |
| Yang MX, 2019 | CHN | 141 | 1053 | 13.39 [11.46, 15.58] |
| Sessa, 2019 | ITA | 1554 | 10091 | 15.40 [14.71, 16.12] |
| Chen CY, 2015 | USA | 3918 | 23111 | 16.95 [16.47, 17.44] |
| Munoz MA, 2020 | ESP | 216 | 1148 | 18.82 [16.66, 21.18] |
| Zheng, 2022 | USA | 105523 | 558322 | 18.90 [18.80, 19.00] |
| Tiili, 2019 | FIN | 10285 | 53953 | 19.06 [18.73, 19.40] |
| Farre N, 2017 | ESP | 18545 | 88195 | 21.03 [20.76, 21.30] |
| Farré N, 2016 | ESP | 18545 | 88195 | 21.03 [20.76, 21.30] |
| Shuvy, 2020 | ISR | 1013 | 4668 | 21.70 [20.54, 22.91] |
| Ou, 2017 | TW | 43428 | 196986 | 22.05 [21.86, 22.23] |
| Kase M, 2023 | JPN | 285 | 1269 | 22.46 [20.25, 24.84] |
| Zhao Q, 2019 | USA | 6536 | 28983 | 22.55 [22.07, 23.04] |
| Christiansen, 2020 | DEN | 64527 | 280002 | 23.05 [22.89, 23.20] |
| Lawson, 2019 | UK | 20500 | 88416 | 23.19 [22.91, 23.47] |
| Douros A, 2021 | GER | 129 | 544 | 23.71 [20.33, 27.47] |
| Goyal P, 2016 | USA | 195 | 796 | 24.50 [21.63, 27.61] |
| Kennel PJ, 2019 | USA | 256 | 947 | 27.03 [24.30, 29.95] |
| Weber, 2023 | AUS | 5389 | 18241 | 29.54 [28.89, 30.21] |
| Manemann SM, 2016 | USA | 525 | 1714 | 30.63 [28.49, 32.85] |
| Lin AH, 2017 | USA | 19284 | 51286 | 37.60 [37.18, 38.02] |
| Median prevalence | Tau² = 0.72 I² = 100% | | | 10.34 [ 8.45, 12.60] |
| Population−averaged prevalence | | | | 12.82 [10.60, 14.89] |

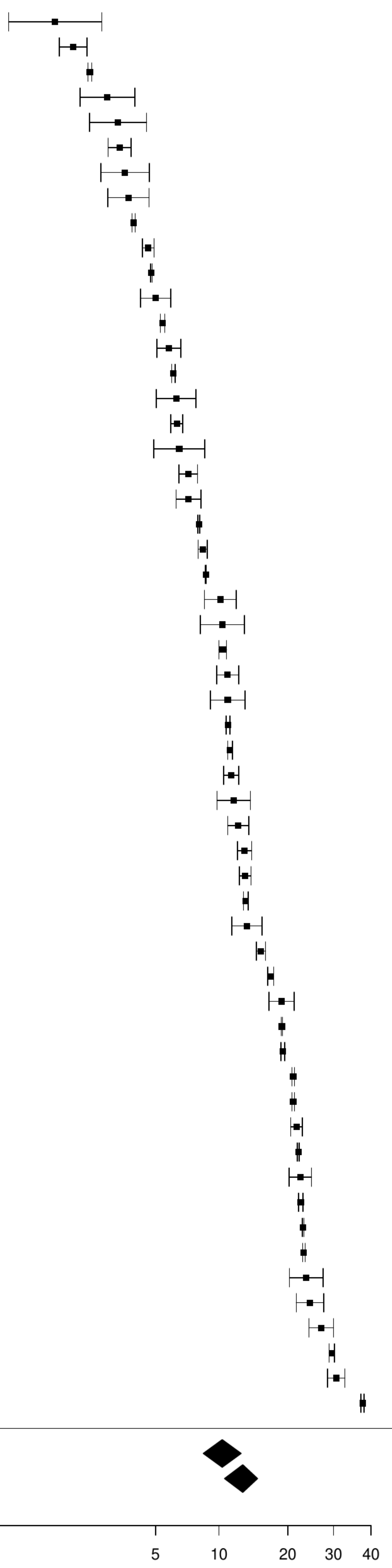

**Figure S46. Pooled prevalence of precious cancer in chronic heart failure**

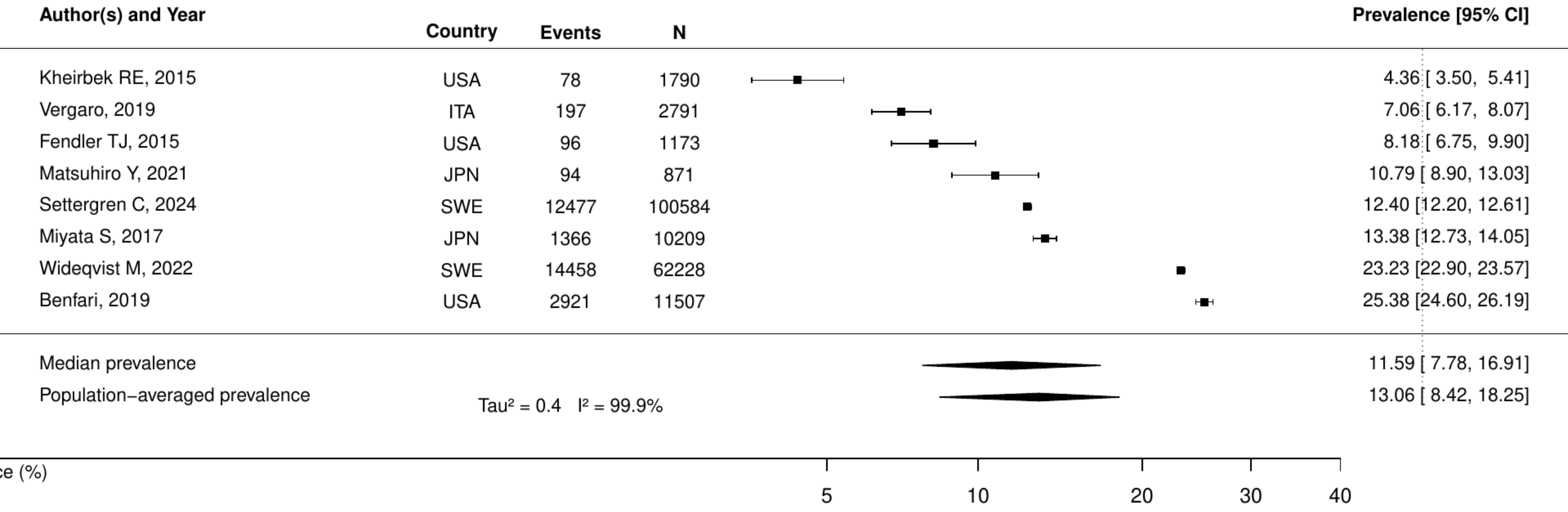

**Figure S47. Pooled prevalence of blood cancer in chronic heart failure**

| Author(s) and Year | Country | Events | N | Prevalence [95% CI] |
|---|---|---|---|---|
| Sessa, 2019 | ITA | 19 | 10091 | 0.19 [0.12, 0.29] |
| Schmidt M, 2016 | DEN | 3374 | 317161 | 1.06 [1.03, 1.10] |
| Cocchieri A, 2015 | ITA | 15 | 1192 | 1.26 [0.76, 2.08] |
| Gilstrap L, 2021 | USA | 4818 | 295494 | 1.63 [1.59, 1.68] |
| Median prevalence | | | | 0.80 [0.35, 1.83] |
| Population−averaged prevalence | | | | 1.11 [0.39, 1.63] |

$Tau^2 = 0.69$   $I^2 = 99.9\%$

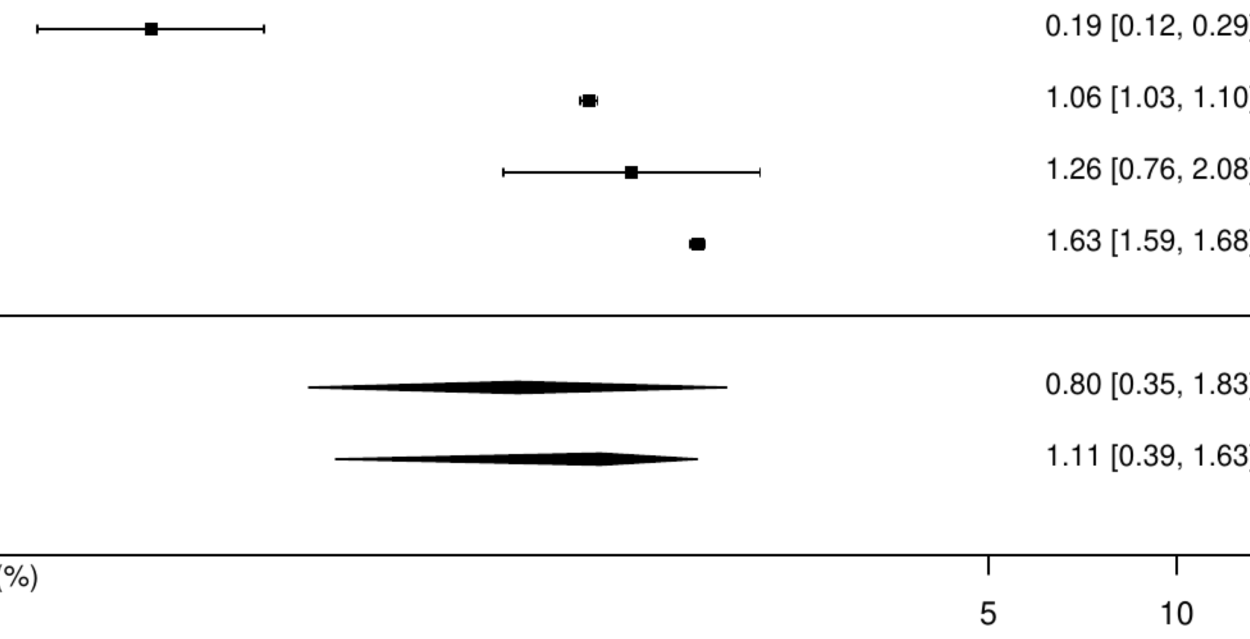

**Figure S48. Pooled prevalence of metastatic cancer in chronic heart failure**

| Author(s) and Year | Country | Events | N | Prevalence [95% CI] |
|---|---|---|---|---|
| Christiansen, 2020 | DEN | 1541 | 280002 | 0.55 [0.52, 0.58] |
| Wiskar K, 2017 | USA | 963 | 102746 | 0.94 [0.88, 1.00] |
| Gilstrap L, 2021 | USA | 4709 | 295494 | 1.59 [1.55, 1.64] |
| Kheirbek RE, 2016 | USA | 1072 | 60666 | 1.77 [1.67, 1.88] |
| Sun LY, 2020 | CAN | 6786 | 352329 | 1.93 [1.88, 1.97] |
| Muhlestein J B, 2016 | USA | 128 | 6414 | 2.00 [1.68, 2.37] |
| Cocchieri A, 2015 | ITA | 26 | 1192 | 2.18 [1.49, 3.18] |
| Chung, 2017 | KOR | 525 | 19128 | 2.74 [2.52, 2.99] |
| Chen CY, 2015 | USA | 666 | 23111 | 2.88 [2.67, 3.11] |
| McCullough, 2019 | USA | 300 | 8757 | 3.43 [3.06, 3.83] |
| Median prevalence | | | | 1.78 [1.28, 2.46] |
| Population−averaged prevalence | | | | 2.02 [1.47, 2.57] |

$Tau^2 = 0.28$ $I^2 = 99.7\%$

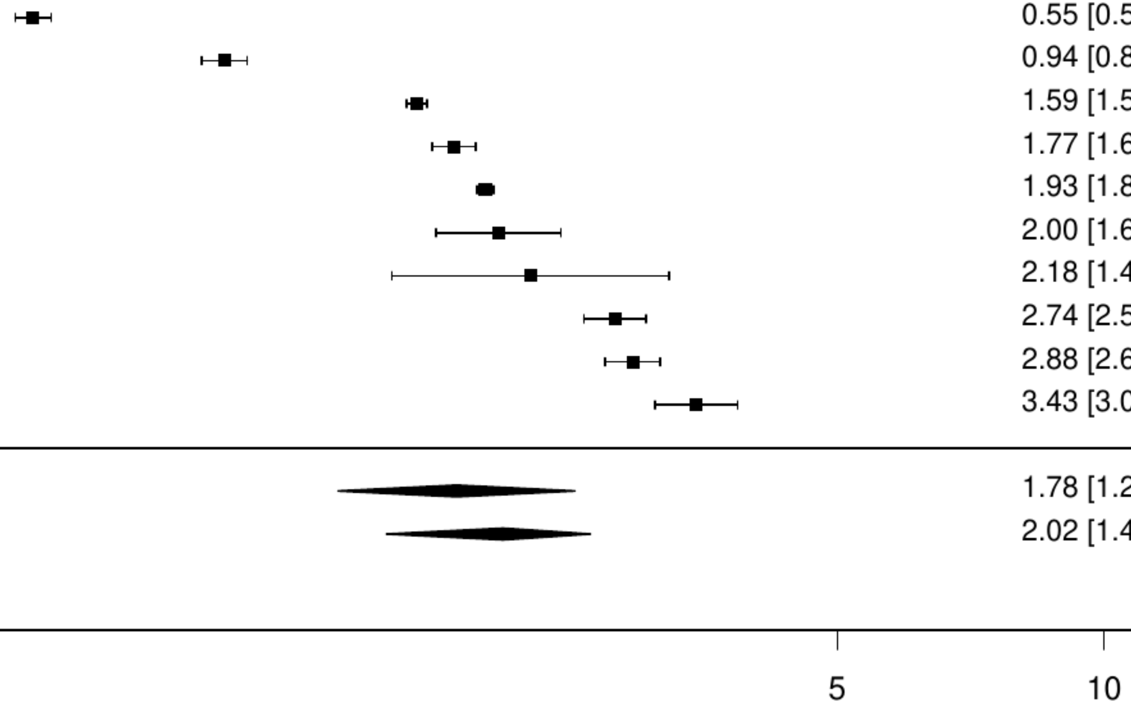

**Figure S49. Pooled prevalence of solid cancer in chronic heart failure**

| Author(s) and Year | Country | Events | N | Prevalence [95% CI] |
|---|---|---|---|---|
| Sessa, 2019 | ITA | 82 | 10091 | 0.81 [ 0.65, 1.01] |
| Schmidt M, 2016 | DEN | 20686 | 317161 | 6.52 [ 6.44, 6.61] |
| Hariri, 2022 | USA | 334 | 4921 | 6.79 [ 6.12, 7.52] |
| Gilstrap L, 2021 | USA | 26391 | 295494 | 8.93 [ 8.83, 9.03] |
| Parajuli DR, 2021 | AUS | 176 | 1184 | 14.86 [12.95, 17.01] |
| Chung, 2017 | KOR | 2892 | 19128 | 15.12 [14.62, 15.63] |
| Median prevalence | | | | 6.63 [ 3.02, 13.94] |
| Population−averaged prevalence | | | | 9.46 [ 5.08, 12.59] |

$Tau^2 = 1.06$   $I^2 = 100\%$

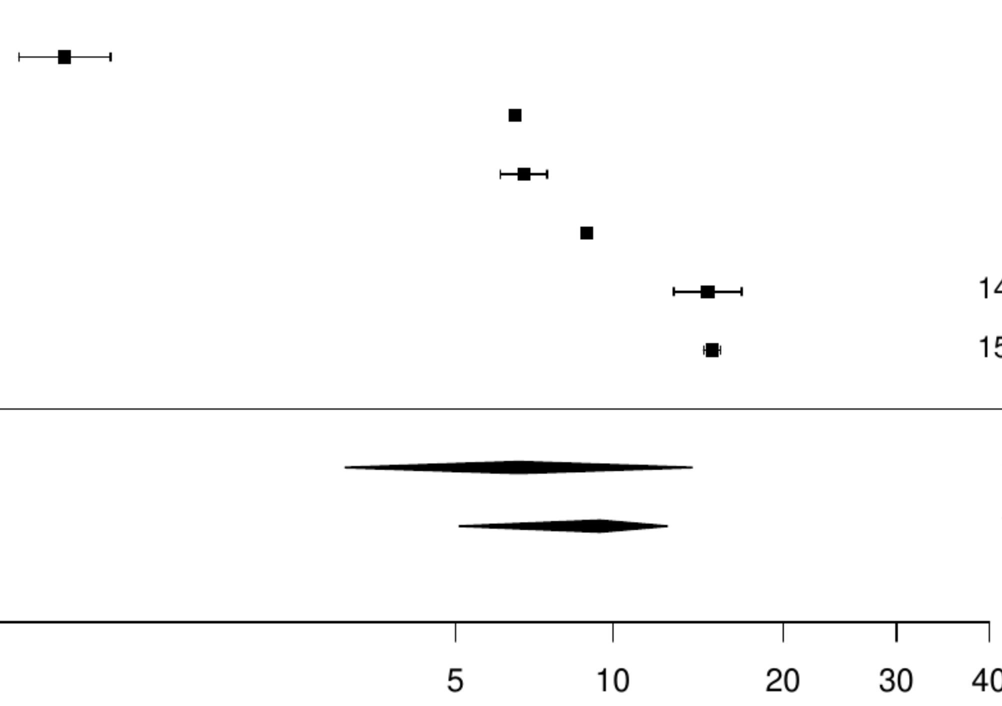

### Figure S50. Pooled prevalence of any cancer in chronic heart failure with reduced ejection fraction

| Author(s) and Year | Country | Events | N | Prevalence [95% CI] |
|---|---|---|---|---|
| Chen YL, 2017 | TW | 38 | 1311 | 2.90 [ 2.12, 3.96] |
| Lam CS, 2016 | Multinational | 162 | 5276 | 3.07 [ 2.64, 3.57] |
| Rivera−Toquica, 2024 | Colombia | 49 | 1384 | 3.54 [ 2.69, 4.65] |
| Kormos, 2019 | USA | 853 | 18539 | 4.60 [ 4.31, 4.91] |
| Braga JR, 2019 | CAN | 811 | 12814 | 6.33 [ 5.92, 6.76] |
| Song HK, 2016 | USA | 205 | 2527 | 8.11 [ 7.11, 9.24] |
| Jawitz, 2020 | USA | 1537 | 18307 | 8.40 [ 8.00, 8.81] |
| Sicras−Mainar, 2022 | ESP | 2055 | 19762 | 10.40 [ 9.98, 10.83] |
| Wienbergen H, 2019 | Multinational | 104 | 949 | 10.96 [ 9.12, 13.11] |
| Vaduganathan M, 2020 | USA | 565 | 4970 | 11.37 [10.52, 12.28] |
| Iorio, 2018 | ITA | 113 | 941 | 12.01 [10.08, 14.25] |
| Schwartz B, 2021 | DEN | 5466 | 39260 | 13.92 [13.58, 14.27] |
| Kapelios CJ, 2021 | SWE | 5244 | 36076 | 14.54 [14.18, 14.90] |
| Median prevalence | | | | 7.52 [ 5.56, 10.09] |
| Population−averaged prevalence | | | | 8.52 [ 6.29, 10.58] |

Tau² = 0.35 I² = 99.7%

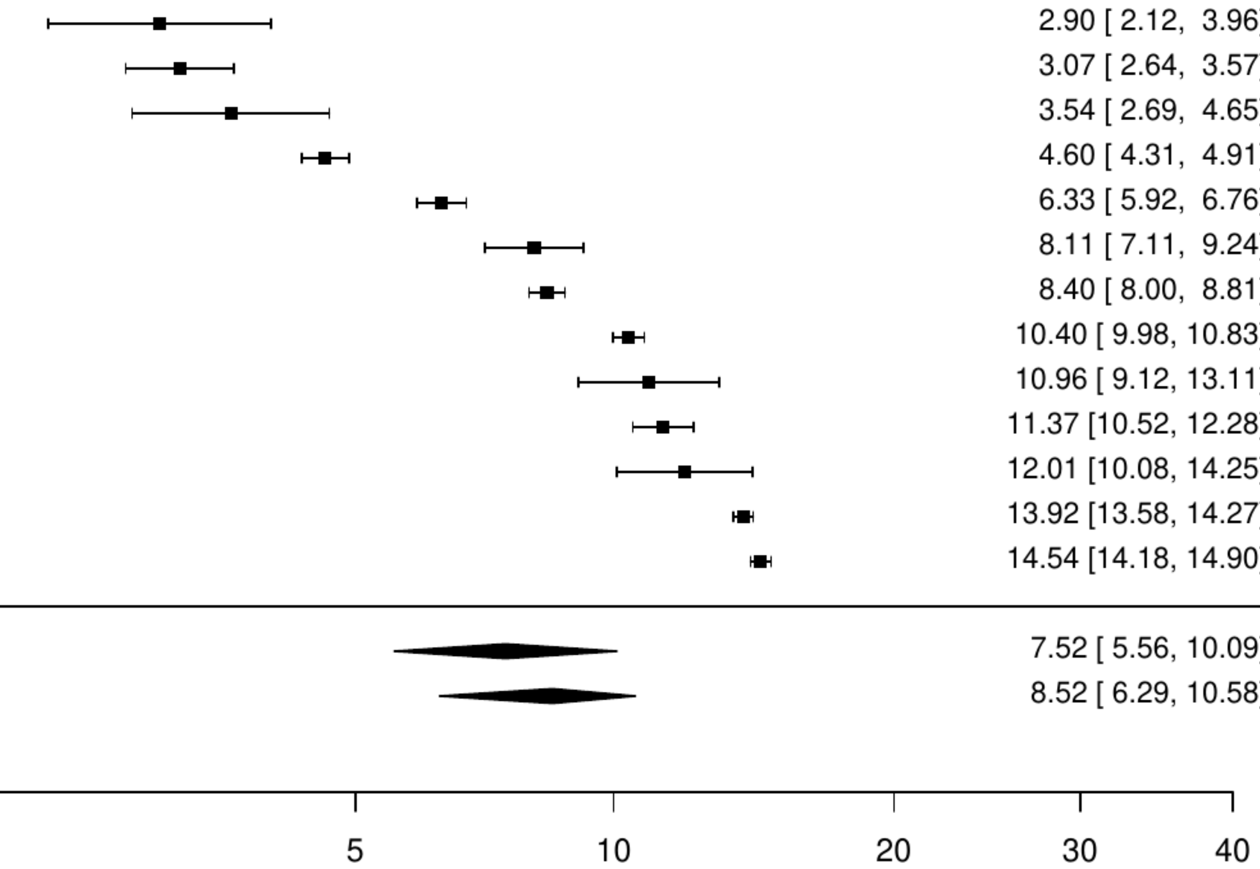

### Figure S51. Pooled prevalence of previous cancer in chronic heart failure with preserved ejection fraction

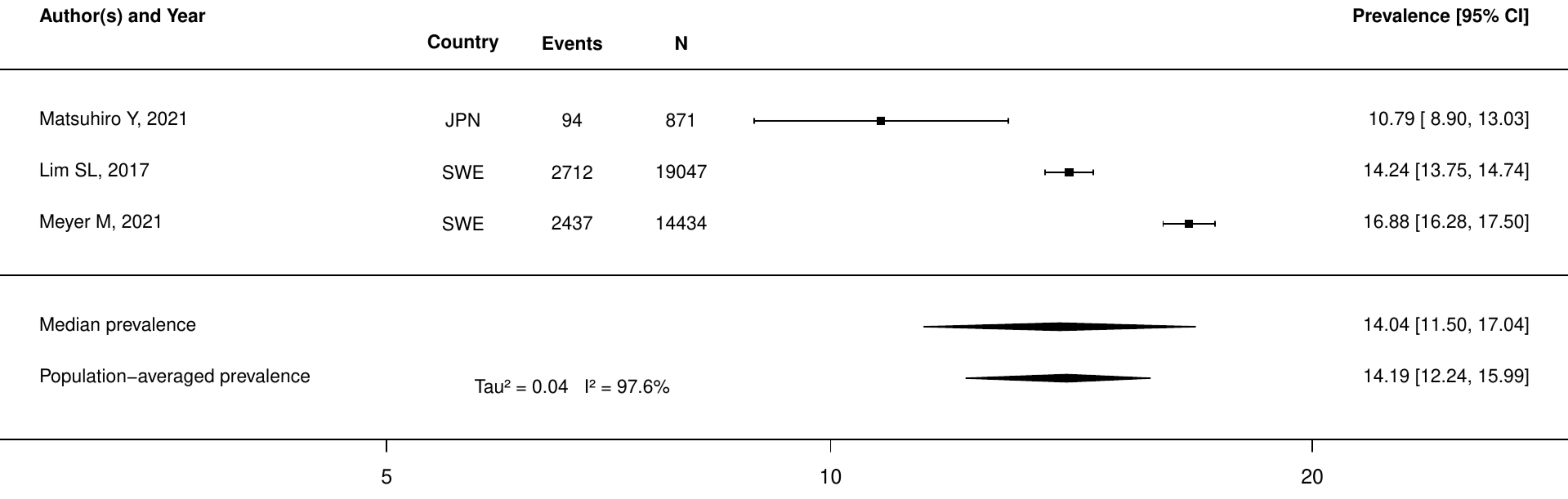

**Figure S52. Pooled prevalence of any cancer in chronic heart failure with preserved ejection fraction**

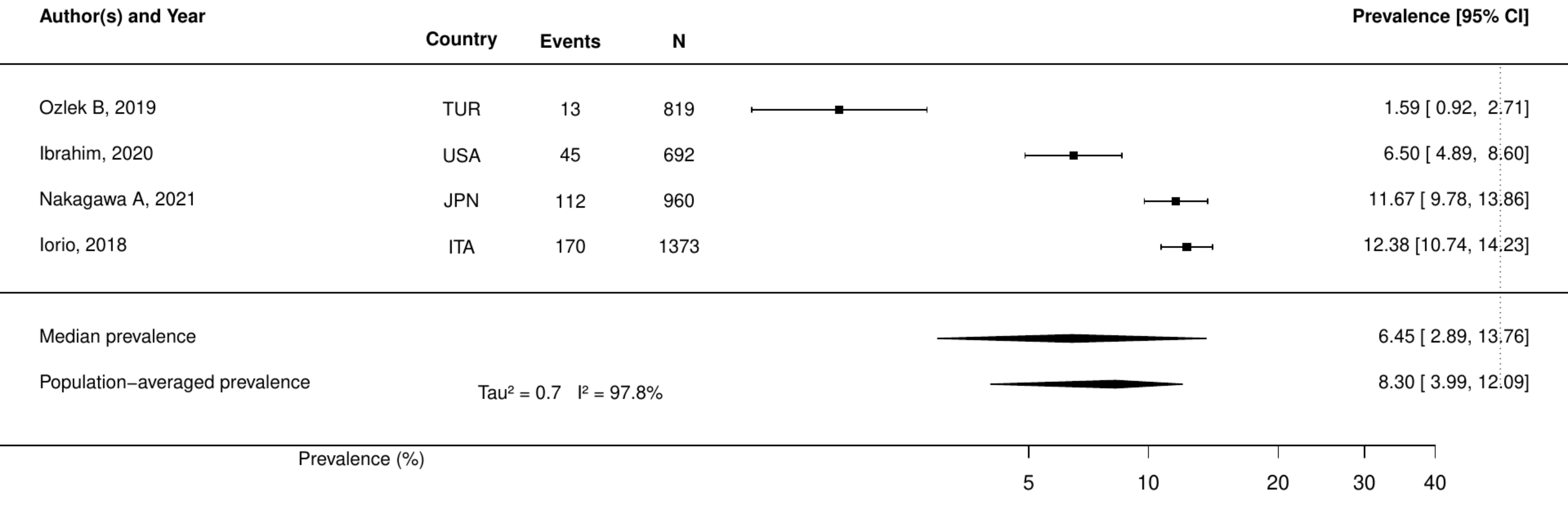

**Figure S53. Pooled prevalence of active cancer in acute heart failure.**

| Author(s) and Year | Country | Events | N | Prevalence [95% CI] |
|---|---|---|---|---|
| Mebazaa, 2018 | Multinational | 276 | 5387 | 5.12 [ 4.57, 5.75] |
| Sax Dana R, 2021 | USA | 2855 | 26189 | 10.90 [10.53, 11.28] |
| Miró, 2017 | ESP | 859 | 6516 | 13.18 [12.38, 14.03] |
| Median prevalence | | | | 9.12 [ 5.73, 14.22] |
| Population−averaged prevalence | | | | 9.75 [ 7.03, 12.36] |

Tau² = 0.19 I² = 99.3%

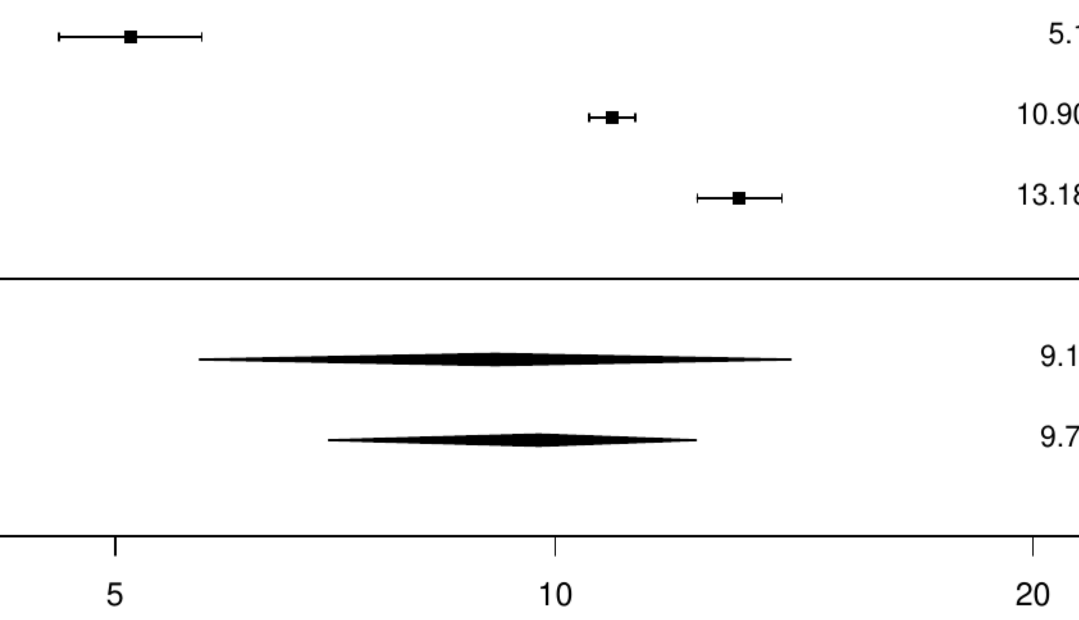

**Figure S54. Pooled prevalence of any cancer in acute heart failure.**

| Author(s) and Year | Country | Events | N | Prevalence [95% CI] |
|---|---|---|---|---|
| Nakamaru, 2023 | JPN | 53 | 3498 | 1.52 [ 1.16, 1.98] |
| Chen YL, 2021 | TW | 37 | 1275 | 2.90 [ 2.11, 3.98] |
| Bistola V, 2018 | Multinational | 167 | 4953 | 3.37 [ 2.90, 3.91] |
| Wang W, 2024 | CHN | 62 | 1454 | 4.26 [ 3.34, 5.43] |
| Chioncel O, 2019 | Multinational | 385 | 7865 | 4.90 [ 4.44, 5.39] |
| Gerhardt, 2023 | Multinational | 998 | 18528 | 5.39 [ 5.07, 5.72] |
| Kaneko H, 2021 | JPN | 25202 | 432620 | 5.83 [ 5.76, 5.90] |
| Braga JR, 2019 | CAN | 811 | 12814 | 6.33 [ 5.92, 6.76] |
| Park JJ, 2017 | KOR | 352 | 4370 | 8.05 [ 7.28, 8.90] |
| Park, 2023 | KOR | 2498 | 29290 | 8.53 [ 8.21, 8.85] |
| Crespo−Leiro MG, 2020 | ESP | 82 | 881 | 9.31 [ 7.56, 11.41] |
| Kamiya K, 2020 | JPN | 336 | 3277 | 10.25 [ 9.26, 11.34] |
| Miles JA, 2019 | USA | 469 | 4343 | 10.80 [ 9.91, 11.76] |
| Trullàs JC, 2016 | ESP | 363 | 3287 | 11.04 [10.02, 12.16] |
| Enzan N, 2023 | JPN | 1162 | 10473 | 11.10 [10.51, 11.71] |
| Nakagawa A, 2021 | JPN | 112 | 960 | 11.67 [ 9.78, 13.86] |
| De Sutter J, 2015 | BEL | 108 | 904 | 11.95 [ 9.99, 14.23] |
| Rossello, 2019 | ESP | 1149 | 9098 | 12.63 [11.96, 13.33] |
| Alsawas M, 2019 | USA | 1566 | 12265 | 12.77 [12.19, 13.37] |
| Llorens P, 2018 | ESP | 1676 | 12308 | 13.62 [13.02, 14.23] |
| Martin−Sanchez FJ, 2017 | ESP | 66 | 465 | 14.19 [11.31, 17.67] |
| Iguchi M, 2021 | JPN | 585 | 4056 | 14.42 [13.38, 15.54] |
| Funabashi S, 2020 | JPN | 108 | 708 | 15.25 [12.79, 18.09] |
| Al−Gobari M, 2019 | FRA | 333 | 2032 | 16.39 [14.84, 18.06] |
| Martin−Sanchez FJ, 2017 | ESP | 98 | 596 | 16.44 [13.68, 19.64] |
| Coles Andrew H, 2015 | USA | 828 | 4025 | 20.57 [19.35, 21.85] |
| Median prevalence | | | | 8.87 [ 7.04, 11.12] |
| Population−averaged prevalence | Tau² = 0.42 I² = 99.7% |  |  | 10.22 [ 8.49, 11.94] |

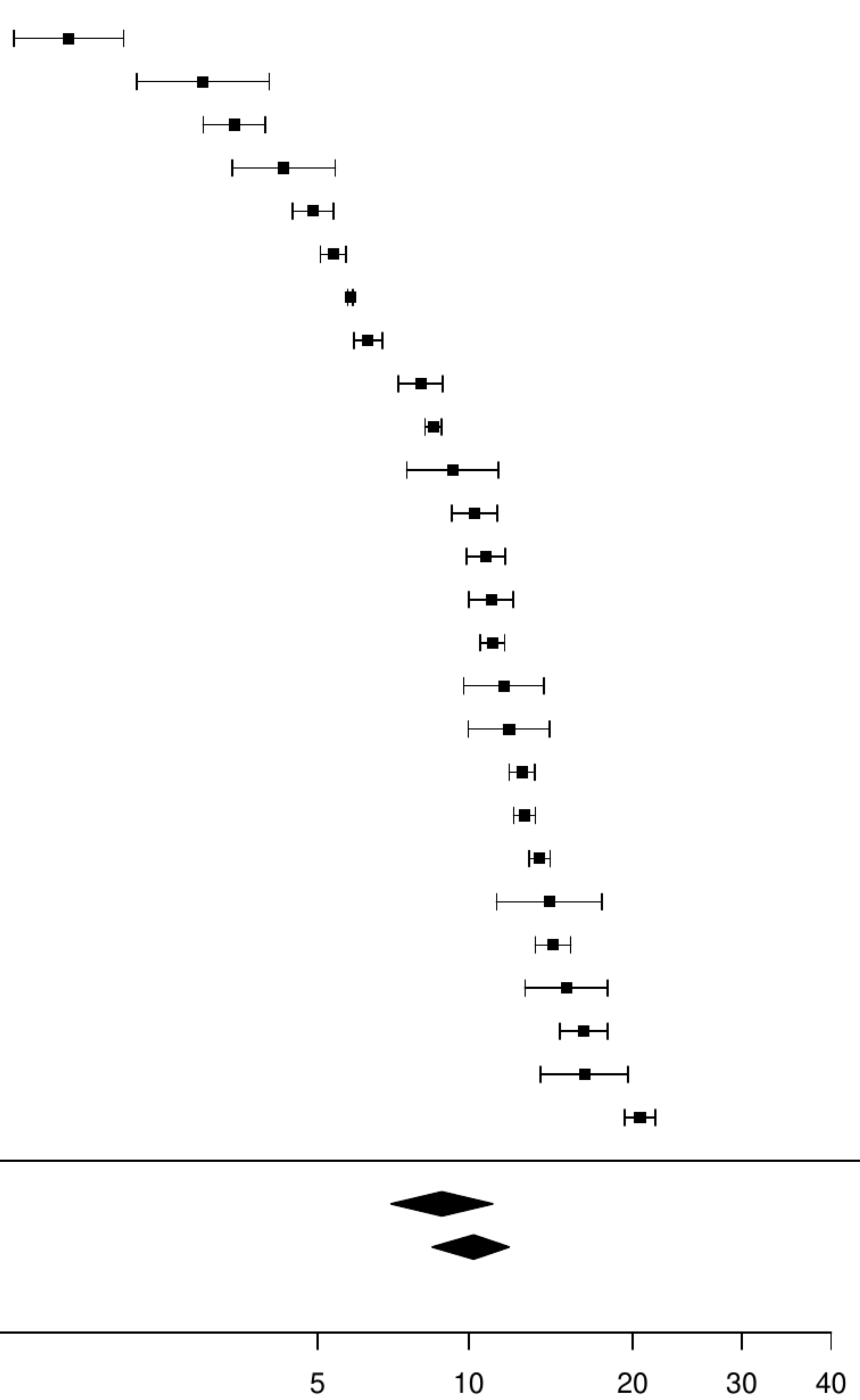

**Figure S55. Pooled prevalence of previous cancer in acute heart failure.**

| Author(s) and Year | Country | Events | N | Prevalence [95% CI] |
|---|---|---|---|---|
| Anguita−Gamez M, 2024 | ESP | 7693 | 354786 | 2.17 [ 2.12, 2.22] |
| Liao CT, 2021 | TW | 83 | 1630 | 5.09 [ 4.12, 6.27] |
| Targher Giovanni, 2016 | ITA | 108 | 1776 | 6.08 [ 5.06, 7.29] |
| Lee SE, 2020 | KOR | 45 | 717 | 6.28 [ 4.72, 8.30] |
| Okano T, 2020 | JPN | 58 | 676 | 8.58 [ 6.69, 10.94] |
| Kubo, 2018 | JPN | 92 | 563 | 16.34 [13.51, 19.63] |
| Median prevalence | | | | 6.25 [ 3.79, 10.14] |
| Population−averaged prevalence | | | | 7.33 [ 4.28, 11.24] |

$Tau^2 = 0.42$ $I^2 = 98.3\%$

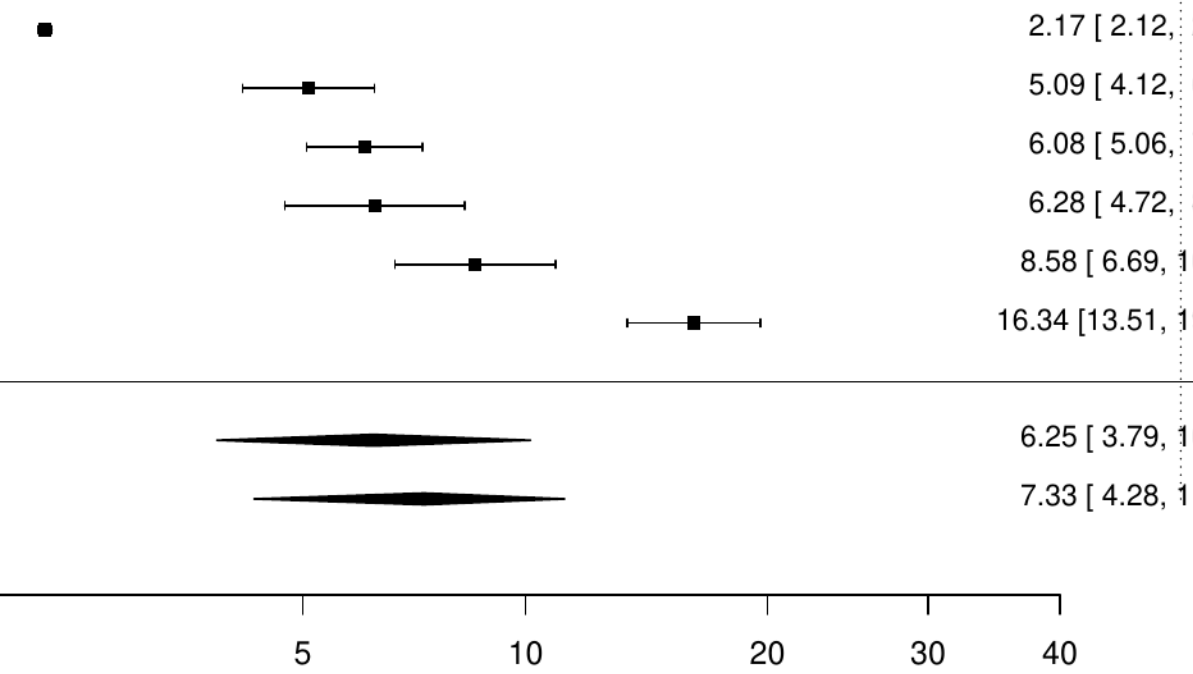

**Figure S56. Pooled prevalence of metastatic cancer in acute heart failure.**

| Author(s) and Year | Country | Events | N | Prevalence [95% CI] |
|---|---|---|---|---|
| Lagu Tara, 2016 | USA | 120 | 13163 | 0.91 [0.76, 1.09] |
| Sun LY, 2019 | CAN | 1283 | 82125 | 1.56 [1.48, 1.65] |
| Arslanian−Engoren C, 2018 | USA | 243 | 9548 | 2.55 [2.25, 2.88] |
| Median prevalence | | | | 1.54 [0.96, 2.46] |
| Population−averaged prevalence | | | | 1.67 [1.14, 2.20] |

$Tau^2 = 0.17$   $I^2 = 98.1\%$

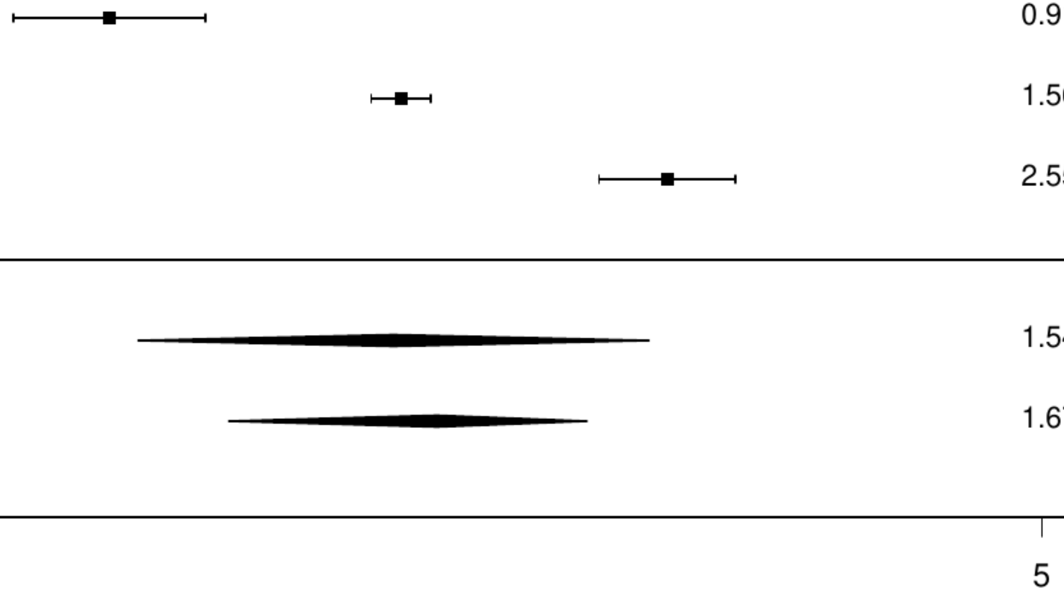

**Figure S57. Pooled prevalence of active cancer in any acute stroke.**

| Author(s) and Year | Country | Events | N | Prevalence [95% CI] |
|---|---|---|---|---|
| Kim, 2020 | USA | 146 | 27739 | 0.53 [ 0.45, 0.62] |
| Aarnio K, 2016 | FIN | 11 | 970 | 1.13 [ 0.63, 2.04] |
| Atterman A, 2020 | SWE | 1518 | 52471 | 2.89 [ 2.75, 3.04] |
| Bejot Y, 2020 | FRA | 45 | 1048 | 4.29 [ 3.22, 5.70] |
| Yamaguchi, 2019 | JPN | 61 | 1012 | 6.03 [ 4.72, 7.67] |
| Gon Y, 2020 | JPN | 185 | 1570 | 11.78 [10.28, 13.47] |
| Median prevalence | | | | 2.89 [ 1.23, 6.60] |
| Population−averaged prevalence | | | | 4.60 [ 1.72, 8.13] |

$Tau^2 = 1.15$  $I^2 = 99.4\%$

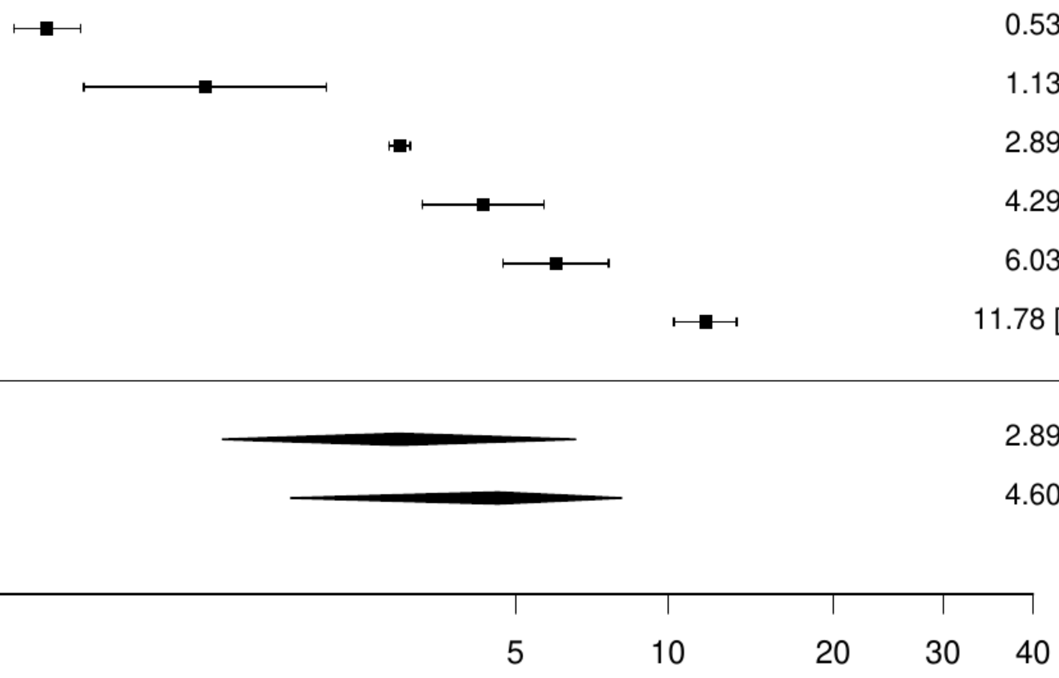

**Figure S58. Pooled prevalence of any cancer in any acute stroke.**

| Author(s) and Year | Country | Events | N | Prevalence [95% CI] |
|---|---|---|---|---|
| Matuja SS, 2024 | Tanzania | 7 | 566 | 1.24 [ 0.59, 2.57] |
| Zhang X, 2018 | CHN | 215 | 12027 | 1.79 [ 1.57, 2.04] |
| Jones EM, 2020 | USA | 20 | 897 | 2.23 [ 1.44, 3.43] |
| Tanaka K, 2020 | Multinational | 30 | 1165 | 2.58 [ 1.81, 3.66] |
| Pandey AS, 2016 | USA | 892 | 33191 | 2.69 [ 2.52, 2.87] |
| Momosaki R, 2016 | JPN | 2372 | 77890 | 3.05 [ 2.93, 3.17] |
| Xian Y, 2017 | USA | 1035 | 31775 | 3.26 [ 3.07, 3.46] |
| Hall, 2019 | CAN | 274 | 6988 | 3.92 [ 3.49, 4.40] |
| Bergman EM, 2015 | SWE | 109 | 2599 | 4.19 [ 3.49, 5.04] |
| Guijarro, 2016 | ESP | 22133 | 495427 | 4.47 [ 4.41, 4.53] |
| Pommier T, 2024 | FRA | 42 | 916 | 4.59 [ 3.41, 6.15] |
| Kaufman BG, 2020 | USA | 5025 | 109420 | 4.59 [ 4.47, 4.72] |
| Zhou LW, 2023 | USA | 13295 | 273811 | 4.86 [ 4.78, 4.94] |
| Correia PN, 2021 | CHE | 183 | 3530 | 5.18 [ 4.50, 5.97] |
| Rea, 2019 | ITA | 597 | 10878 | 5.49 [ 5.08, 5.93] |
| Gardener H, 2019 | USA | 1181 | 16952 | 6.97 [ 6.59, 7.36] |
| Huang HK, 2019 | TW | 4796 | 66727 | 7.19 [ 6.99, 7.39] |
| Liljehult J, 2020 | DEN | 75 | 1031 | 7.27 [ 5.84, 9.03] |
| Gardener, 2019 | USA | 4277 | 58435 | 7.32 [ 7.11, 7.53] |
| Corraini, 2016 | DEN | 16283 | 201025 | 8.10 [ 7.98, 8.22] |
| Saltman AP, 2015 | CAN | 2449 | 29810 | 8.22 [ 7.91, 8.53] |
| Koton S, 2016 | ISR | 552 | 6177 | 8.94 [ 8.25, 9.67] |
| Zhao Y, 2016 | AUS | 334 | 3733 | 8.95 [ 8.07, 9.91] |
| Barlas RS, 2018 | UK | 334 | 3575 | 9.34 [ 8.43, 10.34] |
| Reid JM, 2017 | UK | 58 | 575 | 10.09 [ 7.88, 12.83] |
| Kilkenny MF, 2021 | AUS | 1688 | 15468 | 10.91 [10.43, 11.41] |
| Appelros, 2017 | SWE | 2564 | 23356 | 10.98 [10.58, 11.39] |
| Yasaka, 2018 | JPN | 1105 | 9804 | 11.27 [10.66, 11.91] |
| Wilbers, 2020 | NLD | 321 | 2736 | 11.73 [10.58, 12.99] |
| Phan HT, 2022 | USA | 165 | 1334 | 12.37 [10.71, 14.25] |
| Forti P, 2020 | ITA | 328 | 2428 | 13.51 [12.21, 14.93] |
| Wireklint Sundstrm, 2015 | SWE | 193 | 1376 | 14.03 [12.29, 15.96] |
| Yousufuddin Mohammed, 2017 | USA | 983 | 6771 | 14.52 [13.70, 15.38] |
| Labberton, 2019 | NOR | 129 | 817 | 15.79 [13.45, 18.45] |
| Renoux C, 2017 | UK | 449 | 2553 | 17.59 [16.16, 19.11] |
| Akyea, 2021 | UK | 5627 | 30702 | 18.33 [17.90, 18.76] |
| Gamble DT, 2020 | UK | 373 | 1928 | 19.35 [17.64, 21.17] |
| Beuker C, 2023 | GER | 4007 | 18506 | 21.65 [21.06, 22.25] |
| Aparicio HJ, 2022 | USA | 13360 | 29690 | 45.00 [44.43, 45.56] |
| Median prevalence | | | | 7.57 [ 5.95, 9.59] |
| Population−averaged prevalence | | | | 9.52 [ 7.33, 12.19] |

Tau² = 0.67   I² = 99.9%

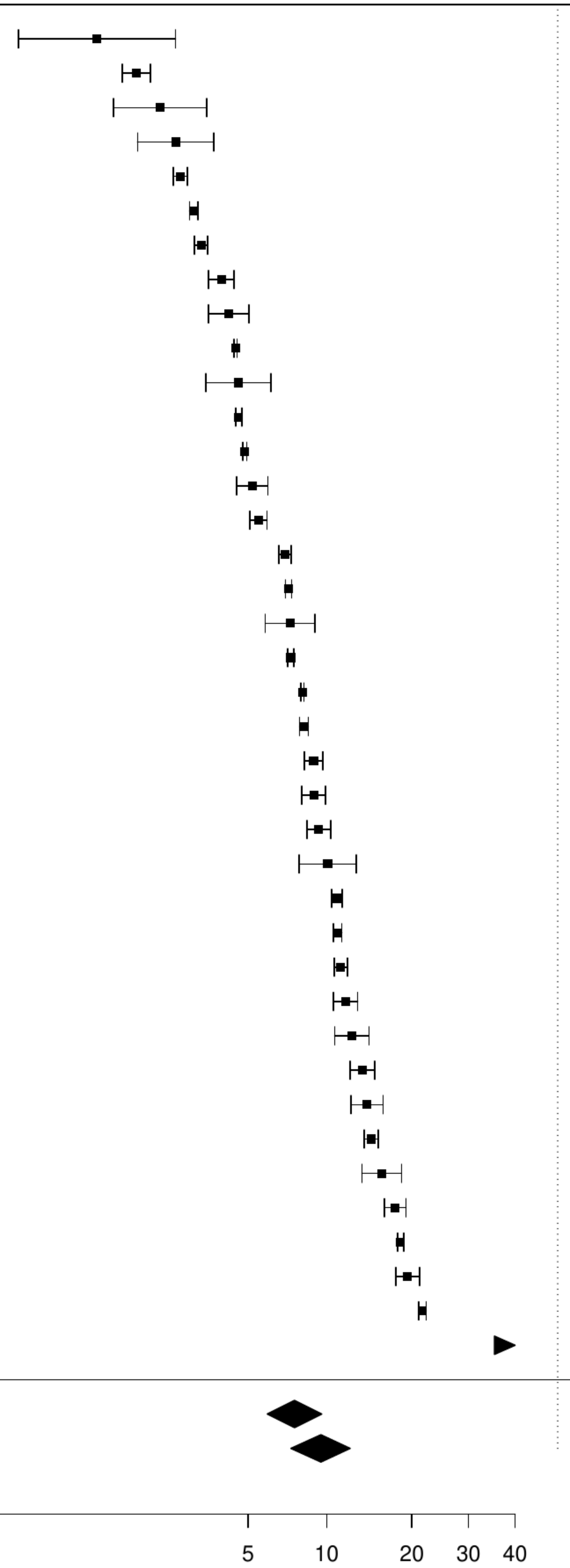

**Figure S59. Pooled prevalence of previous cancer in any acute stroke.**

| Author(s) and Year | Country | Events | N | Prevalence [95% CI] |
|---|---|---|---|---|
| Casetta I, 2020 | ITA | 171 | 3045 | 5.62 [ 4.85, 6.49] |
| Kristiansen MH, 2023 | DEN | 64 | 538 | 11.90 [ 9.42, 14.91] |
| Tybjerg AJ, 2020 | DEN | 11407 | 85893 | 13.28 [13.06, 13.51] |
| O'Neal WT, 2015 | USA | 2248 | 15428 | 14.57 [14.02, 15.14] |
| Ganesh A, 2017 | UK | 221 | 1425 | 15.51 [13.72, 17.48] |
| Bisson A, 2018 | FRA | 40456 | 240459 | 16.82 [16.68, 16.97] |
| Reznik ME, 2018 | USA | 189 | 1003 | 18.84 [16.54, 21.38] |
| Median prevalence | | | | 13.15 [10.03, 17.06] |
| Population−averaged prevalence | | | | 13.81 [10.90, 16.60] |

$Tau^2 = 0.17$ $I^2 = 99.8\%$

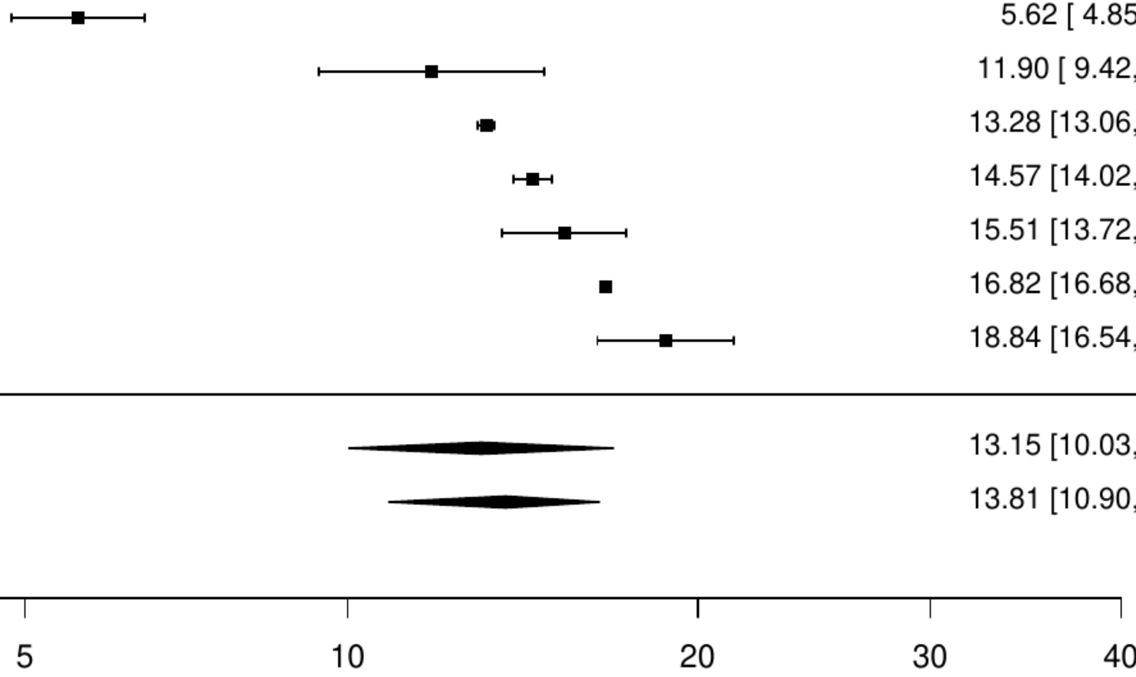

**Figure S60. Pooled prevalence of blood cancer in any acute stroke.**

| Author(s) and Year | Country | Events | N | Prevalence [95% CI] |
|---|---|---|---|---|
| Lindgren A, 2019 | Multinational | 9 | 8525 | 0.11 [0.05, 0.20] |
| Si, 2019 | CHN | 35 | 10331 | 0.34 [0.24, 0.47] |
| Hreha, 2020 | USA | 1369 | 269314 | 0.51 [0.48, 0.54] |
| Corraini P, 2018 | DEN | 1242 | 201691 | 0.62 [0.58, 0.65] |
| Ripley Diane C Cowper, 2015 | USA | 101 | 10430 | 0.97 [0.80, 1.18] |
| Downer, 2024 | UK | 31 | 2454 | 1.26 [0.89, 1.79] |
| Median prevalence | | | | 0.50 [0.27, 0.92] |
| Population−averaged prevalence | | | | 0.65 [0.33, 0.94] |

$Tau^2 = 0.56$   $I^2 = 99.4\%$

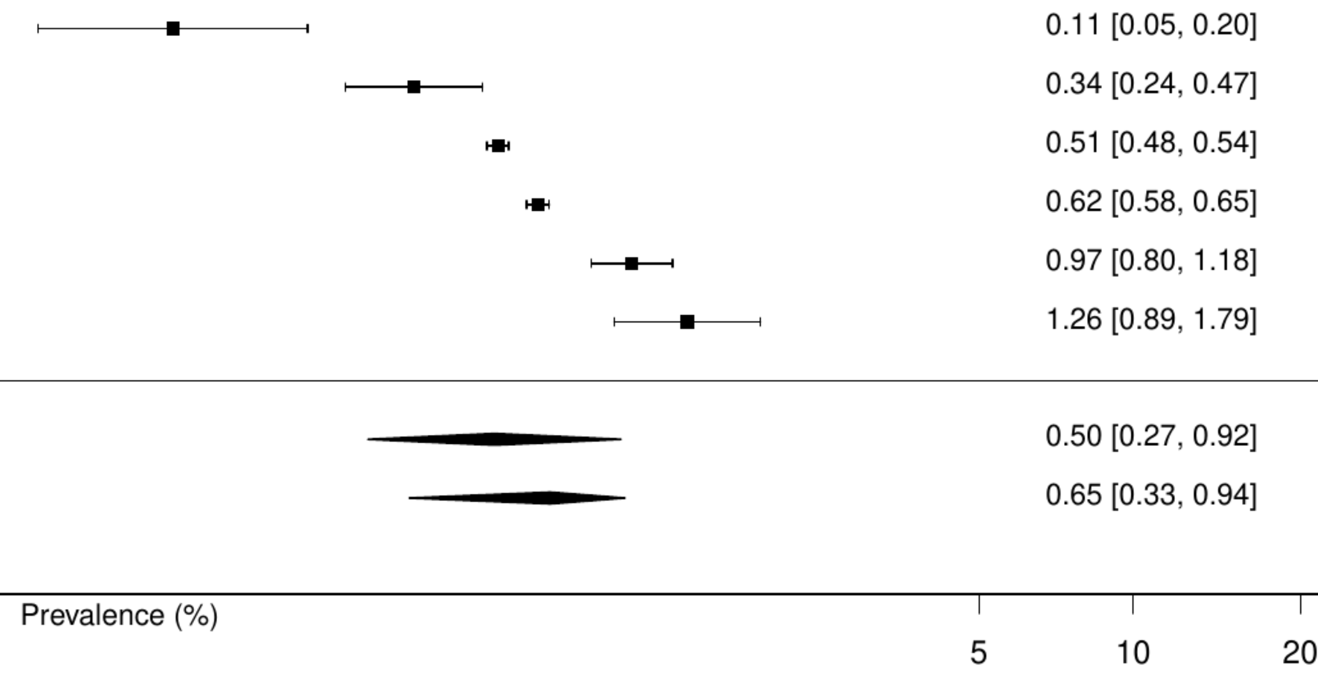

**Figure S61. Pooled prevalence of metastatic cancer in any acute stroke.**

| Author(s) and Year | Country | Events | N | Prevalence [95% CI] |
|---|---|---|---|---|
| Lindgren A, 2019 | Multinational | 15 | 8525 | 0.18 [0.11, 0.29] |
| Corraini P, 2018 | DEN | 1422 | 201691 | 0.71 [0.67, 0.74] |
| Castro, 2017 | BRA | 7 | 959 | 0.73 [0.35, 1.52] |
| Hreha, 2020 | USA | 2268 | 269314 | 0.84 [0.81, 0.88] |
| Downer, 2024 | UK | 26 | 2454 | 1.06 [0.72, 1.55] |
| Si, 2019 | CHN | 129 | 10331 | 1.25 [1.05, 1.48] |
| Hall, 2019 | CAN | 116 | 6988 | 1.66 [1.39, 1.99] |
| Kim, 2020 | USA | 472 | 27739 | 1.70 [1.56, 1.86] |
| Ripley Diane C Cowper, 2015 | USA | 225 | 10430 | 2.16 [1.90, 2.45] |
| Median prevalence | | | | 0.96 [0.62, 1.51] |
| Population−averaged prevalence | | | | 1.19 [0.80, 1.57] |

$Tau^2 = 0.45$ $I^2 = 99.4\%$

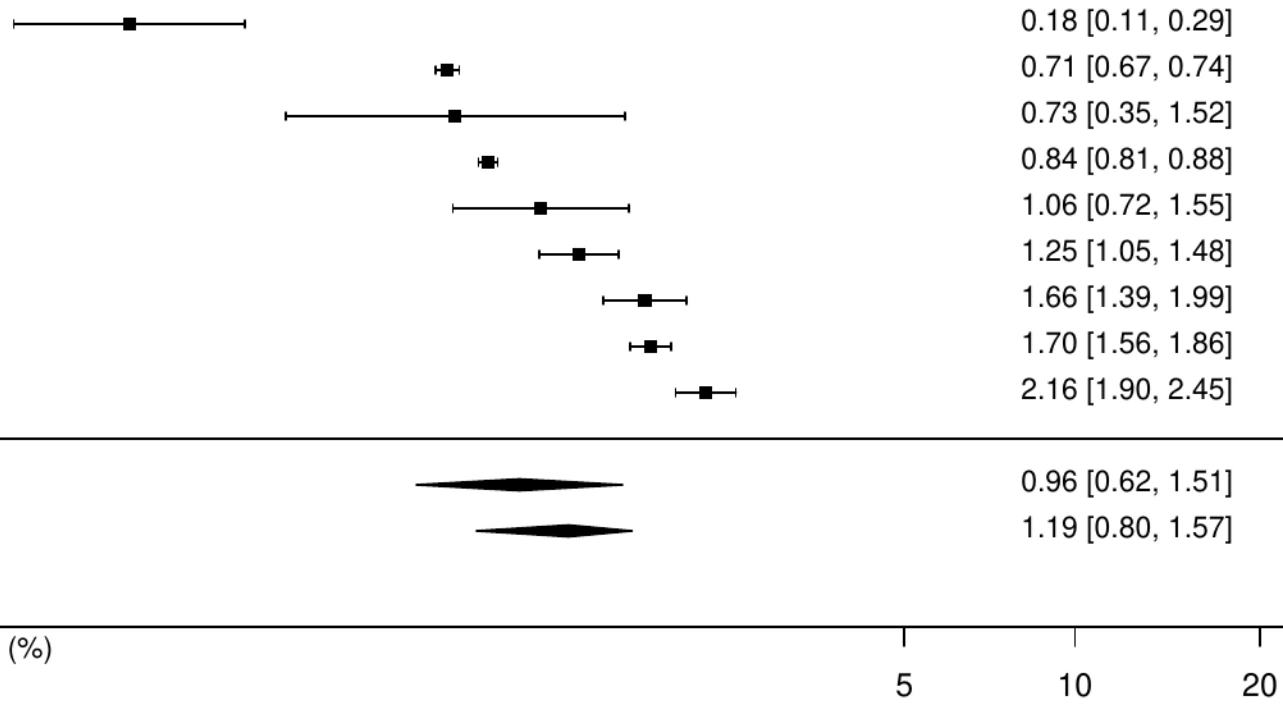

**Figure S62. Pooled prevalence of solid cancer in any acute stroke.**

| Author(s) and Year | Country | Events | N | Prevalence [95% CI] |
|---|---|---|---|---|
| Lindgren A, 2019 | Multinational | 58 | 8525 | 0.68 [ 0.53, 0.88] |
| Hreha, 2020 | USA | 6626 | 269314 | 2.46 [ 2.40, 2.52] |
| Castro, 2017 | BRA | 25 | 959 | 2.61 [ 1.77, 3.83] |
| Si, 2019 | CHN | 368 | 10331 | 3.56 [ 3.22, 3.94] |
| Corraini P, 2018 | DEN | 16652 | 201691 | 8.26 [ 8.14, 8.38] |
| Downer, 2024 | UK | 339 | 2454 | 13.81 [12.50, 15.24] |
| Ripley Diane C Cowper, 2015 | USA | 1760 | 10430 | 16.87 [16.17, 17.61] |
| Median prevalence | | | | 4.49 [ 2.03, 9.65] |
| Population−averaged prevalence | | | | 7.02 [ 2.86, 11.57] |

Tau² = 1.22 I² = 100%

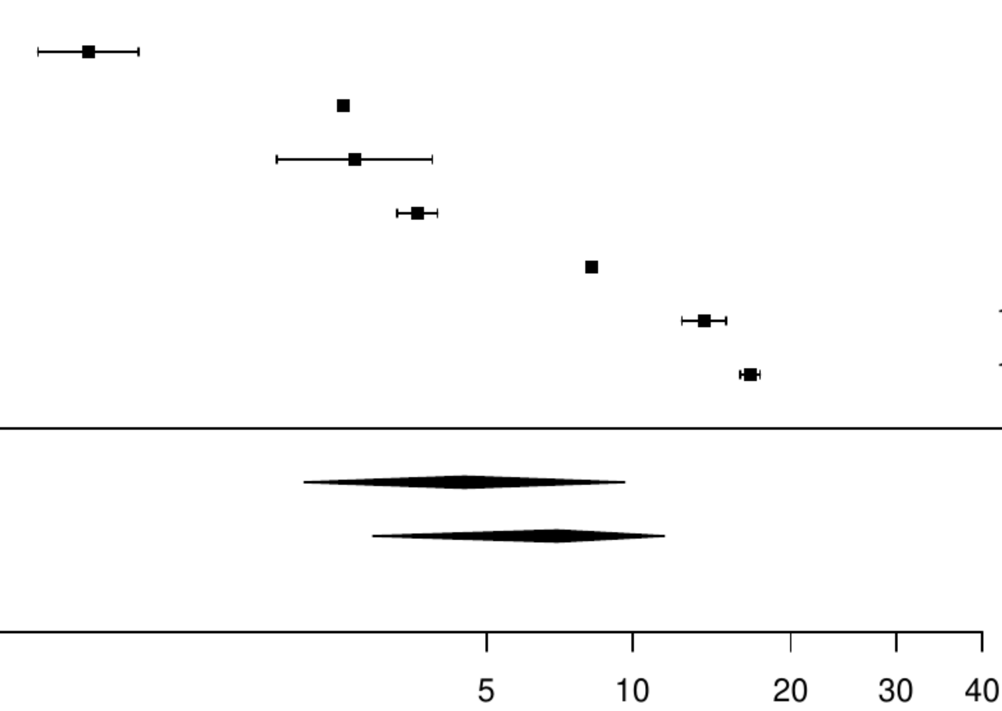

**Figure S63. Pooled prevalence of active cancer in ischemic stroke.**

| Author(s) and Year | Country | Events | N | Prevalence [95% CI] |
|---|---|---|---|---|
| Aarnio K, 2016 | FIN | 11 | 970 | 1.13 [ 0.63, 2.04] |
| Atterman A, 2020 | SWE | 1518 | 52471 | 2.89 [ 2.75, 3.04] |
| Bejot Y, 2020 | FRA | 45 | 1048 | 4.29 [ 3.22, 5.70] |
| Yamaguchi, 2019 | JPN | 61 | 1012 | 6.03 [ 4.72, 7.67] |
| Gon Y, 2020 | JPN | 185 | 1570 | 11.78 [10.28, 13.47] |
| Median prevalence | | | | 4.10 [ 2.08, 7.94] |
| Population−averaged prevalence | | | | 5.26 [ 2.40, 8.91] |

Tau² = 0.61   I² = 98.7%

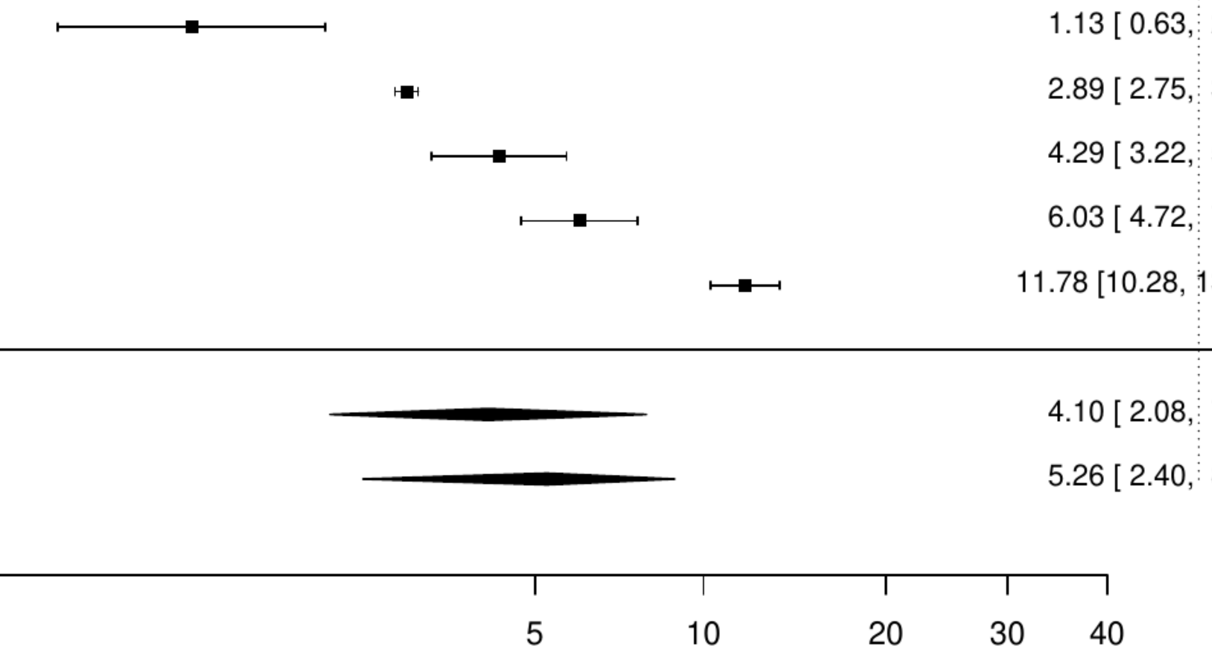

### Figure S64. Pooled prevalence of any cancer in ischemic stroke.

| Author(s) and Year | Country | Events | N | Prevalence [95% CI] |
|---|---|---|---|---|
| Hall RE, 2015 | CAN | 954 | 73368 | 1.30 [ 1.22, 1.38] |
| Sipilä JO, 2016 | FIN | 232 | 14834 | 1.56 [ 1.38, 1.78] |
| Xu J, 2016 | CHN | 174 | 9698 | 1.79 [ 1.55, 2.08] |
| Jones EM, 2020 | USA | 20 | 897 | 2.23 [ 1.44, 3.43] |
| Giang KW, 2016 | SWE | 414 | 17149 | 2.41 [ 2.19, 2.65] |
| Momosaki R, 2016 | JPN | 160 | 6153 | 2.60 [ 2.23, 3.03] |
| Pandey AS, 2016 | USA | 892 | 33191 | 2.69 [ 2.52, 2.87] |
| Yeh SJ, 2021 | TW | 1569 | 50092 | 3.13 [ 2.98, 3.29] |
| Xian Y, 2017 | USA | 1035 | 31775 | 3.26 [ 3.07, 3.46] |
| Guijarro, 2016 | ESP | 22133 | 495427 | 4.47 [ 4.41, 4.53] |
| Pommier T, 2024 | FRA | 42 | 916 | 4.59 [ 3.41, 6.15] |
| Zhou LW, 2023 | USA | 13295 | 273811 | 4.86 [ 4.78, 4.94] |
| Correia PN, 2021 | CHE | 183 | 3530 | 5.18 [ 4.50, 5.97] |
| Gardener, 2019 | USA | 4277 | 58435 | 7.32 [ 7.11, 7.53] |
| Barlas RS, 2018 | UK | 334 | 3575 | 9.34 [ 8.43, 10.34] |
| Dharan AS, 2024 | AUS | 604 | 5994 | 10.08 [ 9.34, 10.86] |
| Koton Silvia, 2017 | ISR | 396 | 3915 | 10.11 [ 9.21, 11.10] |
| Appelros, 2017 | SWE | 2564 | 23356 | 10.98 [10.58, 11.39] |
| Yasaka, 2018 | JPN | 1105 | 9804 | 11.27 [10.66, 11.91] |
| Wilbers, 2020 | NLD | 321 | 2736 | 11.73 [10.58, 12.99] |
| Phan HT, 2022 | USA | 165 | 1334 | 12.37 [10.71, 14.25] |
| Sundbøll, 2018 | DEN | 13764 | 98487 | 13.98 [13.76, 14.19] |
| Pana TA, 2019 | UK | 1683 | 10816 | 15.56 [14.89, 16.26] |
| Renoux C, 2017 | UK | 449 | 2553 | 17.59 [16.16, 19.11] |
| Akyea RK, 2022 | UK | 1018 | 5368 | 18.96 [17.94, 20.04] |
| Beuker C, 2023 | GER | 4007 | 18506 | 21.65 [21.06, 22.25] |
| Aparicio HJ, 2022 | USA | 13360 | 29690 | 45.00 [44.43, 45.56] |
| Median prevalence | | | | 6.67 [ 4.67, 9.44] |
| Population−averaged prevalence | Tau² = 1 I² = 100% | | | 9.35 [ 6.52, 13.14] |

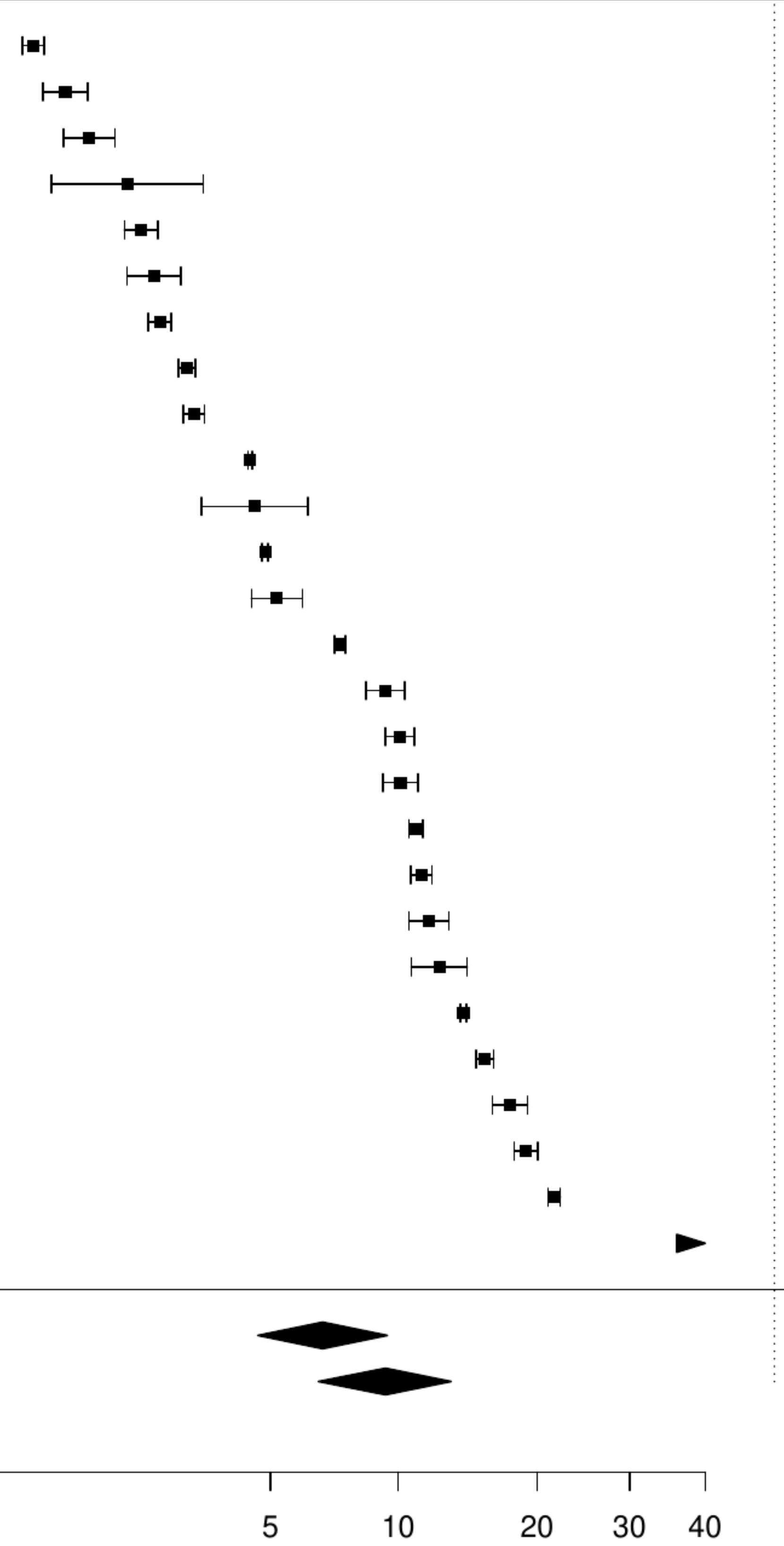

**Figure S65. Pooled prevalence of previous cancer in ischemic stroke.**

| Author(s) and Year | Country | Events | N | Prevalence [95% CI] |
|---|---|---|---|---|
| Casetta I, 2020 | ITA | 171 | 3045 | 5.62 [ 4.85, 6.49] |
| Kristiansen MH, 2023 | DEN | 64 | 538 | 11.90 [ 9.42, 14.91] |
| Ganesh A, 2017 | UK | 221 | 1425 | 15.51 [13.72, 17.48] |
| Bisson A, 2018 | FRA | 40456 | 240459 | 16.82 [16.68, 16.97] |
| Reznik ME, 2018 | USA | 189 | 1003 | 18.84 [16.54, 21.38] |
| Median prevalence | | | | 12.84 [ 8.74, 18.46] |
| Population−averaged prevalence | | | | 13.75 [ 9.34, 16.84] |

$Tau^2 = 0.23$   $I^2 = 98.5\%$

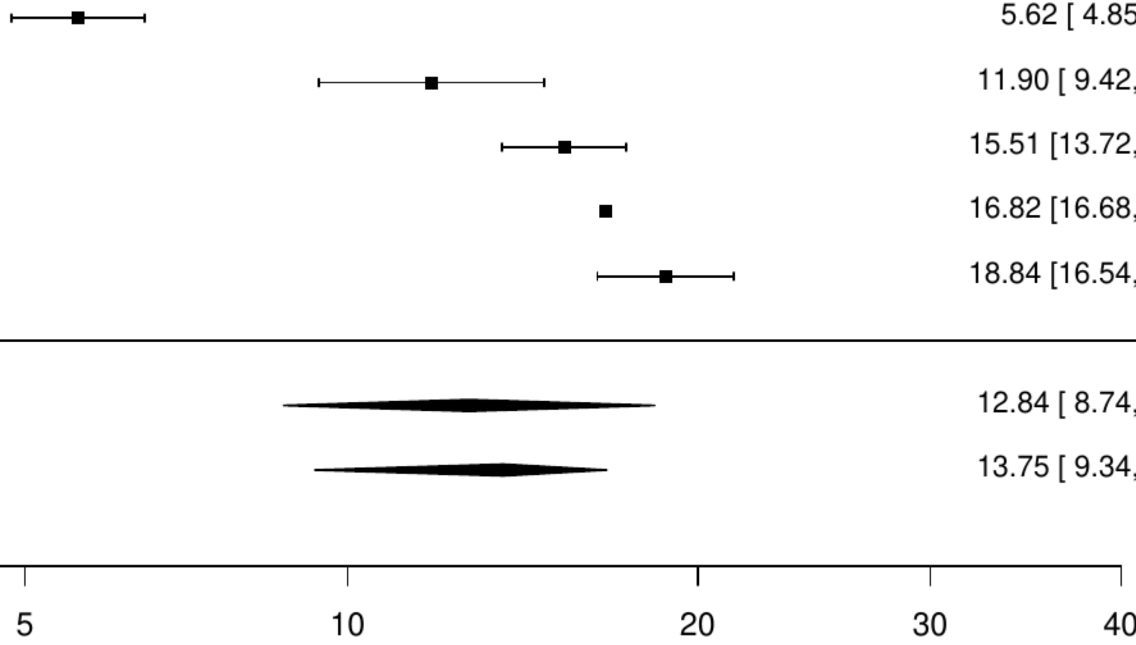

**Figure S66. Pooled prevalence of metastatic cancer in ischemic stroke.**

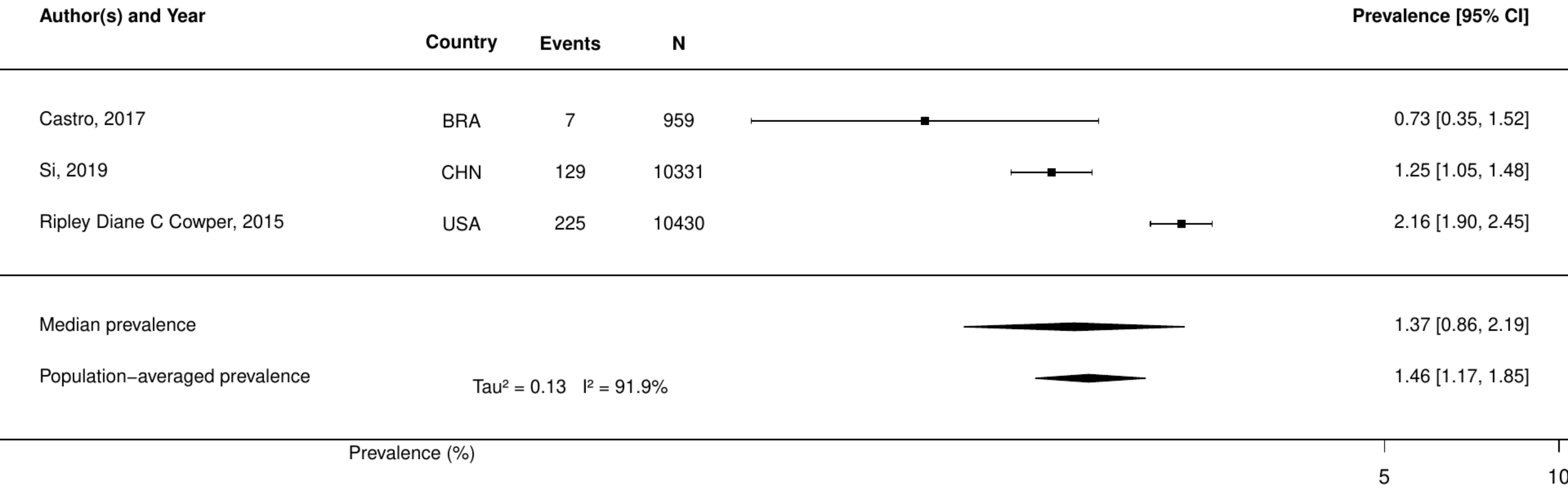

**Figure S67. Pooled prevalence of solid cancer in ischemic stroke.**

| Author(s) and Year | Country | Events | N | Prevalence [95% CI] |
|---|---|---|---|---|
| Castro, 2017 | BRA | 25 | 959 | 2.61 [ 1.77, 3.83] |
| Si, 2019 | CHN | 368 | 10331 | 3.56 [ 3.22, 3.94] |
| Ripley Diane C Cowper, 2015 | USA | 1760 | 10430 | 16.87 [16.17, 17.61] |
| Median prevalence | | | | 5.57 [ 2.09, 14.01] |
| Population−averaged prevalence | | | | 7.46 [ 3.41, 12.55] |

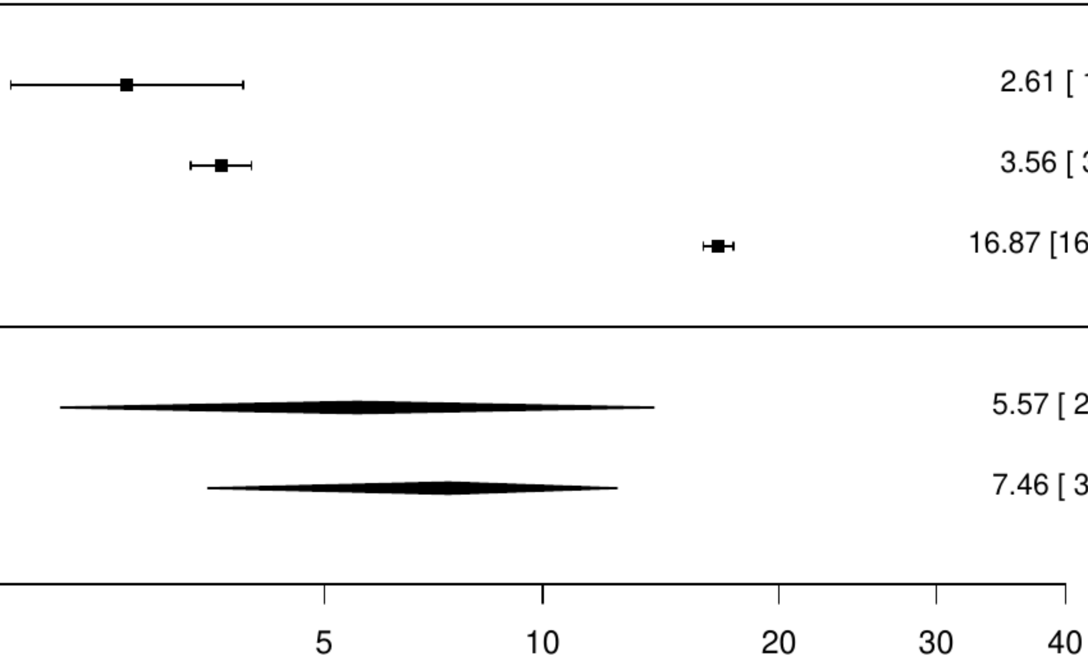

**Figure S68. Pooled prevalence of any cancer in hemorrhagic stroke.**

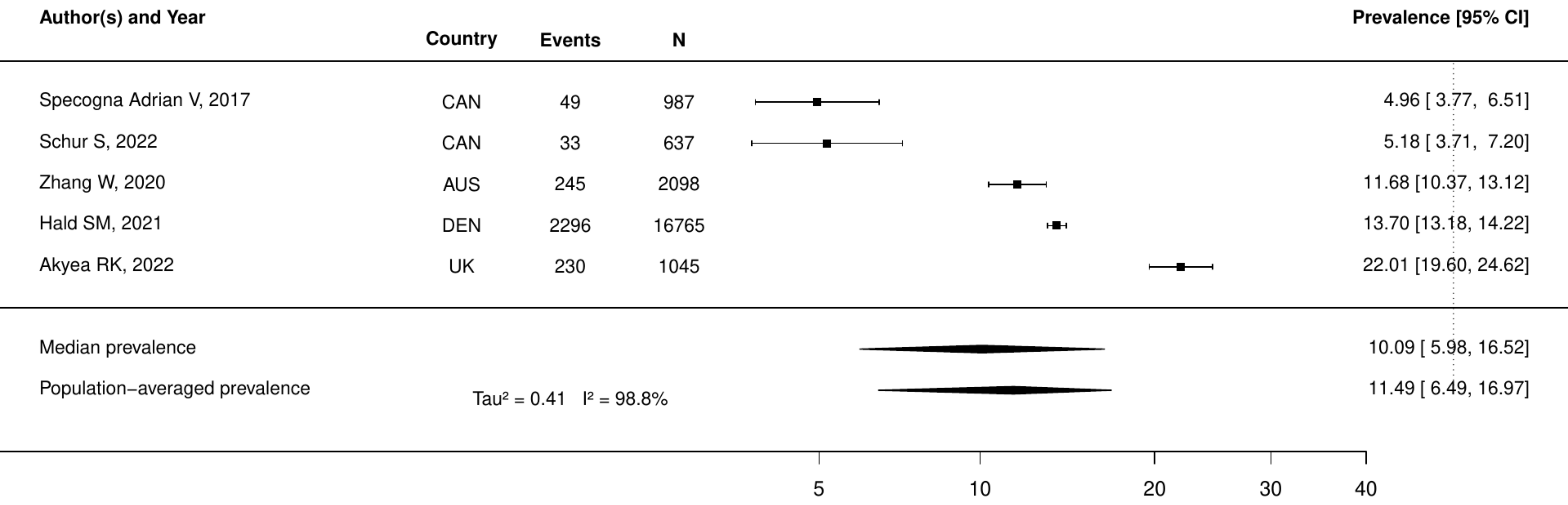

**Figure S69. Pooled prevalence of any cancer in peripheral artery disease.**

| Author(s) and Year | Country | Events | N | Prevalence [95% CI] |
|---|---|---|---|---|
| Lüders F, 2016 | GER | 774 | 41882 | 1.85 [ 1.72, 1.98] |
| Smilowitz, 2020 | USA | 774 | 39441 | 1.96 [ 1.83, 2.10] |
| Yuo, 2019 | USA | 1135 | 20347 | 5.58 [ 5.27, 5.90] |
| Keller K, 2019 | GER | 431808 | 5611827 | 7.69 [ 7.67, 7.72] |
| Perez, 2016 | ESP | 140 | 1663 | 8.42 [ 7.18, 9.85] |
| Lee HF, 2020 | TW | 9470 | 106327 | 8.91 [ 8.74, 9.08] |
| Marone, 2018 | ITA | 1834 | 18344 | 10.00 [ 9.57, 10.44] |
| Jelani QU, 2021 | Multinational | 121 | 1204 | 10.05 [ 8.47, 11.88] |
| Kamil S, 2021 | DEN | 32891 | 299293 | 10.99 [10.88, 11.10] |
| Tepe G, 2021 | Multinational | 100 | 877 | 11.40 [ 9.46, 13.68] |
| Liu FL, 2017 | TW | 951 | 6647 | 14.31 [13.49, 15.17] |
| Vedder IR, 2020 | NLD | 108 | 686 | 15.74 [13.21, 18.66] |
| Sigvant B, 2017 | SWE | 12480 | 66189 | 18.86 [18.56, 19.15] |
| Bourrier, 2020 | CAN | 4512 | 20600 | 21.90 [21.34, 22.47] |
| O'Brien EC, 2019 | USA | 2319 | 9743 | 23.80 [22.97, 24.66] |
| Arruda−Olson AM, 2018 | USA | 648 | 1676 | 38.66 [36.36, 41.02] |
| Median prevalence | | | | 10.50 [ 7.07, 15.32] |
| Population−averaged prevalence | Tau² = 0.78 I² = 100% | | | 13.20 [ 9.11, 17.61] |

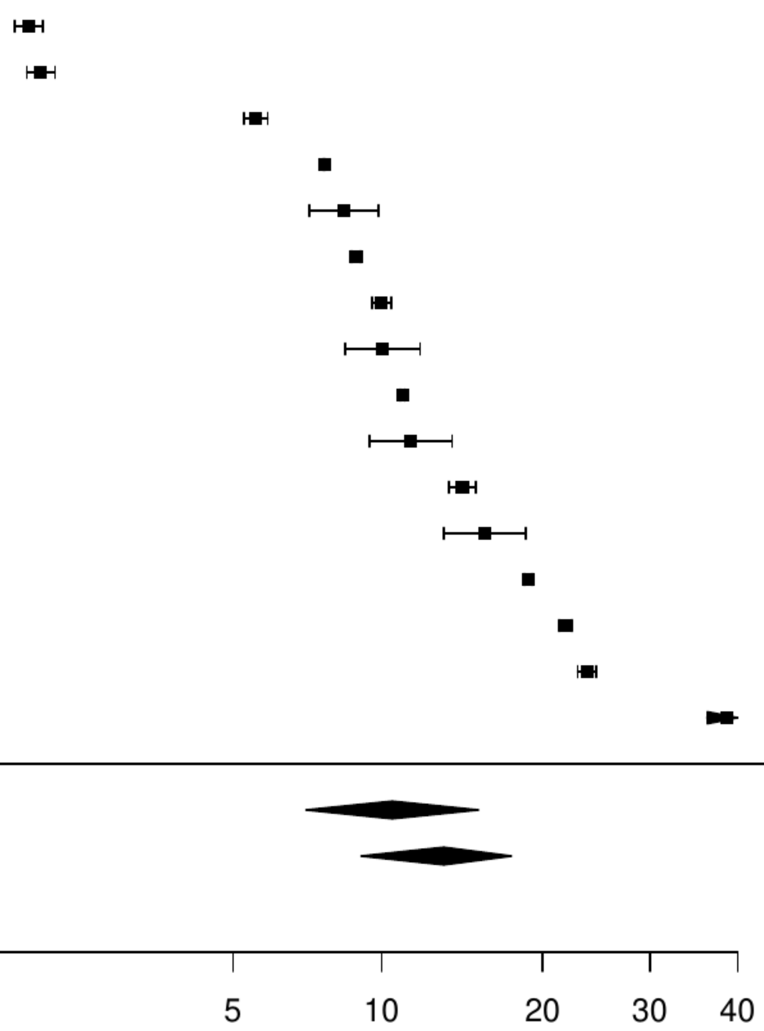

**Figure S70. Pooled prevalence of metastatic cancer in peripheral artery disease.**

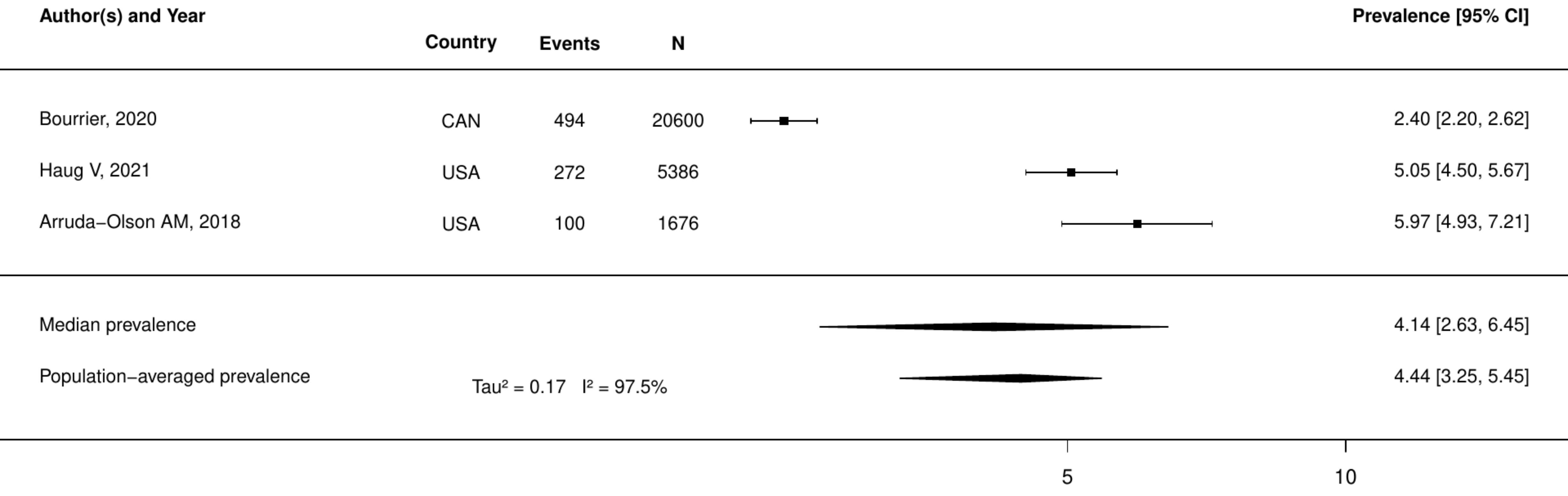

**Figure S71. Pooled prevalence of active cancer in valve heart disease.**

| Author(s) and Year | Country | Events | N | Prevalence [95% CI] |
|---|---|---|---|---|
| Du Y, 2021 | USA | 18 | 729 | 2.47 [1.56, 3.88] |
| Chieffo A, 2016 | Multinational | 36 | 1019 | 3.53 [2.56, 4.86] |
| Minamino−Muta, 2017 | JPN | 149 | 3815 | 3.91 [3.34, 4.57] |
| Zahn, 2017 | GER | 55 | 1378 | 3.99 [3.08, 5.16] |
| Ishizu, 2021 | JPN | 124 | 2588 | 4.79 [4.03, 5.68] |
| Mentias A, 2020 | USA | 6913 | 134717 | 5.13 [5.01, 5.25] |
| Pasceri, 2024 | Multinational | 411 | 6765 | 6.08 [5.53, 6.67] |
| Werner N, 2020 | GER | 73 | 786 | 9.29 [7.45, 11.53] |
| Median prevalence | | | | 4.67 [3.67, 5.93] |
| Population−averaged prevalence | | | | 4.90 [3.84, 6.37] |

$Tau^2 = 0.12$   $I^2 = 96.2\%$

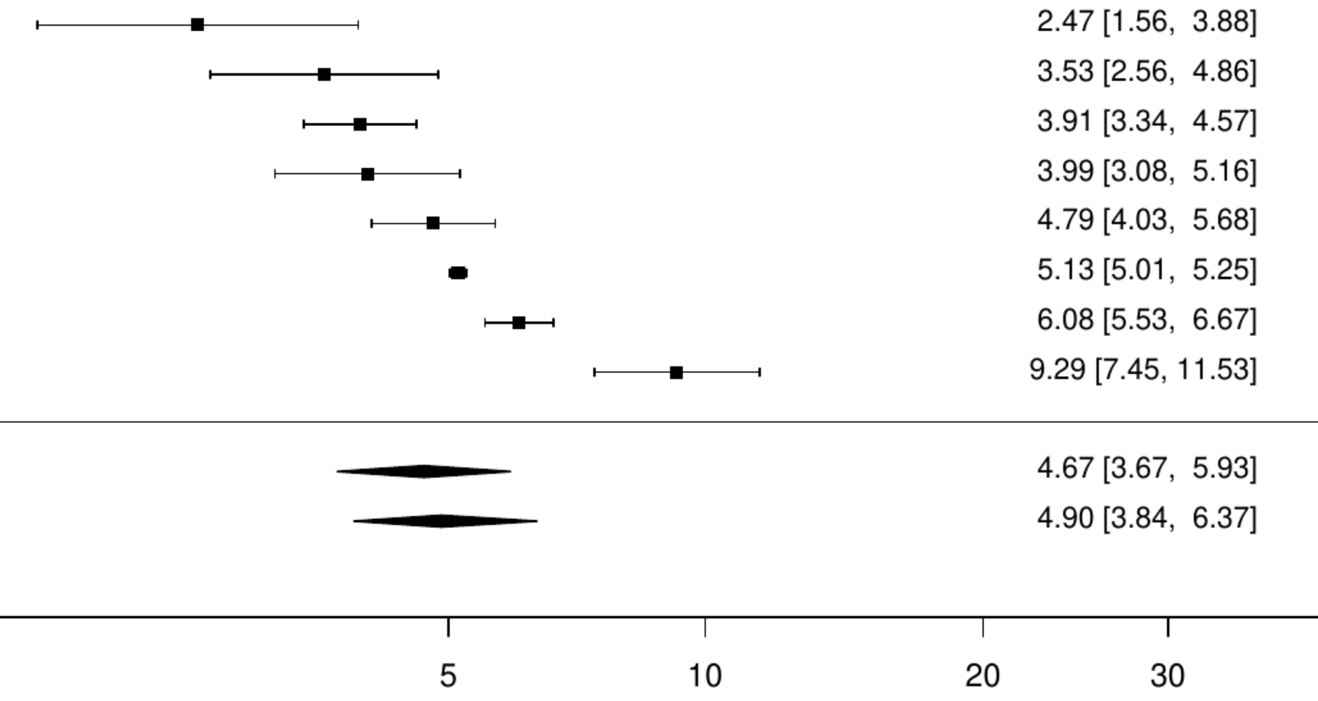

**Figure S72. Pooled prevalence of any cancer in valve heart disease.**

| Author(s) and Year | Country | Events | N | Prevalence [95% CI] |
|---|---|---|---|---|
| Jang SY, 2022 | KOR | 42 | 3773 | 1.11 [ 0.82, 1.50] |
| Tarantini G, 2020 | ITA | 82 | 4801 | 1.71 [ 1.38, 2.12] |
| Percy ED, 2021 | USA | 21 | 617 | 3.40 [ 2.23, 5.16] |
| Khera Sahil, 2017 | USA | 660 | 16252 | 4.06 [ 3.77, 4.38] |
| Shimura, 2017 | JPN | 61 | 1215 | 5.02 [ 3.93, 6.40] |
| Wijeysundera HC, 2020 | CAN | 285 | 4861 | 5.86 [ 5.24, 6.56] |
| Yang H, 2023 | USA | 51 | 785 | 6.50 [ 4.97, 8.45] |
| Zaheer A, 2024 | CAN | 753 | 11077 | 6.80 [ 6.34, 7.28] |
| Kawsara A, 2021 | USA | 132 | 1513 | 8.72 [ 7.40, 10.26] |
| Kalbacher D, 2019 | GER | 71 | 762 | 9.32 [ 7.45, 11.60] |
| Malmberg, 2021 | FIN | 1181 | 10635 | 11.10 [10.52, 11.72] |
| Biancari F, 2020 | FIN | 264 | 2216 | 11.91 [10.63, 13.33] |
| Czarnecki A, 2017 | CAN | 123 | 999 | 12.31 [10.42, 14.50] |
| Dawwas GK, 2021 | USA | 10399 | 81243 | 12.80 [12.57, 13.03] |
| Hiltner E, 2021 | USA | 1981 | 14996 | 13.21 [12.68, 13.76] |
| Fauchier L, 2020 | FRA | 14398 | 107806 | 13.36 [13.15, 13.56] |
| Minamino−Muta, 2017 | JPN | 517 | 3815 | 13.55 [12.50, 14.68] |
| Østergaard, 2019 | DEN | 11934 | 83453 | 14.30 [14.06, 14.54] |
| Young C, 2018 | Multinational | 79 | 516 | 15.31 [12.45, 18.68] |
| Moretti Claudio, 2015 | Multinational | 127 | 811 | 15.66 [13.32, 18.33] |
| Schymik G, 2015 | Multinational | 480 | 2688 | 17.86 [16.45, 19.35] |
| Durand E, 2017 | FRA | 98 | 546 | 17.95 [14.95, 21.40] |
| Median prevalence | | | | 8.46 [ 6.25, 11.34] |
| Population−averaged prevalence | Tau² = 0.6 I² = 99.9% | | | 10.32 [ 8.28, 12.35] |

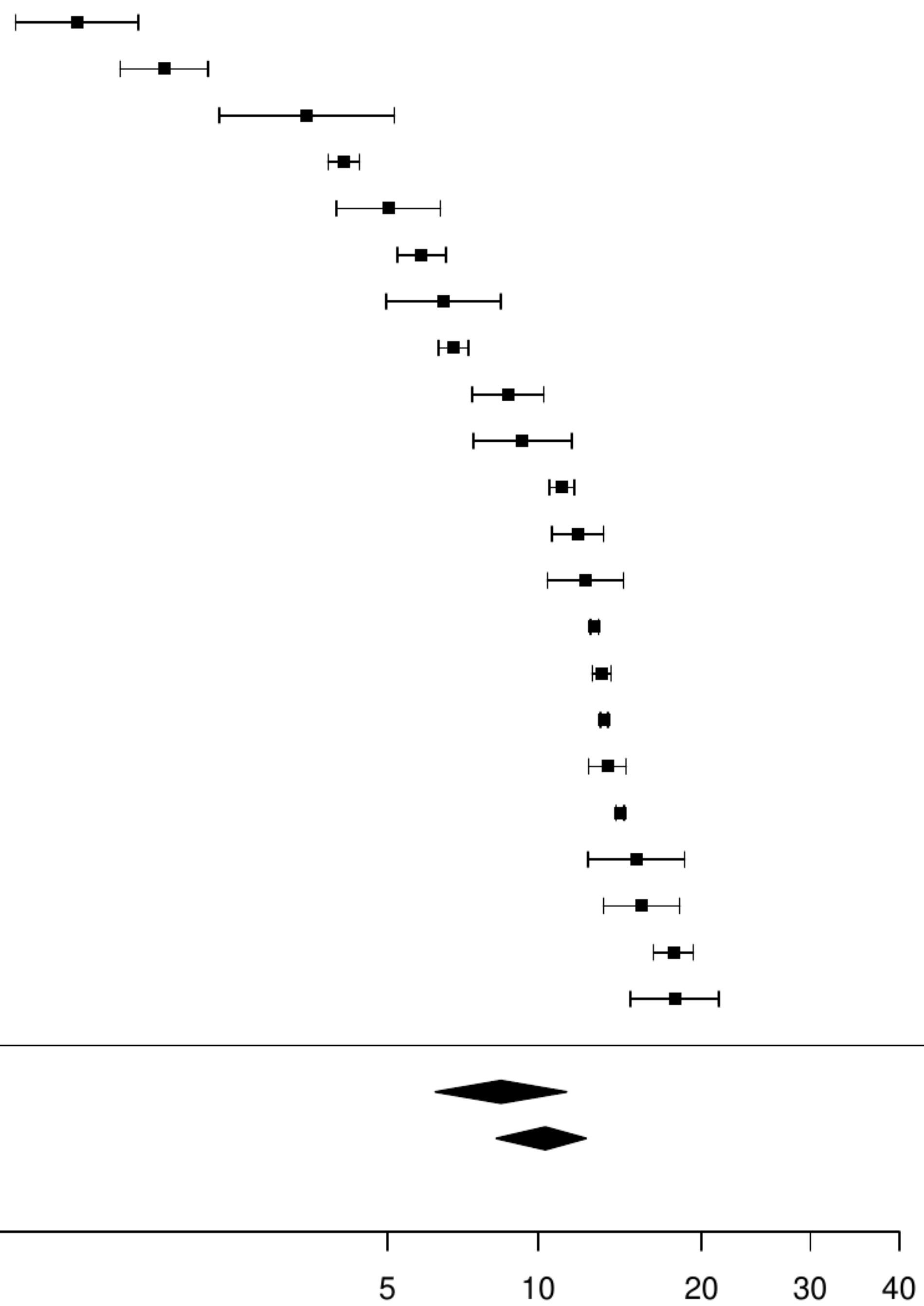

**Figure S73. Pooled prevalence of previous cancer in valve heart disease.**

| Author(s) and Year | Country | Events | N | Prevalence [95% CI] |
|---|---|---|---|---|
| Park SJ, 2023 | KOR | 1457 | 24374 | 5.98 [ 5.69, 6.28] |
| Finkelstein A, 2019 | ISR | 292 | 2336 | 12.50 [11.22, 13.90] |
| Biasco L, 2024 | CHE | 79 | 581 | 13.60 [11.04, 16.63] |
| Glaser N, 2024 | SWE | 3311 | 21002 | 15.77 [15.28, 16.26] |
| Melgaard L, 2020 | DEN | 4887 | 28770 | 16.99 [16.56, 17.42] |
| Lantelme P, 2020 | FRA | 4065 | 20443 | 19.88 [19.34, 20.44] |
| Kiramijyan S, 2016 | USA | 118 | 589 | 20.03 [17.00, 23.46] |
| Frank, 2019 | Multinational | 407 | 1946 | 20.91 [19.17, 22.78] |
| Median prevalence | | | | 14.90 [11.42, 19.21] |
| Population−averaged prevalence | | | | 15.69 [12.31, 18.65] |

Tau² = 0.19   I² = 99.6%

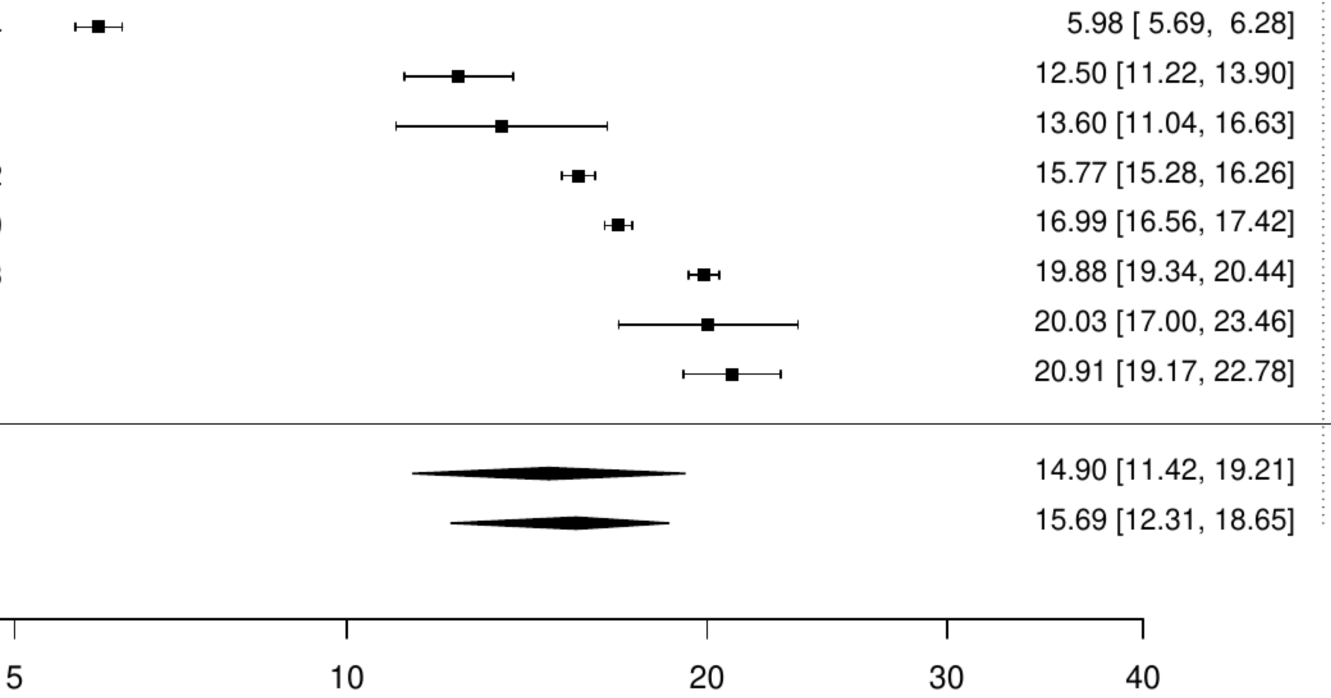

**Figure S74. Pooled prevalence of blood cancer in valve heart disease.**

| Author(s) and Year | Country | Events | N | Prevalence [95% CI] |
|---|---|---|---|---|
| Pack QR, 2016 | USA | 230 | 38532 | 0.60 [0.52, 0.68] |
| Mentias A, 2020 | USA | 2451 | 134717 | 1.82 [1.75, 1.89] |
| McCullough, 2019 | USA | 171 | 8757 | 1.95 [1.68, 2.26] |
| Median prevalence | | | | 1.29 [0.70, 2.37] |
| Population−averaged prevalence | Tau² = 0.3 I² = 99.1% |  |  | 1.48 [0.99, 1.83] |

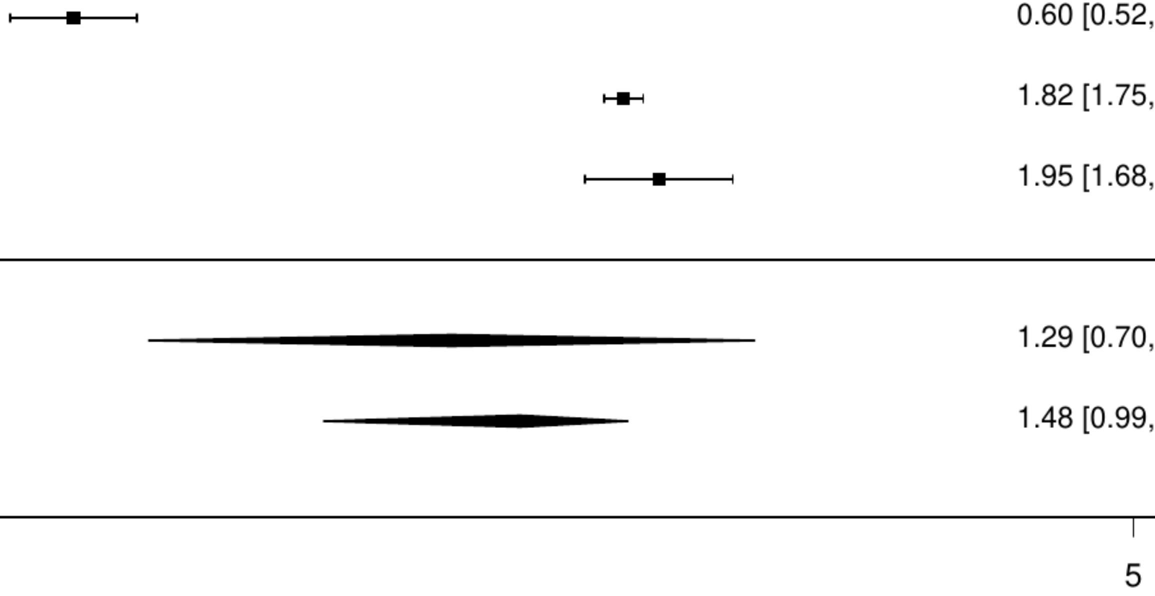


Prevalence (%)

**Figure S75. Pooled prevalence of metastatic cancer in valve heart disease.**

| Author(s) and Year | Country | Events | N | Prevalence [95% CI] |
|---|---|---|---|---|
| Pack QR, 2016 | USA | 57 | 38532 | 0.15 [0.11, 0.19] |
| Yang H, 2023 | USA | 7 | 785 | 0.89 [0.43, 1.86] |
| Mentias A, 2020 | USA | 1593 | 134717 | 1.18 [1.13, 1.24] |
| McCullough, 2019 | USA | 300 | 8757 | 3.43 [3.06, 3.83] |
| Median prevalence | | | | 0.85 [0.27, 2.62] |
| Population−averaged prevalence | Tau² = 1.33 I² = 99.6% |  |  | 1.56 [0.37, 2.94] |

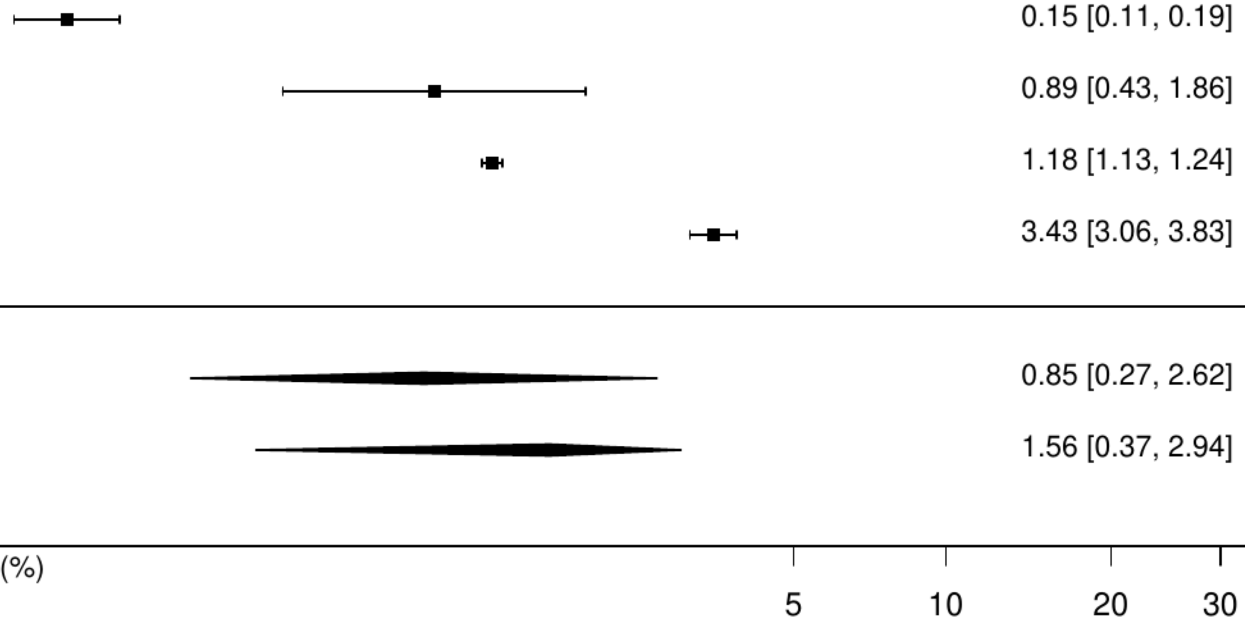

**Figure S76. Pooled prevalence of active cancer in aortic stenosis.**

| Author(s) and Year | Country | Events | N | Prevalence [95% CI] |
|---|---|---|---|---|
| Du Y, 2021 | USA | 18 | 729 | 2.47 [1.56, 3.88] |
| Chieffo A, 2016 | Multinational | 36 | 1019 | 3.53 [2.56, 4.86] |
| Minamino−Muta, 2017 | JPN | 149 | 3815 | 3.91 [3.34, 4.57] |
| Zahn, 2017 | GER | 55 | 1378 | 3.99 [3.08, 5.16] |
| Ishizu, 2021 | JPN | 124 | 2588 | 4.79 [4.03, 5.68] |
| Mentias A, 2020 | USA | 6913 | 134717 | 5.13 [5.01, 5.25] |
| Pasceri, 2024 | Multinational | 411 | 6765 | 6.08 [5.53, 6.67] |
| Median prevalence | | | | 4.33 [3.61, 5.20] |
| Population−averaged prevalence | | | | 4.42 [3.64, 5.21] |

$Tau^2 = 0.05$   $I^2 = 92\%$

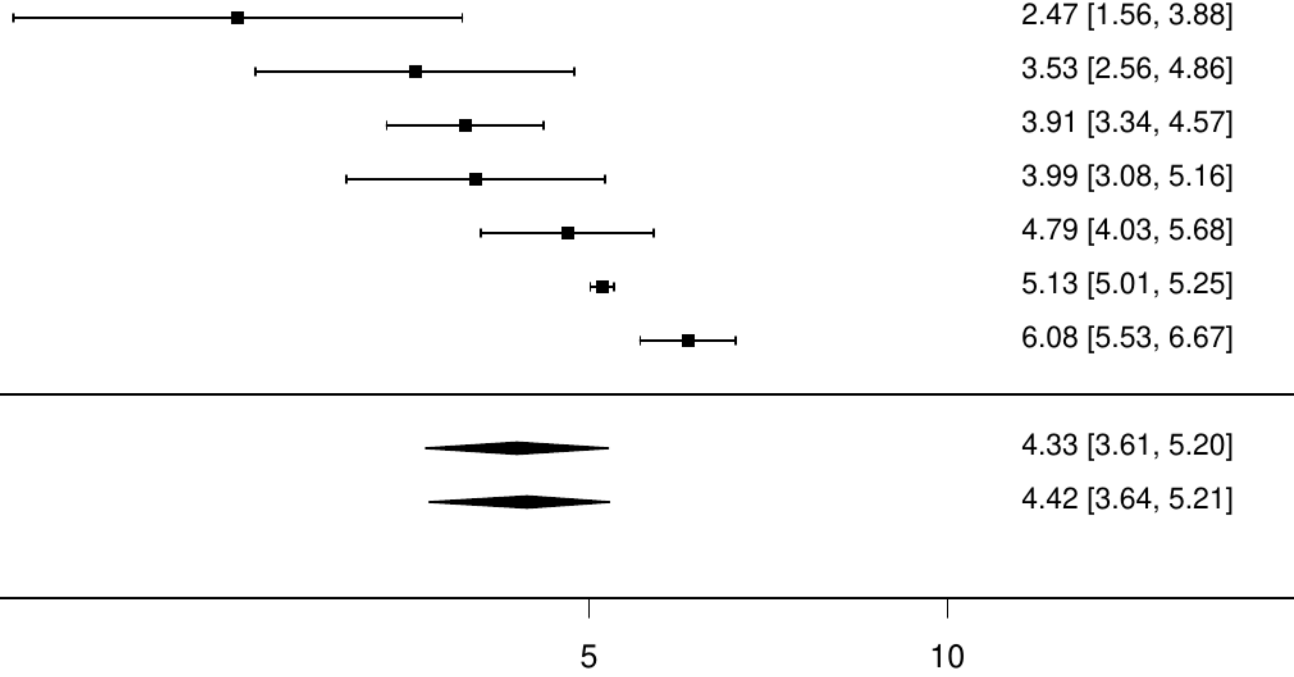

**Figure S77. Pooled prevalence of any cancer in aortic stenosis.**

| Author(s) and Year | Country | Events | N | Prevalence [95% CI] |
|---|---|---|---|---|
| Jang SY, 2022 | KOR | 42 | 3773 | 1.11 [ 0.82, 1.50] |
| Tarantini G, 2020 | ITA | 82 | 4801 | 1.71 [ 1.38, 2.12] |
| Percy ED, 2021 | USA | 21 | 617 | 3.40 [ 2.23, 5.16] |
| Khera Sahil, 2017 | USA | 660 | 16252 | 4.06 [ 3.77, 4.38] |
| Shimura, 2017 | JPN | 61 | 1215 | 5.02 [ 3.93, 6.40] |
| Wijeysundera HC, 2020 | CAN | 285 | 4861 | 5.86 [ 5.24, 6.56] |
| Yang H, 2023 | USA | 51 | 785 | 6.50 [ 4.97, 8.45] |
| Zaheer A, 2024 | CAN | 753 | 11077 | 6.80 [ 6.34, 7.28] |
| Biancari F, 2020 | FIN | 264 | 2216 | 11.91 [10.63, 13.33] |
| Matthew Brennan J, 2020 | USA | 4206 | 32853 | 12.80 [12.45, 13.17] |
| Hiltner E, 2021 | USA | 1981 | 14996 | 13.21 [12.68, 13.76] |
| Fauchier L, 2020 | FRA | 14398 | 107806 | 13.36 [13.15, 13.56] |
| Minamino−Muta, 2017 | JPN | 517 | 3815 | 13.55 [12.50, 14.68] |
| Young C, 2018 | Multinational | 79 | 516 | 15.31 [12.45, 18.68] |
| Moretti Claudio, 2015 | Multinational | 127 | 811 | 15.66 [13.32, 18.33] |
| Schymik G, 2015 | Multinational | 480 | 2688 | 17.86 [16.45, 19.35] |
| Durand E, 2017 | FRA | 98 | 546 | 17.95 [14.95, 21.40] |
| Butt, 2019 | DEN | 1237 | 6409 | 19.30 [18.35, 20.29] |
| Median prevalence | | | | 8.24 [ 5.66, 11.84] |
| Population−averaged prevalence | | | | 10.53 [ 7.70, 13.36] |

Tau² = 0.75 I² = 99.8%

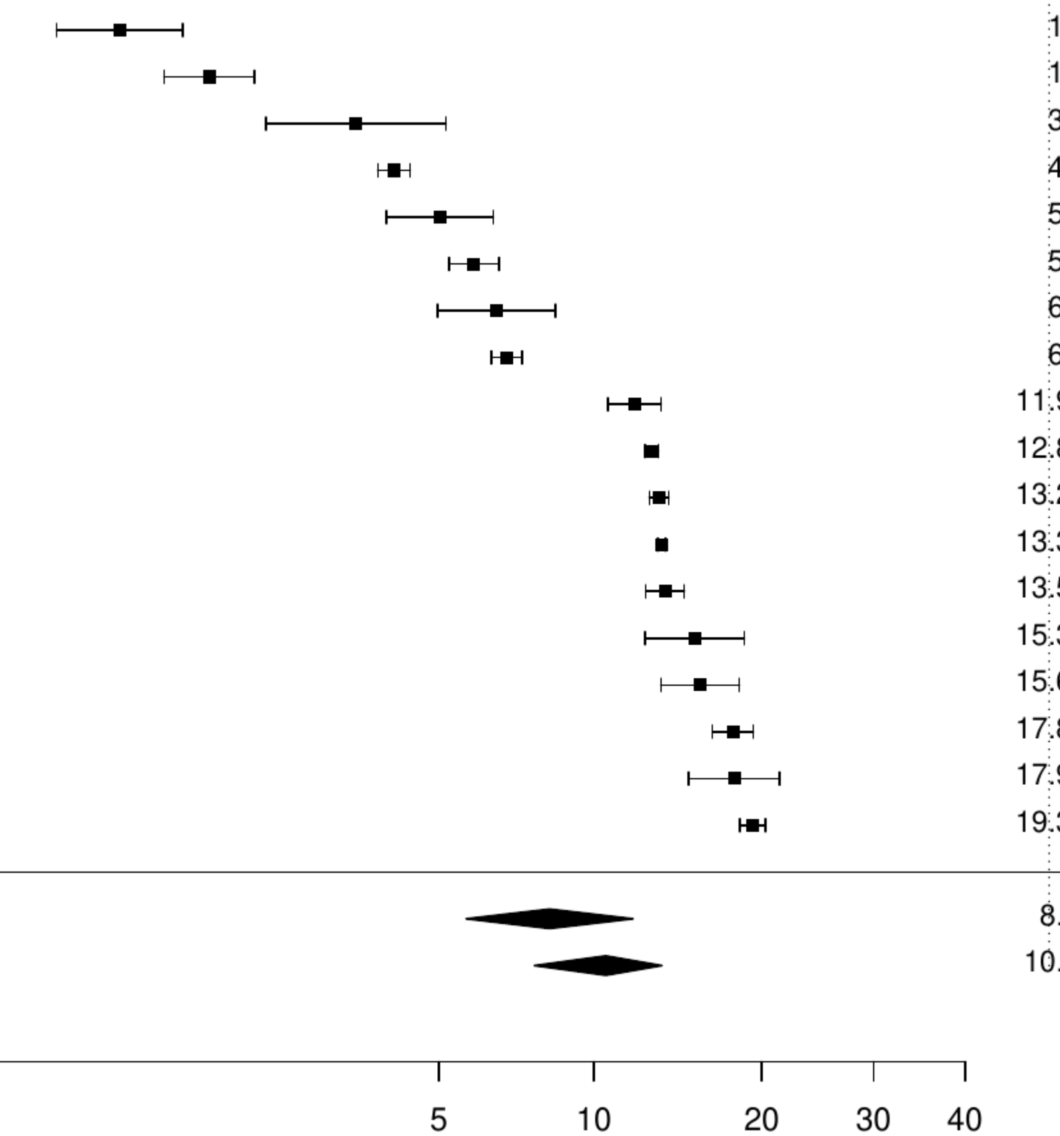

**Figure S78. Pooled prevalence of previous cancer in aortic stenosis.**

| Author(s) and Year | Country | Events | N | Prevalence [95% CI] |
|---|---|---|---|---|
| Finkelstein A, 2019 | ISR | 292 | 2336 | 12.50 [11.22, 13.90] |
| Glaser N, 2024 | SWE | 3311 | 21002 | 15.77 [15.28, 16.26] |
| Clementy N, 2021 | FRA | 5859 | 29422 | 19.91 [19.46, 20.37] |
| Kiramijyan S, 2016 | USA | 118 | 589 | 20.03 [17.00, 23.46] |
| Frank, 2019 | Multinational | 407 | 1946 | 20.91 [19.17, 22.78] |
| Median prevalence | | | | 17.52 [14.77, 20.67] |
| Population−averaged prevalence | | | | 17.74 [14.60, 20.03] |

Tau² = 0.05   I² = 98.3%

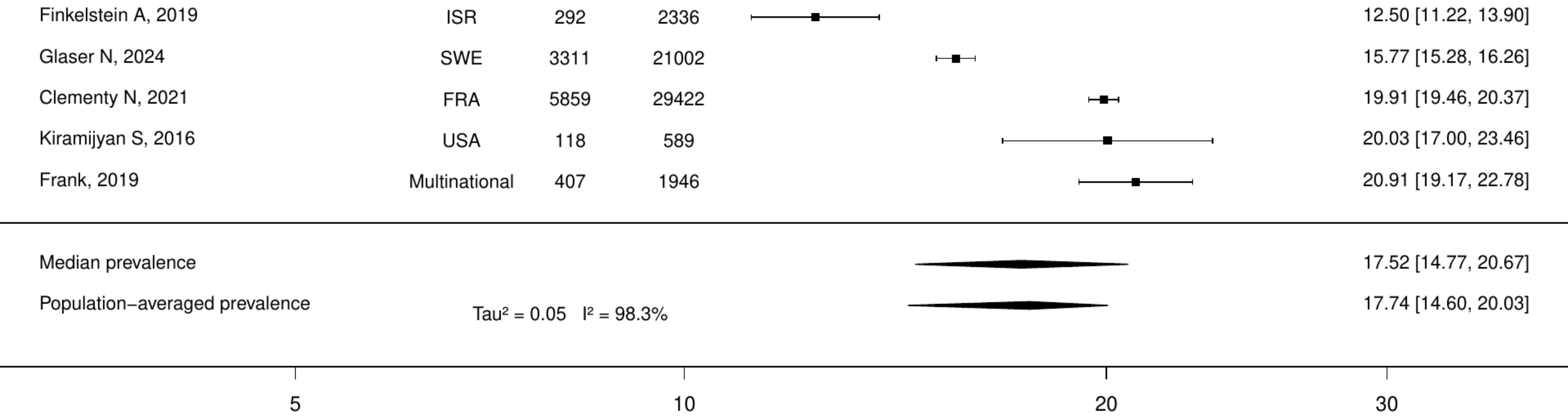

**Figure S79. Pooled prevalence of active cancer in transcatheter aortic valve intervention.**

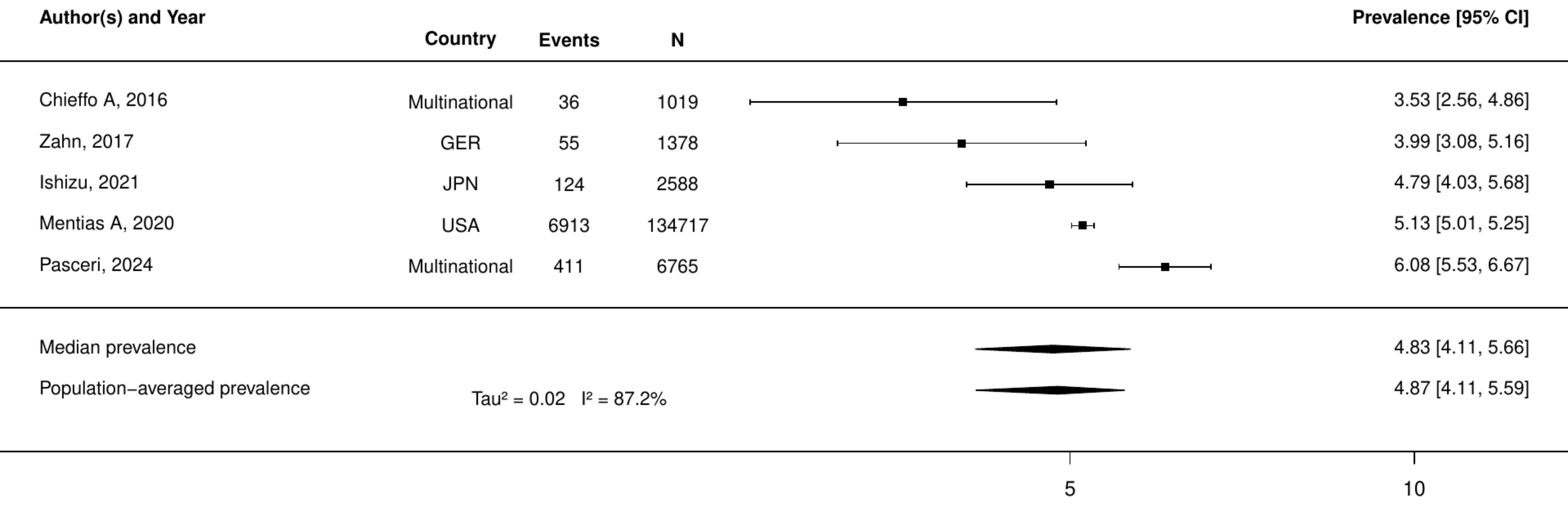

**Figure S80. Pooled prevalence of any cancer in transcatheter aortic valve intervention.**

| Author(s) and Year | Country | Events | N | Prevalence [95% CI] |
|---|---|---|---|---|
| Percy ED, 2021 | USA | 21 | 617 | 3.40 [ 2.23, 5.16] |
| Khera Sahil, 2017 | USA | 660 | 16252 | 4.06 [ 3.77, 4.38] |
| Shimura, 2017 | JPN | 61 | 1215 | 5.02 [ 3.93, 6.40] |
| Wijeysundera HC, 2020 | CAN | 285 | 4861 | 5.86 [ 5.24, 6.56] |
| Yang H, 2023 | USA | 51 | 785 | 6.50 [ 4.97, 8.45] |
| Zaheer A, 2024 | CAN | 753 | 11077 | 6.80 [ 6.34, 7.28] |
| Czarnecki A, 2017 | CAN | 123 | 999 | 12.31 [10.42, 14.50] |
| Young C, 2018 | Multinational | 79 | 516 | 15.31 [12.45, 18.68] |
| Schymik G, 2015 | Multinational | 480 | 2688 | 17.86 [16.45, 19.35] |
| Durand E, 2017 | FRA | 98 | 546 | 17.95 [14.95, 21.40] |
| De Backer Ole, 2020 | DEN | 564 | 2455 | 22.97 [21.35, 24.68] |
| Median prevalence | | | | 9.03 [ 6.12, 13.13] |
| Population−averaged prevalence | | | | 10.63 [ 7.12, 15.01] |

$Tau^2 = 0.49$   $I^2 = 99.1\%$

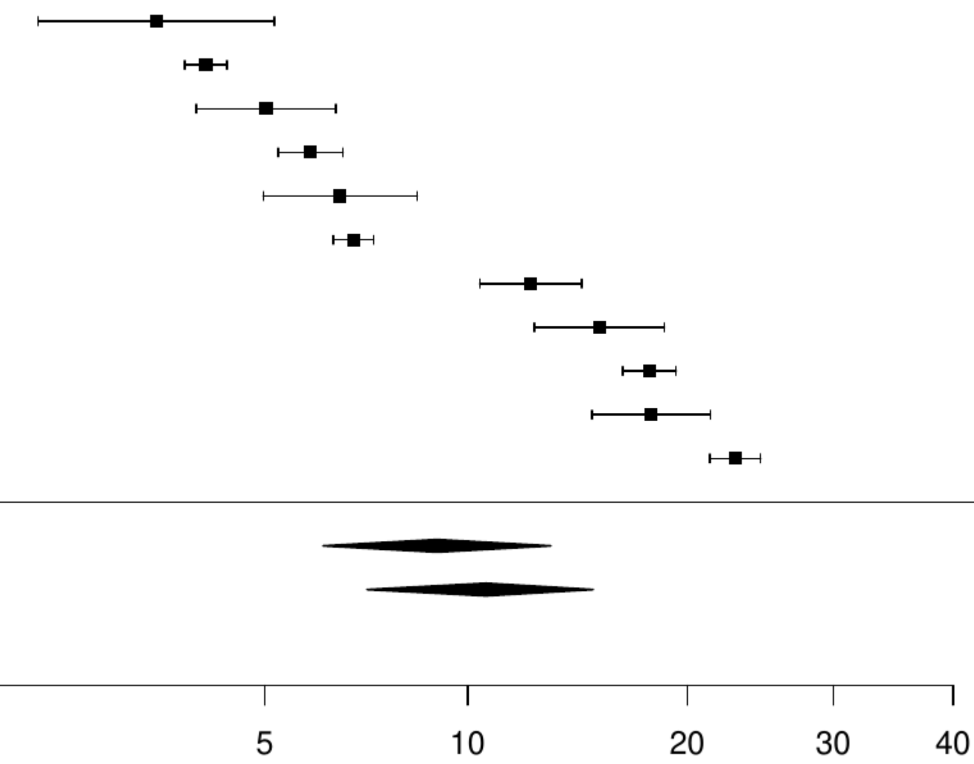

**Figure S81. Pooled prevalence of previous cancer in transcatheter aortic valve intervention.**

| Author(s) and Year | Country | Events | N | Prevalence [95% CI] |
|---|---|---|---|---|
| Finkelstein A, 2019 | ISR | 292 | 2336 | 12.50 [11.22, 13.90] |
| Clementy N, 2021 | FRA | 5859 | 29422 | 19.91 [19.46, 20.37] |
| Kiramijyan S, 2016 | USA | 118 | 589 | 20.03 [17.00, 23.46] |
| Frank, 2019 | Multinational | 407 | 1946 | 20.91 [19.17, 22.78] |
| Median prevalence | | | | 18.02 [14.65, 21.97] |
| Population−averaged prevalence | | | | 18.28 [14.40, 20.71] |

Tau² = 0.06   I² = 96.1%

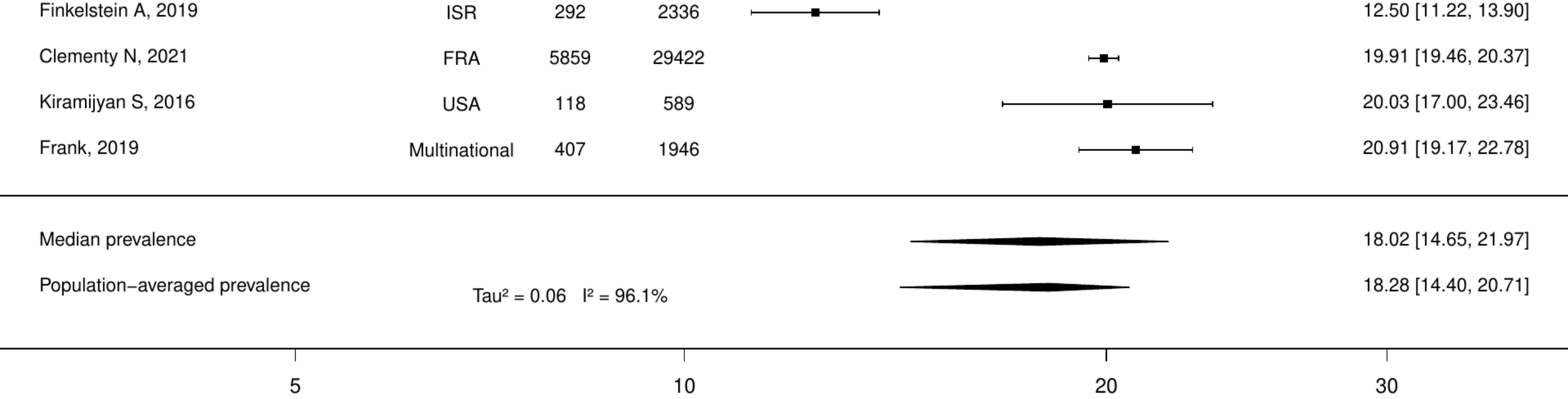

**Figure S82. Pooled prevalence of active cancer in type 2 diabetes mellitus.**

| Author(s) and Year | Country | Events | N | Prevalence [95% CI] |
|---|---|---|---|---|
| Cheung KK, 2017 | CHN | 8 | 1239 | 0.65 [0.32, 1.29] |
| Coleman CI, 2021 | USA | 6170 | 116049 | 5.32 [5.19, 5.45] |
| Goderis Geert, 2017 | BEL | 33 | 578 | 5.71 [4.09, 7.92] |
| Median prevalence | | | | 2.80 [0.90, 8.36] |
| Population−averaged prevalence | | | | 4.22 [2.18, 5.32] |

$Tau^2 = 0.99$   $I^2 = 97.5\%$

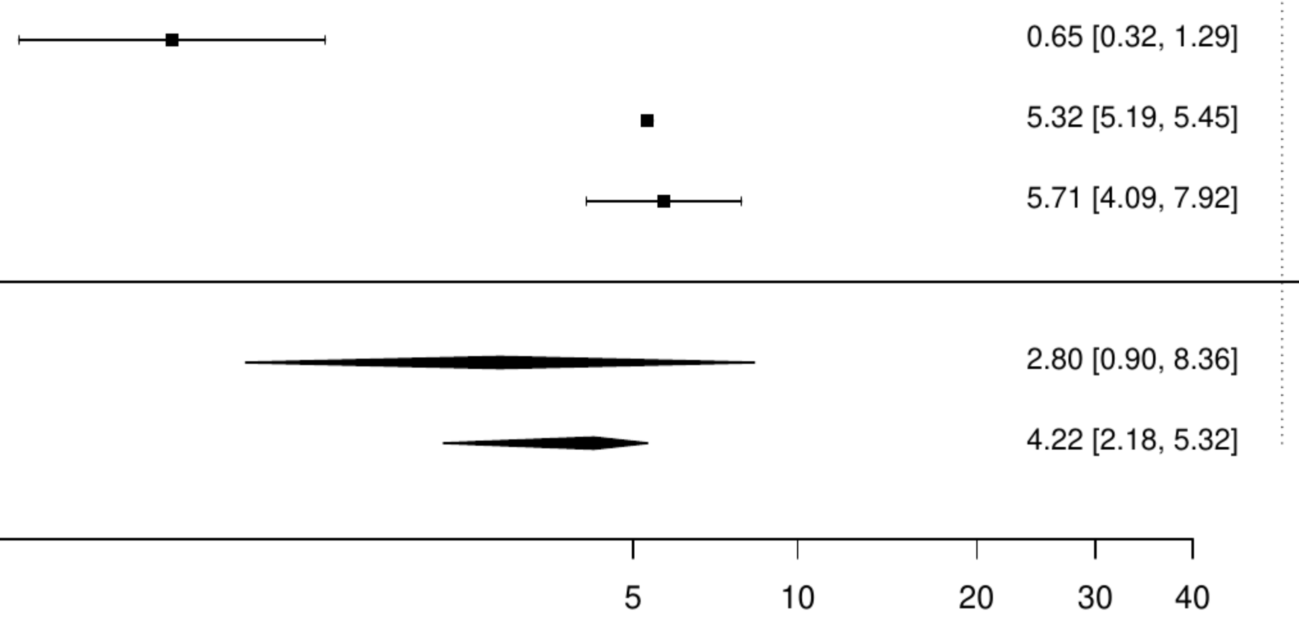

**Figure S83. Pooled prevalence of any cancer in type 2 diabetes mellitus.**

| Author(s) and Year | Country | Events | N | Prevalence [95% CI] |
|---|---|---|---|---|
| Igland, 2024 | NOR | 85 | 42290 | 0.20 [ 0.16, 0.25] |
| Roussel R, 2015 | FRA | 412 | 31652 | 1.30 [ 1.18, 1.43] |
| Pati, 2020 | IND | 12 | 912 | 1.32 [ 0.75, 2.30] |
| Usó−Talamantes, 2021 | ESP | 200 | 14935 | 1.34 [ 1.17, 1.54] |
| Ke C, 2020 | Hong Kong | 5361 | 349215 | 1.54 [ 1.49, 1.58] |
| He S, 2021 | CHN | 398 | 24099 | 1.65 [ 1.50, 1.82] |
| Kameda T, 2020 | JPN | 220 | 11595 | 1.90 [ 1.66, 2.16] |
| Xu Y, 2020 | CHN | 369 | 15752 | 2.34 [ 2.12, 2.59] |
| Chung GE, 2024 | KOR | 54931 | 2007775 | 2.74 [ 2.71, 2.76] |
| Lee S, 2023 | Hong Kong | 1676 | 61223 | 2.74 [ 2.61, 2.87] |
| Zareini B, 2020 | DEN | 5909 | 153405 | 3.85 [ 3.76, 3.95] |
| Wan EYF, 2023 | Hong Kong | 10297 | 263000 | 3.92 [ 3.84, 3.99] |
| Gu Y, 2016 | CHN | 516 | 12792 | 4.03 [ 3.71, 4.39] |
| Seong JM, 2019 | KOR | 55255 | 1140784 | 4.84 [ 4.80, 4.88] |
| Messina R, 2022 | ITA | 1536 | 30815 | 4.98 [ 4.75, 5.23] |
| van Mark G, 2021 | GER | 512 | 9601 | 5.33 [ 4.90, 5.80] |
| Franchi M, 2024 | ITA | 1014 | 18702 | 5.42 [ 5.11, 5.76] |
| Paul SK, 2021 | USA | 188810 | 3293983 | 5.73 [ 5.71, 5.76] |
| Eby EL, 2015 | USA | 6329 | 101728 | 6.22 [ 6.07, 6.37] |
| Koye DN, 2020 | UK | 23516 | 370854 | 6.34 [ 6.26, 6.42] |
| Sancho−Mestre, 2016 | ESP | 22792 | 350015 | 6.51 [ 6.43, 6.59] |
| McCoy RG, 2015 | USA | 2073 | 31545 | 6.57 [ 6.30, 6.85] |
| Satman Ilhan, 2021 | TUR | 1409 | 21180 | 6.65 [ 6.32, 7.00] |
| Luijks H, 2015 | NLD | 42 | 610 | 6.89 [ 5.13, 9.19] |
| Calderón−Larrañaga A, 2015 | ESP | 4821 | 68968 | 6.99 [ 6.80, 7.18] |
| Kai H, 2020 | JPN | 195 | 2718 | 7.17 [ 6.26, 8.21] |
| Blecker S, 2016 | USA | 339 | 4723 | 7.18 [ 6.48, 7.95] |
| Toh, 2018 | USA | 87587 | 1164307 | 7.52 [ 7.47, 7.57] |
| Yang CT, 2020 | TW | 1341651 | 17809183 | 7.53 [ 7.52, 7.55] |
| Bazo−Alvarez JC, 2021 | UK | 1816 | 23837 | 7.62 [ 7.29, 7.96] |
| Sukkar, 2020 | AUS | 745 | 9313 | 8.00 [ 7.47, 8.57] |
| Chen J, 2021 | USA | 52 | 647 | 8.04 [ 6.18, 10.40] |
| Raebel MA, 2017 | USA | 10555 | 129040 | 8.18 [ 8.03, 8.33] |
| Ojeda−Fernandez L, 2022 | ITA | 1975 | 23081 | 8.56 [ 8.20, 8.92] |
| Seyed Ahmadi S, 2020 | SWE | 37135 | 421855 | 8.80 [ 8.72, 8.89] |
| Lee HF, 2020 | TW | 9470 | 106327 | 8.91 [ 8.74, 9.08] |
| Ajmera M, 2015 | USA | 1493 | 16653 | 8.97 [ 8.54, 9.41] |
| Mizani MA, 2024 | UK | 37882 | 420448 | 9.01 [ 8.92, 9.10] |
| Nefs G, 2016 | NLD | 133 | 1465 | 9.08 [ 7.71, 10.66] |
| Ling AW, 2020 | TW | 2547 | 27989 | 9.10 [ 8.77, 9.44] |
| Mikkola I, 2022 | FIN | 477 | 5104 | 9.35 [ 8.58, 10.18] |
| McCoy, 2020 | USA | 18940 | 201705 | 9.39 [ 9.26, 9.52] |
| Sidorenkov G, 2018 | NLD | 143 | 1459 | 9.80 [ 8.38, 11.44] |
| Marone, 2018 | ITA | 1834 | 18344 | 10.00 [ 9.57, 10.44] |
| Hougen I, 2021 | CAN | 3311 | 32576 | 10.16 [ 9.84, 10.50] |
| Hua R, 2024 | UK | 2771 | 26394 | 10.50 [10.13, 10.87] |
| Tancredi M, 2015 | SWE | 45907 | 435369 | 10.54 [10.45, 10.64] |
| Longato E, 2020 | ITA | 1382 | 12996 | 10.63 [10.12, 11.18] |
| Chen, 2020 | USA | 13502 | 123905 | 10.90 [10.72, 11.07] |
| Goldshtein I, 2016 | ISR | 120 | 1069 | 11.23 [ 9.47, 13.26] |
| Jeong HE, 2023 | KOR | 169252 | 1470286 | 11.51 [11.46, 11.56] |
| Shao, 2023 | USA | 2342 | 19025 | 12.31 [11.85, 12.78] |
| Udell JA, 2018 | USA | 14052 | 111576 | 12.59 [12.40, 12.79] |
| Sicras−Mainar A, 2016 | ESP | 164 | 1301 | 12.61 [10.91, 14.52] |
| Creuzot Garcher CP, 2024 | FRA | 1533 | 11901 | 12.88 [12.29, 13.50] |
| Norhammar, 2019 | SWE | 37729 | 287180 | 13.14 [13.01, 13.26] |
| Real J, 2021 | ITA | 32910 | 239733 | 13.73 [13.59, 13.87] |
| Wong, 2024 | AUS | 376 | 2688 | 13.99 [12.73, 15.35] |
| Williams, 2020 | USA | 7824 | 54452 | 14.37 [14.08, 14.67] |
| Beltran−Valls MR, 2023 | USA | 6208 | 41726 | 14.88 [14.54, 15.22] |
| Jimenez−Garcia R, 2020 | ESP | 1418 | 7799 | 18.18 [17.34, 19.05] |
| Blin P, 2021 | FRA | 47066 | 258260 | 18.22 [18.08, 18.37] |
| Chiang, 2020 | AUS | 12733 | 69718 | 18.26 [17.98, 18.55] |
| Xu Y, 2020 | USA | 4671 | 22857 | 20.44 [19.92, 20.96] |
| Tang O, 2020 | USA | 437 | 1853 | 23.58 [21.71, 25.57] |
| Kohsaka S, 2021 | JPN | 6377 | 23340 | 27.32 [26.75, 27.90] |
| Rose, 2019 | USA | 69628 | 240074 | 29.00 [28.82, 29.18] |
| Median prevalence | Tau² = 0.78 I² = 100% | | | 7.29 [ 5.98, 8.86] |
| Population−averaged prevalence | | | | 9.52 [ 8.25, 11.02] |

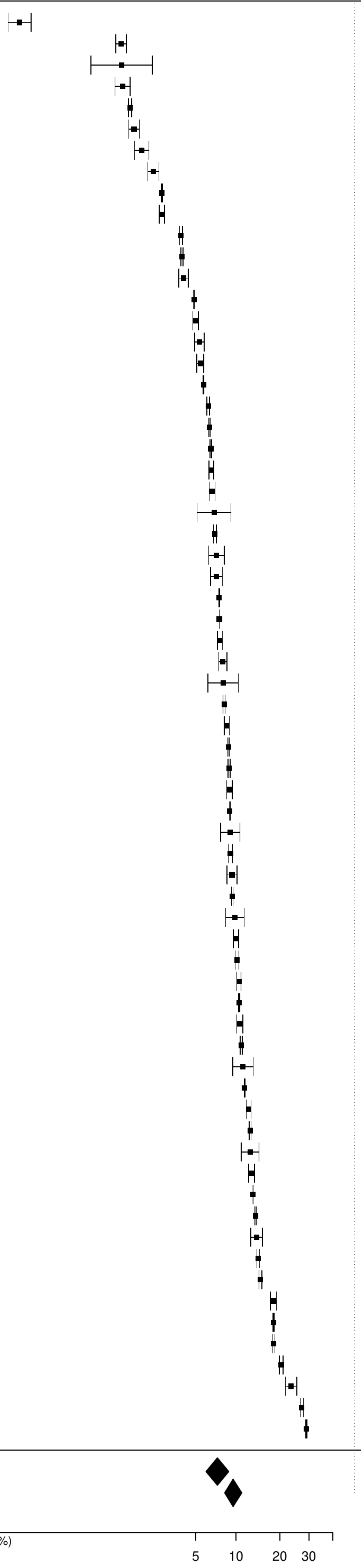

**Figure S84. Pooled prevalence of previous cancer in type 2 diabetes mellitus.**

| Author(s) and Year | Country | Events | N | Prevalence [95% CI] |
|---|---|---|---|---|
| Lavielle P, 2015 | MEX | 12 | 1712 | 0.70 [ 0.40, 1.23] |
| Takao T, 2021 | JPN | 36 | 2948 | 1.22 [ 0.88, 1.69] |
| Tanaka S, 2016 | JPN | 20 | 1532 | 1.31 [ 0.84, 2.01] |
| Captieux M, 2021 | UK | 6174 | 162316 | 3.80 [ 3.71, 3.90] |
| Fujita Y, 2022 | JPN | 370 | 7785 | 4.75 [ 4.30, 5.25] |
| Sattar Naveed, 2019 | SWE | 14988 | 241278 | 6.21 [ 6.12, 6.31] |
| Yokomichi H, 2017 | JPN | 2859 | 30834 | 9.27 [ 8.95, 9.60] |
| Komorita Y, 2020 | JPN | 462 | 4923 | 9.38 [ 8.60, 10.23] |
| Rodionov RN, 2021 | GER | 12790 | 101162 | 12.64 [12.44, 12.85] |
| Al−Salameh, 2020 | FRA | 134 | 983 | 13.63 [11.63, 15.92] |
| Garry Elizabeth M, 2018 | USA | 57181 | 247356 | 23.12 [22.95, 23.28] |
| Hsieh CF, 2015 | TW | 6821 | 9805 | 69.57 [68.65, 70.47] |
| Median prevalence | | | | 6.88 [ 3.05, 14.78] |
| Population−averaged prevalence | | | | 12.72 [ 4.87, 23.94] |

Tau² = 2.26 I² = 100%

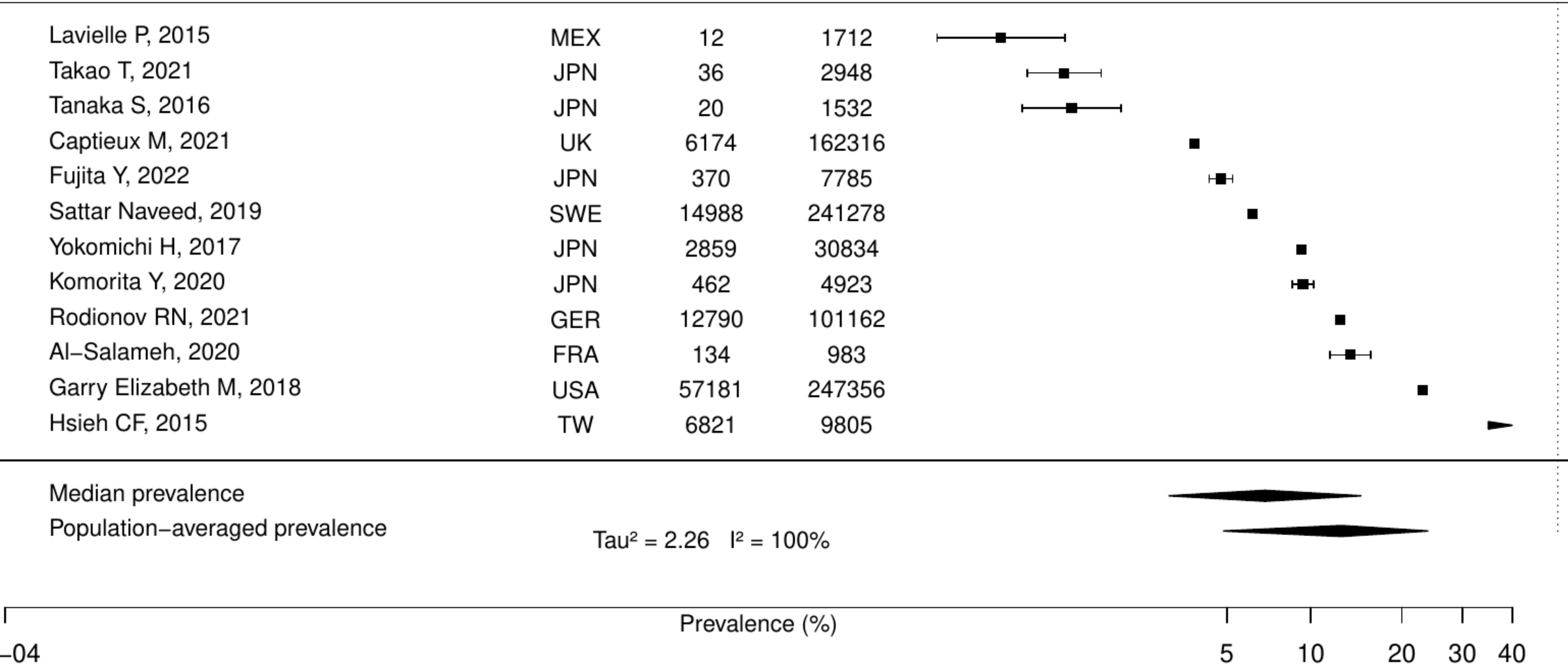

**Figure S85. Pooled prevalence of metastatic cancer in type 2 diabetes mellitus.**

| Author(s) and Year | Country | Events | N | Prevalence [95% CI] |
|---|---|---|---|---|
| Mehta HB, 2017 | UK | 106 | 53055 | 0.20 [0.17, 0.24] |
| Johnston, 2019 | USA | 78 | 13026 | 0.60 [0.48, 0.75] |
| Captieux M, 2021 | UK | 1093 | 162316 | 0.67 [0.63, 0.71] |
| Gomes C, 2020 | PRT | 83 | 6596 | 1.26 [1.02, 1.56] |
| Oh TK, 2020 | KOR | 874 | 63739 | 1.37 [1.28, 1.46] |
| Udell JA, 2018 | USA | 1843 | 111576 | 1.65 [1.58, 1.73] |
| Median prevalence | | | | 0.78 [0.44, 1.38] |
| Population−averaged prevalence | | | | 1.00 [0.53, 1.38] |

$Tau^2 = 0.51$   $I^2 = 99.6\%$

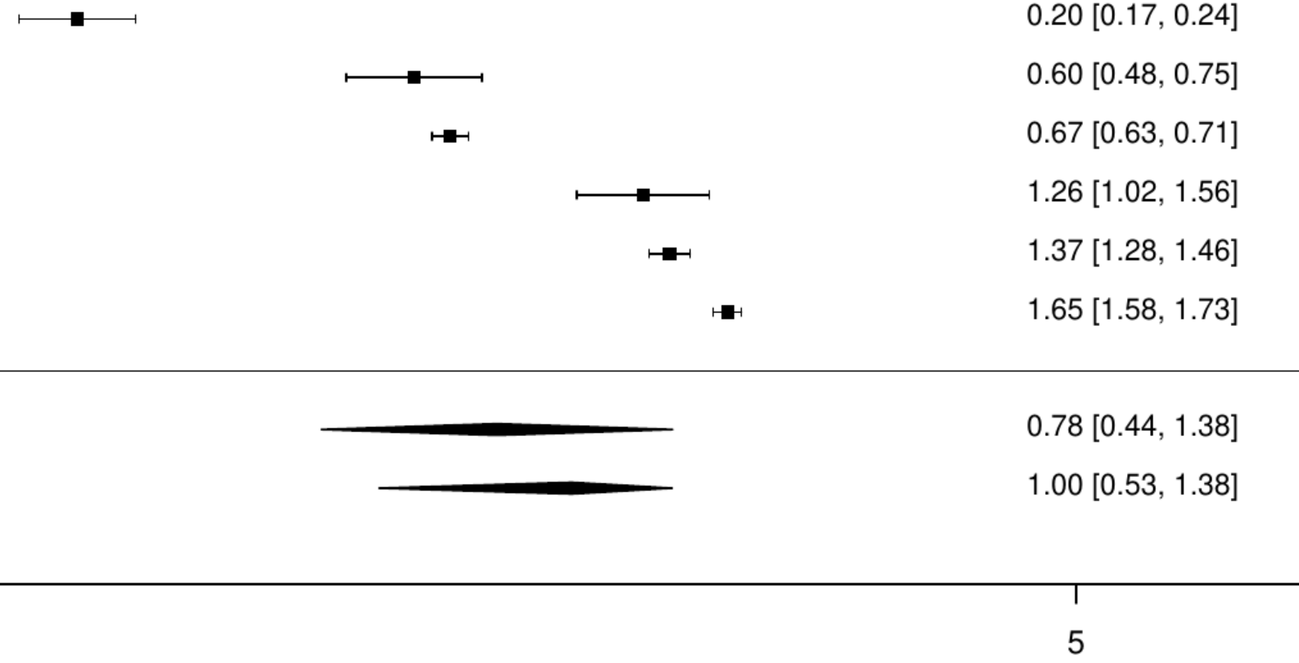

**Figure S86. Pooled prevalence of any cancer in hypertension (one study excluded).**

| Author(s) and Year | Country | Events | N | Prevalence [95% CI] |
|---|---|---|---|---|
| Kang E, 2019 | KOR | 106 | 22853 | 0.46 [ 0.38, 0.56] |
| Zhuang Pan, 2021 | CHN | 57 | 8095 | 0.70 [ 0.54, 0.91] |
| Roussel R, 2015 | FRA | 412 | 31652 | 1.30 [ 1.18, 1.43] |
| Nishimura S, 2020 | JPN | 508 | 31592 | 1.61 [ 1.48, 1.75] |
| Qian J, 2016 | CHN | 52 | 2939 | 1.77 [ 1.35, 2.31] |
| Jorgensen ME, 2015 | DEN | 1418 | 55320 | 2.56 [ 2.43, 2.70] |
| Altin C, 2022 | TUR | 41 | 1015 | 4.04 [ 2.99, 5.44] |
| Rea F, 2020 | ITA | 2885 | 60526 | 4.77 [ 4.60, 4.94] |
| Bugeja A, 2024 | CAN | 24275 | 266273 | 9.12 [ 9.01, 9.23] |
| Sinnott, 2019 | UK | 882 | 8639 | 10.21 [ 9.59, 10.87] |
| Kim Ju Hwan, 2021 | KOR | 138 | 1290 | 10.70 [ 9.12, 12.51] |
| Chien SC, 2015 | TW | 9538 | 79152 | 12.05 [11.83, 12.28] |
| Sheppard, 2021 | UK | 6307 | 45418 | 13.89 [13.57, 14.21] |
| El−Battrawy, 2021 | Multinational | 507 | 2850 | 17.79 [16.43, 19.24] |
| Douros A, 2021 | GER | 223 | 988 | 22.57 [20.07, 25.28] |
| Oh TK, 2021 | KOR | 27963 | 101657 | 27.51 [27.23, 27.78] |
| Median prevalence | | | | 5.12 [ 2.76, 9.31] |
| Population−averaged prevalence | | Tau² = 1.72 I² = 100% | | 9.01 [ 5.16, 13.54] |

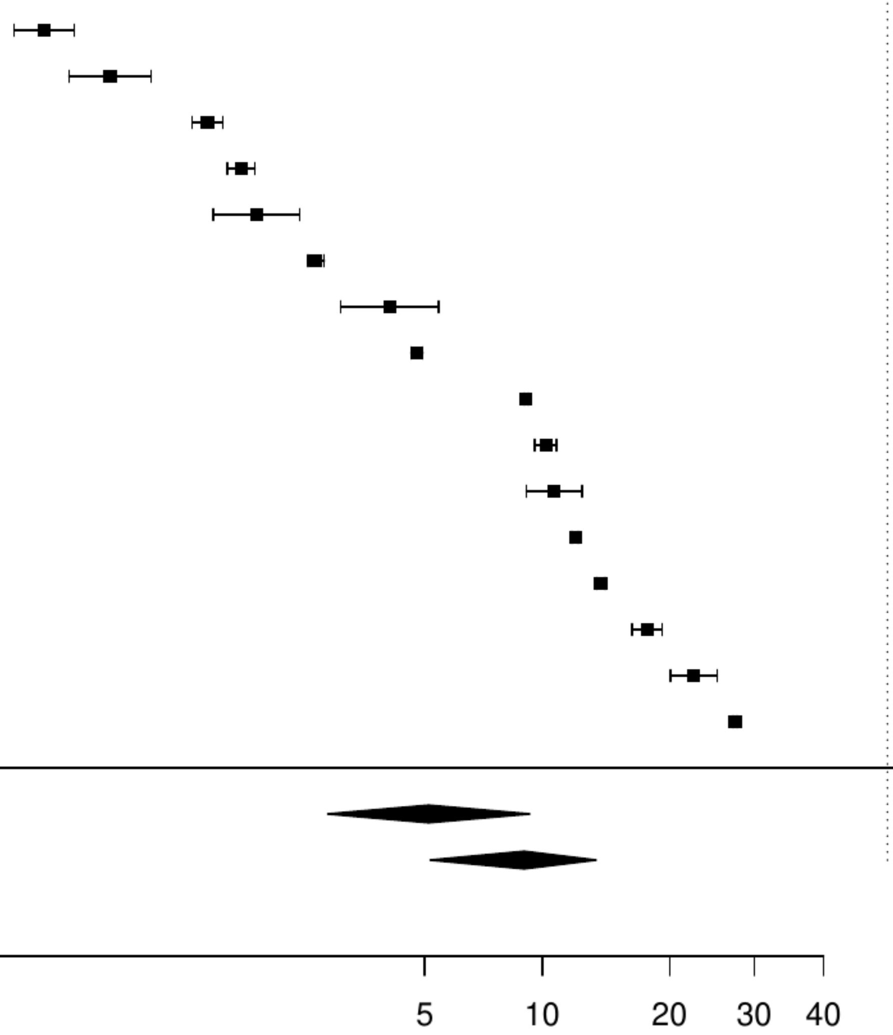

**Figure S87. Pooled prevalence of any cancer in hypertension.**

| Author(s) and Year | Country | Events | N | Prevalence [95% CI] |
|---|---|---|---|---|
| Kang E, 2019 | KOR | 106 | 22853 | 0.46 [ 0.38, 0.56] |
| Zhuang Pan, 2021 | CHN | 57 | 8095 | 0.70 [ 0.54, 0.91] |
| Roussel R, 2015 | FRA | 412 | 31652 | 1.30 [ 1.18, 1.43] |
| Nishimura S, 2020 | JPN | 508 | 31592 | 1.61 [ 1.48, 1.75] |
| Qian J, 2016 | CHN | 52 | 2939 | 1.77 [ 1.35, 2.31] |
| Jorgensen ME, 2015 | DEN | 1418 | 55320 | 2.56 [ 2.43, 2.70] |
| Altin C, 2022 | TUR | 41 | 1015 | 4.04 [ 2.99, 5.44] |
| Rea F, 2020 | ITA | 2885 | 60526 | 4.77 [ 4.60, 4.94] |
| Bugeja A, 2024 | CAN | 24275 | 266273 | 9.12 [ 9.01, 9.23] |
| Sinnott, 2019 | UK | 882 | 8639 | 10.21 [ 9.59, 10.87] |
| Kim Ju Hwan, 2021 | KOR | 138 | 1290 | 10.70 [ 9.12, 12.51] |
| Chien SC, 2015 | TW | 9538 | 79152 | 12.05 [11.83, 12.28] |
| Alanaeme, 2024 | USA | 119960300 | 9.74e+08 | 12.32 [12.31, 12.32] |
| Sheppard, 2021 | UK | 6307 | 45418 | 13.89 [13.57, 14.21] |
| El−Battrawy, 2021 | Multinational | 507 | 2850 | 17.79 [16.43, 19.24] |
| Douros A, 2021 | GER | 223 | 988 | 22.57 [20.07, 25.28] |
| Oh TK, 2021 | KOR | 27963 | 101657 | 27.51 [27.23, 27.78] |
| Median prevalence | | | | 5.40 [ 3.59, 8.05] |
| Population−averaged prevalence | | | | 9.30 [ 5.50, 13.33] |

Tau² = 1.67   I² = 100%

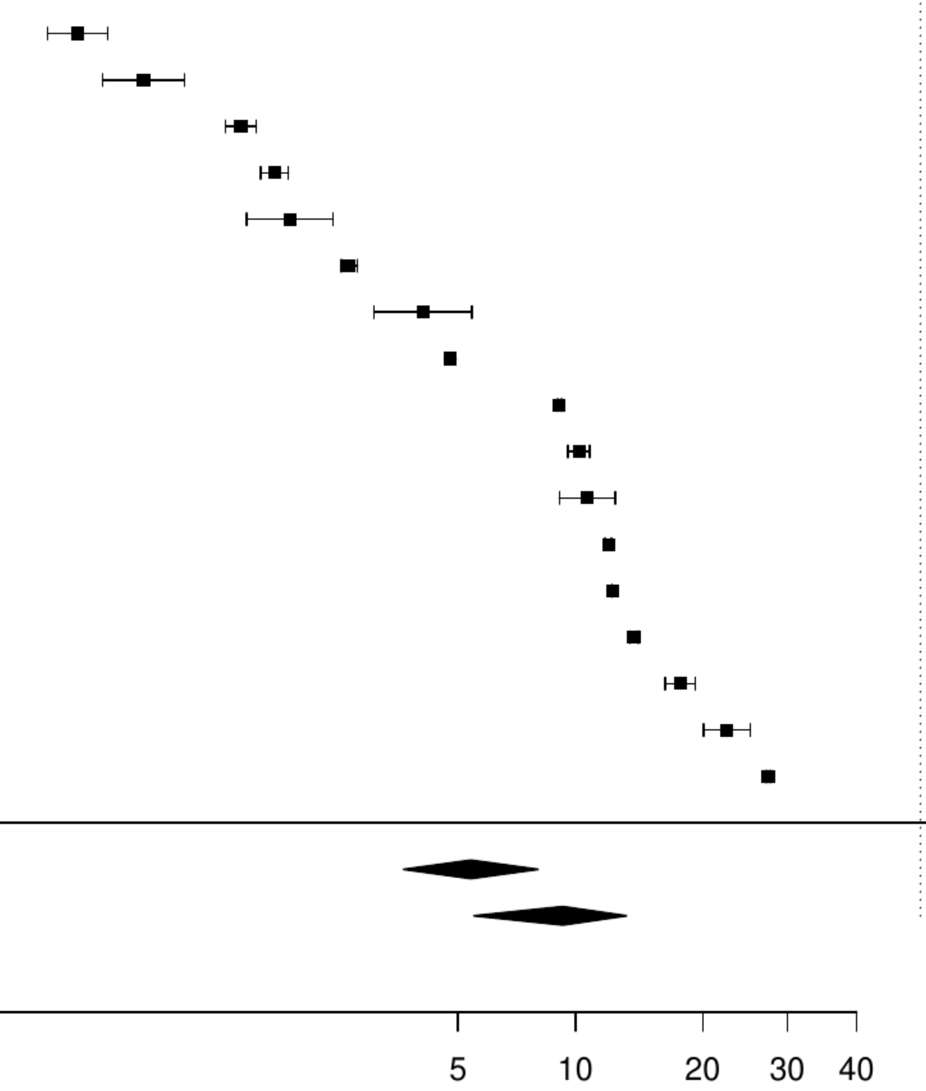

**Figure S88. Pooled prevalence of active cancer in the total population.**

| Author(s) and Year | Country | Events | N | Prevalence [95% CI] |
|---|---|---|---|---|
| Kim, 2020 | USA | 146 | 27739 | 0.53 [ 0.45, 0.62] |
| Ngo L, 2021 | Multinational | 212 | 35211 | 0.60 [ 0.53, 0.69] |
| Cheung KK, 2017 | CHN | 8 | 1239 | 0.65 [ 0.32, 1.29] |
| Aarnio K, 2016 | FIN | 11 | 970 | 1.13 [ 0.63, 2.04] |
| Rodriguez−Padial L, 2021 | ESP | 5425 | 445145 | 1.22 [ 1.19, 1.25] |
| Andersson, 2017 | SWE | 545 | 43145 | 1.26 [ 1.16, 1.37] |
| Lodzinski P, 2020 | POL | 10 | 691 | 1.45 [ 0.78, 2.67] |
| Kirchhof P, 2020 | Multinational | 105 | 6784 | 1.55 [ 1.28, 1.87] |
| Wiskar K, 2017 | USA | 1712 | 102746 | 1.67 [ 1.59, 1.75] |
| Itzhaki Ben Zadok O, 2019 | ISR | 53 | 3089 | 1.72 [ 1.31, 2.24] |
| Vergaro, 2019 | ITA | 50 | 2791 | 1.79 [ 1.36, 2.36] |
| Chandiramani, 2020 | Multinational | 44 | 2397 | 1.84 [ 1.37, 2.46] |
| Yamamoto K, 2024 | JPN | 234 | 12291 | 1.90 [ 1.68, 2.16] |
| Du Y, 2021 | USA | 18 | 729 | 2.47 [ 1.56, 3.88] |
| Halabi A, 2017 | AUS | 409 | 15912 | 2.57 [ 2.34, 2.83] |
| Hautamäki, 2020 | FIN | 41 | 1576 | 2.60 [ 1.92, 3.51] |
| Halvorsen S, 2022 | Multinational | 6102 | 219545 | 2.78 [ 2.71, 2.85] |
| Bernardini A, 2024 | ITA | 310 | 11078 | 2.80 [ 2.51, 3.12] |
| Atterman A, 2020 | SWE | 1518 | 52471 | 2.89 [ 2.75, 3.04] |
| Okamoto, 2022 | JPN | 5294 | 176275 | 3.00 [ 2.92, 3.08] |
| Patel, 2018 | AUS | 121 | 3944 | 3.07 [ 2.57, 3.65] |
| Hanon O, 2017 | Multinational | 233 | 7225 | 3.22 [ 2.84, 3.66] |
| Biccire FG, 2023 | ITA | 18 | 555 | 3.24 [ 2.05, 5.09] |
| Zaprutko J, 2017 | POL | 25 | 765 | 3.27 [ 2.22, 4.79] |
| Hess CN, 2015 | USA | 496 | 15008 | 3.30 [ 3.03, 3.60] |
| Chieffo A, 2016 | Multinational | 36 | 1019 | 3.53 [ 2.56, 4.86] |
| Valero−Masa, 2017 | ESP | 41 | 1111 | 3.69 [ 2.73, 4.97] |
| Minamino−Muta, 2017 | JPN | 149 | 3815 | 3.91 [ 3.34, 4.57] |
| Zahn, 2017 | GER | 55 | 1378 | 3.99 [ 3.08, 5.16] |
| Vinter N, 2023 | DEN | 1159 | 27947 | 4.15 [ 3.92, 4.39] |
| Bejot Y, 2020 | FRA | 45 | 1048 | 4.29 [ 3.22, 5.70] |
| Greene, 2022 | Multinational | 71 | 1619 | 4.39 [ 3.49, 5.50] |
| Lacoin L, 2017 | UK | 1822 | 41541 | 4.39 [ 4.19, 4.59] |
| Rubens FD, 2022 | CAN | 2130 | 45883 | 4.64 [ 4.45, 4.84] |
| Drusch S, 2024 | FRA | 5199 | 109693 | 4.74 [ 4.62, 4.87] |
| Ishizu, 2021 | JPN | 124 | 2588 | 4.79 [ 4.03, 5.68] |
| Mebazaa, 2018 | Multinational | 276 | 5387 | 5.12 [ 4.57, 5.75] |
| Mentias A, 2020 | USA | 6913 | 134717 | 5.13 [ 5.01, 5.25] |
| Coleman CI, 2021 | USA | 6170 | 116049 | 5.32 [ 5.19, 5.45] |
| Goderis Geert, 2017 | BEL | 33 | 578 | 5.71 [ 4.09, 7.92] |
| Yamaguchi, 2019 | JPN | 61 | 1012 | 6.03 [ 4.72, 7.67] |
| Pasceri, 2024 | Multinational | 411 | 6765 | 6.08 [ 5.53, 6.67] |
| Guo, 2022 | CHN | 202 | 3175 | 6.36 [ 5.56, 7.27] |
| Rome, 2021 | USA | 2432 | 35309 | 6.89 [ 6.63, 7.16] |
| Spatz ES, 2016 | USA | 188478 | 2581892 | 7.30 [ 7.27, 7.33] |
| Cao Davide, 2020 | USA | 719 | 9623 | 7.47 [ 6.96, 8.01] |
| Camelo−Castillo A, 2020 | ESP | 80 | 1044 | 7.66 [ 6.20, 9.44] |
| Rivera−Caravaca José Miguel, 2017 | ESP | 105 | 1361 | 7.71 [ 6.41, 9.26] |
| Sun LY, 2020 | CAN | 29291 | 352329 | 8.31 [ 8.22, 8.41] |
| Yamato H, 2020 | JPN | 100 | 1086 | 9.21 [ 7.63, 11.08] |
| Bertomeu−González V, 2015 | ESP | 88 | 948 | 9.28 [ 7.59, 11.30] |
| Werner N, 2020 | GER | 73 | 786 | 9.29 [ 7.45, 11.53] |
| Rutherford OW, 2020 | NOR | 6119 | 65563 | 9.33 [ 9.11, 9.56] |
| Bezin J, 2018 | FRA | 3041 | 31668 | 9.60 [ 9.28, 9.93] |
| Rivera−Caravaca JM, 2022 | Multinational | 147 | 1479 | 9.94 [ 8.51, 11.57] |
| Sax Dana R, 2021 | USA | 2855 | 26189 | 10.90 [10.53, 11.28] |
| Kanemaru K, 2020 | JPN | 3589 | 32726 | 10.97 [10.63, 11.31] |
| Okumura K, 2020 | JPN | 3589 | 32726 | 10.97 [10.63, 11.31] |
| Gon Y, 2020 | JPN | 185 | 1570 | 11.78 [10.28, 13.47] |
| Jiménez−Navarro MF, 2015 | USA | 700 | 5350 | 13.08 [12.21, 14.01] |
| Miró, 2017 | ESP | 859 | 6516 | 13.18 [12.38, 14.03] |
| Oh TK, 2020 | KOR | 1547 | 10440 | 14.82 [14.15, 15.51] |
| Median prevalence | Tau² = 0.66 I² = 99.9% | | | 4.11 [ 3.38, 5.00] |
| Population−averaged prevalence | | | | 5.36 [ 4.39, 6.33] |

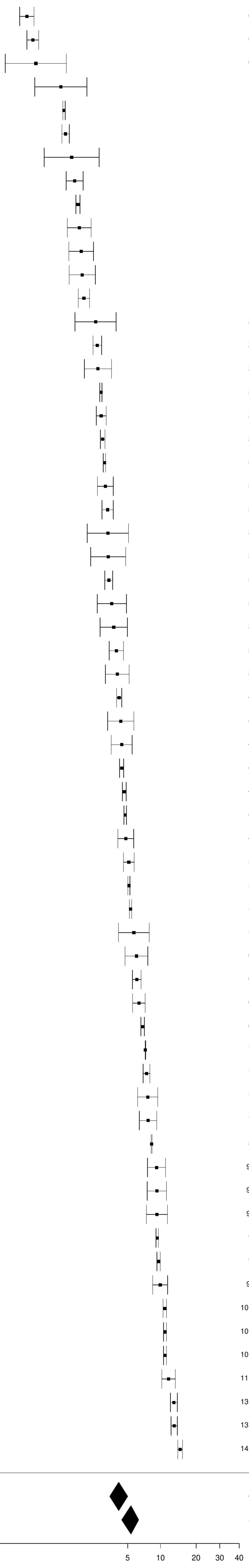

**Figure S89. Pooled prevalence of any cancer in the total population.**

| Author(s) and Year (420 studies omitted) | Country | Events | N | Prevalence [95% CI] |
|---|---|---|---|---|
| Rohla M, 2024 | CHE | | 1996 | 0.15 [ 0.08, 0.30] |
| Teece L, 2023 | UK | | 512388 | 0.41 [ 0.34, 0.48] |
| Yasaitis LC, 2020 | CAN | | 112484 | 1.25 [ 1.12, 1.40] |
| Rossini R, 2017 | ITA | 269 | 1082 | 1.85 [ 1.72, 1.98] |
| Altin C, 2022 | TUR | 41 | 1015 | 1.88 [ 1.60, 2.22] |
| Singh, 2022 | USA | 1864 | 15128 | 1.96 [ 1.83, 2.10] |
| Amann U, 2018 | GER | 90 | 1667 | 1.99 [ 1.90, 2.08] |
| Andersson, 2017 | SWE | | 43145 | 1.99 [ 1.92, 2.06] |
| Szummer, 2018 | SWE | | 205693 | 2.18 [ 1.91, 2.47] |
| Bouisset F, 2021 | FRA | | 6273 | 2.51 [ 2.39, 2.64] |
| Lin GM, 2015 | TW | 30 | 722 | 2.56 [ 2.36, 2.77] |
| Worrall−Carter Linda, 2016 | AUS | | 16771 | 2.80 [ 2.15, 3.64] |
| Wang F, 2016 | USA | | 2346 | 2.90 [ 2.77, 3.04] |
| Shore S, 2016 | USA | | 3283 | 3.03 [ 2.14, 4.28] |
| Forcadell MJ, 2018 | ESP | 162 | 1733 | 3.36 [ 3.17, 3.57] |
| Szummer K, 2017 | SWE | | 105674 | 3.41 [ 2.55, 4.54] |
| Álvarez−Álvarez B, 2015 | ESP | | 5302 | 3.60 [ 3.04, 4.26] |
| Wang Y, 2020 | USA | | 884931 | 4.49 [ 3.12, 6.42] |
| Bezin J, 2018 | FRA | | 31668 | 4.50 [ 4.08, 4.97] |
| Alter, 2018 | CAN | 1251 | 19544 | 4.61 [ 3.86, 5.50] |
| Steele AJ, 2018 | UK | 6576 | 82197 | 4.79 [ 4.62, 4.96] |
| Halabi A, 2017 | AUS | | 15912 | 4.92 [ 4.21, 5.74] |
| Chuang AM, 2018 | AUS | | 3057 | 4.94 [ 4.71, 5.18] |
| Oikawa T, 2019 | JPN | 123 | 870 | 5.09 [ 4.79, 5.40] |
| Rodriguez−Padial L, 2021 | ESP | | 445145 | 5.51 [ 5.29, 5.73] |
| Coll−Fernandez R, 2016 | ESP | 27 | 752 | 5.58 [ 5.27, 5.90] |
| Saia F, 2016 | ITA | 2777 | 39362 | 5.68 [ 5.37, 6.01] |
| Bentzel S, 2024 | SWE | | 3765 | 5.80 [ 5.55, 6.06] |
| Sabbag A, 2015 | ISR | | 11501 | 5.88 [ 5.72, 6.03] |
| Aboal J, 2020 | ESP | | 617 | 6.11 [ 5.12, 7.27] |
| Sanchez F, 2021 | ITA | | 1086 | 6.22 [ 6.10, 6.33] |
| Jamaly S, 2021 | SWE | | 25384 | 6.26 [ 5.96, 6.57] |
| Itzhaki Ben Zadok O, 2019 | ISR | 152 | 3089 | 6.40 [ 6.07, 6.75] |
| Palomäki A, 2021 | FIN | | 60446 | 6.71 [ 5.95, 7.57] |
| Lobo, 2017 | PRT | | 123442 | 7.05 [ 6.98, 7.13] |
| Valero−Masa, 2017 | ESP | | 1111 | 7.07 [ 6.32, 7.90] |
| Skalsky K, 2021 | ISR | 1667 | 15153 | 7.60 [ 6.81, 8.48] |
| Perl L, 2020 | ISR | | 25690 | 7.69 [ 7.67, 7.72] |
| Petersen JK, 2024 | DEN | 14311 | 139137 | 7.75 [ 7.19, 8.36] |
| Vallabhajosyula S, 2020 | USA | 3242 | 163071 | 7.78 [ 6.09, 9.90] |
| Tran, 2018 | CAN | 2269 | 41210 | 7.92 [ 6.78, 9.24] |
| Kyto V, 2022 | FIN | 6723 | 40654 | 8.03 [ 7.97, 8.10] |
| Montoy JCC, 2021 | USA | | 293589 | 8.04 [ 7.91, 8.17] |
| McCarthy CP, 2021 | USA | | 254422 | 8.07 [ 7.26, 8.96] |
| Ohya, 2017 | JPN | 163 | 1607 | 8.27 [ 8.16, 8.37] |
| Seki T, 2019 | JPN | 557 | 21409 | 8.42 [ 7.18, 9.85] |
| Kang, 2020 | KOR | | 108232 | 8.70 [ 8.33, 9.08] |
| Cilia, 2019 | USA | 850 | 10756 | 8.72 [ 7.40, 10.26] |
| Bjorklund E, 2021 | SWE | | 6558 | 8.78 [ 7.36, 10.44] |
| Mackay MH, 2017 | CAN | 2898 | 41792 | 8.98 [ 8.43, 9.56] |
| Sawano M, 2017 | JPN | 487 | 13024 | 9.07 [ 8.59, 9.58] |
| Blin P, 2021 | FRA | 47066 | 258260 | 9.32 [ 7.45, 11.60] |
| Khera R, 2017 | USA | | 478247 | 9.39 [ 8.41, 10.47] |
| Okamoto, 2022 | JPN | | 176275 | 9.47 [ 8.58, 10.44] |
| Marui A, 2015 | JPN | 198 | 1877 | 9.81 [ 9.64, 9.99] |
| Hwang IC, 2015 | KOR | | 8372 | 10.05 [ 8.47, 11.88] |
| Wu VC, 2021 | TW | 8687 | 170775 | 10.31 [ 9.94, 10.70] |
| Williams MJA, 2019 | NZL | 374 | 8305 | 10.67 [ 9.19, 12.37] |
| Hsieh CF, 2015 | TW | | 9805 | 10.99 [10.88, 11.10] |
| Akyea RK, 2022 | UK | 18311 | 143702 | 11.10 [10.52, 11.72] |
| Jacobs JA, 2019 | USA | | 609 | 11.13 [10.26, 12.06] |
| Fujita Y, 2022 | JPN | | 7785 | 11.40 [ 9.46, 13.68] |
| Takeji Y, 2020 | JPN | 1402 | 15231 | 12.31 [10.42, 14.50] |
| Erta. FS, 2016 | TUR | | 1034 | 12.48 [11.79, 13.20] |
| Wong CCY, 2021 | AUS | 43 | 10304 | 12.80 [12.57, 13.03] |
| Hautamäki, 2020 | FIN | | 1576 | 13.28 [12.48, 14.11] |
| De Luca, 2018 | ITA | 331 | 5070 | 13.74 [13.54, 13.94] |
| Spirito A, 2021 | CHE | 1810 | 16821 | 14.30 [14.06, 14.54] |
| Rohla M, 2015 | AUT | | 2890 | 14.31 [13.49, 15.17] |
| Nakatsuma, 2018 | JPN | | 12180 | 15.74 [13.21, 18.66] |
| Biccire FG, 2023 | ITA | | 555 | 16.54 [16.18, 16.90] |
| Chandiramani, 2020 | Multinational | | 2397 | 17.10 [16.85, 17.35] |
| Yamamoto K, 2024 | JPN | 1544 | 12291 | 17.58 [17.30, 17.86] |
| Yamaji K, 2018 | CHE | | 7640 | 18.86 [18.56, 19.15] |
| Lai CL, 2015 | TW | | 6584 | 21.90 [21.34, 22.47] |
| Cao Davide, 2020 | USA | | 9623 | 22.01 [21.42, 22.61] |
| Nakamura M, 2020 | JPN | 367 | 6267 | 22.97 [21.35, 24.68] |
| Tam DY, 2023 | CAN | | 22586 | 23.80 [22.97, 24.66] |
| Hess CN, 2015 | USA | | 15008 | 24.19 [23.18, 25.22] |
| Jiménez−Navarro MF, 2015 | USA | | 5350 | 38.66 [36.36, 41.02] |
| Population−averaged prevalence | | | | 10.27 [ 9.62, 10.92] |

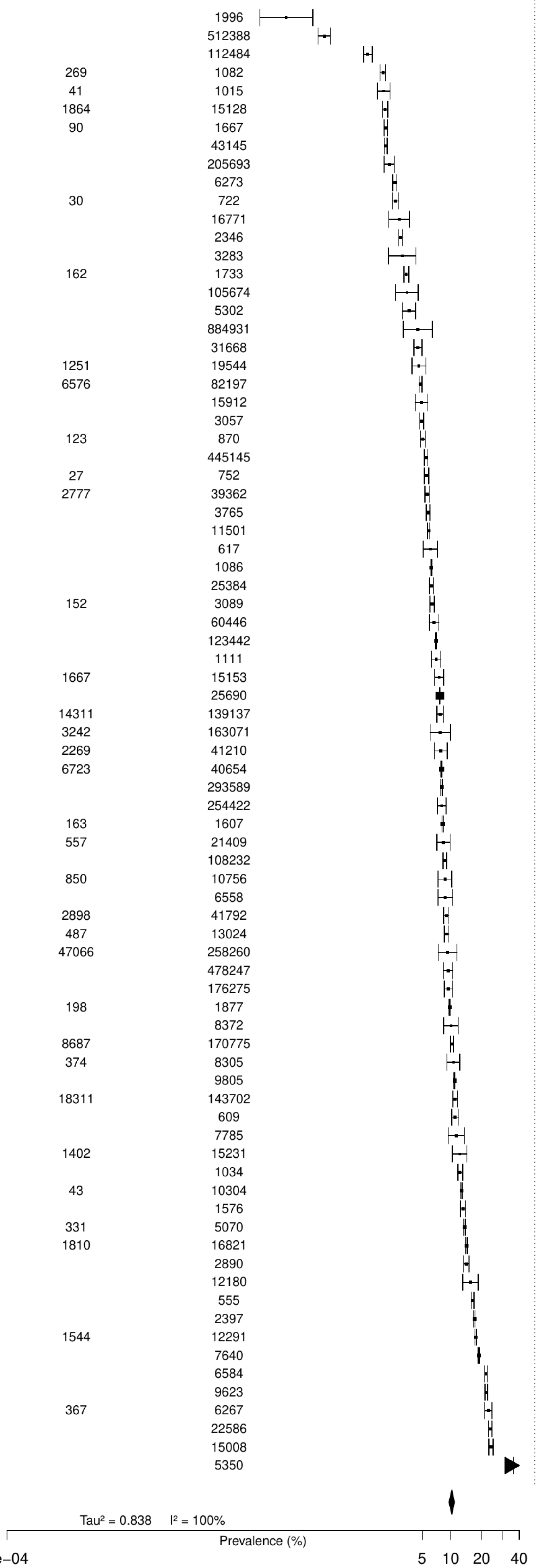

**Figure S90. Pooled prevalence of previous cancer in the total population.**

| Author(s) and Year | Country | Events | N | Prevalence [95% CI] |
|---|---|---|---|---|
| Lavielle P, 2015 | MEX | 12 | 1712 | 0.70 [ 0.40, 1.23] |
| Erta. FS, 2016 | TUR | 11 | 1034 | 1.06 [ 0.59, 1.91] |
| Takao T, 2021 | JPN | 36 | 2948 | 1.22 [ 0.88, 1.69] |
| Tanaka S, 2016 | JPN | 20 | 1532 | 1.31 [ 0.84, 2.01] |
| Kang, 2020 | KOR | 1459 | 108232 | 1.35 [ 1.28, 1.42] |
| Alherbish A, 2017 | CAN | 88 | 6262 | 1.41 [ 1.14, 1.73] |
| Szummer K, 2017 | SWE | 1957 | 105674 | 1.85 [ 1.77, 1.93] |
| Anguita–Gamez M, 2024 | ESP | 7693 | 354786 | 2.17 [ 2.12, 2.22] |
| Jamaly S, 2021 | SWE | 590 | 25384 | 2.32 [ 2.15, 2.52] |
| Szummer, 2018 | SWE | 5346 | 205693 | 2.60 [ 2.53, 2.67] |
| Choi IJ, 2021 | KOR | 38 | 1384 | 2.75 [ 2.00, 3.75] |
| Itzhaki Ben Zadok O, 2019 | ISR | 99 | 3089 | 3.20 [ 2.64, 3.89] |
| Captieux M, 2021 | UK | 6174 | 162316 | 3.80 [ 3.71, 3.90] |
| Sabbag A, 2015 | ISR | 466 | 11501 | 4.05 [ 3.71, 4.43] |
| Pu Aihua, 2017 | CAN | 849 | 20076 | 4.23 [ 3.96, 4.52] |
| Kheirbek RE, 2015 | USA | 78 | 1790 | 4.36 [ 3.50, 5.41] |
| Fujita Y, 2022 | JPN | 370 | 7785 | 4.75 [ 4.30, 5.25] |
| Liao CT, 2021 | TW | 83 | 1630 | 5.09 [ 4.12, 6.27] |
| Rohla M, 2015 | AUT | 152 | 2890 | 5.26 [ 4.50, 6.14] |
| Di Monaco A, 2022 | ITA | 68 | 1260 | 5.40 [ 4.28, 6.79] |
| Casetta I, 2020 | ITA | 171 | 3045 | 5.62 [ 4.85, 6.49] |
| Nystrom T, 2015 | SWE | 3039 | 53820 | 5.65 [ 5.45, 5.84] |
| Park SJ, 2023 | KOR | 1457 | 24374 | 5.98 [ 5.69, 6.28] |
| Puymirat E, 2016 | FRA | 161 | 2679 | 6.01 [ 5.17, 6.98] |
| Targher Giovanni, 2016 | ITA | 108 | 1776 | 6.08 [ 5.06, 7.29] |
| Sattar Naveed, 2019 | SWE | 14988 | 241278 | 6.21 [ 6.12, 6.31] |
| Lee SE, 2020 | KOR | 45 | 717 | 6.28 [ 4.72, 8.30] |
| Ajabnoor AM, 2023 | UK | 11379 | 177065 | 6.43 [ 6.31, 6.54] |
| Vergaro, 2019 | ITA | 197 | 2791 | 7.06 [ 6.17, 8.07] |
| Álvarez–Álvarez B, 2015 | ESP | 381 | 5302 | 7.19 [ 6.52, 7.91] |
| Shore S, 2016 | USA | 240 | 3283 | 7.31 [ 6.47, 8.25] |
| Abu–Assi Emad, 2016 | ESP | 57 | 767 | 7.43 [ 5.78, 9.51] |
| Danchin, 2020 | FRA | 398 | 5147 | 7.73 [ 7.03, 8.49] |
| Bouisset F, 2021 | FRA | 491 | 6243 | 7.86 [ 7.22, 8.56] |
| Fendler TJ, 2015 | USA | 96 | 1173 | 8.18 [ 6.75, 9.90] |
| Teece L, 2023 | UK | 42187 | 512388 | 8.23 [ 8.16, 8.31] |
| Okano T, 2020 | JPN | 58 | 676 | 8.58 [ 6.69, 10.94] |
| Gurusamy VK, 2019 | SWE | 6043 | 68056 | 8.88 [ 8.67, 9.10] |
| Rohla M, 2024 | CHE | 179 | 1996 | 8.97 [ 7.79, 10.30] |
| Nakatsuma, 2018 | JPN | 1109 | 12180 | 9.11 [ 8.61, 9.63] |
| Velders MA, 2020 | SWE | 16237 | 175146 | 9.27 [ 9.14, 9.41] |
| Yokomichi H, 2017 | JPN | 2859 | 30834 | 9.27 [ 8.95, 9.60] |
| Komorita Y, 2020 | JPN | 462 | 4923 | 9.38 [ 8.60, 10.23] |
| Jacobs JA, 2019 | USA | 58 | 609 | 9.52 [ 7.43, 12.12] |
| Ortiz MR, 2020 | ESP | 55 | 574 | 9.58 [ 7.43, 12.28] |
| Corica B, 2024 | Multinational | 2388 | 24574 | 9.72 [ 9.35, 10.09] |
| Aboal J, 2020 | ESP | 60 | 617 | 9.72 [ 7.62, 12.33] |
| Hwang IC, 2015 | KOR | 860 | 8372 | 10.27 [ 9.64, 10.94] |
| Perl L, 2020 | ISR | 2696 | 25690 | 10.49 [10.13, 10.88] |
| Biccire FG, 2023 | ITA | 59 | 555 | 10.63 [ 8.33, 13.48] |
| Friberg Leif, 2018 | SWE | 33238 | 312341 | 10.64 [10.53, 10.75] |
| Yamaji K, 2018 | CHE | 815 | 7640 | 10.67 [ 9.99, 11.38] |
| Matsuhiro Y, 2021 | JPN | 94 | 871 | 10.79 [ 8.90, 13.03] |
| Tam DY, 2023 | CAN | 2468 | 22586 | 10.93 [10.53, 11.34] |
| Okumura K, 2020 | JPN | 3600 | 32726 | 11.00 [10.67, 11.34] |
| Wang F, 2016 | USA | 263 | 2346 | 11.21 [10.00, 12.55] |
| Abdel–Qadir H, 2022 | CAN | 39305 | 347632 | 11.31 [11.20, 11.41] |
| Chuang AM, 2018 | AUS | 348 | 3071 | 11.33 [10.26, 12.50] |
| Kristiansen MH, 2023 | DEN | 64 | 538 | 11.90 [ 9.42, 14.91] |
| Palomäki A, 2021 | FIN | 7263 | 60446 | 12.02 [11.76, 12.28] |
| Settergren C, 2024 | SWE | 12477 | 100584 | 12.40 [12.20, 12.61] |
| Finkelstein A, 2019 | ISR | 292 | 2336 | 12.50 [11.22, 13.90] |
| Chen Q, 2024 | NLD | 40224 | 320139 | 12.56 [12.45, 12.68] |
| Rodionov RN, 2021 | GER | 12790 | 101162 | 12.64 [12.44, 12.85] |
| de la Torre Hernandez Jose M, 2021 | ESP | 161 | 1249 | 12.89 [11.14, 14.87] |
| Vanbeselaere, 2016 | BEL | 237 | 1830 | 12.95 [11.49, 14.57] |
| Gajanana D, 2020 | USA | 975 | 7499 | 13.00 [12.26, 13.78] |
| Tybjerg AJ, 2020 | DEN | 11407 | 85893 | 13.28 [13.06, 13.51] |
| Miyata S, 2017 | JPN | 1366 | 10209 | 13.38 [12.73, 14.05] |
| Sanchez F, 2021 | ITA | 146 | 1086 | 13.44 [11.54, 15.60] |
| Biasco L, 2024 | CHE | 79 | 581 | 13.60 [11.04, 16.63] |
| Al–Salameh, 2020 | FRA | 134 | 983 | 13.63 [11.63, 15.92] |
| Dalen M, 2024 | SWE | 461 | 3267 | 14.11 [12.96, 15.35] |
| Bhonsale A, 2024 | USA | 9510 | 67221 | 14.15 [13.89, 14.41] |
| Lim SL, 2017 | SWE | 2712 | 19047 | 14.24 [13.75, 14.74] |
| O'Neal WT, 2015 | USA | 2248 | 15428 | 14.57 [14.02, 15.14] |
| Sogaard M, 2024 | DEN | 5406 | 36223 | 14.92 [14.56, 15.29] |
| Bjorklund E, 2021 | SWE | 980 | 6558 | 14.94 [14.10, 15.83] |
| Ganesh A, 2017 | UK | 221 | 1425 | 15.51 [13.72, 17.48] |
| Bentzel S, 2024 | SWE | 591 | 3765 | 15.70 [14.57, 16.89] |
| Glaser N, 2024 | SWE | 3311 | 21002 | 15.77 [15.28, 16.26] |
| Kubo, 2018 | JPN | 92 | 563 | 16.34 [13.51, 19.63] |
| Bisson A, 2018 | FRA | 40456 | 240459 | 16.82 [16.68, 16.97] |
| Meyer M, 2021 | SWE | 2437 | 14434 | 16.88 [16.28, 17.50] |
| Melgaard L, 2020 | DEN | 4887 | 28770 | 16.99 [16.56, 17.42] |
| Nielsen PB, 2021 | DEN | 401 | 2285 | 17.55 [16.04, 19.16] |
| Reznik ME, 2018 | USA | 189 | 1003 | 18.84 [16.54, 21.38] |
| Lantelme P, 2020 | FRA | 4065 | 20443 | 19.88 [19.34, 20.44] |
| Clementy N, 2021 | FRA | 5859 | 29422 | 19.91 [19.46, 20.37] |
| Kiramijyan S, 2016 | USA | 118 | 589 | 20.03 [17.00, 23.46] |
| Saliba W, 2015 | ISR | 6710 | 32912 | 20.39 [19.96, 20.83] |
| Frank, 2019 | Multinational | 407 | 1946 | 20.91 [19.17, 22.78] |
| Kim K, 2021 | KOR | 138902 | 615724 | 22.56 [22.45, 22.66] |
| Garry Elizabeth M, 2018 | USA | 57181 | 247356 | 23.12 [22.95, 23.28] |
| Wideqvist M, 2022 | SWE | 14458 | 62228 | 23.23 [22.90, 23.57] |
| Benfari, 2019 | USA | 2921 | 11507 | 25.38 [24.60, 26.19] |
| Noseworthy PA, 2022 | USA | 153663 | 562850 | 27.30 [27.18, 27.42] |
| Abbas SS, 2021 | AUS | 521 | 1206 | 43.20 [40.43, 46.02] |
| Hsieh CF, 2015 | TW | 6821 | 9805 | 69.57 [68.65, 70.47] |
| Median prevalence | | | | 9.18 [ 7.82, 10.76] |
| Population–averaged prevalence | | | | 11.72 [10.10, 13.49] |

Tau² = 0.79 I² = 100%

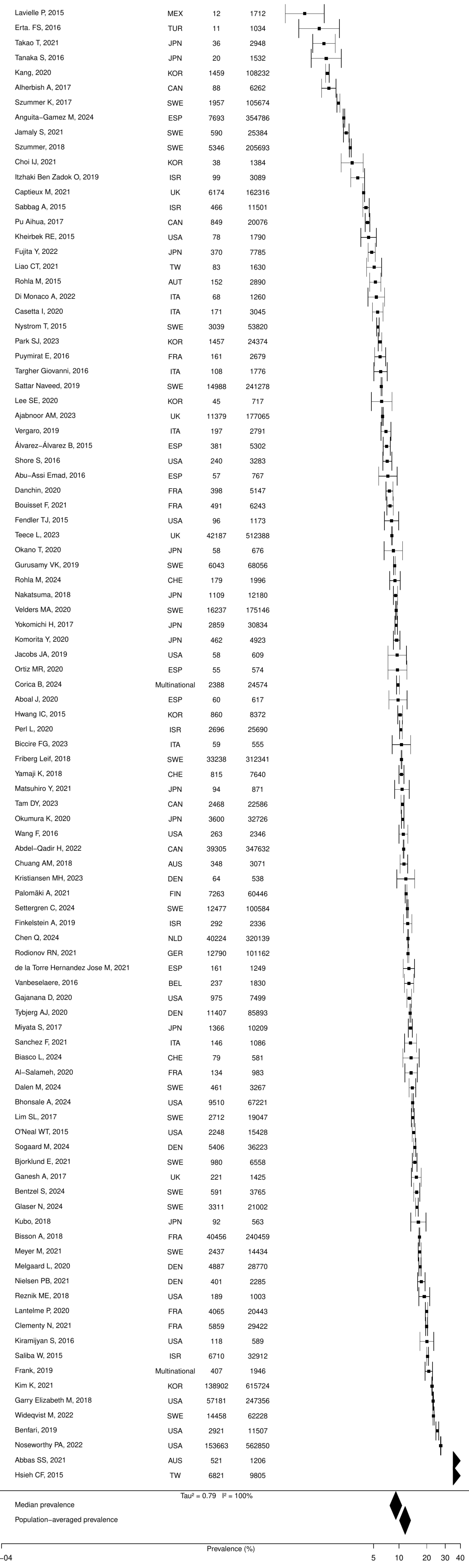

**Figure S91. Pooled prevalence of blood cancer in the total population.**

| Author(s) and Year | Country | Events | N | Prevalence [95% CI] |
|---|---|---|---|---|
| Lindgren A, 2019 | Multinational | 9 | 8525 | 0.11 [0.05, 0.20] |
| Sessa, 2019 | ITA | 19 | 10091 | 0.19 [0.12, 0.29] |
| Lai CL, 2015 | TW | 11 | 5587 | 0.20 [0.11, 0.36] |
| Si, 2019 | CHN | 35 | 10331 | 0.34 [0.24, 0.47] |
| Hess CN, 2015 | USA | 55 | 15008 | 0.37 [0.28, 0.48] |
| Lobo, 2017 | PRT | 619 | 123442 | 0.50 [0.46, 0.54] |
| Hreha, 2020 | USA | 1369 | 269314 | 0.51 [0.48, 0.54] |
| Machado−Duque ME, 2023 | Colombia | 11 | 2076 | 0.53 [0.29, 0.95] |
| Pack QR, 2016 | USA | 230 | 38532 | 0.60 [0.52, 0.68] |
| Montoy JCC, 2021 | USA | 1800 | 293589 | 0.61 [0.59, 0.64] |
| Corraini P, 2018 | DEN | 1242 | 201691 | 0.62 [0.58, 0.65] |
| Vanbeselaere, 2016 | BEL | 12 | 1830 | 0.66 [0.37, 1.15] |
| McCarthy CP, 2021 | USA | 1917 | 254422 | 0.75 [0.72, 0.79] |
| Lagu Tara, 2016 | USA | 115 | 13163 | 0.87 [0.73, 1.05] |
| Munir, 2017 | USA | 3406 | 388340 | 0.88 [0.85, 0.91] |
| Ripley Diane C Cowper, 2015 | USA | 101 | 10430 | 0.97 [0.80, 1.18] |
| Schmidt M, 2016 | DEN | 3374 | 317161 | 1.06 [1.03, 1.10] |
| Proietti, 2019 | ITA | 276 | 24040 | 1.15 [1.02, 1.29] |
| Khera R, 2017 | USA | 5739 | 478247 | 1.20 [1.17, 1.23] |
| Cocchieri A, 2015 | ITA | 15 | 1192 | 1.26 [0.76, 2.08] |
| Downer, 2024 | UK | 31 | 2454 | 1.26 [0.89, 1.79] |
| Pham P, 2020 | USA | 67 | 4869 | 1.38 [1.08, 1.74] |
| Gilstrap L, 2021 | USA | 4818 | 295494 | 1.63 [1.59, 1.68] |
| Mentias A, 2020 | USA | 2451 | 134717 | 1.82 [1.75, 1.89] |
| Nielsen PB, 2021 | DEN | 42 | 2285 | 1.84 [1.36, 2.48] |
| McCullough, 2019 | USA | 171 | 8757 | 1.95 [1.68, 2.26] |
| Gamst J, 2015 | SWE | 117 | 5065 | 2.31 [1.93, 2.76] |
| Median prevalence | | | | 0.76 [0.58, 1.00] |
| Population−averaged prevalence | | | | 0.97 [0.73, 1.20] |

Tau² = 0.51 I² = 99.8%

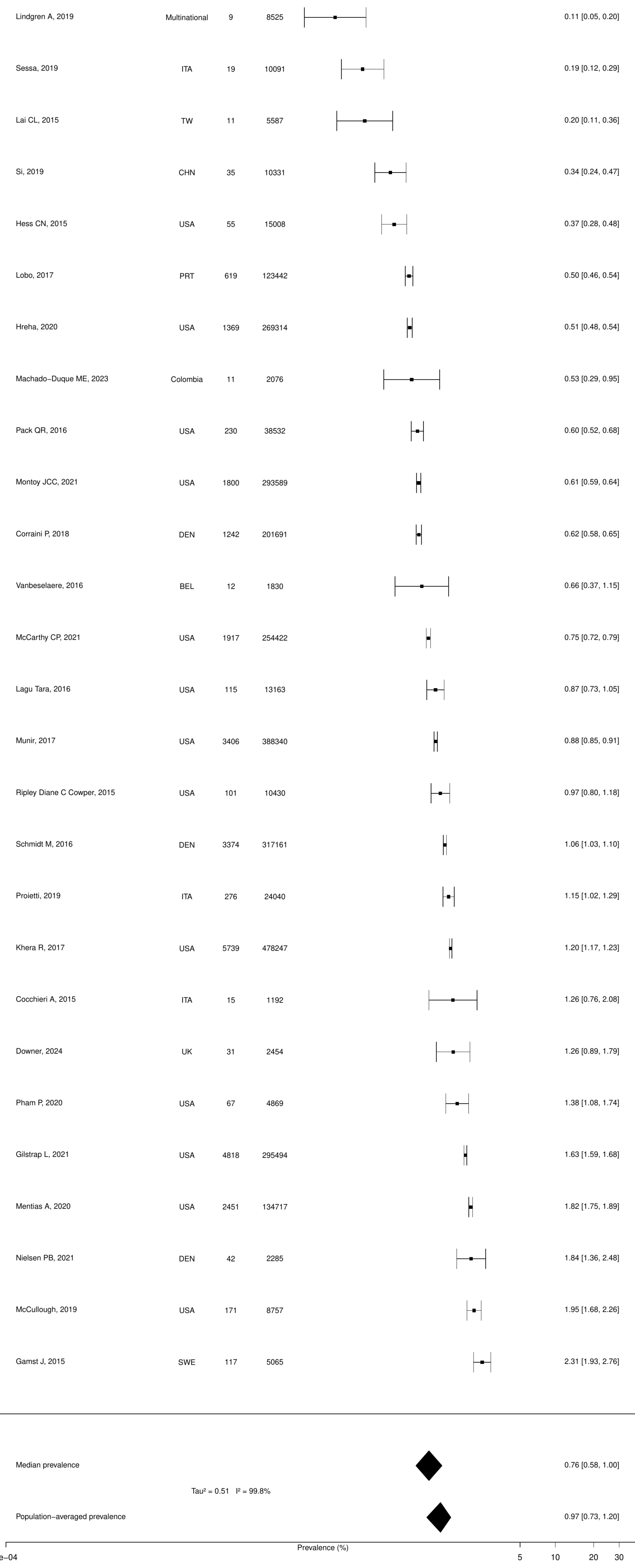

**Figure S92. Pooled prevalence of metastatic cancer in the total population.**

| Author(s) and Year | Country | Events | N | Prevalence [95% CI] |
|---|---|---|---|---|
| Vallabhajosyula S, 2020 | USA | 49 | 163071 | 0.03 [0.02, 0.04] |
| Seki T, 2019 | JPN | 25 | 21409 | 0.12 [0.08, 0.17] |
| Pack QR, 2016 | USA | 57 | 38532 | 0.15 [0.11, 0.19] |
| Lindgren A, 2019 | Multinational | 15 | 8525 | 0.18 [0.11, 0.29] |
| Mehta HB, 2017 | UK | 106 | 53055 | 0.20 [0.17, 0.24] |
| Kerola, 2022 | FIN | 18 | 8934 | 0.20 [0.13, 0.32] |
| Lai CL, 2015 | TW | 16 | 5587 | 0.29 [0.18, 0.47] |
| Kyto V, 2022 | FIN | 142 | 40654 | 0.35 [0.30, 0.41] |
| Kōnig, 2018 | GER | 646 | 161502 | 0.40 [0.37, 0.43] |
| Rubens FD, 2022 | CAN | 208 | 45883 | 0.45 [0.40, 0.52] |
| Lobo, 2017 | PRT | 619 | 123442 | 0.50 [0.46, 0.54] |
| Christiansen, 2020 | DEN | 1541 | 280002 | 0.55 [0.52, 0.58] |
| Johnston, 2019 | USA | 78 | 13026 | 0.60 [0.48, 0.75] |
| Captieux M, 2021 | UK | 1093 | 162316 | 0.67 [0.63, 0.71] |
| Corraini P, 2018 | DEN | 1422 | 201691 | 0.71 [0.67, 0.74] |
| Bezin J, 2018 | FRA | 226 | 31668 | 0.71 [0.63, 0.81] |
| Jiménez−Navarro MF, 2015 | USA | 39 | 5350 | 0.73 [0.53, 1.00] |
| Castro, 2017 | BRA | 7 | 959 | 0.73 [0.35, 1.52] |
| Schmidt M, 2016 | DEN | 2342 | 317161 | 0.74 [0.71, 0.77] |
| Hautamāki, 2020 | FIN | 13 | 1576 | 0.82 [0.48, 1.42] |
| Hreha, 2020 | USA | 2268 | 269314 | 0.84 [0.81, 0.88] |
| Williams MJA, 2019 | NZL | 71 | 8305 | 0.85 [0.68, 1.08] |
| Patel A, 2022 | AUS | 542 | 60953 | 0.89 [0.82, 0.97] |
| Yang H, 2023 | USA | 7 | 785 | 0.89 [0.43, 1.86] |
| Lagu Tara, 2016 | USA | 120 | 13163 | 0.91 [0.76, 1.09] |
| Wiskar K, 2017 | USA | 963 | 102746 | 0.94 [0.88, 1.00] |
| Alter, 2018 | CAN | 186 | 19544 | 0.95 [0.82, 1.10] |
| Rome, 2021 | USA | 342 | 35309 | 0.97 [0.87, 1.08] |
| Singh, 2022 | USA | 158 | 15128 | 1.04 [0.89, 1.22] |
| Downer, 2024 | UK | 26 | 2454 | 1.06 [0.72, 1.55] |
| Pham P, 2020 | USA | 52 | 4869 | 1.07 [0.81, 1.40] |
| Yasaitis LC, 2020 | CAN | 1237 | 112484 | 1.10 [1.04, 1.16] |
| Spoon DB, 2015 | USA | 74 | 6636 | 1.12 [0.89, 1.40] |
| Mentias A, 2020 | USA | 1593 | 134717 | 1.18 [1.13, 1.24] |
| Si, 2019 | CHN | 129 | 10331 | 1.25 [1.05, 1.48] |
| Gomes C, 2020 | PRT | 83 | 6596 | 1.26 [1.02, 1.56] |
| Tran, 2018 | CAN | 556 | 41210 | 1.35 [1.24, 1.47] |
| Munir, 2017 | USA | 5307 | 388340 | 1.37 [1.33, 1.40] |
| Oh TK, 2020 | KOR | 874 | 63739 | 1.37 [1.28, 1.46] |
| Oh TK, 2020 | KOR | 154 | 10440 | 1.48 [1.26, 1.73] |
| Sun LY, 2019 | CAN | 1283 | 82125 | 1.56 [1.48, 1.65] |
| Gilstrap L, 2021 | USA | 4709 | 295494 | 1.59 [1.55, 1.64] |
| Udell JA, 2018 | USA | 1843 | 111576 | 1.65 [1.58, 1.73] |
| Hall, 2019 | CAN | 116 | 6988 | 1.66 [1.39, 1.99] |
| Kim, 2020 | USA | 472 | 27739 | 1.70 [1.56, 1.86] |
| McCarthy CP, 2021 | USA | 4342 | 254422 | 1.71 [1.66, 1.76] |
| Kheirbek RE, 2016 | USA | 1072 | 60666 | 1.77 [1.67, 1.88] |
| Worrall−Carter Linda, 2016 | AUS | 318 | 16771 | 1.90 [1.70, 2.11] |
| Sun LY, 2020 | CAN | 6786 | 352329 | 1.93 [1.88, 1.97] |
| Lutsey PL, 2020 | USA | 3266 | 167275 | 1.95 [1.89, 2.02] |
| Field, 2019 | KOR | 303 | 15276 | 1.98 [1.77, 2.22] |
| Muhlestein J B, 2016 | USA | 128 | 6414 | 2.00 [1.68, 2.37] |
| Proietti, 2019 | ITA | 503 | 24040 | 2.09 [1.92, 2.28] |
| Ripley Diane C Cowper, 2015 | USA | 225 | 10430 | 2.16 [1.90, 2.45] |
| Cocchieri A, 2015 | ITA | 26 | 1192 | 2.18 [1.49, 3.18] |
| Bourrier, 2020 | CAN | 494 | 20600 | 2.40 [2.20, 2.62] |
| Montoy JCC, 2021 | USA | 7404 | 293589 | 2.52 [2.47, 2.58] |
| Arslanian−Engoren C, 2018 | USA | 243 | 9548 | 2.55 [2.25, 2.88] |
| Chung, 2017 | KOR | 525 | 19128 | 2.74 [2.52, 2.99] |
| Jun, 2017 | CAN | 496 | 17988 | 2.76 [2.53, 3.01] |
| Chen CY, 2015 | USA | 666 | 23111 | 2.88 [2.67, 3.11] |
| Gamst J, 2015 | SWE | 149 | 5065 | 2.94 [2.51, 3.44] |
| McCullough, 2019 | USA | 300 | 8757 | 3.43 [3.06, 3.83] |
| Haug V, 2021 | USA | 272 | 5386 | 5.05 [4.50, 5.67] |
| Ryden A, 2018 | SWE | 514 | 10039 | 5.12 [4.71, 5.57] |
| Wang Y, 2020 | USA | 48910 | 884931 | 5.53 [5.48, 5.57] |
| Arruda−Olson AM, 2018 | USA | 100 | 1676 | 5.97 [4.93, 7.21] |
| Nielsen PB, 2021 | DEN | 172 | 2285 | 7.53 [6.51, 8.68] |
| Median prevalence | | | | 1.12 [0.89, 1.41] |
| Population−averaged prevalence | | | | 1.73 [1.38, 2.13] |

Tau² = 0.95  I² = 99.9%

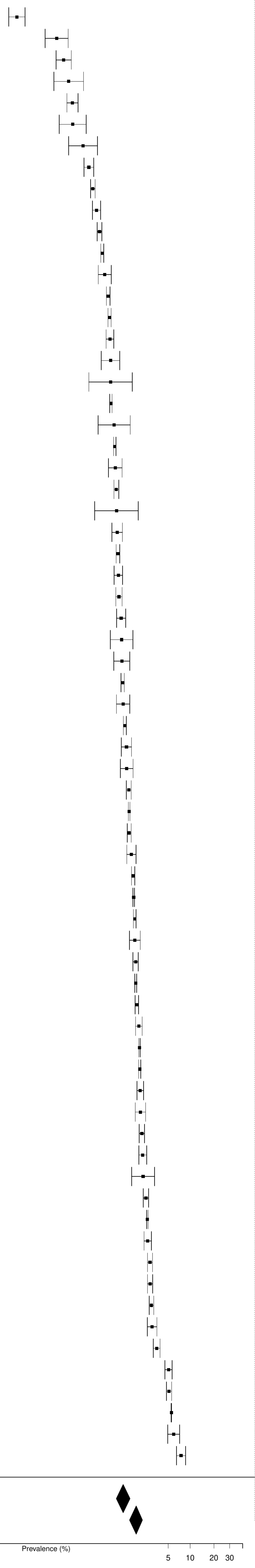

**Figure S93. Pooled prevalence of solid cancer in the total population.**

| Author(s) and Year | Country | Events | N | Prevalence [95% CI] |
|---|---|---|---|---|
| Lindgren A, 2019 | Multinational | 58 | 8525 | 0.68 [ 0.53, 0.88] |
| Sessa, 2019 | ITA | 82 | 10091 | 0.81 [ 0.65, 1.01] |
| Pack QR, 2016 | USA | 327 | 38532 | 0.85 [ 0.76, 0.95] |
| Machado−Duque ME, 2023 | Colombia | 21 | 2076 | 1.01 [ 0.66, 1.55] |
| Montoy JCC, 2021 | USA | 3054 | 293589 | 1.04 [ 1.00, 1.08] |
| Lagu Tara, 2016 | USA | 244 | 13163 | 1.85 [ 1.64, 2.10] |
| McCarthy CP, 2021 | USA | 5204 | 254422 | 2.05 [ 1.99, 2.10] |
| Munir, 2017 | USA | 8169 | 388340 | 2.10 [ 2.06, 2.15] |
| Hreha, 2020 | USA | 6626 | 269314 | 2.46 [ 2.40, 2.52] |
| Castro, 2017 | BRA | 25 | 959 | 2.61 [ 1.77, 3.83] |
| Si, 2019 | CHN | 368 | 10331 | 3.56 [ 3.22, 3.94] |
| Worrall−Carter Linda, 2016 | AUS | 690 | 16771 | 4.11 [ 3.82, 4.43] |
| Lai CL, 2015 | TW | 288 | 6584 | 4.37 [ 3.91, 4.90] |
| Gomes C, 2020 | PRT | 332 | 6596 | 5.03 [ 4.53, 5.59] |
| Johnston, 2019 | USA | 769 | 13026 | 5.90 [ 5.51, 6.32] |
| Schmidt M, 2016 | DEN | 20686 | 317161 | 6.52 [ 6.44, 6.61] |
| Hariri, 2022 | USA | 334 | 4921 | 6.79 [ 6.12, 7.52] |
| Corraini P, 2018 | DEN | 16652 | 201691 | 8.26 [ 8.14, 8.38] |
| Gilstrap L, 2021 | USA | 26391 | 295494 | 8.93 [ 8.83, 9.03] |
| Sun LY, 2019 | CAN | 7505 | 82125 | 9.14 [ 8.94, 9.34] |
| Khera R, 2017 | USA | 49456 | 478247 | 10.34 [10.26, 10.43] |
| Downer, 2024 | UK | 339 | 2454 | 13.81 [12.50, 15.24] |
| Nielsen PB, 2021 | DEN | 323 | 2285 | 14.14 [12.77, 15.63] |
| Parajuli DR, 2021 | AUS | 176 | 1184 | 14.86 [12.95, 17.01] |
| Chung, 2017 | KOR | 2892 | 19128 | 15.12 [14.62, 15.63] |
| McCullough, 2019 | USA | 1374 | 8757 | 15.69 [14.94, 16.47] |
| Pham P, 2020 | USA | 780 | 4869 | 16.02 [15.02, 17.08] |
| Ripley Diane C Cowper, 2015 | USA | 1760 | 10430 | 16.87 [16.17, 17.61] |
| Median prevalence | | | | 4.68 [ 3.19, 6.82] |
| Population−averaged prevalence | | | | 7.15 [ 4.77, 9.41] |

Tau² = 1.15   I² = 100%

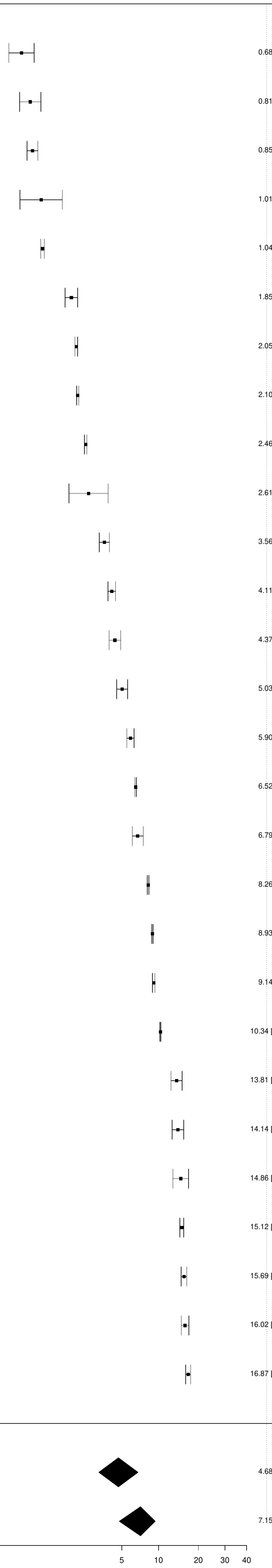


Prevalence (%)
