## Supplementary references for "“Prevalence of cancer in patients with cardiovascular diseases and risk factors: a systematic review and meta-analysis”"

21. Alsawas M, Wang Z, Murad MH, Yousufuddin M. Gender disparities among hospitalised patients with acute myocardial infarction, acute decompensated heart failure or pneumonia:

retrospective cohort study. BMJ open. 2019 Jan 21;9(1):e022782.

31. Anguita-Gamez M, Esteban-Fernandez A, Bonilla-Palomas JL, Bernal JL, Del Prado N,

Fernandez-Perez C, et al. Gender differences in clinical features and outcomes of patients over 75 years presenting with acute heart failure. Results of a nationwide study (2016-2019). Cardiology journal. 2024;31(3):427–33.

41. Arutyunov AG, Dragunov DO, Arutyunov GP, Rylova AK, Pashkevich DD, Viter KV. [First Open Study of Syndrome of Acute Decompensation of Heart Failure and Concomitant

diseases in Russian Federation: Independent Registry ORAKUL]. Kardiologiia. 2015;55(5):12–21.

73. Blecker S, Park H, Katz SD. Association of HbA1c with hospitalization and mortality

among patients with heart failure and diabetes. BMC cardiovascular disorders. 2016 May 20;16:99.

83. Buckley BJR, Harrison SL, Fazio-Eynullayeva E, Underhill P, Lane DA, Thijssen DHJ, et

al. Exercise-Based Cardiac Rehabilitation and All-Cause Mortality Among Patients With Atrial Fibrillation. Journal of the American Heart Association. 2021 June 15;10(12):e020804.

104. Chen J, Borra S, Fan L, Huang A, Patel D, Juneja R. Treatment patterns and outcomes before and after human regular U-500 insulin initiation via KwikPen(R) among US veterans with type 2 diabetes mellitus. Journal of diabetes and its complications. 2021

Oct;35(10):107995.

115. Chiang JI, Furler J, Mair F, Jani BD, Nicholl BI, Thuraisingam S, et al. Associations

between multimorbidity and glycaemia (HbA1c) in people with type 2 diabetes: cross-sectional study in Australian general practice. BMJ open. 2020 Nov 26;10(11):e039625.

136. Coll-Fernandez R, Coll R, Munoz-Torrero JFS, Aguilar E, Ramon Alvarez L, Sahuquillo

JC, et al. Supervised versus non-supervised exercise in patients with recent myocardial infarction: A propensity analysis. European journal of preventive cardiology. 2016 Feb;23(3):245–52.

156. Denas G, Gennaro N, Ferroni E, Fedeli U, Saugo M, Zoppellaro G, et al. Effectiveness and safety of oral anticoagulation with non-vitamin K antagonists compared to well-managed

vitamin K antagonists in naive patients with non-valvular atrial fibrillation: Propensity score matched cohort study. International journal of cardiology. 2017 Dec 15;249:198–203.

177. Erne P, Radovanovic D, Seifert B, Bertel O, Urban P. Outcome of patients admitted with acute coronary syndrome on palliative treatment: insights from the nationwide AMIS Plus

Registry 1997-2014. BMJ open. 2015 Mar 2;5(3):e006218.

188. Ferroni E, Gennaro N, Costa G, Fedeli U, Denas G, Pengo V, et al. Real-world persistence with direct oral anticoagulants (DOACs) in naive patients with non-valvular atrial

fibrillation. International journal of cardiology. 2019 Aug 1;288:72–5.

199. Fu EL, Desai RJ, Paik JM, Kim DH, Zhang Y, Mastrorilli JM, et al. Comparative Safety and Effectiveness of Warfarin or Rivaroxaban Versus Apixaban in Patients With Advanced

CKD and Atrial Fibrillation: Nationwide US Cohort Study. American journal of kidney diseases : the official journal of the National Kidney Foundation. 2024 Mar;83(3):293-305.e1.

209. Gardener H, Leifheit EC, Lichtman JH, Wang K, Wang Y, Gutierrez CM, et al. Race-Ethnic Disparities in 30-Day Readmission After Stroke Among Medicare Beneficiaries in the

Florida Stroke Registry. Journal of stroke and cerebrovascular diseases : the official journal of National Stroke Association. 2019 Dec;28(12):104399.

242. Hanon O, Vidal JS, Le Heuzey JY, Kirchhof P, De Caterina R, Schmitt J, et al. Oral

anticoagulant use in octogenarian European patients with atrial fibrillation: A subanalysis of PREFER in AF. International journal of cardiology. 2017 Apr 1;232:98–104.

273. Jackson LR 2nd, Kim S, Fonarow GC, Freeman JV, Gersh BJ, Go AS, et al. Stroke Risk

and Treatment in Patients with Atrial Fibrillation and Low CHA(2)DS(2)-VASc Scores: Findings From the ORBIT-AF I and II Registries. Journal of the American Heart Association. 2018 Aug 21;7(16):e008764.

304. Kaul P, Welsh RC, Liu W, Savu A, Weiss DR, Armstrong PW. Temporal and Provincial

Variation in Ambulance Use Among Patients Who Present to Acute Care Hospitals With ST-Elevation Myocardial Infarction. The Canadian journal of cardiology. 2016 Aug;32(8):949–55.

326. Kirchhof P, Haas S, Amarenco P, Hess S, Lambelet M, van Eickels M, et al. Impact of

Modifiable Bleeding Risk Factors on Major Bleeding in Patients With Atrial Fibrillation Anticoagulated With Rivaroxaban. Journal of the American Heart Association. 2020 Mar 3;9(5):e009530.

336. Koton S, Tanne D, Grossman E. Prestroke treatment with beta-blockers for hypertension is not associated with severity and poor outcome in patients with ischemic stroke: data from a

national stroke registry. Journal of hypertension. 2017 Apr;35(4):870–6.

379. Loikas D, Forslund T, Wettermark B, Schenck-Gustafsson K, Hjemdahl P, von Euler M.

Sex and Gender Differences in Thromboprophylactic Treatment of Patients With Atrial Fibrillation After the Introduction of Non-Vitamin K Oral Anticoagulants. The American journal of cardiology. 2017 Oct 15;120(8):1302–8.

Mar;40(1):39–47.

421. Mostaza JM, Suarez C, Cepeda JM, Manzano L, Sanchez D. Demographic, clinical, and

functional determinants of antithrombotic treatment in patients with nonvalvular atrial fibrillation. BMC cardiovascular disorders. 2021 Aug 9;21(1):384.

431. Ngo L, Ali A, Ganesan A, Woodman R, Adams R, Ranasinghe I. Gender differences in complications following catheter ablation of atrial fibrillation. European heart journal

Quality of care & clinical outcomes. 2021 Sept 16;7(5):458–67.

441. O’Neal WT, Claxton JS, Sandesara PB, MacLehose RF, Chen LY, Bengtson LGS, et al. Provider Specialty, Anticoagulation, and Stroke Risk in Patients With Atrial Fibrillation and

Cancer. Journal of the American College of Cardiology. 2018 Oct 16;72(16):1913–22.

452. Ojeda-Fernandez L, Foresta A, Macaluso G, Colacioppo P, Tettamanti M, Zambon A, et al. Metformin use is associated with a decrease in the risk of hospitalization and mortality in

COVID-19 patients with diabetes: A population-based study in Lombardy. Diabetes, obesity & metabolism. 2022 May;24(5):891–8.

462. Ou SM, Chen HT, Kuo SC, Chen TJ, Shih CJ, Chen YT. Dipeptidyl peptidase-4 inhibitors and cardiovascular risks in patients with pre-existing heart failure. Heart (British Cardiac

Society). 2017 Mar;103(6):414–20.

473. Pasceri V, Pelliccia F, Mehran R, Dangas G, Porto I, Radico F, et al. Risk Score for Prediction of Dialysis After Transcatheter Aortic Valve Replacement. Journal of the

American Heart Association. 2024 Apr 2;13(7):e032955.

495. Pu A, Ding L, Shin J, Price J, Skarsgard P, Wong DR, et al. Long-term Outcomes of Multiple Arterial Coronary Artery Bypass Grafting: A Population-Based Study of Patients

in British Columbia, Canada. JAMA cardiology. 2017 Nov 1;2(11):1187–96.

548. Sawano M, Kohsaka S, Abe T, Inohara T, Maekawa Y, Ueda I, et al. Patterns of statin non-

prescription in patients with established coronary artery disease: A report from a contemporary multicenter Japanese PCI registry. PloS one. 2017;12(8):e0182687.

569. Shiraishi Y, Kohsaka S, Ueda I, Inohara T, Sawano M, Numasawa Y, et al. Degree of

dyspnoea in patients with non-ST-elevation acute coronary syndrome: A report from Japanese multicenter registry. International journal of clinical practice. 2016 Dec;70(12):978–87.

591. Soldati S, Di Martino M, Rosa AC, Fusco D, Davoli M, Mureddu GF. The impact of in-hospital cardiac rehabilitation program on medication adherence and clinical outcomes in patients with acute myocardial infarction in the Lazio region of Italy. BMC cardiovascular

disorders. 2021 Sept 27;21(1):466.

613. Takeji Y, Shiomi H, Morimoto T, Furukawa Y, Ehara N, Nakagawa Y, et al. Diabetes

Mellitus and Long-Term Risk for Heart Failure After Coronary Revascularization. Circulation journal : official journal of the Japanese Circulation Society. 2020 Feb 25;84(3):471–8.

15;405:131940.

688. Yamaguchi I, Kanematsu Y, Shimada K, Korai M, Miyamoto T, Shikata E, et al. Active

Cancer and Elevated D-Dimer Are Risk Factors for In-Hospital Ischemic Stroke. Cerebrovascular diseases extra. 2019;9(3):129–38.

retrospective cohort study. CMAJ open. 2020 June;8(2):E437–47.
